## Supplementary material for "Distinctive features of lipoprotein profiles in stroke patients": Fig. S1: box.htm

Support

### Supporting information

for **Human lipoproteins as risk factors**  
  
Jump to S3 and S4.

### S3 Fig. Classes

### CM1

| TG | Cholesterol | TG/CH |
| --- | --- | --- |
| TukeyHSD groups \| diff \| lwr \| upr \| p-adj; 1-0 \| -0.6810614 \| -1.2125628 \| -0.1495601 \| 0.0059705; 2-0 \| -0.3072650 \| -0.8558540 \| 0.2413241 \| 0.4673244; 3-0 \| -0.2138531 \| -1.1003327 \| 0.6726266 \| 0.9233624; 2-1 \| 0.3737965 \| -0.1696187 \| 0.9172117 \| 0.2836347; 3-1 \| 0.4672084 \| -0.4160788 \| 1.3504956 \| 0.5175617; 3-2 \| 0.0934119 \| -0.8002617 \| 0.9870855 \| 0.9929748 | TukeyHSD groups \| diff \| lwr \| upr \| p-adj; 1-0 \| -0.44313859 \| -0.8038432 \| -0.08243395 \| 0.0092339; 2-0 \| -0.45547417 \| -0.8277754 \| -0.08317292 \| 0.0096249; 3-0 \| -0.79762781 \| -1.3992393 \| -0.19601635 \| 0.0040872; 2-1 \| -0.01233558 \| -0.3811256 \| 0.35645441 \| 0.9997639; 3-1 \| -0.35448922 \| -0.9539341 \| 0.24495568 \| 0.4185372; 3-2 \| -0.34215364 \| -0.9486473 \| 0.26434003 \| 0.4608995 | TukeyHSD groups \| diff \| lwr \| upr \| p-adj; 1-0 \| -0.2379229 \| -0.65362892 \| 0.1777832 \| 0.4480053; 2-0 \| 0.1482092 \| -0.28086174 \| 0.5772802 \| 0.8062547; 3-0 \| 0.5837748 \| -0.10957239 \| 1.2771219 \| 0.1314504; 2-1 \| 0.3861321 \| -0.03889222 \| 0.8111564 \| 0.0893808; 3-1 \| 0.8216976 \| 0.13084739 \| 1.5125478 \| 0.0126180; 3-2 \| 0.4355655 \| -0.26340826 \| 1.1345394 \| 0.3711799 |
| 0: stroke, 1: statin, 2: control, 3: old. |  |  |

### CM2

| TG | Cholesterol | TG/CH |
| --- | --- | --- |
| TukeyHSD groups \| diff \| lwr \| upr \| p-adj; 1-0 \| -0.35126118 \| -0.6395021 \| -0.06302027 \| 0.0100013; 2-0 \| -0.16347471 \| -0.4609825 \| 0.13403311 \| 0.4842984; 3-0 \| -0.21176669 \| -0.6925175 \| 0.26898408 \| 0.6627548; 2-1 \| 0.18778647 \| -0.1069155 \| 0.48248843 \| 0.3510879; 3-1 \| 0.13949449 \| -0.3395250 \| 0.61851395 \| 0.8737718; 3-2 \| -0.04829198 \| -0.5329442 \| 0.43636019 \| 0.9938985 | TukeyHSD groups \| diff \| lwr \| upr \| p-adj; 1-0 \| -0.64318078 \| -1.0221548 \| -0.26420673 \| 0.0001144; 2-0 \| -0.42568289 \| -0.8168409 \| -0.03452487 \| 0.0270787; 3-0 \| -0.65641967 \| -1.2885023 \| -0.02433704 \| 0.0385035; 2-1 \| 0.21749790 \| -0.1699710 \| 0.60496682 \| 0.4653710; 3-1 \| -0.01323888 \| -0.6430452 \| 0.61656745 \| 0.9999413; 3-2 \| -0.23073678 \| -0.8679489 \| 0.40647534 \| 0.7830123 | TukeyHSD groups \| diff \| lwr \| upr \| p-adj; 1-0 \| 0.29191961 \| 0.032617404 \| 0.5512218 \| 0.0205494; 2-0 \| 0.26220818 \| -0.005430551 \| 0.5298469 \| 0.0571537; 3-0 \| 0.44465298 \| 0.012168447 \| 0.8771375 \| 0.0413494; 2-1 \| -0.02971143 \| -0.294826001 \| 0.2354032 \| 0.9913773; 3-1 \| 0.15273337 \| -0.278193671 \| 0.5836604 \| 0.7938303; 3-2 \| 0.18244480 \| -0.253549441 \| 0.6184390 \| 0.6979962 |
| 0: stroke, 1: statin, 2: control, 3: old. |  |  |

#### VLDL

| TG | Cholesterol | TG/CH |
| --- | --- | --- |
| TukeyHSD groups \| diff \| lwr \| upr \| p-adj; 1-0 \| 0.03166595 \| -0.1848442 \| 0.2481761 \| 0.9812622; 2-0 \| -0.01136363 \| -0.2348345 \| 0.2121073 \| 0.9991736; 3-0 \| 0.14151319 \| -0.2195994 \| 0.5026257 \| 0.7391815; 2-1 \| -0.04302958 \| -0.2643929 \| 0.1783337 \| 0.9578002; 3-1 \| 0.10984724 \| -0.2499649 \| 0.4696593 \| 0.8575162; 3-2 \| 0.15287682 \| -0.2111662 \| 0.5169199 \| 0.6956487 | TukeyHSD groups \| diff \| lwr \| upr \| p-adj; 1-0 \| -0.04682450 \| -0.5901326 \| 0.4964836 \| 0.9960265; 2-0 \| -0.09668225 \| -0.6574577 \| 0.4640932 \| 0.9699495; 3-0 \| -0.43110733 \| -1.3372792 \| 0.4750645 \| 0.6051060; 2-1 \| -0.04985776 \| -0.6053444 \| 0.5056289 \| 0.9955191; 3-1 \| -0.38428284 \| -1.2871914 \| 0.5186257 \| 0.6866548; 3-2 \| -0.33442508 \| -1.2479507 \| 0.5791006 \| 0.7773221 | TukeyHSD groups \| diff \| lwr \| upr \| p-adj; 1-0 \| 0.078490447 \| -0.4392168 \| 0.5961977 \| 0.9792122; 2-0 \| 0.085318622 \| -0.4490328 \| 0.6196700 \| 0.9758736; 3-0 \| 0.572620523 \| -0.2908521 \| 1.4360932 \| 0.3155233; 2-1 \| 0.006828175 \| -0.5224837 \| 0.5361400 \| 0.9999864; 3-1 \| 0.494130076 \| -0.3662330 \| 1.3544931 \| 0.4448841; 3-2 \| 0.487301901 \| -0.3831780 \| 1.3577818 \| 0.4677888 |
| 0: stroke, 1: statin, 2: control, 3: old. |  |  |

### Lp(a)

| TG | Cholesterol | TG/CH |
| --- | --- | --- |
| TukeyHSD groups \| diff \| lwr \| upr \| p-adj; 1-0 \| -0.08940393 \| -0.2878600 \| 0.10905211 \| 0.6465131; 2-0 \| -0.23411607 \| -0.4389524 \| -0.02927970 \| 0.0180614; 3-0 \| -0.26964165 \| -0.6006422 \| 0.06135888 \| 0.1526177; 2-1 \| -0.14471214 \| -0.3476167 \| 0.05819238 \| 0.2530338; 3-1 \| -0.18023772 \| -0.5100462 \| 0.14957080 \| 0.4891051; 3-2 \| -0.03552558 \| -0.3692123 \| 0.29816110 \| 0.9925845 | TukeyHSD groups \| diff \| lwr \| upr \| p-adj; 1-0 \| -0.16726223 \| -0.2823468 \| -0.05217763 \| 0.0012958; 2-0 \| -0.26329709 \| -0.3820816 \| -0.14451254 \| 0.0000003; 3-0 \| -0.23517849 \| -0.4271256 \| -0.04323139 \| 0.0094843; 2-1 \| -0.09603486 \| -0.2136991 \| 0.02162941 \| 0.1513526; 3-1 \| -0.06791626 \| -0.2591721 \| 0.12333959 \| 0.7928844; 3-2 \| 0.02811860 \| -0.1653862 \| 0.22162340 \| 0.9816098 | TukeyHSD groups \| diff \| lwr \| upr \| p-adj; 1-0 \| 0.07785830 \| -0.08931416 \| 0.2450308 \| 0.6215757; 2-0 \| 0.02918102 \| -0.14336601 \| 0.2017281 \| 0.9715558; 3-0 \| -0.03446316 \| -0.31328648 \| 0.2443602 \| 0.9885124; 2-1 \| -0.04867728 \| -0.21959698 \| 0.1222424 \| 0.8808529; 3-1 \| -0.11232145 \| -0.39014066 \| 0.1654978 \| 0.7201495; 3-2 \| -0.06364418 \| -0.34473021 \| 0.2174419 \| 0.9355276 |
| 0: stroke, 1: statin, 2: control, 3: old. |  |  |

### TR

| TG | Cholesterol | TG/CH |
| --- | --- | --- |
| TukeyHSD groups \| diff \| lwr \| upr \| p-adj; 1-0 \| -0.04088578 \| -0.30609983 \| 0.2243283 \| 0.9781889; 2-0 \| 0.13247886 \| -0.14126178 \| 0.4062195 \| 0.5915632; 3-0 \| 0.42198409 \| -0.02036068 \| 0.8643288 \| 0.0674488; 2-1 \| 0.17336465 \| -0.09779429 \| 0.4445236 \| 0.3480804; 3-1 \| 0.46286987 \| 0.02211810 \| 0.9036216 \| 0.0355012; 3-2 \| 0.28950522 \| -0.15642927 \| 0.7354397 \| 0.3343863 | TukeyHSD groups \| diff \| lwr \| upr \| p-adj; 1-0 \| -0.38297755 \| -1.6483013 \| 0.8823462 \| 0.8606114; 2-0 \| -0.04923864 \| -1.3552423 \| 1.2567651 \| 0.9996622; 3-0 \| -1.15107592 \| -3.2614820 \| 0.9593301 \| 0.4908185; 2-1 \| 0.33373891 \| -0.9599476 \| 1.6274254 \| 0.9082053; 3-1 \| -0.76809837 \| -2.8709043 \| 1.3347076 \| 0.7784832; 3-2 \| -1.10183728 \| -3.2293697 \| 1.0256952 \| 0.5356392 | TukeyHSD groups \| diff \| lwr \| upr \| p-adj; 1-0 \| 0.3420918 \| -0.8738824 \| 1.558066 \| 0.8845935; 2-0 \| 0.1817175 \| -1.0733500 \| 1.436785 \| 0.9818014; 3-0 \| 1.5730600 \| -0.4550369 \| 3.601157 \| 0.1870549; 2-1 \| -0.1603743 \| -1.4036050 \| 1.082856 \| 0.9869867; 3-1 \| 1.2309682 \| -0.7898250 \| 3.251761 \| 0.3917229; 3-2 \| 1.3913425 \| -0.6532129 \| 3.435898 \| 0.2929195 |
| 0: stroke, 1: statin, 2: control, 3: old. |  |  |

#### LDL1

| TG | Cholesterol | TG/CH |
| --- | --- | --- |
| TukeyHSD groups \| diff \| lwr \| upr \| p-adj; 1-0 \| -0.006858426 \| -0.116622339 \| 0.10290549 \| 0.9984720; 2-0 \| -0.099270200 \| -0.212563007 \| 0.01402261 \| 0.1081930; 3-0 \| 0.091836667 \| -0.091236186 \| 0.27490952 \| 0.5623834; 2-1 \| -0.092411774 \| -0.204636093 \| 0.01981255 \| 0.1455195; 3-1 \| 0.098695093 \| -0.083718468 \| 0.28110865 \| 0.4978876; 3-2 \| 0.191106867 \| 0.006548337 \| 0.37566540 \| 0.0393150 | TukeyHSD groups \| diff \| lwr \| upr \| p-adj; 1-0 \| -0.060653111 \| -0.12115671 \| -0.0001495081 \| 0.0491831; 2-0 \| -0.013507641 \| -0.07595643 \| 0.0489411446 \| 0.9431594; 3-0 \| -0.007970661 \| -0.10888331 \| 0.0929419892 \| 0.9969319; 2-1 \| 0.047145470 \| -0.01471435 \| 0.1090052885 \| 0.2001798; 3-1 \| 0.052682451 \| -0.04786679 \| 0.1532316883 \| 0.5257234; 3-2 \| 0.005536980 \| -0.09619460 \| 0.1072685586 \| 0.9989877 | TukeyHSD groups \| diff \| lwr \| upr \| p-adj; 1-0 \| 0.05379469 \| -0.062118638 \| 0.16970801 \| 0.6242724; 2-0 \| -0.08576256 \| -0.205402480 \| 0.03387736 \| 0.2488219; 3-0 \| 0.09980733 \| -0.093521995 \| 0.29313665 \| 0.5383291; 2-1 \| -0.13955724 \| -0.258068816 \| -0.02104567 \| 0.0138405; 3-1 \| 0.04601264 \| -0.146620452 \| 0.23864574 \| 0.9253936; 3-2 \| 0.18556989 \| -0.009328346 \| 0.38046812 \| 0.0682376 |
| 0: stroke, 1: statin, 2: control, 3: old. |  |  |

#### LAC1

| TG | Cholesterol | TG/CH |
| --- | --- | --- |
| TukeyHSD groups \| diff \| lwr \| upr \| p-adj; 1-0 \| -0.07654825 \| -0.4669246 \| 0.31382805 \| 0.9567534; 2-0 \| -0.36385807 \| -0.7667849 \| 0.03906878 \| 0.0923626; 3-0 \| 0.04154561 \| -0.6095546 \| 0.69264579 \| 0.9983734; 2-1 \| -0.28730982 \| -0.6864366 \| 0.11181694 \| 0.2453728; 3-1 \| 0.11809386 \| -0.5306615 \| 0.76684927 \| 0.9649356; 3-2 \| 0.40540368 \| -0.2509803 \| 1.06178768 \| 0.3792158 | TukeyHSD groups \| diff \| lwr \| upr \| p-adj; 1-0 \| -0.18286494 \| -0.7183304 \| 0.3526005 \| 0.8115515; 2-0 \| -0.23625637 \| -0.7889369 \| 0.3164242 \| 0.6836754; 3-0 \| 0.19525020 \| -0.6978410 \| 1.0883414 \| 0.9414566; 2-1 \| -0.05339143 \| -0.6008596 \| 0.4940767 \| 0.9942733; 3-1 \| 0.37811514 \| -0.5117598 \| 1.2679901 \| 0.6877653; 3-2 \| 0.43150657 \| -0.4688323 \| 1.3318454 \| 0.5992940 | TukeyHSD groups \| diff \| lwr \| upr \| p-adj; 1-0 \| 0.10631669 \| -0.2205362 \| 0.4331696 \| 0.8328605; 2-0 \| -0.12760170 \| -0.4649629 \| 0.2097595 \| 0.7596116; 3-0 \| -0.15370459 \| -0.6988555 \| 0.3914463 \| 0.8839315; 2-1 \| -0.23391838 \| -0.5680978 \| 0.1002611 \| 0.2687348; 3-1 \| -0.26002128 \| -0.8032089 \| 0.2831664 \| 0.6002441; 3-2 \| -0.02610289 \| -0.5756778 \| 0.5234720 \| 0.9993260 |
| 0: stroke, 1: statin, 2: control, 3: old. |  |  |

#### LDL2+LAC2

| TG | Cholesterol | TG/CH |
| --- | --- | --- |
| TukeyHSD groups \| diff \| lwr \| upr \| p-adj; 1-0 \| 0.05738289 \| -0.05928458 \| 0.1740504 \| 0.5785368; 2-0 \| 0.08063069 \| -0.03978763 \| 0.2010490 \| 0.3070361; 3-0 \| 0.11437133 \| -0.08021583 \| 0.3089585 \| 0.4240512; 2-1 \| 0.02324780 \| -0.09603483 \| 0.1425304 \| 0.9574871; 3-1 \| 0.05698844 \| -0.13689796 \| 0.2508748 \| 0.8707084; 3-2 \| 0.03374064 \| -0.16242563 \| 0.2299069 \| 0.9701517 | TukeyHSD groups \| diff \| lwr \| upr \| p-adj; 1-0 \| 0.030526442 \| -0.0269986 \| 0.0880514 \| 0.514756; 2-0 \| 0.122289460 \| 0.06291500 \| 0.1816639 \| 0.000001; 3-0 \| 0.039432320 \| -0.0565124 \| 0.1353770 \| 0.709733; 2-1 \| 0.091763018 \| 0.03294853 \| 0.1505775 \| 0.000463; 3-1 \| 0.008905878 \| -0.0866937 \| 0.1045051 \| 0.994997; 3-2 \| -0.08285714 \| -0.1795805 \| 0.0138662 \| 0.121057 | TukeyHSD groups \| diff \| lwr \| upr \| p-adj; 1-0 \| 0.02685645 \| -0.1044386 \| 0.15815151 \| 0.9513270; 2-0 \| -0.04165877 \| -0.1771750 \| 0.09385741 \| 0.8549901; 3-0 \| 0.07493901 \| -0.1440452 \| 0.29392320 \| 0.8106009; 2-1 \| -0.06851522 \| -0.2027533 \| 0.06572288 \| 0.5479522; 3-1 \| 0.04808256 \| -0.1701130 \| 0.26627813 \| 0.9401611; 3-2 \| 0.11659778 \| -0.1041635 \| 0.33735907 \| 0.5188353 |
| 0: stroke, 1: statin, 2: control, 3: old. |  |  |

#### mHDL

| TG | Cholesterol | TG/CH |
| --- | --- | --- |
| TukeyHSD groups \| diff \| lwr \| upr \| p-adj; 1-0 \| 0.21225540 \| -0.4984211 \| 0.9229319 \| 0.8652813; 2-0 \| -0.05187019 \| -0.7853948 \| 0.6816544 \| 0.9977914; 3-0 \| 0.22801638 \| -0.9573055 \| 1.4133383 \| 0.9590202; 2-1 \| -0.26412558 \| -0.9907322 \| 0.4624810 \| 0.7810131; 3-1 \| 0.01576098 \| -1.1652923 \| 1.1968142 \| 0.9999850; 3-2 \| 0.27988657 \| -0.9150545 \| 1.4748276 \| 0.9292673 | TukeyHSD groups \| diff \| lwr \| upr \| p-adj; 1-0 \| -0.17429630 \| -1.0671584 \| 0.7185658 \| 0.9572945; 2-0 \| -0.13579989 \| -1.0573674 \| 0.7857676 \| 0.9808515; 3-0 \| 0.09613465 \| -1.3930507 \| 1.5853200 \| 0.9983161; 2-1 \| 0.03849641 \| -0.8743796 \| 0.9513724 \| 0.9995277; 3-1 \| 0.27043096 \| -1.2133915 \| 1.7542534 \| 0.9648147; 3-2 \| 0.23193455 \| -1.2693359 \| 1.7332050 \| 0.9780539 | TukeyHSD groups \| diff \| lwr \| upr \| p-adj; 1-0 \| 0.38655170 \| -0.4470833 \| 1.2201867 \| 0.6249287; 2-0 \| 0.08392971 \| -0.7765065 \| 0.9443659 \| 0.9942700; 3-0 \| 0.13188173 \| -1.2585200 \| 1.5222834 \| 0.9947244; 2-1 \| -0.30262199 \| -1.1549432 \| 0.5496993 \| 0.7929552; 3-1 \| -0.25466997 \| -1.6400645 \| 1.1307245 \| 0.9639489; 3-2 \| 0.04795202 \| -1.3537331 \| 1.4496371 \| 0.9997474 |
| 0: stroke, 1: statin, 2: control, 3: old. |  |  |

#### HDL1

| TG | Cholesterol | TG/CH |
| --- | --- | --- |
| TukeyHSD groups \| diff \| lwr \| upr \| p-adj; 1-0 \| -0.120801405 \| -0.25123361 \| 0.009630805 \| 0.0802563; 2-0 \| 0.210913357 \| 0.07628777 \| 0.345538944 \| 0.0004349; 3-0 \| 0.005797464 \| -0.21174759 \| 0.223342522 \| 0.9998805; 2-1 \| 0.331714762 \| 0.19835886 \| 0.465070667 \| 0.0000000; 3-1 \| 0.126598868 \| -0.09016275 \| 0.343360491 \| 0.4297773; 3-2 \| -0.205115893 \| -0.42442638 \| 0.014194590 \| 0.0758557 | TukeyHSD groups \| diff \| lwr \| upr \| p-adj; 1-0 \| 0.7441361 \| 0.09761364 \| 1.3906585 \| 0.0169904; 2-0 \| 1.6164066 \| 0.94909858 \| 2.2837147 \| 0.0000000; 3-0 \| 1.1949565 \| 0.11663573 \| 2.2732773 \| 0.0234185; 2-1 \| 0.8722705 \| 0.21125601 \| 1.5332851 \| 0.0043130; 3-1 \| 0.4508204 \| -0.62361705 \| 1.5252579 \| 0.6962091; 3-2 \| -0.4214501 \| -1.50852172 \| 0.6656215 \| 0.7455088 | TukeyHSD groups \| diff \| lwr \| upr \| p-adj; 1-0 \| -0.8649375 \| -1.5376399 \| -0.19223506 \| 0.0057547; 2-0 \| -1.4054933 \| -2.0998230 \| -0.71116355 \| 0.0000029; 3-0 \| -1.1891591 \| -2.3111449 \| -0.06717323 \| 0.0331671; 2-1 \| -0.5405558 \| -1.2283371 \| 0.14722558 \| 0.1774533; 3-1 \| -0.3242216 \| -1.4421668 \| 0.79372371 \| 0.8751045; 3-2 \| 0.2163342 \| -0.9147568 \| 1.34742521 \| 0.9596793 |
| 0: stroke, 1: statin, 2: control, 3: old. |  |  |

#### HDL2

| TG | Cholesterol | TG/CH |
| --- | --- | --- |
| TukeyHSD groups \| diff \| lwr \| upr \| p-adj; 1-0 \| 0.01578431 \| -0.4896155 \| 0.5211842 \| 0.9998077; 2-0 \| 0.20819169 \| -0.3134567 \| 0.7298401 \| 0.7281584; 3-0 \| -1.04845570 \| -1.8914012 \| -0.2055102 \| 0.0081549; 2-1 \| 0.19240737 \| -0.3243212 \| 0.7091360 \| 0.7682376; 3-1 \| -1.06424001 \| -1.9041498 \| -0.2243302 \| 0.0067290; 3-2 \| -1.25664738 \| -2.1064335 \| -0.4068612 \| 0.0010203 | TukeyHSD groups \| diff \| lwr \| upr \| p-adj; 1-0 \| 0.2658196 \| -0.03507669 \| 0.5667159 \| 0.1037451; 2-0 \| 0.3705950 \| 0.06002492 \| 0.6811651 \| 0.0122390; 3-0 \| 0.1301791 \| -0.37167933 \| 0.6320375 \| 0.9068519; 2-1 \| 0.1047754 \| -0.20286564 \| 0.4124164 \| 0.8128013; 3-1 \| -0.1356405 \| -0.63569162 \| 0.3644106 \| 0.8951279; 3-2 \| -0.2404159 \| -0.74634701 \| 0.2655152 \| 0.6060134 | TukeyHSD groups \| diff \| lwr \| upr \| p-adj; 1-0 \| -0.25003529 \| -0.7833813 \| 0.28331075 \| 0.6165323; 2-0 \| -0.16240330 \| -0.7128963 \| 0.38808974 \| 0.8694731; 3-0 \| -1.17863478 \| -2.0681911 \| -0.28907846 \| 0.0041174; 2-1 \| 0.08763199 \| -0.4576692 \| 0.63293322 \| 0.9754216; 3-1 \| -0.92859949 \| -1.8149523 \| -0.04224669 \| 0.0361310; 3-2 \| -1.01623148 \| -1.9130067 \| -0.11945622 \| 0.0194384 |
| 0: stroke, 1: statin, 2: control, 3: old. |  |  |

#### Free Glycerol

Glycerol

TukeyHSD

|  |  |  |  |  |
| --- | --- | --- | --- | --- |
| groups | diff | lwr | upr | p-adj |
| 1-0 | -0.10659677 | -0.2374917 | 0.02429814 | 0.1528280 |
| 2-0 | -0.19385482 | -0.3289580 | -0.05875165 | 0.0015393 |
| 3-0 | -0.42482680 | -0.6431436 | -0.20651002 | 0.0000073 |
| 2-1 | -0.08725804 | -0.2210870 | 0.04657093 | 0.3305502 |
| 3-1 | -0.31823003 | -0.5357606 | -0.10069946 | 0.0011850 |
| 3-2 | -0.23097198 | -0.4510605 | -0.01088351 | 0.0356832 |

0: stroke, 1: statin, 2: control, 3: old.

| Glycerol to CM1 ratio | Glycerol to TR | Glycerol to LDL1 | Glycerol to LDL2+LAC2 |
| --- | --- | --- | --- |
| Correlation to glycerol |  |  |  |
| class \| Pearson's r; CM1 \| -0.167529150865999; CM2 \| -0.0841126067569958; VLDL \| -0.0814365817125813; Lp(a) \| -0.216746854774793; TR \| -0.207463680240653; LDL1 \| 0.0162021823361841; LAC1 \| -0.0732322047123116; LDL2 \| -0.175758937353437; LAC2 \| -0.086804455339854; mHDL \| -0.183683185654165; HDL1 \| -0.0980736922032977; HDL2 \| 0.0732104080735861 |  |  |  |

  
  

| Glycerol and age | Glycerol and age of 65 |
| --- | --- |
|  | TukeyHSD groups \| diff \| lwr \| upr \| p-adj; 1-0 \| 0.13071375 \| -0.03943972 \| 0.300867226 \| 0.1941940; 2-0 \| -0.23097198 \| -0.45425102 \| -0.007692946 \| 0.0395941; 3-0 \| 0.14211208 \| 0.01755029 \| 0.266673867 \| 0.0183433; 2-1 \| -0.36168574 \| -0.60503819 \| -0.118333279 \| 0.0009503; 3-1 \| 0.01139833 \| -0.14634364 \| 0.169140295 \| 0.9976448; 3-2 \| 0.37308406 \| 0.15911244 \| 0.587055680 \| 0.0000697 |
|  | 0 and 1: under 65, 2 and 3: over 65, 0 and 2: control, 1 and 3: stroke. |

  
  

### S4 Fig. Current methods

| Total TG | Total Cholesterol |
| --- | --- |
| TukeyHSD groups \| diff \| lwr \| upr \| p-adj; 1-0 \| -0.15526944 \| -0.6214609 \| 0.3109220 \| 0.8227774; 2-0 \| -0.37328102 \| -0.8544604 \| 0.1078984 \| 0.1869324; 3-0 \| -0.07210346 \| -0.8496540 \| 0.7054471 \| 0.9950652; 2-1 \| -0.21801158 \| -0.6946529 \| 0.2586297 \| 0.6352573; 3-1 \| 0.08316598 \| -0.6915845 \| 0.8579164 \| 0.9924024; 3-2 \| 0.30117756 \| -0.4826830 \| 1.0850381 \| 0.7507130 | TukeyHSD groups \| diff \| lwr \| upr \| p-adj; 1-0 \| -0.8275592 \| -1.5841213 \| -0.07099712 \| 0.0260051; 2-0 \| 0.1047257 \| -0.6761597 \| 0.88561117 \| 0.9854271; 3-0 \| -0.5635101 \| -1.8253636 \| 0.69834338 \| 0.6528974; 2-1 \| 0.9322849 \| 0.1587642 \| 1.70580568 \| 0.0111315; 3-1 \| 0.2640491 \| -0.9932602 \| 1.52135832 \| 0.9476227; 3-2 \| -0.6682359 \| -1.9403296 \| 0.60385787 \| 0.5235040 |

0: stroke, 1: statin, 2: control, 3: old.

| LDL | HDL |
| --- | --- |
| TukeyHSD groups \| diff \| lwr \| upr \| p-adj; 1-0 \| -1.0754578 \| -1.790797962 \| -0.3601176 \| 0.0008065; 2-0 \| -0.3540259 \| -1.092364172 \| 0.3843124 \| 0.5989332; 3-0 \| -0.7426145 \| -1.935714921 \| 0.4504859 \| 0.3722322; 2-1 \| 0.7214319 \| -0.009942977 \| 1.4528067 \| 0.0547032; 3-1 \| 0.3328433 \| -0.855960487 \| 1.5216470 \| 0.8860344; 3-2 \| -0.3885886 \| -1.591371295 \| 0.8141941 \| 0.8356614 | TukeyHSD groups \| diff \| lwr \| upr \| p-adj; 1-0 \| -0.15526944 \| -0.6214609 \| 0.3109220 \| 0.8227774; 2-0 \| -0.37328102 \| -0.8544604 \| 0.1078984 \| 0.1869324; 3-0 \| -0.07210346 \| -0.8496540 \| 0.7054471 \| 0.9950652; 2-1 \| -0.21801158 \| -0.6946529 \| 0.2586297 \| 0.6352573; 3-1 \| 0.08316598 \| -0.6915845 \| 0.8579164 \| 0.9924024; 3-2 \| 0.30117756 \| -0.4826830 \| 1.0850381 \| 0.7507130 |

0: stroke, 1: statin, 2: control, 3: old.

  
  

index
