## Supplementary material for "Distinctive features of lipoprotein profiles in stroke patients": Fig. S1: Fitdata.htm

Support


### Supporting information

| id | stroke | age | statin | Total TG | Total cholesterol | HDL | LDL | T CM1 | T CM2 | T Lp(a) | T VLDL1 | T TR | T LDL1 | T LAC1 | T LDL2 | T LAC2 | T mHDL | T HDL1 | T HDL2 | C CM1 | C CM2 | C Lp(a) | C VLDL1 | C TR | C LDL1 | C LAC1 | C LDL2 | C LAC2 | C mHDL | C HDL1 | C HDL2 | time CM1 | time CM2 | time Lp(a) | time VLDL1 | time TR | time LDL1 | time LAC1 | time LDL2 | time LAC2 | time mHDL | time HDL1 | time HDL2 | time glycerol | free glycerol |
| --- | --- | --- | --- | --- | --- | --- | --- | --- | --- | --- | --- | --- | --- | --- | --- | --- | --- | --- | --- | --- | --- | --- | --- | --- | --- | --- | --- | --- | --- | --- | --- | --- | --- | --- | --- | --- | --- | --- | --- | --- | --- | --- | --- | --- | --- |
| 1 | 1 | 83 | 1 | 68 | 195 | 64 | 104 | 0.360538678 | 3.245139988 | 10.07085321 | 11.60857992 | 22.27796631 | 10.27394729 | 1.543876354 | 8.730780951 | 3.209155879 | 0.089473451 | 0.692762263 | 0.185060911 | 0.052429368 | 0.492245596 | 3.417777605 | 14.56957108 | 3.264466255 | 100.5941058 | 4.514453189 | 35.98961628 | 22.64729039 | 5.78E-05 | 0.90853562 | 0.571455851 | 17.94921875 | 18.75 | 20.95703125 | 22.109375 | 23.28125 | 23.90625 | 25.35625 | 26.903125 | 27.41875 | 28.49492188 | 28.92363281 | 29.8875 | 33.32 | 0.43302932 |
| 2 | 1 | 45 | 0 | 175 | 186 | 41 | 120 | 5.860891314 | 35.89111893 | 45.79729405 | 45.1618549 | 17.27799015 | 18.36022759 | 3.282849022 | 6.729758639 | 6.385894509 | 1.024435145 | 1.511883115 | 0.421434915 | 0.90666211 | 5.549700048 | 7.344471778 | 22.71852833 | 3.751821211 | 96.99678998 | 4.014754922 | 12.30832641 | 20.38708305 | 5.249424588 | 7.37E-05 | 0.631419183 | 18.1640625 | 19.921875 | 21.0546875 | 22.2265625 | 23.59375 | 24.0625 | 24.996875 | 26.94024576 | 27.4052202 | 28.08330792 | 28.7625 | 29.684375 | 33.32 | 0.498871375 |
| 6 | 1 | 84 | 1 | 174 | 116 | 33 | 61 | 2.240601673 | 4.899762494 | 4.545858826 | 6.363776934 | 4.255465115 | 4.0902889 | 0.394087842 | 3.032973528 | 5.78E-05 | 0.144885483 | 0.241814897 | 5.78E-05 | 2.720630446 | 5.809739744 | 5.176317033 | 13.16898361 | 6.64E-05 | 59.02871632 | 1.72884041 | 26.1170342 | 3 | 5.78E-05 | 1.13761478 | 0.03098589 | 17.44730469 | 19.4089156 | 20.69243878 | 21.85487488 | 22.94465871 | 23.74383353 | 25.42907884 | 26.94024576 | 27.61833348 | 28.2964212 | 28.83889138 | 29.73009238 | 33.02 | 2.110588669 |
| 7 | 1 | 80 | 0 | 112 | 266 | 60 | 189 | 0.947898404 | 3.282523457 | 20.87320703 | 18.02757447 | 25.63856349 | 18.70501853 | 1.967507813 | 10.32596877 | 1.333633728 | 0.061702103 | 1.872435348 | 0.009076064 | 0.412583875 | 1.109617254 | 9.645680482 | 23.45840728 | 6.64E-05 | 178.7535189 | 6.406917502 | 37.79524542 | 11.54448274 | 2.843337464 | 1.648547013 | 0.399205898 | 18.00430532 | 19.65108978 | 21.15256974 | 22.1454839 | 23.25948516 | 23.88913804 | 24.996875 | 26.94024576 | 27.4052202 | 28.08330792 | 28.66452597 | 30.00132747 | 33.32 | 3.698001055 |
| 8 | 1 | 71 | 0 | 79 | 158 | 7 | 55 | 1.027155647 | 5.360693979 | 17.58050709 | 12.24144691 | 21.59110017 | 12.11107583 | 1.420415135 | 12.44314272 | 1.362018974 | 0.868029698 | 1.113015352 | 0.084286198 | 0.29671934 | 1.155794316 | 5.182927625 | 10.73593331 | 1.456402902 | 59.64214716 | 1.446322912 | 45.76909729 | 15.00225061 | 5.713304741 | 8.10E-05 | 0.652646295 | 18.00430532 | 19.65108978 | 21.15256974 | 22.1454839 | 23.25948516 | 23.88913804 | 24.996875 | 26.94024576 | 27.4052202 | 28.08330792 | 28.746875 | 29.7625 | 33.32 | 1.703846121 |
| 9 | 1 | 62 | 0 | 150 | 160 | 50 | 90 | 4.868211827 | 18.86570956 | 56.09217008 | 30.70259901 | 11.83026662 | 7.557500444 | 2.19619059 | 8.952126485 | 5.545986426 | 0.190894176 | 1.992382775 | 0.126776933 | 1.13487209 | 3.86491016 | 11.36521978 | 12.86858677 | 6.64E-05 | 80.69311615 | 2.567250382 | 20.82163388 | 21.57150935 | 5.340607892 | 7.37E-05 | 0.54699129 | 18.10546875 | 19.55078125 | 20.9375 | 22.109375 | 23.37890625 | 24.00390625 | 24.996875 | 26.94024576 | 27.4052202 | 28.08330792 | 28.66452597 | 30.00132747 | 33.32 | 1.39595689 |
| 10 | 1 | 72 | 0 | 104 | 252 | 64 | 176 | 0.718594193 | 7.419660344 | 29.6943532 | 26.07844148 | 18.36966371 | 13.96646768 | 1.737655254 | 6.112811568 | 3.597674473 | 5.78E-05 | 2.142649403 | 0.005791418 | 0.180375041 | 1.17200888 | 6.307779303 | 21.02141248 | 12.45323617 | 139.480962 | 5.939923237 | 27.23913548 | 23.21716964 | 6.482249263 | 6.63E-05 | 0.810527177 | 18.02734375 | 19.82421875 | 21.0546875 | 22.12890625 | 23.30078125 | 24.04296875 | 24.996875 | 26.94024576 | 27.4052202 | 28.08330792 | 28.66452597 | 29.66875 | 33.32 | 0.715234092 |
| 13 | 1 | 65 | 0 | 116 | 146 | 54 | 65 | 2.920451202 | 13.88337105 | 35.39364012 | 19.63899069 | 22.13745993 | 10.83717359 | 2.343824889 | 6.867415157 | 3.938458352 | 6.49E-05 | 1.909991349 | 0.122457019 | 0.382600598 | 1.682420263 | 5.783146452 | 8.444395996 | 6.64E-05 | 68.21393815 | 2.43115617 | 26.74471566 | 21.67858338 | 4.200487771 | 0.500225277 | 0.487350437 | 18.00430532 | 19.7963943 | 21.15256974 | 22.1454839 | 23.25948516 | 23.88913804 | 24.996875 | 26.94024576 | 27.4052202 | 28.08330792 | 28.66452597 | 30.00132747 | 33.32 | 0.609386931 |
| 14 | 1 | 84 | 0 | 132 | 248 | 65 | 146 | 0.911553219 | 5.633032813 | 26.62070074 | 23.68359565 | 25.89391103 | 21.90648702 | 1.312675373 | 18.18448579 | 2.505615777 | 1.158070414 | 0.953919012 | 0.230541875 | 0.215606997 | 2.014453518 | 14.18353692 | 26.82552263 | 12.23330039 | 106.6005501 | 2.786873386 | 43.20854676 | 10.98809118 | 5.026545427 | 6.78E-05 | 0.801281765 | 18.00430532 | 19.65108978 | 21.15256974 | 22.1454839 | 23.25948516 | 23.88913804 | 24.996875 | 26.79375 | 27.4052202 | 28.08330792 | 28.66452597 | 29.746875 | 33.29 | 1.105720871 |
| 17 | 1 | 49 | 0 | 244 | 258 | 40 | 192 | 12.34136751 | 35.10778235 | 89.17864955 | 38.49954588 | 19.74786328 | 23.1755899 | 4.851781965 | 3.959121338 | 7.289279263 | 2.07364918 | 1.5807673 | 0.743647899 | 1.860286165 | 5.817829051 | 17.55276886 | 22.73357959 | 8.863986588 | 160.0062541 | 8.380066244 | 5.992068792 | 21.8029721 | 7.557994505 | 7.37E-05 | 0.658927751 | 18.0859375 | 19.43359375 | 20.9765625 | 22.16796875 | 23.33984375 | 24.0625 | 24.996875 | 26.94024576 | 27.4052202 | 28.08330792 | 28.66452597 | 30.00132747 | 33.32 | 0.69079665 |
| 20 | 1 | 65 | 0 | 610 | 288 | 50 | 156 | 48.95087446 | 90.47750558 | 198.6060036 | 99.99995521 | 28.26343538 | 40.73656184 | 10.46282895 | 10.19326001 | 6.787959301 | 1.768637977 | 3.500647142 | 1.509962896 | 6.013628214 | 13.56055703 | 20.76511179 | 35.21957413 | 1.532180692 | 144.5089166 | 8.507248782 | 16.51294745 | 21.45607601 | 6.197717208 | 7.37E-05 | 0.943168865 | 17.9296875 | 19.3359375 | 21.0546875 | 22.109375 | 23.37890625 | 24.12109375 | 24.996875 | 26.85625 | 27.4052202 | 28.08330792 | 28.66452597 | 30.00132747 | 33.29 | 1.949583507 |
| 24 | 1 | 77 | 1 | 52 | 224 | 56 | 153 | 0.242450854 | 0.501554429 | 11.92277842 | 4.335366484 | 12.38752679 | 11.58877306 | 0.75 | 4.721832528 | 1.061792472 | 0.43 | 0.555411213 | 0.1616983 | 0.096539362 | 0.042782521 | 5.254855126 | 15.18984785 | 6.64E-05 | 140.7431709 | 4.696740829 | 35.91073006 | 15.06858218 | 0.8 | 0.910544786 | 0.561929816 | 17.91 | 18.82 | 21.34243272 | 22.1484375 | 23.41796875 | 23.92578125 | 25.86623023 | 27.075 | 27.559375 | 28.49492188 | 28.92363281 | 29.8875 | 33.32 | 1.311406364 |
| 25 | 1 | 69 | 0 | 52 | 173 | 46 | 110 | 0.268113253 | 0.989055841 | 6.534207131 | 7.560659615 | 10.17588601 | 17.71228765 | 1.109518028 | 4.0428175 | 2.487837437 | 0.167622675 | 1.612124592 | 0.076583972 | 0.155351793 | 0.61198016 | 3.467678469 | 12.75303602 | 4.521454267 | 97.28684528 | 4.690537864 | 21.03795269 | 18.32811299 | 5.604394408 | 7.37E-05 | 0.675347311 | 18.00430532 | 19.65108978 | 21.15256974 | 22.1454839 | 23.25948516 | 24.04296875 | 24.996875 | 26.94024576 | 27.4052202 | 28.08330792 | 28.66452597 | 29.778125 | 33.29 | 1.441336808 |
| 26 | 1 | 72 | 0 | 144 | 221 | 51 | 134 | 2.257652446 | 13.27871915 | 37.42817631 | 19.20682608 | 22.85286945 | 33.18275444 | 2.548166406 | 13.03046319 | 3.541740589 | 0.316272682 | 2.419833267 | 0.185209491 | 0.527819295 | 3.021538939 | 12.0608649 | 18.8727293 | 6.64E-05 | 132.7057315 | 3.123701916 | 28.57044095 | 14.24752095 | 4.284047299 | 0.639934159 | 0.485434596 | 18.00430532 | 19.65108978 | 21.15256974 | 22.1454839 | 23.25948516 | 23.88913804 | 24.996875 | 26.94024576 | 27.4052202 | 28.08330792 | 28.66452597 | 30.00132747 | 33.29 | 0.885550917 |
| 27 | 1 | 74 | 0 | 462 | 202 | 49 | 71 | 47.89939436 | 42.3755009 | 252.4039333 | 30.68574886 | 9.435420973 | 42.69987178 | 4.797398201 | 21.54866399 | 10.99621053 | 2.8 | 0.25 | 1.165202219 | 8.786964544 | 8.489617247 | 43.5177411 | 15.77302284 | 6.64E-05 | 73.78734551 | 2.599376262 | 19.55889174 | 20.65524353 | 3.018523382 | 2.17E-05 | 0.610409299 | 17.8125 | 19.140625 | 21.171875 | 22.421875 | 23.30188 | 24.08203125 | 25.87924 | 26.965625 | 27.496875 | 28.49478 | 28.93554 | 29.88838 | 33.32 | 1.538273242 |
| 29 | 1 | 89 | 0 | 91 | 209 | 70 | 116 | 1.103259942 | 3.867729558 | 35.0319336 | 7.717380799 | 15.9486405 | 13.9927188 | 0.8 | 7.959315589 | 0.989476334 | 0.22 | 0.370742235 | 0.064028981 | 0.344653906 | 0.699977293 | 10.26937761 | 14.60081859 | 8.531651302 | 98.1373108 | 2.2 | 53.0768864 | 10.84367041 | 2 | 7.37E-05 | 0.461082339 | 17.91 | 19.1796875 | 21.15234375 | 22.08984375 | 23.30177132 | 23.97137633 | 25.87943088 | 26.8875 | 27.496875 | 28.49553223 | 28.91966553 | 29.8875 | 33.32 | 2.050612382 |
| 31 | 1 | 82 | 1 | 57 | 151 | 52 | 82 | 0.093649594 | 0.226124426 | 8.489404629 | 4.619381046 | 5.84924291 | 18.52710336 | 0.651534881 | 10.08138235 | 5.78E-05 | 0.7 | 0.4 | 0.272158407 | 0.140206938 | 0.074764338 | 5.990478748 | 11.05928722 | 6.64E-05 | 74.82182447 | 0.328097194 | 46.00599238 | 5.853236313 | 5.78E-05 | 0.220935198 | 0.515009249 | 17.91 | 18.82 | 21.2890625 | 22.2265625 | 23.359375 | 23.92578125 | 25.86 | 27.04375 | 27.6 | 28.5125 | 28.903125 | 29.903125 | 33.29 | 2.708522142 |
| 33 | 1 | 68 | 0 | 144 | 190 | 44 | 124 | 5.698777119 | 6.688426624 | 48.22009986 | 14.14631352 | 22.24194917 | 12.87126241 | 1.878397205 | 7.802809937 | 3.602393129 | 0.620283282 | 0.4 | 0.242067445 | 1.280843882 | 1.512651398 | 13.97738562 | 18.84174398 | 4.560713535 | 97.729588 | 4.265267702 | 24.42891331 | 13.40804491 | 1 | 0.5 | 0.464966986 | 17.91 | 19.2878285 | 21.11328125 | 22.16773499 | 23.30177132 | 23.97137633 | 25.79375 | 27.05189631 | 27.54375 | 28.49553223 | 28.91966553 | 29.8875 | 33.32 | 0.992799659 |
| 35 | 1 | 96 | 0 | 56 | 133 | 41 | 72 | 0.014619386 | 0.06010712 | 4.003338408 | 1.126710688 | 5.384495573 | 16.16766664 | 1.399787966 | 3.028345837 | 0.3 | 0.01 | 0.12 | 0.004913028 | 0.058147692 | 0.082844644 | 3.504115319 | 5.085716351 | 6.64E-05 | 69.43872819 | 8.656768426 | 33.70533826 | 2.425902898 | 0.5 | 8.58E-05 | 0.350198979 | 17.91 | 18.82 | 21.6015625 | 22.1875 | 23.30177132 | 23.88671875 | 25.7625 | 26.965625 | 27.54375 | 28.49553223 | 29.028125 | 29.8875 | 33.32 | 9.322873269 |
| 36 | 1 | 92 | 0 | 68 | 141 | 50 | 69 | 0.586457076 | 3.062607373 | 4.347674912 | 7.410252802 | 4.195853095 | 19.51564092 | 0.322940449 | 5.628315131 | 5.78E-05 | 0.055318608 | 0.620232176 | 5.78E-05 | 0.407762161 | 1.959610002 | 3.529986827 | 12.78571344 | 6.64E-05 | 76.18813986 | 5.78E-05 | 47.12274651 | 3.622346342 | 5.78E-05 | 1.454121927 | 0.14405572 | 17.64104404 | 19.62687237 | 20.69243878 | 21.73378778 | 22.87200646 | 23.50165934 | 25.42907884 | 26.70775854 | 27.23085479 | 28.02518612 | 28.66452597 | 29.73009238 | 33.02 | 1.145174647 |
| 37 | 1 | 62 | 1 | 46 | 252 | 68 | 164 | 6.64E-05 | 1.311927932 | 10.10647465 | 4.867014705 | 8.575156343 | 14.85576238 | 0.922960832 | 2.528443555 | 0.874447247 | 0.5 | 0.6 | 0.222650326 | 0.179094154 | 0.374769103 | 5.800184758 | 21.06322722 | 6.64E-05 | 150.7299195 | 5.178412926 | 42.45349157 | 20.71320893 | 0.5 | 1.3 | 0.720042238 | 17.9143 | 19.07239 | 21.21323 | 22.16796875 | 23.1640625 | 24.04296875 | 25.746875 | 27.059375 | 27.58228 | 28.49478 | 28.93554 | 29.88838 | 33.29 | 1.641720193 |
| 38 | 1 | 65 | 0 | 70 | 212 | 67 | 140 | 1.128268328 | 7.431313029 | 8.207398704 | 15.5076182 | 7.908569254 | 13.09968611 | 1.078561525 | 6.134346392 | 0.093399166 | 5.78E-05 | 1.022711908 | 0.106314253 | 0.610204509 | 4.934265126 | 4.767005621 | 34.72068278 | 3.605589669 | 97.72364481 | 4.236625297 | 52.30346635 | 6.076797429 | 1.638653499 | 7.37E-05 | 1.116643527 | 17.7136963 | 19.74795946 | 20.78930846 | 21.78222262 | 22.99309355 | 23.67118127 | 25.33220917 | 26.86275002 | 27.56021168 | 27.96706431 | 28.49016055 | 29.38136156 | 33.02 | 0.851202187 |
| 40 | 1 | 52 | 0 | 281 | 226 | 40 | 138 | 7.105478138 | 9.446205897 | 5.60444135 | 6.467555756 | 2.549118436 | 3.340901967 | 0.712464985 | 2.082686342 | 0.3191275 | 0.07918545 | 0.29570061 | 0.129320422 | 13.08798492 | 18.39527019 | 11.80184883 | 30.29789965 | 6.64E-05 | 105.2275386 | 5.515034489 | 29.9108014 | 6 | 5.78E-05 | 1.85192602 | 0.40756103 | 17.47152211 | 19.26361109 | 20.64400395 | 21.90330971 | 23.13839806 | 23.76805095 | 25.2934613 | 26.94024576 | 27.69582922 | 28.21892546 | 28.80014351 | 29.94320567 | 33.02 | 1.443420092 |
| 41 | 1 | 47 | 0 | 118 | 192 | 45 | 133 | 1.936774819 | 7.653370323 | 10.34140776 | 6.179997308 | 4.506792448 | 6.233983318 | 0.54669982 | 3.905324294 | 5.78E-05 | 5.78E-05 | 0.587994734 | 0.020385649 | 1.927483755 | 7.388576707 | 12.18285627 | 23.53746036 | 6.64E-05 | 102.9581525 | 2.905931494 | 36.21412911 | 4 | 5.78E-05 | 1.936338566 | 0.267248986 | 17.421875 | 19.19921875 | 20.72265625 | 21.796875 | 22.79296875 | 23.61328125 | 25.325 | 26.95 | 27.54375 | 28.215625 | 28.5125 | 28.98125 | 33.02 | 1.243059967 |
| 42 | 1 | 66 | 0 | 79 | 153 | 81 | 54 | 4.801953827 | 6.38019319 | 13.46182083 | 2.362452823 | 4.342395576 | 10.65290601 | 7.45E-05 | 9.575245459 | 0.139858903 | 0.027995534 | 0.961073227 | 0.083355649 | 1.563422664 | 1.88872761 | 6.402934028 | 5.502048801 | 6.64E-05 | 48.4695386 | 5.78E-05 | 81.51241109 | 7.223784428 | 0.383771965 | 2.128499795 | 0.282009825 | 17.51953125 | 19.00390625 | 20.83984375 | 21.796875 | 22.75390625 | 23.49609375 | 24.934375 | 26.5125 | 27.04375 | 27.60625 | 28.465625 | 29.7 | 33.02 | 5.795027418 |
| 44 | 1 | 85 | 1 | 59 | 244 | 82 | 152 | 6.64E-05 | 0.940044794 | 13.01892836 | 4.82899052 | 8.136569119 | 18.73842855 | 1.211909963 | 8.378865092 | 1.978729779 | 0.05 | 0.856803234 | 0.234175938 | 0.204394687 | 6.98E-05 | 7.510217697 | 15.89267495 | 6.64E-05 | 137.4463885 | 5.541280048 | 55.61081291 | 17.33768592 | 0.557980982 | 0.881794603 | 0.638935891 | 17.91 | 18.82 | 21.23046875 | 22.08984375 | 23.23526774 | 23.86492062 | 25.903125 | 26.90149789 | 27.653125 | 28.496875 | 28.91875 | 29.8875 | 33.32 | 1.386903421 |
| 46 | 1 | 86 | 0 | 180 | 249 | 72 | 150 | 8.6462171 | 15.47870784 | 10.8394701 | 15.9822023 | 4.386031997 | 5.571375345 | 0.83868052 | 6.456864962 | 0.042030099 | 5.78E-05 | 0.890633625 | 0.139922423 | 5.083547189 | 11.09121756 | 6.414555817 | 43.29407449 | 6.64E-05 | 106.7963765 | 1.126646366 | 62.1416921 | 8.252817937 | 5.78E-05 | 2.615786346 | 0.348410802 | 17.51953125 | 19.4921875 | 20.625 | 21.7578125 | 22.96875 | 23.59375 | 24.965625 | 26.7 | 27.2625 | 27.934375 | 28.559375 | 30.090625 | 33.02 | 0.878540821 |
| 53 | 1 | 71 | 0 | 112 | 239 | 51 | 166 | 3.103582861 | 9.805806286 | 7.944893375 | 11.65293229 | 6.082082272 | 5.281743668 | 0.555534884 | 3.758357961 | 0.053177548 | 0.073972098 | 0.489234137 | 0.040205856 | 2.504899876 | 8.657289759 | 7.051804169 | 37.21550622 | 6.64E-05 | 131.9379104 | 3.93189002 | 38.4068513 | 6 | 5.78E-05 | 1.73457337 | 0.413115928 | 17.55859375 | 19.609375 | 20.80078125 | 21.9140625 | 23.0859375 | 23.75 | 25.371875 | 26.91875 | 27.465625 | 28.059375 | 28.575 | 29.85625 | 33.02 | 0.776741989 |
| 54 | 1 | 60 | 0 | 65 | 223 | 58 | 159 | 0.327515328 | 1.638357755 | 11.51941568 | 3.826064388 | 3.58865687 | 22.51432259 | 0.79141956 | 5.28720762 | 0.100066847 | 5.78E-05 | 0.846762246 | 0.020400312 | 0.361345954 | 2.290547605 | 24.00090971 | 21.75741981 | 6.64E-05 | 123.5378485 | 3.027066147 | 44.39772704 | 7.065917513 | 5.78E-05 | 1.916061242 | 0.474069731 | 18.02734375 | 19.51171875 | 21.46484375 | 21.89453125 | 22.83203125 | 23.69140625 | 25.1375 | 26.809375 | 27.434375 | 27.903125 | 28.5125 | 29.809375 | 33.02 | 1.574147527 |
| 56 | 1 | 74 | 1 | 69 | 129 | 59 | 50 | 0.22401491 | 0.688744984 | 8.959403309 | 14.69969594 | 16.42668754 | 15.5760393 | 1.091373783 | 13.51264848 | 1.293435211 | 0.273531597 | 0.76848851 | 0.262488504 | 0.039406305 | 0.060998242 | 2.045919454 | 7.122553309 | 6.64E-05 | 53.76953785 | 2.646779558 | 45.16031056 | 9.710202689 | 0.344270561 | 0.8 | 0.422504083 | 17.91 | 18.82 | 21.32209167 | 22.31500583 | 23.13839806 | 23.92578125 | 25.903125 | 27.09523724 | 27.75395103 | 28.496875 | 28.91875 | 29.8875 | 33.32 | 0.390117495 |
| 60 | 1 | 85 | 0 | 72 | 229 | 79 | 133 | 2.503290847 | 5.566381068 | 9.656070799 | 9.509072997 | 13.79888828 | 14.3868449 | 0.5 | 12.56838953 | 5.78E-05 | 5.78E-05 | 1.10195335 | 0.10548209 | 0.820059396 | 2.774602469 | 6.362174191 | 23.14502473 | 15.43600491 | 96.21019526 | 6 | 80.60278895 | 5.78E-05 | 1.246038377 | 1.618319307 | 0.512200667 | 17.56839178 | 19.26361109 | 20.66822137 | 21.58848327 | 22.60561485 | 23.5743116 | 24.59599964 | 26.572141 | 27.07586331 | 27.67645529 | 28.52890842 | 29.84633599 | 33.02 | 1.244324684 |
| 61 | 1 | 63 | 0 | 159 | 182 | 42 | 116 | 6.863254294 | 8.392181029 | 4.110522498 | 9.010164265 | 2.216133892 | 3.723370807 | 0.722835953 | 2.99662765 | 0.259226726 | 0.303629001 | 0.203856896 | 0.069124475 | 7.197201884 | 9.566395092 | 3.929191301 | 25.7303433 | 2.902673574 | 82.60794482 | 4.210549285 | 34.05054885 | 7.301216143 | 6.89E-05 | 1.501791647 | 0.019577811 | 17.44730469 | 19.57843753 | 20.49869943 | 21.78222262 | 22.99309355 | 23.64696385 | 25.06097408 | 26.92087183 | 27.59511719 | 28.54828236 | 28.99388286 | 29.8875 | 33.02 | 1.756094799 |
| 62 | 1 | 59 | 1 | 73 | 198 | 88 | 88 | 6.68E-05 | 0.956701916 | 20.26775887 | 6.30694486 | 14.66945997 | 22.90411108 | 1.468505553 | 9.471665928 | 2.524155248 | 0.62 | 0.6 | 0.275548414 | 0.082125733 | 0.033434282 | 3.310985826 | 8.293437164 | 6.64E-05 | 91.88350633 | 4.404316612 | 57.5428417 | 23.28944956 | 1.8 | 0.6 | 0.571671281 | 17.91 | 18.92578125 | 21.171875 | 22.04861423 | 23.2421875 | 23.92578125 | 25.7971836 | 26.88212396 | 27.559375 | 28.496875 | 28.996875 | 29.8875 | 33.32 | 0.55793248 |
| 63 | 1 | 82 | 0 | 195 | 225 | 48 | 138 | 5.769197068 | 10.71310751 | 7 | 8.2 | 4.343510568 | 4.139584227 | 0.8646412 | 4.260034908 | 0.100194162 | 0.263168667 | 0.312913369 | 0.009237451 | 7.378955855 | 15.40917914 | 10.76051142 | 37.64434606 | 6.64E-05 | 107.7427824 | 4.27532641 | 38.03442622 | 5 | 5.78E-05 | 1.881112847 | 4.14E-05 | 17.39886985 | 19.43313302 | 20.7166562 | 21.80644004 | 23.1868329 | 23.67118127 | 24.9641044 | 26.86275002 | 27.61833348 | 28.37391694 | 28.89701319 | 29.78821419 | 33.02 | 0.955735124 |
| 67 | 1 | 78 | 1 | 101 | 166 | 64 | 90 | 2.327275134 | 4.830396247 | 39.65599368 | 12.03932058 | 17.46884801 | 7.696390699 | 1.083244613 | 6.216809664 | 3.084022857 | 0.65 | 0.4 | 0.2234437 | 0.312173197 | 0.62816131 | 6.485144922 | 8.81416298 | 2.851721395 | 81.84774678 | 2.943495292 | 33.14330705 | 22.75132811 | 2.5 | 7.37E-05 | 0.413964672 | 18.02734375 | 19.35546875 | 21.11328125 | 22.109375 | 23.30401087 | 23.96538989 | 25.73125 | 26.825 | 27.48125 | 28.49492188 | 28.92363281 | 29.8875 | 33.32 | 0.412843581 |
| 70 | 1 | 67 | 0 | 100 | 258 | 60 | 176 | 6.979665891 | 16.94060092 | 7.231545167 | 21.50221602 | 7.290058381 | 22.27812687 | 1.443919625 | 5.721566576 | 4.384041202 | 0.625953934 | 0.296006998 | 0.082544471 | 2.090119638 | 9.928895996 | 6.439695426 | 50.0686085 | 6.64E-05 | 117.2995868 | 4.656145941 | 22.87497083 | 29.44558889 | 1.395350485 | 0.23690117 | 0.265067811 | 17.88321823 | 20.08700332 | 20.91039555 | 22.04861423 | 23.23526774 | 23.88913804 | 25.71968786 | 26.76588035 | 27.38584626 | 28.6838999 | 29.09075253 | 29.98195354 | 33.23 | 1.769339505 |
| 88 | 1 | 84 | 0 | 146 | 188 | 42 | 118 | 18.10732088 | 35.32126673 | 17.14520396 | 27.71778859 | 11.30803787 | 20.60897033 | 2.00600683 | 4.696293357 | 5.912847615 | 0.821081518 | 0.694335938 | 0.162524994 | 4.429052517 | 11.01903176 | 6.571223291 | 30.61363185 | 6.64E-05 | 88.44888271 | 3.592639378 | 10.6112894 | 24.6481822 | 1.294233632 | 0.4 | 0.194067435 | 17.88321823 | 19.91748139 | 20.93461297 | 22.04861423 | 23.23526774 | 23.88913804 | 25.71968786 | 26.88212396 | 27.34709839 | 28.56765629 | 29.09075253 | 29.98195354 | 33.23 | 1.559340195 |
| 89 | 1 | 65 | 1 | 145 | 166 | 55 | 84 | 3.636998375 | 6.508411037 | 53.74913485 | 21.01603998 | 26.9445045 | 16.57055449 | 2.158323662 | 18.33578558 | 3.293714879 | 0.371389068 | 0.6 | 0.436408867 | 0.825252513 | 1.206589415 | 10.64534867 | 10.96179427 | 2.111891536 | 78.37662309 | 2.943645478 | 32.2437161 | 14.30940215 | 0.223876619 | 0.75 | 0.563795442 | 17.91 | 19.16015625 | 21.171875 | 22.28515625 | 23.35635483 | 24.05865997 | 25.87943088 | 27.05189631 | 27.54083774 | 28.49553223 | 28.91966553 | 29.8875 | 33.32 | 0.448424502 |
| 95 | 1 | 83 | 1 | 88 | 185 | 56 | 107 | 3.712251558 | 2.002563556 | 20.54532112 | 4.978226208 | 10.27685735 | 16.16780469 | 1.26667121 | 11.02329385 | 1.472766632 | 0.448903766 | 0.35 | 0.3 | 0.636658894 | 0.274597296 | 6.599902894 | 8.78635215 | 6.64E-05 | 97.80298256 | 2.860540814 | 41.9026088 | 10.05283335 | 1.7 | 0.15 | 0.620914302 | 17.91 | 19.19921875 | 21.15234375 | 22.1484375 | 23.28370257 | 23.86492062 | 25.71968786 | 26.90149789 | 27.52146381 | 28.43203875 | 28.93576105 | 30.090625 | 33.32 | 0.866672514 |
| 99 | 1 | 69 | 1 | 151 | 234 | 64 | 127 | 0.380747051 | 7.615207741 | 51.68814871 | 16.84063023 | 29.27704153 | 27.48702759 | 1.939027495 | 21.03392515 | 3.901443528 | 0.7 | 0.4 | 0.234779634 | 0.287901621 | 1.942658485 | 18.33861367 | 23.73754449 | 12.82828235 | 109.0061464 | 3.692292087 | 39.96126597 | 19 | 0.7 | 0.7 | 0.524634915 | 17.91 | 18.82 | 21.15234375 | 22.16773499 | 23.30177132 | 23.97137633 | 25.87943088 | 26.95 | 27.528125 | 28.49553223 | 28.91966553 | 29.8875 | 33.29 | 1.542503302 |
| 100 | 1 | 70 | 1 | 113 | 193 | 54 | 117 | 6.88E-05 | 6.205657005 | 52.70322308 | 15.07287692 | 17.15837894 | 16.22117616 | 1.415696476 | 10.99819208 | 2.606148595 | 0.7 | 0.4 | 0.170904069 | 0.116492002 | 1.689077356 | 15.02169632 | 18.32985443 | 6.64E-05 | 98.50019332 | 4.209462601 | 33.22409941 | 13.7528985 | 1.8 | 0.4 | 0.520188571 | 17.91 | 18.92578125 | 21.3671875 | 22.28515625 | 23.30177132 | 23.97137633 | 25.87943088 | 27.11461118 | 27.67645529 | 28.49553223 | 28.91966553 | 29.8875 | 33.32 | 0.637325474 |
| 104 | 1 | 78 | 0 | 61 | 166 | 54 | 87 | 1.215461542 | 5.766065245 | 3.668698554 | 9.519864443 | 4.590427816 | 21.04010135 | 1.214080967 | 7.249598126 | 3.499927149 | 0.637804515 | 0.276827509 | 0.038587798 | 0.340403714 | 2.03555747 | 2.305310963 | 16.27795949 | 6.64E-05 | 79.36635919 | 3.717745265 | 28.25383496 | 19.45453418 | 1.154306751 | 0.225013715 | 0.14034403 | 17.88321823 | 20.08700332 | 20.91039555 | 22.04861423 | 23.23526774 | 23.88913804 | 25.71968786 | 26.76588035 | 27.38584626 | 28.6838999 | 29.09075253 | 29.98195354 | 33.23 | 0.69944351 |
| 109 | 1 | 70 | 1 | 110 | 241 | 57 | 161 | 0.361124843 | 2.270757761 | 39.79778136 | 19.23469167 | 30.62179486 | 12.17431268 | 1.338156165 | 11.8533813 | 1.283725805 | 0.636695355 | 0.610369436 | 0.266555373 | 0.120884008 | 0.628152305 | 12.79200006 | 24.36301548 | 4.40E-05 | 145.3497358 | 3.865062384 | 35.04131762 | 14.97591255 | 1.422817501 | 0.398955521 | 0.658905346 | 17.91 | 18.984375 | 21.34765625 | 22.20703125 | 23.33984375 | 24.00390625 | 25.903125 | 27.121875 | 27.54375 | 28.496875 | 28.91875 | 29.73125 | 33.32 | 0.578571569 |
| 113 | 1 | 79 | 1 | 134 | 231 | 72 | 140 | 0.890080904 | 9.272039365 | 53.56392034 | 14.26950348 | 21.51400997 | 13.53214367 | 1.730391068 | 14.64407518 | 1.557219781 | 0.837428566 | 0.3 | 0.245905147 | 0.240989233 | 1.154319647 | 12.64315403 | 17.10395501 | 13.79100241 | 110.6221966 | 5.10672923 | 49.95286823 | 12.30031016 | 2.7 | 0.1 | 0.6 | 17.91 | 19.21875 | 21.0546875 | 22.109375 | 23.30177132 | 23.97137633 | 25.87943088 | 27.05189631 | 27.59511719 | 28.49553223 | 28.91966553 | 29.8875 | 33.32 | 2.270121034 |
| 120 | 1 | 79 | 1 | 148 | 217 | 46 | 135 | 2.697845394 | 7.071351605 | 65.78788268 | 13.22495059 | 22.57629257 | 29.85162559 | 1.04495507 | 18.00724799 | 2.105745208 | 0.266446103 | 0.517571257 | 0.234188853 | 0.857695519 | 1.393893821 | 18.80224295 | 18.96420592 | 8.743164602 | 116.5443279 | 2.147595309 | 31.81217668 | 10.33444507 | 0.341835108 | 0.611628786 | 0.504200964 | 17.79296875 | 19.39453125 | 21.38671875 | 22.16773499 | 23.30177132 | 23.97137633 | 25.715625 | 26.965625 | 27.59511719 | 28.49553223 | 28.91966553 | 29.8875 | 33.32 | 0.546252757 |
| 126 | 1 | 66 | 0 | 90 | 175 | 36 | 115 | 5.299663928 | 15.56389942 | 6.107470427 | 20.15528921 | 14.85688359 | 11.11887444 | 1.48682493 | 2.235112121 | 5.971419443 | 0.829071534 | 0.520810969 | 0.01973012 | 1.746343715 | 7.024713177 | 3.651283116 | 36.04089111 | 4.293059398 | 79.27782973 | 4.139702956 | 5.653376505 | 24.16428745 | 1.297040403 | 0.18 | 0.135758443 | 17.9800879 | 20.13543816 | 20.88617813 | 22.09704906 | 23.47744192 | 23.9617903 | 25.71968786 | 26.84337609 | 27.38584626 | 28.6838999 | 29.09075253 | 29.98195354 | 33.23 | 1.180170937 |
| 128 | 1 | 63 | 1 | 788 | 196 | 54 | 45 | 99.30616073 | 108.7449903 | 336.6433004 | 42.90129535 | 37.78666074 | 28.9188074 | 9.750565352 | 22.83796221 | 13.62359874 | 1.463888079 | 1.5 | 2.38012419 | 13.22175888 | 14.66705571 | 41.90883967 | 11.30504782 | 6.64E-05 | 42.89765699 | 2.706656527 | 20.9580653 | 23.90743191 | 5.78E-05 | 1.2 | 0.870301831 | 17.87109375 | 19.31640625 | 21.03515625 | 22.24609375 | 23.26171875 | 23.984375 | 25.825 | 27.23125 | 27.559375 | 28.49478 | 28.93554 | 29.88838 | 33.32 | 1.042001957 |
| 130 | 1 | 69 | 1 | 80 | 170 | 56 | 85 | 0.64784829 | 1.266336886 | 17.13144898 | 12.35146276 | 20.542704 | 6.852107636 | 1.68380876 | 12.60931123 | 0.913121903 | 0.6 | 0.25 | 0.066507205 | 0.174333522 | 0.184138909 | 5.31565903 | 13.71280937 | 6.616813129 | 70.24821573 | 5.037701639 | 36.79172399 | 7.434206932 | 2 | 7.37E-05 | 0.404218383 | 17.91 | 18.82 | 21.17550851 | 22.16773499 | 23.30177132 | 23.97137633 | 25.87943088 | 27.05189631 | 27.59511719 | 28.49553223 | 28.996875 | 29.8875 | 33.32 | 0.713673306 |
| 133 | 1 | 84 | 1 | 103 | 179 | 57 | 104 | 0.104205016 | 0.970188745 | 20.54684459 | 22.06321429 | 36.42242215 | 13.68132815 | 1.223028595 | 12.98223234 | 2.181304274 | 0.518946806 | 0.45 | 0.13 | 0.064703674 | 0.352129645 | 6.695198603 | 17.02410592 | 28.9350485 | 64.65996026 | 3.053127741 | 39.24283849 | 12.08921329 | 2.0296842 | 0.2 | 0.64898113 | 17.91 | 19.375 | 21.17550851 | 22.16773499 | 23.30177132 | 23.97137633 | 25.87943088 | 27.0125 | 27.59511719 | 28.49553223 | 28.91966553 | 29.8875 | 33.32 | 1.150720037 |
| 139 | 1 | 81 | 0 | 111 | 153 | 45 | 88 | 6.416962981 | 24.63826397 | 19.42330477 | 26.99487794 | 15.85096172 | 9.901375305 | 0.859322937 | 7.135765281 | 8.2479749 | 1.031465191 | 0.206735271 | 0.146225626 | 0.982977685 | 5.565052902 | 1.408184342 | 26.21727751 | 0.227472543 | 66.03139759 | 3.17889703 | 8.289114446 | 32.50395927 | 1.307285566 | 0.17733505 | 0.219346287 | 18.10117499 | 20.0627859 | 21.12835232 | 22.19391874 | 23.55009418 | 24.03444255 | 25.71968786 | 26.84337609 | 27.28897659 | 28.6838999 | 29.09075253 | 29.98195354 | 33.23 | 0.884358331 |
| 140 | 1 | 63 | 0 | 64 | 221 | 73 | 129 | 2.486413428 | 11.24905678 | 9.349271005 | 9.95890938 | 7.430683274 | 11.7277381 | 1.068476573 | 3.366957359 | 3.409097964 | 0.619259553 | 0.384993594 | 0.005161246 | 1.013220324 | 4.331582036 | 5.003614282 | 26.90778333 | 6.64E-05 | 99.75161248 | 5.787156075 | 20.21506633 | 43.50112406 | 1.781627534 | 0.5 | 0.26179597 | 18.00430532 | 20.08700332 | 21.03148264 | 22.04861423 | 23.23526774 | 23.88913804 | 25.71968786 | 26.86275002 | 27.25022872 | 28.49016055 | 29.09075253 | 29.98195354 | 33.23 | 1.405126211 |
| 141 | 1 | 91 | 0 | 103 | 189 | 47 | 115 | 3.075861705 | 10.15045402 | 10.22892175 | 30.9855272 | 11.84047109 | 21.54311381 | 1.346730224 | 5.482143191 | 4.48856407 | 0.620411213 | 0.228027313 | 0.069179728 | 1.016684861 | 4.349427056 | 3.196985523 | 31.29546038 | 0.58583267 | 88.20736669 | 4.1384376 | 19.8711549 | 22.6165845 | 1.281322268 | 0.119305419 | 0.182008674 | 17.88321823 | 20.08700332 | 20.91039555 | 22.04861423 | 23.23526774 | 23.88913804 | 25.71968786 | 26.76588035 | 27.38584626 | 28.6838999 | 29.09075253 | 29.98195354 | 33.23 | 1.165649699 |
| 149 | 1 | 81 | 0 | 104 | 141 | 39 | 80 | 7.192115928 | 24.61274411 | 22.15909041 | 33.90619133 | 12.66390712 | 10.50659932 | 1.312948433 | 4.071506154 | 8.213534501 | 0.774357683 | 0.457950704 | 0.354951686 | 1.949151553 | 6.328348029 | 3.229913762 | 24.9120805 | 1.373436232 | 63.55119341 | 2.545851579 | 6.462935709 | 27.53831622 | 1.614199743 | 0.22236496 | 0.168426682 | 17.88321823 | 19.94169881 | 20.91039555 | 21.97596197 | 23.33213741 | 23.9617903 | 25.71968786 | 26.82400215 | 27.30835052 | 28.47078662 | 29.01325679 | 29.98195354 | 33.23 | 0.890572524 |
| 157 | 1 | 67 | 1 | 515 | 235 | 38 | 99 | 55.44473046 | 61.49046269 | 148.5329881 | 99.99995521 | 9.850866146 | 31.57703853 | 10 | 9.740106859 | 8.069301461 | 1.5 | 0.25 | 1.505038235 | 8.180022434 | 13.26841947 | 23.18082762 | 35.37938248 | 3.725681929 | 97.74850414 | 4.603177661 | 15.2894749 | 14.02422189 | 1.076899207 | 0.2 | 0.729075083 | 17.87109375 | 19.23828125 | 20.91796875 | 22.24609375 | 23.359375 | 24.08203125 | 26.090625 | 27.1375 | 27.621875 | 28.6375 | 29.2 | 29.871875 | 33.32 | 0.70823933 |
| 161 | 1 | 77 | 1 | 89 | 160 | 61 | 80 | 1.991156652 | 3.584680548 | 32.96781546 | 4.759258801 | 12.26620438 | 11.66252366 | 1.065567023 | 6.146603236 | 3.111179613 | 0.638407759 | 0.261147908 | 0.046321296 | 0.380973523 | 0.54423038 | 7.98839461 | 9.733350637 | 1.76498467 | 70.69391889 | 4.253314324 | 28.82772905 | 22.80928344 | 2.3 | 7.37E-05 | 0.45 | 18.046875 | 19.70703125 | 21.46484375 | 22.16773499 | 23.30177132 | 23.97137633 | 25.934375 | 27.04375 | 27.48125 | 28.575 | 29.10625 | 29.840625 | 33.32 | 0.906707918 |
| 174 | 1 | 79 | 1 | 247 | 277 | 50 | 186 | 19.16950895 | 17.96068079 | 105.0552126 | 26.03278109 | 20.06988138 | 28.89914628 | 4.91663554 | 10.52111429 | 8.064995115 | 0.5 | 0.6 | 0.7 | 4.658089827 | 3.796037528 | 27.04611453 | 27.00725996 | 0.675356486 | 156.1397349 | 5.416074666 | 20.70084836 | 21.72088287 | 0.802118732 | 0.7 | 0.798343189 | 17.9296875 | 19.35546875 | 20.9765625 | 22.16773499 | 23.30177132 | 23.97137633 | 26.23125 | 27.184375 | 27.746875 | 28.465625 | 28.996875 | 29.825 | 33.32 | 2.117382001 |
| 175 | 1 | 93 | 1 | 118 | 171 | 34 | 107 | 4.103705198 | 3.788234606 | 55.61382483 | 9.029758187 | 16.78439387 | 14.24916188 | 1.498819049 | 4.807850309 | 3.560854647 | 0.17 | 0.5 | 0.206398524 | 1.570955953 | 0.903509199 | 17.37236456 | 14.89615366 | 6.64E-05 | 93.35776911 | 3.574183049 | 14.16190322 | 16.02022922 | 1.200008757 | 0.35 | 0.45016646 | 17.91 | 19.453125 | 21.5234375 | 22.3046875 | 23.30177132 | 23.97137633 | 25.934375 | 27.0125 | 27.54375 | 28.49553223 | 28.91966553 | 29.8875 | 33.32 | 1.683623697 |
| 177 | 1 | 72 | 1 | 62 | 198 | 52 | 125 | 0.881473929 | 2.659465018 | 19.01761502 | 4.335516144 | 9.840484604 | 12.24508255 | 0.856821865 | 3.889838455 | 1.94820603 | 6.81E-05 | 0.547151231 | 0.094334294 | 1.623367217 | 6.64E-05 | 11.02810941 | 19.44881344 | 1.458455785 | 109.5825109 | 4.574571523 | 26.57800998 | 18.76181433 | 0.296133595 | 0.9 | 0.494060416 | 17.91 | 19.04296875 | 21.4453125 | 22.16773499 | 23.26171875 | 23.97137633 | 25.934375 | 27.05189631 | 27.59511719 | 28.49553223 | 28.91966553 | 29.8875 | 33.32 | 2.287632299 |
| 187 | 1 | 80 | 0 | 85 | 235 | 68 | 137 | 1.842942742 | 10.8910459 | 5.454860665 | 18.50671521 | 6.202011493 | 22.43104469 | 2.782963145 | 9.656082813 | 6.375216412 | 0.62461074 | 0.443475788 | 0.031163006 | 0.590275807 | 5.166632449 | 4.76018147 | 40.82766915 | 6.64E-05 | 98.80073967 | 4.812340272 | 27.98892942 | 33.4638783 | 1.704069033 | 0.349606379 | 0.307352505 | 17.88321823 | 20.08700332 | 20.91039555 | 22.04861423 | 23.23526774 | 23.79226836 | 25.58407032 | 26.70775854 | 27.30835052 | 28.52890842 | 29.01325679 | 29.71071845 | 33.23 | 1.983402364 |
| 189 | 1 | 70 | 1 | 44 | 211 | 76 | 108 | 1.602627392 | 9.693930215 | 56.7000607 | 16.76730099 | 19.39659084 | 9.546334736 | 1.055502086 | 9.354741702 | 1.671868037 | 0.583979138 | 0.45 | 0.190489964 | 0.365789348 | 1.507957549 | 11.78131711 | 14.45706141 | 5.896309405 | 92.08295978 | 3.696555707 | 52.67284943 | 14.95109481 | 2.726815887 | 0.2 | 0.534020177 | 17.91 | 19.35546875 | 21.09375 | 22.05078125 | 23.22265625 | 23.92578125 | 25.87943088 | 27.028125 | 27.559375 | 28.434375 | 28.98125 | 29.8875 | 33.32 | 2.346042866 |
| 198 | 1 | 66 | 1 | 246 | 153 | 47 | 74 | 4.745185476 | 12.07992942 | 4.85590454 | 14 | 0.818411043 | 3.842930773 | 1.272290054 | 2.849215039 | 1.103594615 | 5.78E-05 | 0.358682918 | 0.049657637 | 3.933744737 | 10.01656998 | 2.902154406 | 23.35922849 | 6.64E-05 | 67.05029082 | 4.682185383 | 28.74345437 | 8 | 0.941863997 | 0.502152193 | 0.25344059 | 17.47152211 | 19.31204592 | 20.4502646 | 21.83065746 | 22.96887613 | 23.81648578 | 25.99092295 | 26.84337609 | 27.52146381 | 28.49553223 | 28.91966553 | 29.8875 | 33.02 | 0.50578427 |
| 200 | 1 | 76 | 1 | 90 | 145 | 59 | 68 | 1.10070457 | 2.341704347 | 28.56010224 | 15.12141104 | 29.04542638 | 6.650135089 | 0.996269193 | 8.463546862 | 1.768585982 | 0.35 | 0.48 | 0.10268544 | 0.130761364 | 0.259597912 | 4.219093328 | 6.888721259 | 6.64E-05 | 71.84071146 | 3.206384531 | 37.99334363 | 14.81847836 | 1.812355492 | 0.25 | 0.418565265 | 17.91 | 19.53125 | 21.2890625 | 22.24609375 | 23.30177132 | 23.97137633 | 25.87943088 | 27.05189631 | 27.54375 | 28.49553223 | 28.91966553 | 29.8875 | 33.32 | 0.383441578 |
| 206 | 1 | 77 | 1 | 83 | 133 | 49 | 61 | 1.380616758 | 1.997691335 | 13.76333571 | 3.230550988 | 4.539762841 | 9.678599184 | 0.51512741 | 8.025173072 | 1.854875082 | 0.421431129 | 0.441900459 | 5.78E-05 | 0.936417019 | 1.043808563 | 8.558375106 | 7.971389299 | 6.64E-05 | 63.16952786 | 0.57778851 | 35.22016107 | 12.98998488 | 3.172573153 | 0.564154262 | 0.054225746 | 17.35043502 | 19.11830657 | 21.15256974 | 21.90330971 | 22.9204413 | 23.62274643 | 25.44845278 | 26.6302628 | 27.30835052 | 28.06393399 | 28.74202171 | 29.34261369 | 33.02 | 0.848878585 |
| 207 | 1 | 73 | 1 | 249 | 208 | 54 | 120 | 20.28153946 | 74.08550176 | 25 | 65 | 9.724254422 | 20.25994747 | 3.057737925 | 5.283170095 | 9.807245331 | 0.943502368 | 1.054556121 | 0.20209144 | 3.404108849 | 15.17511344 | 2.078669558 | 32.64342312 | 6.64E-05 | 93.07593603 | 5.029299195 | 11.24851618 | 34.82628103 | 2 | 0.48382545 | 0.22727387 | 18.02852274 | 20.13543816 | 20.95883039 | 22.00017939 | 23.40478967 | 24.01022513 | 25.71968786 | 26.82400215 | 27.34709839 | 28.39329088 | 29.01325679 | 29.84633599 | 33.23 | 0.479201292 |
| 209 | 1 | 71 | 1 | 50 | 150 | 63 | 64 | 0.128137614 | 0.810396404 | 1.890204533 | 4.656315547 | 4.556722369 | 32.95432144 | 1.175486742 | 3.135431432 | 2.131990753 | 0.360670978 | 0.215984991 | 0.009438833 | 0.032971075 | 0.271549797 | 0.609697775 | 6.566312361 | 6.64E-05 | 70.42799622 | 5.865993334 | 30.47864229 | 26.00677286 | 1.336450097 | 0.251202112 | 0.168764967 | 17.88321823 | 20.08700332 | 20.91039555 | 22.04861423 | 22.96875 | 23.984375 | 25.71968786 | 26.76588035 | 27.278125 | 28.590625 | 29.09075253 | 29.840625 | 33.23 | 0.856451838 |
| 226 | 1 | 82 | 1 | 180 | 201 | 66 | 113 | 4.437729818 | 10.44911504 | 17.79646967 | 5.931431664 | 6.896286834 | 6.940436998 | 1.147703561 | 6.778644942 | 0.742440459 | 0.669314904 | 0.397442862 | 5.78E-05 | 1.846946225 | 4.027772044 | 10.88960317 | 14.47642236 | 6.64E-05 | 110.2314673 | 3.554525406 | 51.97345522 | 8.866055575 | 0.747441927 | 1.003095984 | 0.18219269 | 17.49573953 | 18.90034981 | 20.7166562 | 21.78222262 | 23.04152839 | 23.64696385 | 25.58407032 | 26.78525428 | 27.52146381 | 28.49553223 | 28.91966553 | 29.8875 | 33.02 | 1.324207194 |
| 227 | 1 | 50 | 1 | 84 | 131 | 61 | 53 | 2.677966659 | 1.071657619 | 18.82431037 | 6.198101781 | 5.504124919 | 9.558004292 | 0.026416498 | 9.028970519 | 0.156945894 | 0.261112682 | 0.666750934 | 0.018670444 | 1.840748739 | 0.461547626 | 11.36603314 | 7.137931863 | 6.64E-05 | 51.97154597 | 0.045320887 | 55.19798832 | 5 | 5.78E-05 | 1.18154396 | 0.404430501 | 17.61682662 | 19.31204592 | 21.12835232 | 21.80644004 | 22.84778904 | 23.59852902 | 25.40970491 | 26.78525428 | 27.56021168 | 28.2964212 | 28.80014351 | 29.80758812 | 33.02 | 1.657529025 |
| 229 | 1 | 81 | 0 | 70 | 208 | 61 | 132 | 2.223056043 | 10.69447449 | 8.027865461 | 13.35689437 | 16.30447199 | 10.06366992 | 1.214378372 | 2.879557345 | 3.494297694 | 0.671729941 | 0.384953013 | 0.088418345 | 0.515792592 | 3.143828168 | 3.021291675 | 23.41143695 | 8.550739203 | 99.58999489 | 6.034051197 | 17.20864959 | 35.69584787 | 2.5 | 7.37E-05 | 0.298700294 | 17.98828125 | 20.08700332 | 20.99609375 | 22.04861423 | 23.45703125 | 23.96484375 | 25.71968786 | 26.825 | 27.340625 | 28.5125 | 28.934375 | 29.559375 | 33.23 | 1.4345572 |
| 236 | 1 | 89 | 0 | 93 | 217 | 39 | 148 | 20.24906608 | 37.83301702 | 15.99802349 | 31.67303902 | 8.480404526 | 18.9314508 | 2.663459532 | 5.610397674 | 6.847232916 | 0.938312746 | 0.627736175 | 0.168269848 | 5.930164849 | 13.09087695 | 4.703796313 | 29.09346561 | 1.796267705 | 110.4921461 | 5.700948433 | 9.870967847 | 22.97954206 | 1.419547427 | 0.562374171 | 0.025653414 | 17.7734375 | 19.9609375 | 20.87890625 | 22.04861423 | 23.23526774 | 23.9453125 | 25.71968786 | 26.76588035 | 27.2625 | 28.528125 | 29.028125 | 29.621875 | 33.23 | 1.832261345 |
| 237 | 1 | 82 | 0 | 116 | 236 | 42 | 169 | 3.255560083 | 20.49018923 | 9.18526686 | 46.13091789 | 6.163051976 | 22.37912173 | 1.785623945 | 3.696643982 | 4.720094658 | 0.443647543 | 0.656680044 | 5.78E-05 | 1.008583367 | 6.780731255 | 6.20E-05 | 49.13069994 | 6.64E-05 | 126.2671187 | 5.655627316 | 11.44716468 | 23.7250052 | 1.91599805 | 0.172663769 | 0.359677013 | 17.8125 | 20.20809041 | 20.83984375 | 22.08984375 | 23.23526774 | 23.9617903 | 25.71968786 | 26.778125 | 27.325 | 28.371875 | 28.903125 | 29.575 | 33.23 | 1.544096214 |
| 241 | 1 | 83 | 1 | 86 | 169 | 61 | 90 | 0.717715915 | 3.730013048 | 10.75117835 | 9.949955008 | 10.84126771 | 6.647739859 | 5.78E-05 | 10.13285767 | 1.961555325 | 0.699842756 | 0.023412711 | 0.032657273 | 0.568967415 | 2.669645264 | 7.733518103 | 16.57685325 | 6.64E-05 | 83.71895053 | 5.78E-05 | 39.6605422 | 19.70352878 | 6.49E-05 | 1.182417623 | 0.244993638 | 17.7136963 | 19.72374204 | 20.86196071 | 21.78222262 | 22.94465871 | 23.55009418 | 25.2934613 | 26.61088887 | 27.11461118 | 28.49553223 | 28.91966553 | 29.8875 | 33.02 | 1.279375842 |
| 248 | 1 | 61 | 1 | 74 | 213 | 68 | 122 | 0.66295045 | 2.0188111 | 13.86303403 | 7.5463268 | 4.206395074 | 22.02767596 | 0.76488457 | 9.180037674 | 5.78E-05 | 5.78E-05 | 0.724234875 | 0.008801115 | 1.150108493 | 1.076182961 | 16.26122976 | 25.76785345 | 6.64E-05 | 108.8236065 | 5.78E-05 | 62.43299576 | 5.775187869 | 5.78E-05 | 1.776699149 | 0.513393808 | 17.3262176 | 19.72374204 | 21.00726523 | 21.90330971 | 22.84778904 | 23.59852902 | 25.70031393 | 26.80462822 | 27.30835052 | 27.90894251 | 28.58703023 | 29.01325679 | 33.02 | 1.725522159 |
| 261 | 1 | 82 | 1 | 54 | 156 | 53 | 86 | 0.511309213 | 2.391157261 | 3.927699412 | 7.045595183 | 6.64E-05 | 16.82220459 | 0.095084306 | 8.13687324 | 0.163761936 | 5.78E-05 | 0.882140859 | 0.067465078 | 0.892606077 | 2.787961218 | 4.977417836 | 20.69041386 | 6.64E-05 | 76.35510283 | 5.78E-05 | 48.21023604 | 6.662418593 | 0.052553518 | 0.458279019 | 0.596277295 | 17.51995695 | 19.60265495 | 20.66822137 | 21.90330971 | 22.89622388 | 23.59852902 | 25.42907884 | 26.70775854 | 27.23085479 | 27.94769038 | 28.52890842 | 29.73009238 | 33.02 | 3.003037681 |
| 262 | 1 | 77 | 1 | 213 | 179 | 46 | 104 | 0.746980578 | 13.71837797 | 9.377320626 | 10 | 2.426927225 | 4.7464875 | 0.66652406 | 2.438385103 | 5.78E-05 | 5.78E-05 | 0.528148142 | 0.007873161 | 0.438032495 | 9.835107573 | 9.422289835 | 26.55821143 | 6.64E-05 | 92.51857206 | 3.676063772 | 35.57411205 | 6.565679402 | 5.78E-05 | 0.266901751 | 0.264451548 | 17.76213113 | 19.62687237 | 20.74087362 | 21.90330971 | 22.9204413 | 23.74383353 | 25.42907884 | 27.0177415 | 27.463342 | 27.94769038 | 28.52890842 | 29.73009238 | 33.02 | 0.880769965 |
| 277 | 1 | 71 | 1 | 63 | 238 | 81 | 140 | 0.622685728 | 4.88549371 | 12.22064787 | 12.25970418 | 9.647593756 | 16.21684074 | 1.119312148 | 1.815054133 | 3.094941323 | 0.696235914 | 0.242705332 | 6.91E-05 | 0.227619509 | 1.347945236 | 4.702012729 | 25.33061759 | 5.653105706 | 111.6908595 | 7.049135611 | 19.54101475 | 48.27705179 | 2.418904987 | 0.274133239 | 0.214299888 | 17.87109375 | 19.74609375 | 20.95703125 | 22.109375 | 23.30078125 | 23.984375 | 25.71968786 | 26.825 | 27.29375 | 28.60625 | 29.278125 | 29.98195354 | 33.23 | 1.269919181 |
| 284 | 1 | 78 | 1 | 88 | 122 | 39 | 64 | 1.4176139 | 10.82334198 | 10.13001618 | 21.69263523 | 12.67121572 | 18.44919565 | 1.00410409 | 4.841717553 | 4.186311717 | 0.600720818 | 0.210338599 | 0.04823802 | 0.243331332 | 1.745945456 | 1.565194374 | 8.356970682 | 6.64E-05 | 65.99762317 | 2.174334744 | 15.5660394 | 20.90654653 | 1.234446556 | 0.096782954 | 0.09616057 | 17.88321823 | 20.08700332 | 20.91039555 | 22.04861423 | 23.23526774 | 23.88913804 | 25.71968786 | 26.76588035 | 27.38584626 | 28.6838999 | 29.09075253 | 29.98195354 | 33.23 | 0.740728326 |
| 288 | 1 | 84 | 1 | 103 | 133 | 51 | 55 | 1.25326753 | 12.27288029 | 10.65790125 | 24.7602159 | 14.86487642 | 19.59406366 | 2.07511701 | 12.86654695 | 7.517926382 | 0.98212536 | 0.274678566 | 0.084083718 | 0.370029105 | 2.947580244 | 2.523660603 | 14.25254373 | 4.204995914 | 50.03538753 | 3.379448368 | 23.73227744 | 20.71728873 | 1.817252991 | 0.087294636 | 0.144402286 | 17.88321823 | 20.08700332 | 20.91039555 | 22.04861423 | 23.23526774 | 23.88913804 | 25.71968786 | 26.76588035 | 27.38584626 | 28.56765629 | 29.09075253 | 29.98195354 | 33.23 | 0.857910377 |
| 291 | 1 | 70 | 0 | 110 | 235 | 79 | 127 | 3.809349753 | 18.61377945 | 10.74243122 | 30.22330916 | 7.82839887 | 18.91244786 | 2.734141672 | 7.192262361 | 4.916328802 | 0.611234741 | 0.293065747 | 0.103497294 | 1.014267187 | 5.136298864 | 6.20E-05 | 32.36481653 | 6.64E-05 | 110.5540851 | 7.127447869 | 35.8537855 | 34.81451928 | 1.599816892 | 0.326434834 | 0.232217251 | 17.88321823 | 20.15965557 | 21.07991748 | 22.12126648 | 23.23526774 | 23.98600771 | 25.71968786 | 26.76588035 | 27.38584626 | 28.6838999 | 29.09075253 | 29.98195354 | 33.23 | 1.095612907 |
| 307 | 1 | 51 | 0 | 67 | 216 | 71 | 129 | 2.81363577 | 9.993018058 | 6.419079273 | 10.88015424 | 7.332989958 | 14.50425271 | 0.978280966 | 3.130638043 | 4.163933775 | 0.843507751 | 0.336568784 | 0.060598147 | 0.536547388 | 2.625665263 | 2.798102983 | 25.71339569 | 6.481716883 | 100.4733864 | 4.26716544 | 18.63092989 | 44.02315987 | 2.242844722 | 0.650270019 | 7.99E-05 | 18.07695757 | 20.08700332 | 21.05570006 | 22.09704906 | 23.23526774 | 23.98600771 | 25.71968786 | 26.80462822 | 27.28897659 | 28.56765629 | 29.05200466 | 29.82696206 | 33.23 | 2.213493694 |
| 308 | 1 | 84 | 0 | 118 | 180 | 62 | 99 | 5.912350064 | 24.31652525 | 15.60815151 | 32.88591959 | 10.26745873 | 14.78761585 | 1.736660299 | 4.13080979 | 5.603521438 | 1.058351545 | 0.332387979 | 0.064602677 | 1.021358294 | 4.604645565 | 1.281236272 | 22.31375491 | 6.64E-05 | 79.3130356 | 4.588697335 | 13.73718946 | 39.7283189 | 2.447871291 | 7.37E-05 | 4.14E-05 | 17.9800879 | 20.08700332 | 20.91039555 | 22.04861423 | 23.23526774 | 23.98600771 | 25.71968786 | 26.76588035 | 27.19210692 | 28.54828236 | 29.09075253 | 29.98195354 | 33.23 | 0.777369935 |
| 309 | 1 | 66 | 0 | 86 | 225 | 70 | 133 | 0.257649041 | 16.20891402 | 14.12144727 | 14.28636719 | 7.882690494 | 24.47314828 | 1.020821942 | 2.028729074 | 2.56254308 | 0.625185652 | 0.243956726 | 0.044402202 | 0.079099564 | 3.860796269 | 6.180154883 | 25.32390192 | 6.64E-05 | 113.5165407 | 5.110169247 | 22.23118542 | 38.78363806 | 1.777377138 | 0.582761994 | 0.197173533 | 17.88321823 | 20.18387299 | 21.07991748 | 22.04861423 | 23.1868329 | 23.88913804 | 25.71968786 | 26.76588035 | 27.32772446 | 28.6838999 | 29.09075253 | 29.98195354 | 33.23 | 2.266401764 |
| 310 | 1 | 82 | 1 | 70 | 135 | 51 | 67 | 1.237031477 | 4.865610002 | 5.538841331 | 18.21135109 | 11.21710902 | 14.01891179 | 1.468608848 | 6.318730672 | 3.659148241 | 0.580749178 | 0.192346559 | 0.044020035 | 0.258875579 | 1.07691035 | 0.462579067 | 10.1531136 | 6.64E-05 | 61.27286291 | 4.017035334 | 24.91344784 | 19.40598765 | 1.152581587 | 0.156098066 | 0.120742265 | 17.88321823 | 20.08700332 | 21.00726523 | 22.12126648 | 23.23526774 | 23.91335546 | 25.71968786 | 26.76588035 | 27.38584626 | 28.6838999 | 29.09075253 | 29.98195354 | 33.23 | 0.541540952 |
| 317 | 1 | 78 | 0 | 142 | 262 | 53 | 179 | 7.458736272 | 33.1247263 | 19.11292665 | 33.25 | 14.97534812 | 17.79994356 | 1.939976708 | 3.451543424 | 6.446659699 | 0.78777208 | 0.509781415 | 0.030108886 | 2.515006202 | 11.7312779 | 6.835953441 | 45 | 3.584045266 | 127.7397956 | 6.113871602 | 11.00292197 | 34.71121474 | 2.345428807 | 0.089688658 | 0.244101002 | 17.94921875 | 20.15625 | 20.9375 | 22.08984375 | 23.33984375 | 23.9453125 | 25.71968786 | 26.809375 | 27.340625 | 28.54375 | 29.0125 | 29.7 | 33.23 | 1.765713098 |
| 321 | 1 | 78 | 0 | 74 | 165 | 66 | 69 | 2.723794968 | 7.787038316 | 7.804738646 | 9.209415205 | 14.07837664 | 10.42684307 | 1.941598874 | 10.06837043 | 4.39065151 | 0.652017234 | 0.333395589 | 0.043935608 | 1.022415916 | 2.664967344 | 5.001665704 | 13.86648685 | 10.09119946 | 53.62086161 | 5.981696409 | 35.84833873 | 25.82445519 | 1.644957619 | 0.400669994 | 0.153851434 | 17.9800879 | 20.08700332 | 21.05570006 | 22.12126648 | 23.40478967 | 23.81648578 | 25.71968786 | 26.76588035 | 27.38584626 | 28.54828236 | 29.09075253 | 29.98195354 | 33.23 | 2.83820626 |
| 328 | 1 | 68 | 1 | 374 | 202 | 56 | 84 | 79.81565604 | 89.44101692 | 14.14544218 | 52.28407758 | 9.882691648 | 37.3841093 | 6.147018901 | 25.11971941 | 18.57046048 | 1.99633742 | 0.604478791 | 0.5 | 19.37085135 | 21.36130831 | 1.985839539 | 21.31610396 | 0.086531235 | 60.00599483 | 4.341774008 | 22.27797254 | 27.57764728 | 1.081381399 | 1.123866464 | 0.103092664 | 17.68947888 | 20.08700332 | 20.91039555 | 22.04861423 | 23.23526774 | 23.98600771 | 25.71968786 | 26.70775854 | 27.38584626 | 28.60640416 | 29.09075253 | 29.78821419 | 33.23 | 1.807234535 |
| 330 | 1 | 81 | 1 | 79 | 127 | 40 | 67 | 0.350449432 | 3.953468908 | 7.367746979 | 19.55588309 | 14.25404698 | 22.54020346 | 1.374837735 | 4.188024364 | 4.581316727 | 0.722850067 | 0.407222404 | 0.00184263 | 0.175022275 | 1.753648028 | 2.635425069 | 15.83871596 | 6.64E-05 | 56.62748886 | 4.192971261 | 12.9868512 | 20.92235679 | 1.395812389 | 0.424926887 | 0.083494637 | 17.88321823 | 20.08700332 | 21.015625 | 22.12890625 | 23.3203125 | 23.88913804 | 25.71968786 | 26.825 | 27.325 | 28.496875 | 29.090625 | 29.825 | 33.23 | 0.998458266 |
| 333 | 1 | 57 | 0 | 70 | 241 | 58 | 172 | 0.841520911 | 10.83896973 | 9.119465872 | 18.34756088 | 8.321850747 | 14.25380587 | 1.012778865 | 2.068206839 | 4.182714404 | 0.663404383 | 0.34063094 | 0.038250795 | 0.333611601 | 3.533691611 | 2.366450058 | 29.91350767 | 6.64E-05 | 139.8742496 | 6.956296264 | 9.915453512 | 38.40265861 | 2 | 0.503851183 | 0.228819297 | 17.88321823 | 20.20809041 | 20.93461297 | 22.04861423 | 23.23526774 | 23.9617903 | 25.71968786 | 26.84337609 | 27.28897659 | 28.47078662 | 29.09075253 | 29.98195354 | 33.23 | 1.164800192 |
| 356 | 1 | 42 | 1 | 73 | 146 | 56 | 68 | 1.738152323 | 10.77342624 | 5.409492823 | 23.66623633 | 11.62887326 | 11.44993152 | 1.295116309 | 2.66385478 | 4.310348702 | 0.495137859 | 0.497243888 | 0.034942743 | 0.602391918 | 2.777768513 | 0.241985964 | 16.32586596 | 6.64E-05 | 63.68593801 | 3.984770246 | 12.76855839 | 35.38037467 | 2.24753225 | 0.474896943 | 0.21867782 | 17.88321823 | 20.20809041 | 20.93461297 | 22.04861423 | 23.23526774 | 23.9617903 | 25.71968786 | 26.84337609 | 27.28897659 | 28.35454301 | 29.09075253 | 29.98195354 | 33.23 | 1.414349941 |
| 360 | 1 | 70 | 0 | 86 | 151 | 47 | 90 | 4.46141037 | 14.01072735 | 10.90850339 | 14.42051564 | 15.88964705 | 8.468425207 | 1.744225746 | 3.77753325 | 7.832892346 | 0.932322777 | 0.363671282 | 0.113153989 | 0.744455634 | 3.118789678 | 0.732747117 | 18.49773003 | 7.046723318 | 69.90980699 | 4.480542452 | 7.555608085 | 31.92272062 | 1.953933204 | 0.296436459 | 0.211403841 | 18.12539241 | 20.08700332 | 21.27365683 | 22.16970132 | 23.50165934 | 23.98600771 | 25.71968786 | 26.84337609 | 27.28897659 | 28.47078662 | 29.09075253 | 29.82696206 | 33.23 | 1.226160688 |
| 372 | 1 | 71 | 0 | 206 | 256 | 52 | 175 | 10.98871567 | 21.86635728 | 64.38273715 | 42.16934397 | 21.24360376 | 17.90076502 | 2.845493107 | 4.452154264 | 9.504652201 | 0.853202365 | 0.515958026 | 0.287901032 | 2.901349203 | 6.588346214 | 21.08580556 | 34.73840507 | 14.96239268 | 112.4990598 | 6.13746085 | 8.441564741 | 33.67440823 | 2.563863516 | 0.265875309 | 0.488094079 | 18.02852274 | 19.50578527 | 21.00726523 | 22.1454839 | 23.23526774 | 23.98600771 | 25.71968786 | 26.84337609 | 27.28897659 | 28.37391694 | 29.09075253 | 29.74946632 | 33.23 | 0.573184322 |
| 373 | 1 | 64 | 0 | 189 | 238 | 46 | 148 | 11.36614851 | 43.38993893 | 26.99508495 | 60.33695349 | 14.56271916 | 23.1864838 | 1.983579395 | 4.817871196 | 9.537058227 | 0.907889068 | 0.396053208 | 0.245495208 | 4.069047751 | 15.95468841 | 4.778289331 | 50.08628489 | 6.64E-05 | 111.3133756 | 4.039728452 | 6.408185201 | 29.04544266 | 2.209072845 | 0.06646441 | 0.387186704 | 17.95587048 | 20.08700332 | 21.00726523 | 22.04861423 | 23.23526774 | 23.9617903 | 25.71968786 | 26.84337609 | 27.28897659 | 28.47078662 | 29.09075253 | 29.71071845 | 33.23 | 0.574332125 |
| 374 | 1 | 50 | 1 | 43 | 157 | 53 | 93 | 0.408865345 | 2.404672065 | 4.345472904 | 6.031802609 | 7.369812468 | 9.856571576 | 0.628776306 | 1.167623382 | 2.794908501 | 0.281603375 | 0.57209638 | 0.041634219 | 0.217682058 | 1.13304355 | 1.787172707 | 11.78725709 | 6.64E-05 | 82.34002336 | 4.577931884 | 7.462388479 | 34.57451044 | 2.853928164 | 6.65E-05 | 0.489361164 | 17.88321823 | 20.20809041 | 21.15256974 | 22.16970132 | 23.23526774 | 23.9617903 | 25.71968786 | 26.84337609 | 27.28897659 | 28.35454301 | 28.89701319 | 29.67197058 | 33.23 | 1.51253796 |
| 383 | 1 | 51 | 0 | 427 | 211 | 39 | 119 | 75.88885668 | 92.45401142 | 52.81154743 | 52.60243052 | 30.26460191 | 15.95886289 | 5.689713556 | 5.78E-05 | 15.73812185 | 2.346379951 | 0.847918975 | 1.088963393 | 12.16249839 | 15.55904785 | 8.662593848 | 15.81900976 | 10.21091909 | 97.19481369 | 4.48156557 | 4.14E-05 | 30.66586702 | 3.43893246 | 5.37E-05 | 0.376693504 | 17.90743564 | 19.82061171 | 20.98304781 | 22.00017939 | 23.30791999 | 24.15552964 | 25.87467934 | 26.84337609 | 27.28897659 | 28.39329088 | 29.05200466 | 29.9625796 | 33.23 | 1.77530994 |
| 391 | 1 | 89 | 1 | 74 | 197 | 55 | 112 | 0.641243559 | 5.526988171 | 8.985008274 | 12.26614568 | 29.72855399 | 4.473094071 | 1.565886577 | 4.833929297 | 4.723021138 | 0.459942562 | 0.464463479 | 0.088193215 | 0.500431816 | 4.743218053 | 0.025607584 | 25.727878 | 24.20883268 | 78.82454939 | 5.506809948 | 18.38836683 | 28.82213846 | 1.970926544 | 0.173775805 | 0.462791186 | 17.88321823 | 20.30496009 | 21.29787425 | 22.04861423 | 23.62274643 | 23.9617903 | 25.71968786 | 26.80462822 | 27.28897659 | 28.39329088 | 28.95513499 | 29.65259665 | 33.23 | 1.980398378 |
| 442 | 1 | 79 | 1 | 130 | 211 | 55 | 124 | 3.827492797 | 23.23073869 | 11.34958295 | 44.29734179 | 14.40684749 | 18.08394689 | 1.900382059 | 6.721304528 | 6.044977809 | 0.810787655 | 0.496118354 | 0.143631 | 1.378562027 | 6.364653099 | 0.048845978 | 32.59089487 | 6.64E-05 | 106.4329883 | 6.007743269 | 19.40921477 | 28.43371789 | 2.1 | 0.338853936 | 0.413842147 | 17.88321823 | 20.30496009 | 20.98304781 | 22.12126648 | 23.23526774 | 24.03444255 | 25.71968786 | 26.76588035 | 27.28897659 | 28.37391694 | 29.03263073 | 29.74946632 | 33.23 | 0.948078948 |
| 453 | 1 | 70 | 1 | 68 | 184 | 63 | 97 | 0.635474915 | 5.467662358 | 5.07676571 | 13.60161688 | 8.083196453 | 19.9264281 | 1.840042296 | 8.356991371 | 5.888579795 | 0.678866738 | 0.45871772 | 0.066952797 | 0.151790243 | 1.52942458 | 1.785598803 | 18.82122413 | 1.367050292 | 88.35898868 | 5.013763509 | 28.95740664 | 30.90563865 | 2.718775036 | 7.37E-05 | 0.292392368 | 17.88321823 | 20.20809041 | 20.93461297 | 22.12126648 | 23.23526774 | 23.9617903 | 25.66156606 | 26.64963674 | 27.28897659 | 28.35454301 | 28.95513499 | 29.59447484 | 33.23 | 1.106104496 |
| 455 | 1 | 69 | 1 | 89 | 125 | 41 | 67 | 1.947351199 | 16.27476393 | 6.353792714 | 38.37924633 | 12.88624307 | 8.012038801 | 0.90138532 | 2.122397779 | 5.064699181 | 0.735963379 | 0.435111625 | 0.176372738 | 0.284512948 | 2.888643895 | 6.20E-05 | 16.83485131 | 3.83672288 | 56.52397873 | 2.899878883 | 5.513818099 | 31.26904193 | 2.493961147 | 0.181235886 | 0.245260955 | 18.00430532 | 20.20809041 | 20.93461297 | 22.04861423 | 23.23526774 | 23.9617903 | 25.71968786 | 26.80462822 | 27.28897659 | 28.37391694 | 28.91638712 | 29.82696206 | 33.23 | 0.54362187 |
| 461 | 1 | 82 | 1 | 57 | 206 | 71 | 118 | 0.286598041 | 1.542227979 | 2.431752015 | 19.54927499 | 4.957801053 | 17.30149507 | 1.506716497 | 4.008185304 | 2.269226462 | 0.358838907 | 0.486472282 | 0.067053628 | 4.32E-05 | 2.425534733 | 0.305386946 | 27.24965511 | 6.64E-05 | 97.61919183 | 8.819836454 | 34.69945704 | 25.90911921 | 2.209409572 | 0.524776645 | 0.352646319 | 17.88321823 | 20.20809041 | 20.93461297 | 22.29078841 | 23.23526774 | 23.9617903 | 25.64219212 | 26.76588035 | 27.28897659 | 28.33516907 | 28.91638712 | 29.71071845 | 33.23 | 1.301212364 |
| 466 | 1 | 75 | 1 | 194 | 200 | 69 | 96 | 0.9360633 | 26.00477056 | 21.38542581 | 46.02502673 | 26.77812104 | 42.69783895 | 3.156910909 | 24.96535202 | 19.55336946 | 1.318491486 | 0.696758254 | 0.179965575 | 0.137759758 | 3.30535906 | 3.082137511 | 20.10639651 | 10.00713524 | 81.13493503 | 5.793410217 | 25.31520863 | 38.08120469 | 2.349110225 | 0.720723472 | 0.235893273 | 17.88321823 | 20.20809041 | 20.93461297 | 22.04861423 | 23.23526774 | 23.9617903 | 25.66156606 | 26.70775854 | 27.28897659 | 28.33516907 | 29.01325679 | 29.61384878 | 33.23 | 1.235830188 |
| 480 | 1 | 88 | 1 | 294 | 199 | 50 | 107 | 8.619119081 | 76.28249554 | 65.79510877 | 99.99995521 | 18.16678528 | 11.45788845 | 2.872958427 | 3.045908575 | 11.1415875 | 0.966291378 | 0.785957162 | 0.288993673 | 1.44637783 | 11.27201861 | 5.721735207 | 26.1812825 | 12.91634172 | 93.65720456 | 4.612843597 | 3.546479613 | 38.4000753 | 2.873965577 | 0.316970584 | 0.336113548 | 17.88321823 | 20.20809041 | 21.07991748 | 22.04861423 | 23.59852902 | 24.20396448 | 25.71968786 | 26.84337609 | 27.28897659 | 28.37391694 | 28.97450892 | 29.86570993 | 33.23 | 1.29371141 |
| 1001 | 0 | 49 | 0 | 88 | 195 | 75 | 96 | 6.64E-05 | 6.472422655 | 26.39091044 | 5.370834812 | 18.32120968 | 16.24564197 | 0.918992137 | 8.959385627 | 7.960775812 | 0.182376344 | 0.781284395 | 0.056343979 | 0.434773071 | 0.630489293 | 6.151854974 | 11.73296068 | 6.64E-05 | 92.40875456 | 2.963751692 | 22.73616187 | 45.15875602 | 0.122669168 | 1.30253597 | 0.38758286 | 17.40234375 | 19.12109375 | 20.91796875 | 21.71875 | 22.55859375 | 23.6328125 | 25.325 | 26.54375 | 26.934375 | 27.903125 | 28.60625 | 29.73125 | 33.05 | 0.913564777 |
| 1002 | 0 | 44 | 0 | 50 | 169 | 85 | 67 | 0.098744629 | 1.885920907 | 7.342694592 | 2.06445862 | 6.876474145 | 11.87398638 | 0.159573775 | 6.476863145 | 3.439980213 | 5.78E-05 | 0.699138757 | 0.018114217 | 0.019919492 | 0.205121882 | 1.413662208 | 3.781883361 | 6.64E-05 | 72.89026793 | 0.254714993 | 43.79451368 | 39.27545552 | 5.78E-05 | 1.192360731 | 0.353913155 | 17.68947888 | 18.7550453 | 20.88617813 | 21.70957037 | 22.72670195 | 23.5743116 | 25.33220917 | 26.49464526 | 26.9596197 | 27.71520316 | 28.6838999 | 29.78821419 | 33.05 | 2.614971992 |
| 1003 | 0 | 43 | 0 | 39 | 205 | 79 | 112 | 0.02356598 | 0.646390201 | 6.14691394 | 3.478444605 | 5.106473219 | 11.51642152 | 0.03737073 | 3.759722991 | 2.847177018 | 0.254482218 | 0.604685064 | 5.78E-05 | 0.02778351 | 0.149171971 | 2.393790694 | 12.47457703 | 6.64E-05 | 103.4268636 | 0.243655667 | 34.28022991 | 40.25531367 | 0.633902974 | 1.572881653 | 0.240956564 | 17.68947888 | 19.21875 | 20.72265625 | 21.70957037 | 22.72670195 | 23.5743116 | 25.33220917 | 26.49464526 | 26.9596197 | 27.71520316 | 28.621875 | 29.78821419 | 33.05 | 2.436741835 |
| 1004 | 0 | 48 | 0 | 34 | 160 | 66 | 90 | 0.079965747 | 3.320681599 | 8.41381268 | 2.431587284 | 4.622338514 | 8.301269145 | 6.60E-05 | 2.686753469 | 2.345052101 | 0.217338459 | 0.542280911 | 0.092525311 | 0.033011605 | 0.332188275 | 2.058925783 | 5.665144576 | 6.64E-05 | 87.65118366 | 5.78E-05 | 24.1252641 | 40.65186657 | 5.78E-05 | 1 | 0.526402706 | 17.75390625 | 19.82421875 | 21.09375 | 21.9140625 | 22.79296875 | 23.671875 | 25.33220917 | 26.49464526 | 26.9596197 | 27.71520316 | 28.590625 | 29.60625 | 33.05 | 1.345229097 |
| 1005 | 0 | 42 | 0 | 276 | 182 | 53 | 96 | 20.64443098 | 55.45512615 | 33.88235919 | 64.47829143 | 10.55162199 | 15.05368805 | 2.536374032 | 8.122715151 | 7.983656187 | 0.684964565 | 1.941502385 | 0.489046045 | 3.148661579 | 8.149864241 | 3.124647244 | 19.69815289 | 6.64E-05 | 83.92330667 | 3.538637061 | 10.63863251 | 33.48325713 | 2.359872177 | 1.575891872 | 0.40288467 | 17.75390625 | 19.7265625 | 20.68359375 | 21.66015625 | 22.7734375 | 23.7109375 | 25.246875 | 26.66875 | 27.0125 | 27.684375 | 28.3875 | 29.54375 | 33.05 | 0.763301886 |
| 1006 | 0 | 60 | 0 | 186 | 261 | 77 | 163 | 12.40305205 | 21.86142151 | 21.60140954 | 28.97471888 | 12.4775074 | 13.72209826 | 0.984351152 | 6.142089119 | 6.193362938 | 1.330044785 | 0.85 | 0.242312963 | 1.717333695 | 3.570287562 | 6.20E-05 | 40.15828051 | 6.64E-05 | 130.177253 | 3.332528548 | 19.79918719 | 47.8657471 | 0.185297247 | 1.5 | 0.582042316 | 17.75390625 | 19.82421875 | 20.9765625 | 21.9140625 | 22.96875 | 23.57421875 | 25.04375 | 26.49464526 | 26.9596197 | 27.71520316 | 28.496875 | 29.60625 | 33.05 | 1.290007932 |
| 1007 | 0 | 43 | 0 | 31 | 223 | 85 | 125 | 1.166960834 | 1.803072547 | 2.182663203 | 2.317959356 | 6.847346191 | 10.16693143 | 0.555357134 | 3.248075564 | 2.018256426 | 5.78E-05 | 0.769373317 | 5.57E-05 | 0.131420607 | 0.495340168 | 3.636444865 | 10.31540369 | 6.64E-05 | 115.8887578 | 3.437598556 | 37.11910073 | 40.62322121 | 5.78E-05 | 1.52065101 | 0.569514838 | 17.85900081 | 19.82421875 | 21.09375 | 21.9140625 | 22.79296875 | 23.671875 | 24.92535653 | 26.49464526 | 26.9596197 | 27.71520316 | 28.590625 | 29.60625 | 33.05 | 0.232944749 |
| 1008 | 0 | 59 | 0 | 75 | 201 | 78 | 111 | 2.606796523 | 5.558683427 | 9.973141563 | 9.994042458 | 18.76068921 | 12.65541761 | 0.594065288 | 7.38391217 | 4.895749985 | 0.620472625 | 0.915426205 | 0.306988104 | 0.209633446 | 0.919113095 | 4.84918703 | 8.854062695 | 7.647856921 | 99.10087955 | 1.926422558 | 32.00206937 | 40.05962247 | 0.000128261 | 1.440515461 | 0.518734927 | 17.75390625 | 19.82421875 | 21.09375 | 21.9140625 | 22.79296875 | 23.671875 | 25.00285227 | 26.49464526 | 26.9596197 | 27.71520316 | 28.590625 | 29.73009238 | 33.05 | 0.5410032 |
| 1009 | 0 | 50 | 0 | 44 | 238 | 100 | 122 | 0.136985889 | 0.746765526 | 3.331979808 | 3.458640249 | 10.89318057 | 19.14951269 | 0.63122398 | 6.497559964 | 3.401469864 | 5.78E-05 | 0.888210105 | 0.028943716 | 0.058451233 | 0.377121497 | 3.625919529 | 11.66180634 | 12.66371578 | 103.2005064 | 2.193043805 | 51.38238789 | 43.61179022 | 5.78E-05 | 1.436114648 | 0.569437398 | 17.75390625 | 19.82421875 | 21.09375 | 21.77734375 | 22.890625 | 23.671875 | 24.996875 | 26.49464526 | 26.9596197 | 27.71520316 | 28.590625 | 29.60625 | 33.05 | 0.53991782 |
| 1010 | 0 | 51 | 0 | 111 | 170 | 74 | 85 | 2.699911483 | 20.79465635 | 19.97818992 | 33.22104509 | 12.54808073 | 9.965265042 | 0.45898658 | 8.707674474 | 6.852353601 | 5.78E-05 | 1.543846643 | 0.141416714 | 0.449884849 | 3.233355313 | 2.893020011 | 15.10750742 | 6.64E-05 | 70.60971769 | 0.948759277 | 22.51618295 | 44.81693917 | 5.78E-05 | 2.156820826 | 0.345725412 | 17.75390625 | 19.82421875 | 20.80078125 | 21.77734375 | 22.890625 | 23.671875 | 24.996875 | 26.49464526 | 26.9596197 | 27.71520316 | 28.3875 | 29.60625 | 33.05 | 0.798331599 |
| 1011 | 0 | 53 | 0 | 53 | 192 | 54 | 123 | 0.3278862 | 4.5850522 | 11.0293290 | 3.8584812 | 10.5074042 | 11.0851578 | 0.8737118 | 7.0215274 | 0.0000578 | 0.2751319 | 0.6010984 | 0.0150763 | 0.1072824 | 0.7998532 | 4.8375558 | 12.6941529 | 0.0000664 | 115.8637952 | 4.0357160 | 43.9284147 | 4.3760065 | 1.4383390 | 0.9208388 | 0.3233912 | 17.75390625 | 19.82421875 | 21.09375 | 21.77734375 | 22.890625 | 23.671875 | 24.996875 | 26.49464526 | 26.9596197 | 27.71520316 | 28.590625 | 29.60625 | 33.05 | 1.526654133 |
| 1012 | 0 | 49 | 0 | 98 | 220 | 75 | 131 | 2.388450274 | 14.73570657 | 16.9254361 | 17.32133293 | 16.36391815 | 8.918034272 | 0.721688663 | 3.364088918 | 5.930166227 | 0.205671855 | 1.066675957 | 0.059324824 | 0.351303881 | 2.374841331 | 3.345161302 | 14.873933 | 0.791129721 | 117.9536108 | 2.641775698 | 13.12214454 | 51.59375681 | 2.019329339 | 1.520343108 | 0.376624959 | 17.75390625 | 19.82421875 | 20.87890625 | 21.77734375 | 22.890625 | 23.671875 | 25.23125 | 26.49464526 | 26.9596197 | 27.71520316 | 28.590625 | 29.60625 | 33.05 | 1.965314488 |
| 1013 | 0 | 43 | 0 | 69 | 165 | 64 | 88 | 0.191828724 | 5.097119968 | 19.18270939 | 5.338386419 | 16.34307304 | 11.748869 | 0.598385897 | 3.046311068 | 5.002589459 | 0.765732198 | 0.780143477 | 0.009638909 | 0.042338769 | 0.611041021 | 4.087660999 | 6.23460229 | 7.593687262 | 71.86121807 | 2.713087494 | 12.19073724 | 38.50307929 | 4 | 0.8515306 | 0.414943836 | 17.75390625 | 19.82421875 | 21.09375 | 21.77734375 | 22.890625 | 23.671875 | 25.2 | 26.49464526 | 26.9596197 | 27.71520316 | 28.590625 | 29.60625 | 33.05 | 1.919004944 |
| 1014 | 0 | 48 | 0 | 67 | 226 | 73 | 138 | 2.232759343 | 3.558256804 | 7.243785926 | 7.269916059 | 14.79952536 | 10.95600276 | 0.471559079 | 5.466592156 | 4.216581143 | 5.78E-05 | 1.216349593 | 0.151376551 | 0.255994396 | 0.675276246 | 4.219953575 | 11.05931084 | 7.785837304 | 119.3201282 | 2.190149629 | 29.90683288 | 38.82004618 | 1.040874504 | 1.794590683 | 0.536969827 | 17.75390625 | 19.82421875 | 21.09375 | 21.77734375 | 22.79296875 | 23.671875 | 24.996875 | 26.49464526 | 26.9596197 | 27.71520316 | 28.48125 | 29.60625 | 33.05 | 0.664772443 |
| 1015 | 0 | 44 | 0 | 54 | 173 | 80 | 77 | 1.127728809 | 1.565466341 | 5.364794958 | 4.027495477 | 16.33211991 | 9.493461399 | 5.78E-05 | 13.47631968 | 3.064351543 | 0.194133506 | 0.929041198 | 0.012145625 | 0.154711782 | 0.336959601 | 2.417915979 | 3.474182258 | 13.35764864 | 64.53080872 | 5.78E-05 | 51.81372977 | 28.38806569 | 5.78E-05 | 1.672688502 | 0.241467956 | 17.75390625 | 19.82421875 | 21.09375 | 21.77734375 | 22.890625 | 23.671875 | 24.996875 | 26.49464526 | 26.9596197 | 27.71520316 | 28.590625 | 29.60625 | 33.05 | 0.328776668 |
| 1016 | 0 | 47 | 0 | 83 | 194 | 67 | 117 | 2.508396825 | 4.472547569 | 13.51283104 | 7.403688777 | 16.67526404 | 14.99643932 | 0.96741548 | 4.086839929 | 6.335259909 | 5.78E-05 | 0.999815597 | 0.181753781 | 0.305902517 | 0.755168489 | 3.627348493 | 9.586305534 | 10.71055162 | 92.10908497 | 3.513572759 | 14.48727383 | 43.58338405 | 0.300427487 | 1.549929453 | 0.418985086 | 17.421875 | 19.58984375 | 21.09375 | 21.77734375 | 22.890625 | 23.671875 | 25.090625 | 26.49464526 | 26.9596197 | 27.71520316 | 28.48125 | 29.60625 | 33.05 | 0.530027867 |
| 1017 | 0 | 43 | 0 | 293 | 198 | 49 | 123 | 19.10148868 | 47.08089918 | 59.82405975 | 33.03472882 | 19.97914924 | 9.784595096 | 3.551615413 | 0.485660529 | 11.07327355 | 0.557217874 | 1.336089638 | 0.414082133 | 1.978868292 | 6.026045385 | 9.710598113 | 13.81514702 | 3.700425765 | 100.0426105 | 3.936 | 4.14E-05 | 45.59297128 | 5.78E-05 | 1.540187338 | 0.214261802 | 17.75390625 | 19.66796875 | 20.83984375 | 21.77734375 | 22.890625 | 23.671875 | 25.246875 | 26.49464526 | 26.9596197 | 27.71520316 | 28.559375 | 29.60625 | 33.05 | 0.440240466 |
| 1018 | 0 | 43 | 0 | 62 | 223 | 92 | 108 | 0.83192979 | 2.469439501 | 10.33876779 | 7.449930844 | 14.74153261 | 11.3826582 | 0.57686673 | 7.315842062 | 3.734842254 | 5.78E-05 | 0.749271714 | 0.059949444 | 0.092013511 | 0.642990533 | 2.606922528 | 13.98095268 | 0.330488053 | 102.3366734 | 2.369728783 | 47.31640386 | 36.97414817 | 5.78E-05 | 1.345152146 | 0.48422371 | 17.75390625 | 19.82421875 | 21.09375 | 21.640625 | 22.79296875 | 23.61328125 | 24.996875 | 26.49464526 | 26.9596197 | 27.71520316 | 28.590625 | 29.60625 | 33.05 | 1.640125402 |
| 1019 | 0 | 57 | 0 | 42 | 199 | 77 | 105 | 0.81808516 | 1.98408453 | 4.599826302 | 2.336193048 | 6.461301425 | 10.91249289 | 0.565291882 | 2.923887691 | 2.67619499 | 0.223266334 | 0.618225202 | 0.047253569 | 0.345625712 | 0.561421816 | 4.302421705 | 5.135958233 | 21.41608475 | 80.79692527 | 2.797286387 | 24.46694997 | 41.40327518 | 3.582072696 | 1.33316031 | 0.414712501 | 17.75390625 | 19.82421875 | 21.09375 | 21.77734375 | 22.890625 | 23.671875 | 24.996875 | 26.49464526 | 26.9596197 | 27.71520316 | 28.590625 | 29.60625 | 33.05 | 1.302385577 |
| 1020 | 0 | 55 | 0 | 165 | 244 | 64 | 142 | 3.538899574 | 15.61386887 | 45.83301644 | 20.85604453 | 40.58268738 | 19.70637562 | 1.46875026 | 11.08891458 | 8.514639279 | 0.691306538 | 1.008121781 | 0.190896934 | 0.528538073 | 2.847031908 | 12.60750954 | 13.28467867 | 27.05491256 | 110.9740752 | 3.788822305 | 24.50283836 | 32.61429545 | 2.740830307 | 1.377928968 | 0.396315311 | 17.75390625 | 19.91748139 | 21.09375 | 21.77734375 | 22.890625 | 23.671875 | 24.996875 | 26.49464526 | 26.9596197 | 27.71520316 | 28.590625 | 29.60625 | 33.05 | 1.361841462 |
| 1021 | 0 | 47 | 0 | 105 | 252 | 63 | 168 | 1.909550745 | 15.19587733 | 14.27842763 | 30.39946695 | 17.71619195 | 11.08653922 | 0.791169018 | 4.702986576 | 7.648840891 | 0.455107829 | 0.84569981 | 0.111867395 | 0.359285177 | 3.698213186 | 6.20E-05 | 39.45209446 | 31.45337119 | 110.956747 | 3.821510474 | 13.34086436 | 41.93040197 | 2.5621902 | 1.274954084 | 0.401429601 | 17.75390625 | 20.01953125 | 21.09375 | 21.71875 | 23.06640625 | 23.671875 | 24.996875 | 26.49464526 | 26.9596197 | 27.71520316 | 28.590625 | 29.60625 | 33.05 | 1.143123771 |
| 1022 | 0 | 42 | 0 | 151 | 218 | 66 | 131 | 9.595642425 | 16.25702321 | 44.24177988 | 20.35579203 | 18.71892278 | 9.531283276 | 1.202986394 | 5.814857754 | 5.209520435 | 0.557977636 | 1.181291828 | 0.345234345 | 1.611251897 | 3.27243866 | 11.33025056 | 19.27457376 | 0.478291119 | 106.1429566 | 2.946168527 | 21.44608152 | 35.23028567 | 3.822602211 | 1.424185182 | 0.543516757 | 17.75390625 | 19.3359375 | 20.703125 | 21.77734375 | 22.96875 | 23.671875 | 24.996875 | 26.49464526 | 26.9596197 | 27.71520316 | 28.5125 | 29.60625 | 33.05 | 0.612048479 |
| 1023 | 0 | 45 | 0 | 178 | 207 | 47 | 139 | 18.34754582 | 33.28547686 | 38.97131373 | 24.76323121 | 15.9819739 | 11.93765223 | 2.390297512 | 6.016207971 | 9.530761883 | 1.706774224 | 1.316611462 | 0.354179759 | 4.028305942 | 7.706304821 | 8.14905197 | 17.81587829 | 2.764599383 | 107.2564854 | 4.794440102 | 8.296716341 | 29.73839109 | 5.060132906 | 1.299548037 | 0.283996675 | 17.75390625 | 19.55078125 | 20.76171875 | 21.6015625 | 22.890625 | 23.57421875 | 24.996875 | 26.49464526 | 26.9596197 | 27.71520316 | 28.590625 | 29.60625 | 33.05 | 0.945161988 |
| 1024 | 0 | 41 | 0 | 481 | 256 | 58 | 132 | 61.52870323 | 67.42851361 | 54.5380821 | 71.86130317 | 27.05310246 | 20.46205173 | 2.747985613 | 5.428277339 | 16.32322725 | 2.27930061 | 1.77436746 | 1.110068749 | 9.014561929 | 12.11303227 | 7.938392362 | 31.61745478 | 6.64E-05 | 119.3966407 | 2.770926426 | 0.32313975 | 43.95487805 | 5.506541681 | 1.504396503 | 0.604317926 | 17.6953125 | 19.6875 | 20.7421875 | 21.6796875 | 22.98828125 | 23.75 | 24.996875 | 26.49464526 | 26.9596197 | 27.71520316 | 28.590625 | 29.60625 | 33.05 | 1.245127952 |
| 1025 | 0 | 64 | 0 | 157 | 223 | 67 | 131 | 4.923319412 | 18.33395569 | 42.71115978 | 15.83717735 | 32.1779252 | 16.24490344 | 1.341965501 | 11.60641688 | 8.610052805 | 0.9044514 | 1.132411057 | 0.208577062 | 0.91084299 | 3.530127568 | 11.32886652 | 14.14359249 | 16.68902995 | 101.3001656 | 3.188371592 | 23.83674127 | 32.74862655 | 3.301161073 | 1.334860606 | 0.390282103 | 17.75390625 | 19.7265625 | 21.09375 | 21.77734375 | 22.890625 | 23.671875 | 24.996875 | 26.49464526 | 26.9596197 | 27.71520316 | 28.590625 | 29.60625 | 33.05 | 0.46467642 |
| 1026 | 0 | 61 | 0 | 98 | 189 | 62 | 107 | 3.322535313 | 7.773824583 | 22.37675843 | 7.619996762 | 14.83532516 | 7.92285895 | 0.62669015 | 7.726315634 | 3.458099157 | 0.496386996 | 0.79806065 | 0.13707512 | 0.579248772 | 1.809297812 | 7.155068642 | 8.785491493 | 6.64E-05 | 95.71227635 | 2.269845104 | 32.34506407 | 21.52958527 | 3.058005055 | 1.160516701 | 0.379120189 | 17.75390625 | 19.82421875 | 21.09375 | 21.77734375 | 22.96875 | 23.515625 | 24.996875 | 26.49464526 | 26.9596197 | 27.71520316 | 28.5125 | 29.60625 | 33.05 | 0.60765084 |
| 1027 | 0 | 38 | 0 | 95 | 232 | 135 | 63 | 5.341996784 | 12.73945662 | 22.43532803 | 9.430049381 | 14.56894362 | 15.5725983 | 1.095961265 | 7.047128521 | 2.009161657 | 0.230454766 | 1.143423544 | 0.163530119 | 0.646212481 | 1.540842624 | 3.466797126 | 5.829851075 | 1.319106228 | 62.10098446 | 3.34866845 | 82.25874812 | 40.89840478 | 3.775565083 | 2.049527056 | 0.510880803 | 17.8515625 | 19.6875 | 20.91796875 | 21.77734375 | 22.890625 | 23.671875 | 24.996875 | 26.49464526 | 26.9596197 | 27.71520316 | 28.590625 | 29.60625 | 33.02 | 0.474640814 |
| 1028 | 0 | 52 | 0 | 78 | 209 | 75 | 124 | 4.050068884 | 6.151378109 | 10.96352286 | 7.755653626 | 14.55449241 | 10.42731564 | 0.686810314 | 9.057786083 | 4.737115613 | 0.356886909 | 0.866616597 | 0.176503579 | 0.580410021 | 1.490838656 | 5.484395444 | 14.65060499 | 6.64E-05 | 105.9790834 | 2.418294901 | 37.10404395 | 31.4364756 | 2.194989536 | 1.406200933 | 0.429584479 | 17.75390625 | 19.82421875 | 21.09375 | 21.77734375 | 22.96875 | 23.59375 | 24.996875 | 26.49464526 | 26.9596197 | 27.71520316 | 28.5125 | 29.60625 | 33.05 | 0.547280318 |
| 1029 | 0 | 41 | 0 | 72 | 171 | 68 | 89 | 3.73776564 | 3.475767926 | 8.465348321 | 6.561948504 | 12.56569879 | 10.09982657 | 0.668084439 | 7.314784825 | 4.565745676 | 0.365844005 | 0.832065006 | 0.165070807 | 0.558464013 | 0.912766719 | 4.317512558 | 10.46010188 | 6.64E-05 | 75.05507274 | 1.975801695 | 31.108969 | 31.69602128 | 2.48939113 | 1.341219166 | 0.410701257 | 17.65625 | 19.70703125 | 21.03515625 | 21.81640625 | 22.94921875 | 23.5546875 | 24.996875 | 26.49464526 | 26.9596197 | 27.71520316 | 28.5125 | 29.60625 | 33.02 | 0.733686443 |
| 1030 | 0 | 50 | 0 | 34 | 208 | 106 | 83 | 0.173674391 | 2.635794008 | 4.802248105 | 1.786290032 | 5.588794401 | 10.18803403 | 0.539496305 | 4.304690996 | 1.481217844 | 0.182700115 | 0.743514042 | 0.036582129 | 0.059504494 | 0.571521397 | 3.169755868 | 6.556676358 | 9.938563163 | 69.08658186 | 2.930946982 | 62.26076234 | 32.65328282 | 4.416788506 | 1.657341594 | 0.470307915 | 17.75390625 | 19.82421875 | 21.09375 | 21.77734375 | 22.890625 | 23.671875 | 24.996875 | 26.49464526 | 26.9596197 | 27.71520316 | 28.590625 | 29.60625 | 33.05 | 1.880924893 |
| 1031 | 0 | 55 | 0 | 67 | 181 | 59 | 100 | 2.054242614 | 4.695726776 | 13.46094769 | 6.309586012 | 14.21897753 | 11.5756911 | 0.67084287 | 6.777322139 | 6.424066435 | 0.44139262 | 0.87192933 | 0.151383078 | 0.325008781 | 1.152788969 | 5.037386041 | 11.17268018 | 1.917684391 | 84.15891547 | 2.365414013 | 22.84206227 | 38.24549503 | 2.822683054 | 1.387517785 | 0.441972603 | 17.75390625 | 19.82421875 | 21.09375 | 21.77734375 | 22.94921875 | 23.57421875 | 24.996875 | 26.49464526 | 26.9596197 | 27.71520316 | 28.5125 | 29.60625 | 33.05 | 0.344362479 |
| 1032 | 0 | 43 | 0 | 72 | 189 | 72 | 98 | 1.065122238 | 3.81829595 | 14.01879243 | 8.866775585 | 23.87799439 | 8.662354748 | 0.761520968 | 11.75942802 | 6.240499409 | 0.474751877 | 0.936576694 | 0.181958066 | 0.206254155 | 0.925011081 | 7.838992988 | 12.32406581 | 10.87587214 | 71.8545799 | 2.405282007 | 35.18740872 | 29.64546049 | 2.506150349 | 1.364044895 | 0.408007706 | 17.75390625 | 19.82421875 | 21.09375 | 21.77734375 | 23.02734375 | 23.57421875 | 24.996875 | 26.49464526 | 26.9596197 | 27.71520316 | 28.48125 | 29.60625 | 33.05 | 0.573063199 |
| 1033 | 0 | 48 | 0 | 297 | 183 | 45 | 113 | 23.98712902 | 51.75955939 | 42.2890327 | 34.45613146 | 17.36238357 | 12.53606346 | 2.062691094 | 4.84793516 | 11.98784948 | 1.224297752 | 1.370524817 | 0.794101173 | 3.376340624 | 8.127614221 | 7.33280981 | 13.89967928 | 6.64E-05 | 99.04245058 | 3.117847729 | 2.765134396 | 35.89282669 | 3.366352639 | 1.391117572 | 0.444405607 | 17.71484375 | 19.6484375 | 20.72265625 | 21.6796875 | 23.06640625 | 23.65234375 | 24.91875 | 26.49464526 | 26.9596197 | 27.71520316 | 28.48125 | 29.60625 | 33.05 | 0.746850244 |
| 1034 | 0 | 47 | 0 | 112 | 200 | 56 | 125 | 4.06555892 | 16.36773604 | 23.57666589 | 15.03430212 | 15.54328662 | 7.901564466 | 0.76233707 | 4.388734832 | 7.208745792 | 0.729913582 | 0.902669645 | 0.239989747 | 0.975789665 | 4.420513587 | 8.602488227 | 20.90244099 | 6.64E-05 | 107.3670455 | 2.241816239 | 10.39732264 | 37.72043595 | 4.153187798 | 1.151815818 | 0.537909613 | 17.44140625 | 19.8046875 | 20.91796875 | 21.77734375 | 23.125 | 23.5546875 | 24.996875 | 26.49464526 | 26.9596197 | 27.71520316 | 28.48125 | 29.60625 | 33.05 | 1.141250215 |
| 1035 | 0 | 52 | 0 | 138 | 209 | 64 | 120 | 5.951077207 | 24.07242066 | 29.97123338 | 22.66874875 | 23.29386316 | 7.20355416 | 1.051794707 | 8.661660914 | 8.318581556 | 0.747948189 | 0.98836075 | 0.300839055 | 1.015881996 | 5.883619963 | 10.62400108 | 19.3315094 | 8.592300602 | 88.41610012 | 2.953235197 | 20.84519332 | 34.19064062 | 3.149976639 | 1.350346891 | 0.445359919 | 17.4609375 | 19.921875 | 20.87890625 | 21.77734375 | 23.0078125 | 23.59375 | 24.996875 | 26.49464526 | 26.9596197 | 27.71520316 | 28.48125 | 29.60625 | 33.05 | 0.939133959 |
| 1036 | 0 | 51 | 0 | 46 | 193 | 80 | 94 | 0.393172001 | 1.541841858 | 5.62052614 | 2.463975128 | 9.063746485 | 11.09782045 | 0.531907021 | 6.962513205 | 4.136672014 | 0.438307228 | 0.689378659 | 0.092963309 | 0.10083792 | 0.213839674 | 3.860748591 | 6.650848752 | 10.5955916 | 78.03481655 | 2.093494687 | 38.87287932 | 33.19879192 | 3.895035569 | 1.471067338 | 0.458540603 | 17.75390625 | 19.82421875 | 21.09375 | 21.77734375 | 23.02734375 | 23.57421875 | 24.996875 | 26.49464526 | 26.9596197 | 27.71520316 | 28.48125 | 29.60625 | 33.05 | 2.025785961 |
| 1037 | 0 | 41 | 0 | 91 | 188 | 51 | 122 | 2.812042459 | 14.30664509 | 31.97581025 | 11.07016077 | 12.17048768 | 7.127312829 | 0.761793432 | 1.983007372 | 4.352818951 | 0.307333661 | 1.025583519 | 0.321781809 | 0.520075862 | 2.857597986 | 8.106034891 | 16.04208817 | 6.64E-05 | 100.3998203 | 3.137995689 | 6.670913016 | 34.9140615 | 3.460454952 | 1.225993486 | 0.469039024 | 17.75390625 | 19.82421875 | 20.91796875 | 21.77734375 | 23.02734375 | 23.57421875 | 24.996875 | 26.49464526 | 26.9596197 | 27.71520316 | 28.48125 | 29.60625 | 33.05 | 0.328411291 |
| 1038 | 0 | 52 | 0 | 106 | 207 | 102 | 86 | 7.062308297 | 14.76238245 | 14.96901758 | 10.60946566 | 13.07661133 | 11.41228035 | 1.349800164 | 8.927843752 | 2.071560311 | 0.332440885 | 0.950097391 | 0.268088619 | 0.957855532 | 2.56753538 | 4.031919279 | 10.67822334 | 6.64E-05 | 73.95310134 | 3.455279268 | 63.93476409 | 23.21704629 | 3.012858286 | 1.412745659 | 0.583149496 | 17.75390625 | 19.82421875 | 20.87890625 | 21.77734375 | 23.02734375 | 23.57421875 | 24.91875 | 26.49464526 | 26.9596197 | 27.71520316 | 28.48125 | 29.60625 | 33.05 | 0.630341482 |
| 1039 | 0 | 63 | 0 | 265 | 215 | 64 | 114 | 17.77972094 | 55.09165714 | 38.78069143 | 45.3624764 | 37.74007198 | 21.37359529 | 2.214949276 | 17.51433599 | 10.37877834 | 1.212930727 | 1.504636453 | 0.697661425 | 2.229317912 | 7.723021458 | 4.241327503 | 26.33050668 | 6.64E-05 | 104.044416 | 2.609449562 | 22.07488231 | 32.99496718 | 2.679156115 | 1.399836606 | 0.515081782 | 17.75390625 | 19.82421875 | 20.78125 | 21.85546875 | 23.02734375 | 23.671875 | 24.996875 | 26.49464526 | 26.9596197 | 27.71520316 | 28.48125 | 29.60625 | 33.08 | 0.512543494 |
| 1040 | 0 | 42 | 0 | 38 | 175 | 70 | 95 | 0.201650247 | 3.061851642 | 8.176753912 | 3.873693853 | 7.190635183 | 9.42675826 | 0.540110762 | 2.550392693 | 2.476722091 | 0.212166497 | 0.771651504 | 0.065452426 | 0.081091345 | 0.725323087 | 3.462034743 | 10.93737344 | 6.64E-05 | 81.91107775 | 2.515310139 | 21.84094448 | 37.56137959 | 4.046272968 | 1.563273989 | 0.469215698 | 17.75390625 | 19.89326397 | 21.03148264 | 21.77734375 | 23.02734375 | 23.57421875 | 24.996875 | 26.49464526 | 26.9596197 | 27.71520316 | 28.48125 | 29.60625 | 33.05 | 0.87253192 |
| 1041 | 0 | 41 | 0 | 72 | 199 | 55 | 129 | 1.108923537 | 10.61236091 | 16.56868312 | 10.06005423 | 13.11655087 | 7.929573817 | 0.76305257 | 3.974957108 | 4.055666029 | 0.385715341 | 0.963649853 | 0.146367281 | 0.372225136 | 2.958679255 | 7.710022941 | 18.96800071 | 6.64E-05 | 104.8604209 | 3.348891138 | 15.33154691 | 30.93120911 | 3.823348995 | 1.360875669 | 0.47280184 | 17.75390625 | 19.94169881 | 20.95883039 | 21.77734375 | 23.02734375 | 23.57421875 | 24.996875 | 26.49464526 | 26.9596197 | 27.71520316 | 28.48125 | 29.60625 | 33.05 | 1.401095573 |
| 1042 | 0 | 46 | 0 | 92 | 253 | 77 | 153 | 1.036434858 | 4.890013099 | 23.18141015 | 12.61707234 | 20.36340646 | 9.348991012 | 0.847188004 | 6.62119438 | 4.794190024 | 0.497504 | 0.880361527 | 0.116530227 | 0.348400263 | 2.184230553 | 14.43569509 | 25.09931325 | 6.64E-05 | 121.9518066 | 4.082402855 | 30.9439037 | 35.36894867 | 4.175910593 | 1.602254618 | 0.551278478 | 17.75390625 | 19.82421875 | 21.09375 | 21.875 | 23.02734375 | 23.57421875 | 24.996875 | 26.49464526 | 26.9596197 | 27.71520316 | 28.48125 | 29.60625 | 33.08 | 1.871806647 |
| 1043 | 0 | 48 | 0 | 112 | 211 | 53 | 150 | 4.916641804 | 20.95931305 | 23.73211715 | 15.71093967 | 14.61108099 | 9.139961861 | 0.891457354 | 0.409990365 | 10.06863769 | 0.862046642 | 1.0174709 | 0.2698809 | 1.442252951 | 6.029228055 | 10.37900444 | 19.04171923 | 6.64E-05 | 124.804592 | 2.71548488 | 4.14E-05 | 44.32829325 | 4.942933731 | 1.453997507 | 0.476389359 | 17.75390625 | 19.82421875 | 20.87890625 | 21.77734375 | 23.02734375 | 23.57421875 | 24.996875 | 26.49464526 | 26.9596197 | 27.71520316 | 28.48125 | 29.60625 | 33.08 | 2.018311953 |
| 1044 | 0 | 40 | 0 | 202 | 161 | 57 | 88 | 8.62012025 | 25.26686483 | 24.80254323 | 30.32861361 | 18.75383225 | 8.632190766 | 1.286714001 | 5.696308581 | 6.845838665 | 0.631525355 | 1.141247554 | 0.381197726 | 1.513365843 | 5.273205378 | 4.881803836 | 15.85396539 | 6.64E-05 | 69.62278403 | 2.494134868 | 14.50605796 | 32.71683906 | 2.972660512 | 1.251651109 | 0.407040682 | 17.67578125 | 19.84375 | 20.76509104 | 21.77734375 | 23.02734375 | 23.57421875 | 24.996875 | 26.49464526 | 26.9596197 | 27.71520316 | 28.48125 | 29.60625 | 33.08 | 0.654420455 |
| 1045 | 0 | 46 | 0 | 36 | 210 | 85 | 116 | 0.349432739 | 1.248638499 | 3.325152953 | 3.127148887 | 8.188115454 | 11.84734654 | 0.544055081 | 5.372523013 | 3.697206935 | 0.198509242 | 0.879317004 | 0.291869545 | 0.086935185 | 0.311356964 | 3.941729326 | 11.89594387 | 6.64E-05 | 99.00852771 | 2.895064649 | 34.03721936 | 37.6346179 | 3.463929161 | 1.531678148 | 0.569945943 | 17.75390625 | 19.82421875 | 21.09375 | 21.77734375 | 23.02734375 | 23.57421875 | 24.996875 | 26.49464526 | 26.9596197 | 27.71520316 | 28.48125 | 29.60625 | 33.08 | 0.203670803 |
| 2001 | 0 | 85 | 0 | 109 | 153 | 79 | 52 | 0.753424337 | 10.95678404 | 29.12721473 | 8.57886321 | 22.75543692 | 20.50859849 | 1.218276734 | 13.04533246 | 4.362868954 | 5.46E-05 | 0.844775966 | 0.034951045 | 0.042069504 | 0.870126541 | 2.736586553 | 7.473458463 | 0.14721108 | 53.51363813 | 2.349353177 | 47.34954406 | 27.0082354 | 5.78E-05 | 1.128441629 | 0.256167948 | 18.41600144 | 19.89326397 | 21.41896134 | 22.46031034 | 23.4532245 | 24.13131223 | 25.7971836 | 26.9596197 | 27.61833348 | 28.6838999 | 29.09075253 | 30.09819715 | 33.38 | 0.895168033 |
| 2002 | 0 | 91 | 0 | 75 | 220 | 66 | 130 | 0.234397598 | 1.390282218 | 9.39117864 | 11.89952747 | 18.9313879 | 24.40495329 | 0.851200878 | 7.105798018 | 5.764872321 | 0.357514962 | 0.57885251 | 0.000515762 | 0.009201389 | 0.289012052 | 4.127445319 | 15.31230231 | 6.64E-05 | 123.1184549 | 4.146098045 | 28.7433116 | 32.52787744 | 0.611107422 | 0.982526072 | 0.306578244 | 18.41600144 | 19.89326397 | 21.41896134 | 22.46031034 | 23.4532245 | 24.03444255 | 25.7971836 | 26.9596197 | 27.54083774 | 28.6838999 | 29.09075253 | 30.09819715 | 33.38 | 1.048187545 |
| 2003 | 0 | 74 | 0 | 54 | 141 | 69 | 55 | 0.778626917 | 2.986866971 | 11.58527787 | 5.740028576 | 4.698593186 | 17.14922124 | 0.825623512 | 6.992451449 | 1.741243201 | 0.442444685 | 0.549039299 | 0.002565288 | 0.071684447 | 0.205108575 | 1.588600772 | 3.520406477 | 6.64E-05 | 54.84419516 | 2.315923696 | 47.19075944 | 16.91826411 | 1.045536727 | 0.390526988 | 0.322396445 | 18.41600144 | 19.99013364 | 21.39474392 | 22.46031034 | 23.4532245 | 24.05865997 | 25.7971836 | 26.88212396 | 27.56021168 | 28.6838999 | 29.09075253 | 29.88508386 | 33.38 | 0.579504092 |
| 2004 | 0 | 73 | 0 | 237 | 229 | 46 | 154 | 10.03048448 | 34.55412924 | 78.70175403 | 6.126630057 | 72.32795104 | 25.43658283 | 2.270718602 | 5.474347478 | 9.298754695 | 0.635610769 | 0.923506454 | 0.17859874 | 1.140983429 | 4.289634522 | 10.13420626 | 26.63700585 | 6.64E-05 | 128.1334894 | 4.566800197 | 10.17628543 | 28.46516175 | 1.170461254 | 0.66658184 | 0.226848154 | 18.41600144 | 20.0627859 | 21.29787425 | 22.46031034 | 23.4532245 | 24.13131223 | 25.7971836 | 27.03711544 | 27.52146381 | 28.6838999 | 29.09075253 | 30.09819715 | 33.38 | 0.561846784 |
| 2005 | 0 | 70 | 0 | 83 | 209 | 74 | 112 | 1.932497581 | 2.442831415 | 9.853830475 | 14.05057106 | 27.47321045 | 14.02808902 | 1.422376658 | 4.031082045 | 2.421310356 | 0.473971904 | 0.469806062 | 5.78E-05 | 0.204204474 | 0.357440704 | 1.853282705 | 14.19666087 | 6.64E-05 | 106.1263776 | 6.466336608 | 34.54499312 | 29.88412891 | 1.869110999 | 0.430047517 | 0.26888509 | 18.41600144 | 20.0627859 | 21.29787425 | 22.46031034 | 23.4532245 | 24.13131223 | 25.715625 | 27.03711544 | 27.52146381 | 28.60625 | 29.184375 | 29.91875 | 33.38 | 0.412352061 |
| 2006 | 0 | 79 | 0 | 91 | 182 | 61 | 99 | 0.70904729 | 2.083134694 | 12.5530492 | 11.94201698 | 34.98446105 | 18.63018581 | 1.365179179 | 8.568899235 | 6.500706927 | 0.429397889 | 0.499272628 | 0.029363277 | 0.056490019 | 0.333622081 | 6.20E-05 | 15.108913 | 6.509506985 | 88.34836501 | 4.476043166 | 23.21376494 | 30.41604496 | 1.149157457 | 0.616239165 | 0.231869401 | 18.41600144 | 20.0627859 | 21.29787425 | 22.46031034 | 23.4532245 | 24.13131223 | 25.7971836 | 27.03711544 | 27.52146381 | 28.6838999 | 29.04375 | 30.028125 | 33.38 | 0.522193955 |
| 2007 | 0 | 71 | 0 | 236 | 177 | 34 | 115 | 15.48357666 | 42.20365046 | 66.67945178 | 3.237314994 | 73.87076368 | 16.12869845 | 1.813765738 | 1.961790378 | 9.749979099 | 0.725909622 | 0.778685492 | 0.214048763 | 2.189164251 | 6.105410434 | 7.602207145 | 22.2630404 | 4.983846809 | 97.6269119 | 3.824450466 | 1.450926016 | 26.67939645 | 1.51406791 | 0.350221136 | 0.172608834 | 18.2421875 | 20.0627859 | 21.29787425 | 22.36328125 | 23.49609375 | 24.2578125 | 25.7971836 | 27.03711544 | 27.52146381 | 28.6838999 | 29.09075253 | 30.09819715 | 33.38 | 0.787598975 |
| 2008 | 0 | 76 | 0 | 61 | 173 | 42 | 111 | 0.329602274 | 1.062983384 | 6.55388102 | 28.19666027 | 8.07106604 | 13.15962876 | 0.946690444 | 4.328066915 | 3.89557511 | 0.432669196 | 0.38124176 | 5.78E-05 | 0.031886197 | 0.186161324 | 3.525668039 | 9.588783738 | 6.64E-05 | 105.9062619 | 4.657072323 | 16.90456719 | 20.37912828 | 1.093845519 | 0.372359152 | 0.204351444 | 18.41600144 | 20.0627859 | 21.640625 | 22.3828125 | 23.515625 | 24.0234375 | 25.7971836 | 26.965625 | 27.52146381 | 28.6838999 | 29.09075253 | 30.09819715 | 33.38 | 0.253924885 |
| 2009 | 0 | 70 | 0 | 168 | 198 | 59 | 117 | 7.13490457 | 22.01727311 | 51.43586637 | 2.425368494 | 46.21137424 | 14.33594387 | 1.670462867 | 6.38664227 | 7.450090635 | 0.871781677 | 0.47286672 | 0.075437892 | 0.816528066 | 2.840506277 | 6.86573211 | 17.10204829 | 6.64E-05 | 100.8429154 | 5.100073121 | 17.59131443 | 32.0730716 | 1.796984992 | 0.368803136 | 0.258001724 | 18.28125 | 20.05859375 | 21.30859375 | 22.34375 | 23.4532245 | 24.13131223 | 25.7971836 | 27.03711544 | 27.52146381 | 28.6838999 | 29.16875 | 29.996875 | 33.38 | 0.304422045 |
| 2010 | 0 | 81 | 0 | 185 | 175 | 39 | 115 | 10.0161432 | 39.0595905 | 60.79750096 | 9.397687455 | 81.91486706 | 26.64358776 | 2.540867542 | 4.955424494 | 9.948884099 | 0.556203265 | 0.947989348 | 0.179170127 | 1.115849026 | 4.970919715 | 6.12805221 | 28.33384571 | 6.64E-05 | 129.4123767 | 5.247772064 | 9.274202411 | 29.62136846 | 1.112488411 | 0.646425402 | 0.234168439 | 18.41600144 | 20.1171875 | 21.25 | 22.3046875 | 23.4532245 | 24.13131223 | 25.7971836 | 27.03711544 | 27.52146381 | 28.6838999 | 29.09075253 | 30.09819715 | 33.38 | 0.492432842 |
| 2011 | 0 | 81 | 0 | 86 | 205 | 80 | 107 | 2.011945059 | 5.039535553 | 10.77638635 | 24.39448864 | 18.55318094 | 10.025552 | 1.012096635 | 8.768797585 | 4.610651111 | 0.352536433 | 0.39514251 | 5.78E-05 | 0.182204748 | 0.670599338 | 1.825393178 | 10.71303127 | 6.64E-05 | 96.67648056 | 5.30981599 | 42.52864663 | 28.93963754 | 1.354959888 | 0.400706399 | 0.297809229 | 18.3203125 | 20.25390625 | 21.2890625 | 22.28515625 | 23.4532245 | 24.04296875 | 25.7971836 | 26.871875 | 27.52146381 | 28.6838999 | 29.09075253 | 29.996875 | 33.38 | 0.240339363 |
