## Supplementary material for "Distinctive features of lipoprotein profiles in stroke patients": Fig. S1: PC.htm

Support

Jump to S1 and  S2 .

### S1 Fig. TG and Cholesterol

#### PC\_samples

| PC1, 2 | PC3, 4 | PC5, 6 |
| --- | --- | --- |
| PC7, 8 | PC9, 10 |  |

#### PC\_items

| PC1, 2 | PC3, 4 | PC5, 6 |
| --- | --- | --- |
| PC7, 8 | PC9, 10 |  |

### S2 Fig. Current Methods

  

#### PC\_samples

| PC1, 2 | PC3, 4 |
| --- | --- |

  

#### PC\_items

  
  

index
