## Supplementary material for "Distinctive features of lipoprotein profiles in stroke patients": Fig. S1: PCitem.htm

Support


### Supporting information

| item | PC1 | PC2 | PC3 | PC4 | PC5 | PC6 | PC7 | PC8 | PC9 | PC10 | PC11 | PC12 | PC13 | PC14 | PC15 | PC16 | PC17 | PC18 | PC19 | PC20 | PC21 |
| --- | --- | --- | --- | --- | --- | --- | --- | --- | --- | --- | --- | --- | --- | --- | --- | --- | --- | --- | --- | --- | --- |
| g VLDL | -0.062540421 | 0.726403527 | -0.029102469 | 0.300717732 | -0.098622603 | -0.038391771 | -0.426008087 | -0.049726056 | -0.039198422 | -0.120496976 | 0.150100199 | 0.111437988 | 0.049049029 | 0.12346817 | -0.020698357 | 0.150432184 | -0.140763512 | -0.050022181 | -0.305738271 | -0.129758947 | -0.000616841 |
| g Lp(a) | 0.286366227 | 0.446324442 | -0.242278804 | 0.113766497 | 0.23070838 | 0.073928108 | -0.144626779 | -0.004948561 | -0.039930553 | 0.190691131 | 0.050926301 | -0.22719553 | 0.149158836 | 0.28298246 | -0.034372969 | 0.059589919 | -0.026733078 | -0.190920813 | -0.083111821 | 0.244066656 | 0.002436729 |
| g TR | -0.455111544 | 1.501916824 | -0.220078494 | -0.20471362 | -0.072171551 | 0.236257749 | -0.561807545 | 0.311894639 | 0.288461218 | -0.759396284 | -0.366876368 | -0.142264769 | -0.312204059 | -0.060546623 | 0.132427308 | 0.061930662 | -0.092695752 | -0.018860252 | 0.05116262 | 0.041896095 | 0.000383173 |
| g LDL1 | 1.271184415 | 1.348067545 | 1.072295842 | 0.548480446 | 0.645841268 | -0.106071097 | 0.69238483 | -0.274452906 | 0.013446139 | -0.218143737 | 0.178671455 | -0.10238395 | -0.090675305 | -0.085283813 | 0.107283294 | -0.050141722 | -0.020649012 | 0.01354076 | -0.020599959 | 0.020639077 | -0.045092604 |
| g LAC1 | 1.028797292 | 1.188247697 | -0.429911504 | 0.419039048 | 0.189069216 | 0.418981634 | -0.106858638 | -0.089682654 | -0.105871301 | 0.330700443 | -0.081504073 | 0.253843312 | -0.032220355 | 0.314711981 | -0.11063474 | 0.036494173 | -0.199453056 | 0.259339957 | 0.098338046 | 0.003384006 | 0.002875289 |
| g LDL2+LAC2 | -0.265119144 | 1.120905402 | 0.333635587 | -0.005188922 | 0.487200519 | -0.114875106 | -0.463930856 | -0.015730266 | 0.128057231 | -0.067951646 | -0.334362685 | -0.04272753 | 0.332917071 | 0.206665021 | -0.05742349 | -0.006937871 | 0.362576027 | 0.096601759 | -0.013992068 | -0.039838688 | -0.001834847 |
| g mHDL | 0.059183477 | 0.948332518 | -0.57622492 | -0.591387344 | 0.241940155 | -0.833907212 | -0.18243057 | -0.587392043 | -0.006420257 | -0.075567011 | 0.132107666 | -0.167190642 | -0.10805043 | -0.108024907 | -0.490784446 | -0.071449854 | -0.03962666 | -0.038575483 | 0.039072159 | -0.01517082 | -0.000453585 |
| g HDL1 | -1.919835675 | 0.75378512 | 0.042849987 | 0.622907759 | 0.307481395 | 0.455081453 | 0.156676625 | 0.614159539 | -0.731962392 | 0.04103225 | -0.027879435 | -0.220795704 | 0.064234569 | -0.180939801 | -0.271120536 | -0.097327266 | -0.022000544 | -0.027829982 | 0.005244269 | -0.018266911 | -0.000773428 |
| g HDL2 | -0.844769399 | 1.053423524 | -0.237162058 | 0.38343164 | 0.012252815 | -0.180223876 | -0.568158753 | -0.32653903 | 0.169669005 | 0.693708317 | -0.131913353 | -0.084232892 | 0.051733443 | -0.408508036 | 0.294857367 | -0.105235248 | -0.041031207 | 0.031058561 | -0.026529366 | 0.025608199 | -0.001170955 |
| VLDL | -0.231257747 | 0.22486672 | 0.009155188 | 0.850610739 | -0.204481458 | -0.153188409 | -0.500431178 | -0.135001928 | -0.110348926 | -0.402077162 | 0.344620663 | 0.728767868 | 0.210115216 | -0.091350398 | -0.015006006 | -0.075979442 | 0.028729323 | -0.098243521 | 0.095554397 | 0.048164956 | 0.00023692 |
| Lp(a) | 0.55909367 | 0.126975488 | -0.507944943 | 0.471628452 | 0.161699578 | 0.214331957 | -0.022135394 | -0.240095306 | 0.052571228 | -0.031352845 | 0.036435059 | -0.420693011 | 0.27999308 | 0.138988361 | 0.158222869 | 0.056265661 | -0.075144845 | -0.221835451 | 0.164993399 | -0.140942881 | -2.27E-05 |
| LDL1 | -0.227010282 | 0.235600878 | -0.375428877 | 0.433562951 | -0.248349432 | 0.238107672 | 0.411841041 | -0.337226151 | 0.154894626 | -0.446848968 | 0.37348678 | -0.364650535 | 0.198988772 | -0.0960615 | 0.049647274 | -0.059758975 | 0.03119709 | 0.196118763 | -0.054095329 | 0.042704855 | 0.047254123 |
| LAC1 | 1.161026244 | 0.724865595 | -0.999503859 | 0.106862203 | -0.229155287 | 0.824295141 | 0.635729771 | -0.260943124 | 0.113199825 | 0.058129041 | -0.313962958 | 0.297656143 | -0.11112023 | -0.162453965 | -0.113186371 | -0.035988691 | 0.164351154 | -0.127517191 | -0.070741704 | -0.012276117 | -0.000549957 |
| LDL2+LAC2 | -1.07338972 | -0.239277291 | 0.50210754 | -0.315465973 | 0.040610229 | 0.043656528 | 0.472226318 | -0.157589601 | 0.414115844 | -0.138703154 | -0.517705985 | 0.181811407 | 0.470666 | -0.058143649 | -0.136797726 | -0.087177986 | -0.235681598 | -0.013487545 | -0.016564676 | 0.016761197 | -0.001027682 |
| mHDL | -0.934148288 | 1.0260927 | -0.946200119 | -0.488069593 | -0.023819673 | -0.832962633 | 0.738157748 | 0.136277315 | -0.478183932 | -0.103050342 | -0.081250361 | 0.201181317 | 0.099768084 | 0.130036344 | 0.326599213 | 0.008408267 | -2.62E-05 | 0.015000178 | -0.006496575 | -0.001199817 | -0.002528628 |
| HDL1 | -2.908960023 | -0.813058636 | -0.020679938 | 0.251332914 | 0.707112273 | 0.339338486 | 0.086090079 | -0.780588577 | -0.029199324 | -0.059779429 | -0.043917157 | 0.106198584 | -0.299815463 | 0.182774211 | 0.065169571 | 0.081358849 | 0.028531123 | -0.012382914 | 0.001754426 | 0.001514223 | -0.00099366 |
| HDL2 | -1.337224494 | 0.702802494 | 0.147009456 | 1.005551896 | -0.85097524 | -0.538963644 | 0.476807193 | 0.29002812 | 0.627412281 | 0.243968406 | 0.018339381 | -0.047209654 | -0.175215333 | 0.172503008 | -0.141318802 | 0.071300383 | 0.0480454 | -0.038144444 | 0.033934981 | -0.012629106 | 0.000276251 |
| CM1 | -1.023152209 | 1.086199165 | -0.040886122 | -1.006309682 | 0.510233421 | 0.506525753 | 0.156997584 | 0.343113521 | 0.491346333 | 0.206762731 | 0.606957641 | 0.179790986 | 0.103539246 | -0.070701767 | 0.00260512 | 0.172908366 | 0.012319924 | -0.023842611 | 0.038706225 | -0.015943761 | -0.001227582 |
| CM2 | -0.542559734 | 1.208220181 | 0.969691578 | -0.719790026 | -1.286165623 | 0.426205808 | -0.044204805 | -0.507524578 | -0.435727383 | 0.088443201 | 0.048253945 | -0.063988809 | 0.022388647 | 0.090278957 | 0.023993979 | 0.03048575 | 0.019135865 | -0.035623806 | 0.042206833 | 0.000229625 | -7.57E-05 |
| Lp(a) | 0.751696882 | 0.71777828 | 0.897234016 | 0.008496401 | 0.579209179 | -0.251273435 | 0.189268107 | 0.042285319 | -0.093975262 | 0.145323393 | -0.140009992 | 0.17984823 | -0.170272312 | -0.010738063 | 0.046356857 | 0.003095443 | -0.01871154 | -0.121118738 | 0.017083343 | -0.025707567 | 0.073120036 |
| free Gly | 0.291157202 | -0.207075911 | 0.047185058 | 0.275539484 | -0.013364727 | -0.194490178 | 0.097490218 | -0.124190755 | -0.198691175 | -0.02166997 | -0.173883964 | -0.00652024 | 0.112046593 | -0.252133886 | -0.07175619 | 0.652458016 | 0.020260287 | 0.045849676 | 0.036450831 | 0.033147483 | 0.000181856 |
