## Supplementary material for "Distinctive features of lipoprotein profiles in stroke patients": Fig. S1: PCsample.htm

Support


### Supporting information

| sample | PC1 | PC2 | PC3 | PC4 | PC5 | PC6 | PC7 | PC8 | PC9 | PC10 | PC11 | PC12 | PC13 | PC14 | PC15 | PC16 | PC17 | PC18 | PC19 | PC20 |
| --- | --- | --- | --- | --- | --- | --- | --- | --- | --- | --- | --- | --- | --- | --- | --- | --- | --- | --- | --- | --- |
| 1 | 0.88619945 | 0.236778685 | 0.221797092 | 0.130778911 | -0.676982138 | 0.802780381 | -0.334092884 | 0.4073979 | 0.78289477 | 0.200619117 | -0.071878146 | -0.164306899 | -0.026016339 | -0.06294699 | -0.224284545 | -0.168730527 | 0.047668018 | 0.091358319 | 0.137198118 | 0.067880642 |
| 2 | 0.839413343 | 1.547368256 | -0.235636764 | 0.541292069 | -0.180566202 | -0.132396668 | 0.061108638 | 0.468259491 | -0.409163026 | 0.37818046 | 0.410887889 | -0.161516154 | -0.117956285 | 0.133095884 | -0.100826734 | -0.166178058 | -0.007582405 | -0.096477608 | 0.055318591 | -0.035199892 |
| 6 | 1.603116662 | -2.86426041 | -0.407084945 | -0.108223149 | -0.088002869 | -0.228923423 | -0.817821396 | 0.247578884 | -0.390670688 | 0.014956201 | 0.395000604 | 0.031848435 | -0.414959626 | 0.102689918 | -0.207884627 | 0.241652225 | -0.016862572 | -0.036579599 | 0.136284995 | 0.038331371 |
| 7 | 0.373260934 | 0.191081337 | -0.392881606 | 1.172814729 | 0.32474823 | 0.832119499 | 0.708040931 | 0.006581571 | -0.630805182 | -1.075624002 | -0.257768574 | -0.090589248 | -0.168832079 | 0.200225473 | -0.127086837 | 0.402696265 | 0.162106608 | 0.184925554 | -0.040769258 | 0.154509052 |
| 8 | 0.490450087 | 0.55915424 | 0.352465158 | 0.088040163 | -0.227731186 | -0.984019367 | -0.15531897 | 0.865569832 | -0.266802434 | -0.009797799 | -0.317541592 | -0.044434831 | 0.053086722 | 0.21685457 | -0.241213062 | 0.172106937 | -0.33354016 | 0.078651993 | 0.299680743 | 1.30E-02 |
| 9 | 0.334266421 | 0.475481585 | -0.4049871 | 0.471149134 | -0.495821727 | -0.16543197 | -0.32617684 | 1.025340668 | -0.761120538 | 0.317892848 | 0.100094769 | 0.085125488 | 0.442837755 | 0.236265631 | -0.312370276 | 0.138559132 | 0.011643971 | 0.141707835 | -0.066287876 | -0.014528251 |
| 10 | 0.600331429 | 0.604465799 | -0.185369497 | 0.868222255 | -0.886999677 | 0.687422209 | 1.022578564 | 1.259039149 | -0.479969988 | -0.475167797 | 0.014725277 | -0.052213716 | 0.236398897 | 0.551473579 | -0.017739963 | 0.025217371 | 0.122739711 | -0.060366079 | -0.110655848 | -0.000534507 |
| 13 | 0.339609911 | 0.717521468 | 0.368184142 | 0.160210151 | -0.48584714 | 0.489565549 | -0.115641937 | 1.416197069 | -0.600878773 | 0.26631701 | -0.009294265 | 0.237425858 | 0.11406771 | 0.247098526 | 0.370474057 | 0.043551969 | -0.248754797 | 0.143115276 | -0.026931839 | 0.083791869 |
| 14 | 0.802953658 | 1.198275338 | 0.027084238 | 0.792320411 | 0.081321437 | -0.826419475 | 0.123452594 | 0.477603288 | 0.307497292 | -0.322076725 | -0.03072861 | -0.151126559 | 0.220505358 | 0.022083712 | -3.89E-02 | -0.034271934 | 0.039978126 | -0.11437838 | 0.180251889 | 0.014074283 |
| 17 | 1.018116933 | 2.036435919 | -0.8087661 | 0.933005394 | -0.310164424 | 0.09894682 | 0.352385674 | 0.106052379 | -0.39915074 | 0.127671107 | 0.636200751 | 0.00010436 | -0.181509435 | -0.195414148 | -0.146122956 | -0.151911817 | 0.107900452 | 2.50E-02 | -0.139567905 | 0.015981162 |
| 20 | 0.787978516 | 3.018621938 | -0.265539519 | 1.593826822 | 0.103595187 | 0.255809229 | 0.57544315 | 0.308987091 | -0.471831158 | 0.225765932 | 0.303192975 | -0.085167483 | 0.025362992 | -0.254123704 | -0.428442853 | 0.169614177 | -0.182284341 | -0.135749518 | -0.0618416 | 0.012463183 |
| 24 | 1.031694226 | -0.052507148 | 0.050792071 | 0.207161043 | -1.344077149 | 0.136856598 | 0.137227583 | -0.327732205 | 0.13096694 | -0.022096623 | 0.14101912 | -0.268070413 | -0.082716618 | -0.380030676 | -0.145010283 | -0.053211205 | 0.002053174 | 0.069601149 | 0.044729985 | 0.01445212 |
| 25 | 1.087596202 | 0.061711101 | -0.345206431 | 1.062712875 | -0.052366645 | -0.500586633 | 0.898835484 | 0.926509475 | -0.495024605 | 0.115865437 | -0.181878043 | -0.159647245 | -0.34912101 | -0.321891462 | -0.112493935 | -0.195192018 | 0.084847398 | 0.035813551 | -0.00423378 | 0.021159093 |
| 26 | 0.877513235 | 1.53360165 | 0.350113274 | 0.994347097 | 0.333452167 | -0.052885855 | 0.332295481 | 0.645530646 | -0.660725713 | -0.340396164 | 0.225702499 | -0.258495638 | 0.128086135 | -0.195137382 | -0.02681728 | -0.236457763 | -0.083061838 | 0.175804838 | 0.012847171 | 0.023762539 |
| 27 | 1.781073489 | 1.778402842 | 0.829190298 | 0.757092237 | 0.451019791 | -1.18786319 | -0.284947117 | -0.55394353 | 0.314249961 | 0.469067867 | 0.45515522 | 0.703235475 | 0.151962347 | 0.314909798 | 0.191909361 | 0.221703291 | 0.09864309 | 0.053275897 | -0.171691156 | -0.134529884 |
| 29 | 1.198583143 | -0.059519108 | 0.391580243 | 0.043430227 | -0.615313454 | -0.545095207 | -0.09749654 | 0.076288088 | 0.278097904 | -0.37346824 | 0.05375821 | 0.146306061 | 0.25230181 | 0.027715399 | 0.248867107 | 0.276182498 | -0.278167155 | 0.057668105 | -0.02805699 | 0.023291415 |
| 31 | 1.513010908 | -0.775104316 | 1.24447543 | 0.661356217 | -0.095435666 | -1.215955172 | -0.514959561 | -0.086408237 | 0.252671351 | 0.383321047 | 0.090966326 | -0.338806409 | 0.000975111 | -0.287918314 | -0.181299108 | 0.012544941 | -0.170967525 | 0.27456962 | 0.194494708 | 0.028798544 |
| 33 | 1.468189733 | 0.520182151 | -0.251801102 | 0.284054111 | -0.450300616 | -0.088835635 | -0.563549007 | -0.01519727 | 0.452129537 | -0.084589698 | 0.217794657 | 0.178520797 | -0.158538591 | -0.023675989 | 0.065766216 | 0.11385901 | 0.030075764 | -8.49E-05 | -6.80E-03 | -0.013564873 |
| 35 | 2.631032246 | -1.53462331 | 0.101450096 | 0.710550463 | -0.157700392 | 0.198586069 | 0.889200361 | 0.461612435 | -0.031464683 | 0.070907048 | -0.32622217 | 0.801977275 | -0.816007136 | -0.379773725 | 0.013422919 | 0.571440327 | 0.235681406 | 0.279752957 | -0.116676191 | 0.010164118 |
| 36 | 1.237905506 | -2.082850485 | 1.271108739 | 0.34568409 | 1.031939701 | -0.244007977 | 0.020023092 | -0.003304248 | -0.521859468 | -0.325875035 | 0.334687249 | -0.265529357 | -0.225002791 | 0.133183844 | 0.059522167 | -0.129331631 | -0.211445948 | 0.012095423 | 0.030770352 | 0.081792618 |
| 37 | 0.84186343 | -0.642468553 | -0.132338208 | 1.127000752 | -0.682212956 | -0.019031031 | 0.552087483 | -0.690011251 | 0.394762957 | 0.048494991 | -0.0594077 | -0.232548265 | -0.379563356 | -0.432434955 | -0.289494547 | -0.12529785 | -0.36514502 | -0.006093733 | -0.000349084 | 0.057178331 |
| 38 | 0.979769314 | -0.47986747 | -0.143366276 | 1.529582414 | -0.519111213 | -0.110417032 | 0.782560643 | 1.042011009 | 0.404850787 | 0.422350227 | -0.262456093 | -0.135144122 | 0.149720076 | 0.097855972 | 0.200130719 | -0.173509825 | -0.101926622 | -0.214372418 | 0.068114078 | -0.041366841 |
| 40 | 0.724207742 | -2.465233269 | -1.270137691 | 0.991683856 | -0.463233977 | 0.432522421 | -0.343354802 | -0.753091146 | 0.384082489 | 0.435842335 | 0.235038825 | 0.206091108 | -0.426538394 | -0.034574701 | 0.00514974 | 0.078785277 | 0.164606368 | -0.068343065 | 0.062146935 | 0.015912339 |
| 41 | 0.58289613 | -2.553545805 | -0.384479605 | 1.008607526 | 0.131374548 | 0.661388209 | -0.308607814 | -0.216099529 | -0.197839168 | -0.087412871 | 0.234080296 | 0.174789912 | -0.251515093 | 0.186153455 | 0.2353636 | 0.06833488 | 0.050513574 | -0.004293466 | -0.097477952 | -0.032528145 |
| 42 | -0.504865892 | -1.910703264 | 1.560734699 | 0.11719287 | 0.674846781 | -0.23568593 | -0.265681902 | -0.121626885 | -0.379290703 | 0.191033671 | -0.20667009 | 0.302853713 | 0.096726336 | -0.391482892 | 0.0792603 | 0.501324826 | 0.052370173 | -0.237073873 | -0.221282343 | -0.092363553 |
| 44 | 0.915420258 | 0.824066132 | 1.606927818 | 0.310874826 | -2.513023558 | 0.868200454 | 0.431303891 | -1.012999963 | -0.75675627 | 0.307484482 | -0.307845359 | -0.235042701 | 0.404522741 | -0.06739071 | -0.048273014 | -0.187772705 | 0.171882794 | -0.007662738 | 0.152632879 | -0.007488122 |
| 46 | -0.588742601 | -2.387659016 | -0.035560012 | 1.157500545 | 0.496815964 | 0.54341824 | -0.586306693 | -0.366515035 | 0.026319755 | 0.315061946 | 0.038605324 | -0.286259097 | 0.304308619 | 0.544803317 | 0.292255227 | 0.009682879 | -0.418870174 | 0.213287429 | 0.102113481 | -0.012450122 |
| 53 | 0.572746779 | -2.213016846 | -0.940367523 | 0.973776034 | -0.277962555 | 0.456668134 | -0.251484662 | -0.400720621 | 0.367090118 | -0.057816132 | 0.164188048 | -0.229894886 | -0.242865521 | 0.165193372 | -0.10600973 | -0.110736029 | 0.008917148 | -0.103054502 | -0.019415393 | 0.019935722 |
| 54 | 0.743616799 | -1.607918692 | 0.678720258 | 1.893687253 | 0.610618692 | 0.378061396 | 0.613235969 | -0.328802844 | -0.051897868 | -0.190159354 | 0.302865661 | 0.301983124 | -0.283451979 | 0.070386845 | 0.107142794 | -0.032646205 | 0.03145763 | 0.004187 | -0.055962438 | -0.089031687 |
| 56 | 1.075302653 | 0.507100316 | 1.168485563 | -0.352883757 | -0.275362573 | 0.198497661 | -0.445109922 | 0.368054053 | 0.138315426 | 0.557460808 | -0.198180145 | -0.11492719 | -0.257366919 | -0.092734187 | -0.216867754 | -0.302908358 | 0.040832267 | -0.116394361 | 0.096744796 | 0.096124038 |
| 60 | 0.154881146 | -0.504639864 | 0.10739098 | 0.982711573 | 0.340501351 | 0.643718532 | 0.591923628 | 0.197447549 | 0.295462733 | -0.191800312 | -0.663755051 | 0.20408113 | 0.087694969 | -0.12218788 | 0.345901014 | 0.006465657 | 0.286626101 | -0.339368268 | -0.141372696 | -0.017044146 |
| 61 | 1.266340289 | -2.538938529 | -1.091760184 | 0.170725519 | 0.012321495 | 0.310268483 | -0.568554332 | -0.671631982 | -0.048933339 | 0.387358771 | 0.011615312 | 0.005334157 | -0.195042425 | -0.177893658 | -0.182693011 | 0.02763827 | -0.07452402 | -0.013176881 | 0.045492711 | 0.05174361 |
| 62 | 1.203573256 | 0.933685516 | 1.072269688 | 0.080080305 | -1.206459604 | -0.225702434 | 0.375394339 | -0.591710691 | -0.227599799 | 0.105772738 | -0.697613424 | -0.035943276 | -0.062593085 | -0.016696951 | -0.088414863 | -0.49673498 | -0.175995849 | 0.064453694 | -0.129018197 | 0.030659743 |
| 63 | 1.021506804 | -2.613192755 | -1.150617795 | 0.349185442 | 0.1271158 | 0.566712957 | -0.524269293 | -0.886602862 | -0.029835225 | -0.238784544 | 0.196385449 | 0.174602925 | -0.111901219 | 0.35703558 | -0.301774907 | -0.043202043 | 0.046653976 | -0.010669942 | 0.094338692 | -0.083137574 |
| 67 | 0.818662618 | 0.238339019 | -0.214408261 | -0.680363782 | -0.812454425 | -0.215078944 | -0.513673882 | 0.186017438 | 0.370819663 | 0.14253151 | 0.217631767 | 0.29289411 | 0.124102138 | 0.07400419 | 0.097337525 | -0.10784586 | -0.192606813 | 0.055758101 | -0.139791334 | 0.040170536 |
| 70 | 2.179211834 | -0.18419088 | -0.218322206 | 0.324144136 | 0.536429546 | -0.265233874 | 0.355127802 | -0.378625877 | 0.301892814 | -0.059726112 | 0.176881388 | -0.036684944 | 0.42652651 | -0.158935982 | 0.177982699 | 0.001744015 | -0.008656362 | -0.076693227 | 0.172284427 | 0.030185041 |
| 88 | 1.904333648 | 0.343738324 | -0.014484595 | 0.133009744 | 0.602222461 | 0.010510334 | -0.25662572 | -0.070223761 | -0.497212465 | 0.069062603 | 0.300660733 | -0.11706856 | 0.088647642 | -0.266685035 | 0.093065644 | 0.020003273 | -0.080640162 | -0.071274646 | 0.113769605 | 0.024812516 |
| 89 | 1.247198489 | 0.835236205 | 0.673119446 | 0.504205347 | -0.209225364 | 0.075301073 | -0.908991791 | 0.242522745 | 0.640300668 | 0.115762021 | -0.050975787 | 0.031006305 | -0.179243507 | 0.063312721 | -0.223554351 | -0.192860685 | 0.057970198 | 0.063936931 | -0.078748496 | 0.056366194 |
| 95 | 1.15406101 | 0.441696441 | 0.464420531 | -0.034773149 | -0.633963344 | -0.504895559 | 0.024856925 | -0.082568337 | 0.557277534 | 0.330877039 | 0.314850017 | 0.239373334 | 0.042474956 | -0.017540738 | 0.147652226 | 0.002858257 | 0.112278148 | 0.28646934 | -0.010111962 | -0.041524617 |
| 99 | 1.774693443 | 0.855328282 | 0.549937789 | 0.984837255 | -0.096053343 | -0.489734397 | -0.392096669 | -0.391164945 | 0.517965279 | -0.577536159 | -0.538902855 | -0.017361442 | 0.085334487 | 0.036617097 | 0.017262507 | -0.049237053 | 0.03125024 | 0.051034084 | -0.0501719 | 0.050971275 |
| 100 | 1.603043164 | 0.346105347 | -0.084371152 | 0.694648193 | -0.472139318 | -0.611137948 | -0.260733855 | -0.203295875 | 0.217632435 | -0.276514422 | -0.273037842 | 0.139521825 | -0.068578418 | 0.152980702 | 0.074332933 | -0.295176997 | 0.051931198 | -0.058236905 | -0.211466865 | -0.015631447 |
| 104 | 2.25075957 | -0.477570986 | 0.339184838 | -0.41588117 | 0.601262339 | -0.238362022 | 0.316176435 | -0.289794477 | -0.112334196 | 0.272735716 | 0.17506551 | 0.097817358 | 1.34E-01 | -0.046652374 | 0.062581139 | -0.318901333 | 0.171602364 | 0.094128309 | 0.084401879 | -0.02275149 |
| 109 | 0.964289799 | 0.590201467 | -0.461161484 | 0.662574926 | -0.653024436 | -0.19587805 | -0.374998273 | 0.175649159 | 0.594261144 | -0.433717018 | 0.010033972 | -0.253129625 | 0.117722017 | -0.01573342 | -0.036704868 | -0.209583124 | -0.031957353 | 0.071605087 | -0.0444689 | 0.078418268 |
| 113 | 1.239477176 | 0.833599543 | -0.141425867 | 0.137244074 | -0.875323982 | -0.342462376 | -0.115818559 | -0.371220418 | 0.550243127 | -0.189128753 | -0.237501449 | 0.293908289 | 0.280148248 | 0.062592711 | 0.049267133 | 0.325582836 | 0.064418065 | 0.059090057 | -0.075950911 | 0.039663078 |
| 120 | 1.498357515 | 0.745605913 | 1.045857199 | 0.817910344 | -0.041771651 | -0.245913009 | -0.588588499 | -0.021508771 | 0.479219168 | -0.411788873 | 0.344996945 | -0.212897107 | -0.036291257 | -0.024874315 | 0.182442824 | -0.223141844 | 0.114733465 | 0.019503549 | -0.112897122 | -0.034987824 |
| 126 | 2.027874763 | -0.408582552 | -0.639396381 | -0.214360439 | 0.333450438 | 0.068939592 | -0.349210366 | 0.20827902 | -0.397230411 | -0.252446564 | 0.12281736 | -0.209992516 | -0.206285521 | 0.040771514 | -0.039110864 | 0.040875668 | 0.071876547 | -0.142203016 | 0.327271671 | -0.013467672 |
| 128 | 0.556927175 | 2.270488948 | 1.269424532 | 1.244276474 | 0.574168174 | 0.057172834 | -1.158143551 | 0.087630896 | 0.269332902 | 0.590792472 | -0.157568664 | 0.558994753 | -0.571194174 | 0.219049525 | -0.651678651 | 0.235822762 | -2.21E-01 | -0.224568302 | 0.091048344 | -0.111697799 |
| 130 | 1.342283947 | 0.120052508 | -0.520001101 | -0.495192841 | -0.860088156 | -0.043624947 | -0.531642631 | 0.073431044 | 0.261971805 | 0.014521902 | -0.294687674 | 0.306223605 | -0.104086413 | 0.36390416 | -0.048219699 | 0.057976998 | 0.236971033 | -0.00310289 | 0.106966337 | -0.0226936 |
| 133 | 1.321496222 | 0.394376903 | 0.028958661 | 0.489856107 | -0.278153119 | -0.79622347 | -0.404125469 | 0.395881872 | 0.445461345 | -0.307819782 | -0.722701573 | -0.001875275 | -0.327607132 | 0.231295428 | 0.087307576 | 0.040987801 | -0.04745989 | -0.196341622 | 0.108694183 | 0.121259792 |
| 139 | 1.597382703 | 0.021364452 | -0.199189158 | -0.879274131 | 0.127267026 | -0.120858964 | -0.683137831 | 0.00502305 | 0.204752463 | 0.203530339 | 0.000282597 | -0.269535987 | 0.086069345 | 0.080953681 | 0.209836543 | 0.15855763 | 0.162359274 | -0.196680301 | -0.019185084 | -0.112495157 |
| 140 | 1.929858552 | -0.768412505 | -0.454853064 | -0.176323139 | -0.146712321 | -0.053397439 | 0.512361452 | -0.106298577 | -0.015494837 | -0.405725124 | -0.07409293 | 0.20775813 | 0.331474028 | 0.123127889 | -0.331101971 | -0.008025647 | -0.087639937 | -0.145305935 | 0.039953881 | -0.090237988 |
| 141 | 2.306199029 | -0.090418945 | -0.030343729 | -0.120647002 | 0.623841944 | -0.150969416 | -0.023892758 | -0.269656742 | -0.01608383 | -0.140227261 | 0.01638309 | -0.117312332 | 0.010748684 | -0.079561639 | 0.31179137 | -0.044508654 | -0.05271467 | -0.138068607 | 0.021173647 | 0.104578035 |
| 149 | 1.645798145 | -0.029209129 | -0.223488945 | -0.437854288 | 0.239153357 | -0.253426765 | -0.893230435 | 0.014589913 | -0.365848712 | 0.348584646 | 0.063244448 | -0.182364105 | 0.070662401 | -0.018441874 | 0.336430762 | -0.016482939 | -0.030765343 | -0.098652949 | 0.003167079 | -0.007644168 |
| 157 | 1.986661968 | 1.661085388 | 0.10777459 | 1.036044866 | 0.105757373 | -0.36015899 | -0.291751847 | -0.549615933 | 0.643488201 | 0.806024731 | 0.746612026 | 0.342480717 | -0.115434457 | 0.461290948 | 0.117754575 | 0.112029279 | 0.007455051 | -0.022734878 | -0.027013206 | -0.021323095 |
| 161 | 1.374632462 | 0.133254116 | 0.074305575 | -0.417552105 | -0.646753167 | -0.344021427 | 0.00812365 | 0.023862949 | 0.285935027 | 0.014404586 | 0.116464819 | 0.573950227 | -0.110015091 | 0.11235435 | -0.049454583 | 0.109185182 | 0.043883943 | -0.106646986 | -0.072270934 | -1.83E-01 |
| 174 | 1.514942366 | 1.488856662 | 0.126739016 | 1.368900788 | -0.290364576 | 0.001832114 | -0.083074394 | -0.277798424 | 0.59713904 | -0.021489259 | 0.2640367 | 0.077875841 | 0.103027111 | -0.057183025 | -0.075258798 | 0.245360119 | 0.057269777 | 0.2193018 | -0.071574758 | 0.035386476 |
| 175 | 1.532176216 | 0.205421935 | -0.045019287 | 0.643851619 | -0.51339895 | -0.201745332 | -0.466902154 | 0.165464063 | -0.044409494 | -0.058466542 | 0.274914181 | 0.207428349 | -0.321562721 | -0.132865977 | 0.28578236 | 0.20636685 | 0.04420574 | 0.040554976 | -0.102356516 | -0.025481283 |
| 177 | 1.095214832 | 0.334845432 | 1.807884178 | -0.077887231 | -3.28209854 | 1.31222998 | -0.148509144 | -1.082976851 | -1.092512299 | 0.239588976 | 0.025983215 | -0.154721175 | 0.054334488 | 0.146793228 | 0.400463309 | 0.289965516 | 0.188962022 | -0.261000464 | 0.296289301 | -0.040454989 |
| 187 | 2.089014584 | 0.022733875 | -0.009540014 | 0.330315375 | 0.575690987 | -0.22343683 | 0.564104686 | -0.22701946 | 0.107374954 | 0.062752814 | -0.135870697 | 0.091182164 | 0.542866549 | 0.183449836 | -0.178437097 | 0.031821676 | 0.090946751 | 0.093397228 | 0.299278279 | -0.059409099 |
| 189 | 0.897697685 | 0.373810376 | -0.202813568 | -0.051307212 | -0.841082657 | -0.36461687 | -0.258081323 | 0.044395538 | 0.367130271 | -0.121733535 | -0.150888704 | 0.279124986 | 0.366578108 | -0.058025669 | -0.026610945 | 0.356813825 | -0.171547191 | -0.086533365 | -0.122501709 | 0.073703229 |
| 198 | 1.872675566 | -2.100180117 | -1.066673403 | 0.27864357 | -0.510479214 | 0.285093792 | -0.250214473 | 0.594503677 | -0.130876879 | 0.709766437 | -0.158806511 | 0.227864225 | 0.106740351 | 0.254645096 | 0.44210915 | -0.300141108 | 0.008804606 | 0.053520025 | -0.07715529 | 0.010481628 |
| 200 | 0.714878031 | 0.283176774 | -0.149715811 | -0.853133619 | -0.899700755 | 0.10958326 | -0.6296992 | 0.521906262 | 0.367104007 | -0.016197841 | 0.019572272 | 0.216895338 | -0.104243521 | 0.162461775 | 0.011501311 | -0.030650245 | -0.079243341 | -0.007254096 | -0.090111714 | 0.127748866 |
| 206 | 1.51921162 | -1.515738398 | 0.343962561 | -0.413837058 | 0.260887755 | -1.20899873 | -0.359932912 | 0.469997574 | -0.776601577 | -0.362460331 | 0.235036801 | 0.277763627 | 0.367398897 | 0.346659537 | -0.014691164 | -0.244398984 | -0.069906499 | 0.315286753 | -0.052014695 | -0.177209163 |
| 207 | 1.575184815 | 0.878142168 | -0.09318223 | -0.071540235 | 0.548764051 | 0.367492271 | 0.102573859 | -0.003021828 | -0.493621298 | 0.412214005 | 0.120326927 | -0.287005067 | 0.339417805 | 0.029074822 | -0.13169206 | -0.321654243 | -0.074469369 | -0.089773341 | -0.106633629 | -0.054314639 |
| 209 | 2.561768335 | -0.522343357 | 0.715290338 | -0.68806444 | 0.692134009 | -0.028381706 | 1.251301351 | -0.169603419 | -0.144366173 | 0.24125756 | -0.030840654 | 0.285525833 | -0.438750374 | -0.240234544 | -0.012963157 | -0.26696952 | -0.048084739 | 0.085798496 | -0.072507275 | 0.072903097 |
| 226 | 1.517312455 | -1.34981804 | -0.523933692 | -0.310218378 | -0.187703209 | 0.129195678 | -0.279780991 | -0.214353209 | -0.158467093 | -0.552901156 | 0.167055106 | 0.333884397 | 0.461253842 | 0.154355655 | -0.537244683 | -0.008799023 | -0.116996442 | 0.276742063 | -0.077606498 | -0.059668226 |
| 227 | 0.593566542 | -1.792315613 | 1.162495035 | 0.301932447 | 0.044571401 | -0.924923333 | -0.629615803 | 0.359720636 | -0.096441251 | -0.140067915 | 0.056755307 | -0.012704967 | 0.002217055 | -0.12476223 | -0.469050822 | 0.007326514 | 0.115979235 | -0.436485502 | -0.207873666 | -0.052083312 |
| 229 | 1.559417375 | -0.130771408 | -0.766926196 | -0.374331093 | -0.336138983 | 0.039016502 | 0.128853431 | -0.033012919 | 0.24553242 | -0.09276473 | -0.122211212 | 0.0190689 | 0.037323113 | -0.23295309 | 0.099148863 | 0.100878329 | -0.149016493 | -0.072051434 | 0.110169929 | 0.093450125 |
| 236 | 2.100808807 | 0.289024305 | -0.429292555 | 0.202840822 | 0.521695905 | 0.17870149 | -0.08464128 | -0.295361559 | -0.49800296 | 0.168924891 | 0.222009101 | -0.129355567 | 0.176267282 | -0.253079354 | -0.003596352 | 0.013144054 | 0.184338598 | 0.127613996 | -0.04688655 | 0.049432047 |
| 237 | 2.128511463 | -0.0908275 | -0.222076786 | 0.114833683 | 0.254386784 | 0.161596082 | 1.124222142 | 0.122175838 | -0.278799774 | 0.117478319 | 0.212711628 | -0.900656124 | -0.125352607 | 0.498049777 | -0.257147374 | 0.233513408 | 0.168480547 | -0.019114273 | -0.082668716 | -0.162668147 |
| 241 | 0.848559137 | -2.041457597 | 0.264236376 | -0.067314238 | 0.164092572 | -0.870626378 | -1.195266223 | -0.064881506 | 0.377850762 | -0.670922675 | -0.056420823 | -0.483204404 | 0.285166418 | -0.110399208 | -0.128141065 | -0.07936377 | 0.137307162 | -0.1531021 | -0.089467714 | 0.086876717 |
| 248 | 0.535120966 | -1.622973312 | 1.422225788 | 1.57697131 | 0.580630215 | -0.129207779 | 0.24063281 | -0.144311688 | 0.001038654 | -0.321330033 | 0.267070224 | -0.040252003 | 0.17081059 | 0.643814345 | 0.133600674 | 0.132951921 | -0.215629808 | 0.166889215 | 0.131054995 | -0.059942291 |
| 261 | 1.097918898 | -1.752722373 | 1.361600235 | 1.25070781 | 0.111273418 | -0.825750839 | 0.092718237 | 1.017966195 | 0.114520645 | 0.251217338 | 0.100644766 | -0.618043072 | 0.390646085 | -0.466786664 | 0.218924583 | 0.022873199 | 0.209602446 | -0.303457911 | 0.035554839 | -0.006462114 |
| 262 | 1.688892746 | -2.384389551 | -0.624373075 | 0.52536652 | -0.628336817 | 0.602977713 | -0.259371435 | 0.716943981 | -0.10551752 | 0.125934113 | 0.378762867 | 0.144716903 | 0.323041761 | 0.038939894 | -0.072564466 | -0.106626366 | -0.139841994 | -0.123374169 | -0.070203526 | -0.041862254 |
| 277 | 2.139034114 | -0.470627819 | -0.34375819 | -0.379999814 | -0.141821216 | 0.093128088 | 0.756880316 | -0.362922495 | -0.079806197 | -0.631001588 | -0.018043382 | 0.263527486 | 0.200264545 | 0.024835655 | -0.128183049 | -0.028641414 | -0.31359797 | -0.225252792 | -0.061684198 | 0.017283473 |
| 284 | 1.900148871 | -0.021698958 | 0.652351064 | -0.948657751 | 0.279939161 | -0.224103479 | -0.290897523 | 0.006239765 | -0.213413531 | 0.090356656 | 0.356406276 | -0.078617741 | -0.360026898 | 0.067628989 | 0.30827707 | 0.02301842 | -0.055661608 | 0.053028932 | -0.02464018 | 0.20827052 |
| 288 | 2.140793035 | 0.378509521 | 0.41766203 | -0.608147452 | 0.656447397 | -0.357165608 | -0.381977891 | -0.152802465 | -0.190894148 | 0.124158141 | -0.413897001 | 0.214945176 | -0.133177123 | 0.236666967 | 0.210349732 | -0.065056465 | 0.122088743 | -0.045510212 | 0.092013306 | 0.049173244 |
| 291 | 2.15201862 | 0.117412874 | -0.240615417 | -0.460070245 | 0.31340761 | 0.271751552 | 0.662216146 | -0.516948169 | 0.311301184 | 0.422459631 | -0.483068988 | -0.444741326 | 0.344611109 | 0.114156599 | 0.056803239 | -0.031673333 | -0.151623563 | 0.232560116 | -0.24964923 | -0.057359308 |
| 307 | 1.835843565 | -0.368689107 | -0.200358883 | -0.665671815 | 0.149745766 | -0.090675021 | 0.322984687 | -0.376382899 | -0.020093772 | 0.010062696 | 0.139633631 | 0.038356597 | 0.478581403 | -0.321916478 | 0.139786126 | 0.142458761 | -0.172643291 | -0.003391585 | 0.109258079 | 0.022805658 |
| 308 | 1.920648113 | 0.14004871 | -0.154852887 | -0.894003085 | 0.206634352 | 0.048187643 | 0.088460524 | -0.314077127 | -0.064912245 | 0.191874531 | 0.006178855 | -0.01719001 | 0.181534931 | 0.141801596 | 0.1526786 | -0.00921696 | -0.184894693 | -0.076570826 | -0.090550802 | 0.017597709 |
| 309 | 2.185302585 | -0.199317832 | 0.124850249 | -0.170031219 | 0.102695985 | -0.070602931 | 5.94E-01 | -0.607129995 | -0.141512902 | -0.268811067 | 0.244952649 | 0.170243541 | 0.224920606 | -0.407441333 | 0.214369723 | 0.077000079 | -0.417799569 | -0.204209675 | -0.071167382 | 0.118247035 |
| 310 | 2.025538689 | -0.2058399 | 0.163230333 | -1.037814674 | 0.294107614 | 0.09010697 | -0.034531546 | 0.012419704 | -0.025490614 | 0.34766574 | -0.193057599 | -0.121932101 | -0.333089749 | 0.134374677 | 0.089373823 | -0.135798673 | 0.022900008 | 0.094608332 | -0.057615463 | 0.0818834 |
| 317 | 1.979468388 | 0.225859228 | -0.580551153 | 0.206253985 | 0.179941352 | 0.099705974 | 0.222052729 | -0.211783819 | -0.159752749 | -0.511959775 | 0.044203643 | -0.095098096 | 0.25958393 | 0.011552557 | 0.002431113 | 0.100458164 | -0.057649027 | -0.103953597 | 0.038668076 | 0.052865799 |
| 321 | 2.014042451 | -0.263466741 | -0.284435531 | -0.525936337 | 0.221766805 | -0.067233206 | -0.274753017 | -0.106669616 | -0.073370114 | -0.065820919 | -0.795214254 | 0.605141426 | 0.16075923 | -0.082871862 | -0.120026772 | 0.214725479 | 0.077662614 | -0.01121867 | 0.142427987 | 0.019852453 |
| 328 | 1.921502656 | 1.156103737 | 0.729972989 | 0.042766308 | 1.518047881 | 0.006576913 | -0.154626677 | -0.76806692 | -0.349769815 | 0.590173268 | -0.537350401 | 0.055881261 | 0.084036311 | 0.147383701 | -0.068095038 | 0.051534533 | 0.244298499 | 0.161681414 | 0.070287961 | 0.015736093 |
| 330 | 2.531708669 | -0.265748689 | 0.237320065 | -0.245456974 | 0.713724311 | -0.263128058 | 0.130360309 | 0.151507889 | -0.469385468 | -0.424574747 | -0.176376581 | 0.109018818 | -0.6300983 | 0.243482123 | -0.046393569 | -0.044323701 | 0.051460427 | -0.272079066 | 0.095683151 | 0.052007469 |
| 333 | 2.056094767 | -0.449033843 | -0.678188427 | -0.164057821 | -0.170499999 | 0.078415094 | 0.475911056 | -0.437633284 | 0.001517958 | -0.209053244 | 0.084938832 | -0.206050986 | 0.186681893 | -0.164353342 | 0.099193363 | -0.111227332 | 0.013537884 | -0.017107815 | -0.149099722 | 0.079781112 |
| 356 | 1.786089889 | -0.40036463 | -0.187040604 | -0.783817734 | 0.098706428 | -0.039909737 | 0.236700132 | 0.221480081 | -0.347887533 | 0.320092034 | -0.491928562 | -0.457084076 | -0.262037912 | 0.087189629 | 0.027295479 | 0.121746117 | -0.210798789 | 0.001527663 | -0.026164638 | 0.060672837 |
| 360 | 1.657624127 | -0.005197386 | -0.58618966 | -0.970258107 | -0.031427638 | 0.10775939 | -0.405675946 | 0.0474757 | 0.023611917 | 0.316701774 | -0.186157246 | -0.179300607 | -0.107307943 | 0.047605604 | 0.111254384 | 0.232141597 | 0.081259132 | 0.141228847 | 0.04873486 | -0.088653924 |
| 372 | 1.554341368 | 1.023212544 | -0.545584555 | 0.666555283 | -0.125606024 | -0.131004519 | -0.160403994 | -0.106247791 | 0.322052831 | -0.162009024 | 0.111040681 | 0.214970431 | 0.079803085 | 0.133794218 | 0.112343964 | -0.164532845 | -0.004854539 | -0.192213352 | -0.070905619 | 0.014366326 |
| 373 | 1.931205749 | 0.646683639 | -0.248913104 | 0.511306696 | 0.26747663 | -0.422981659 | -0.063972942 | -0.245835537 | 0.224035127 | 0.004198124 | 0.159953158 | -0.344375687 | -0.00893479 | 0.247550509 | 0.36725048 | -0.190871392 | 0.067685446 | -0.143771437 | -0.018520069 | 0.016656104 |
| 374 | 1.412414353 | -0.873659738 | -0.528433485 | 0.173750744 | -0.506960174 | -0.504648462 | 0.590431866 | 0.481711049 | -0.10795335 | 0.256105087 | -0.105563156 | -0.077834876 | -0.387496207 | -0.248375808 | -0.068081922 | -0.031720988 | 0.009009756 | -0.063583578 | -0.006163379 | 0.034035406 |
| 383 | 1.256558218 | 1.605387956 | -0.516103167 | 0.214287345 | -0.021826369 | -0.054495835 | -0.580916602 | 0.035279013 | -0.278024757 | 0.208611658 | 0.151543915 | 0.00482944 | -0.18230239 | -0.121177633 | 0.001911507 | 0.2545928 | -0.055560445 | 0.237023367 | 0.047237036 | 0.214597942 |
| 391 | 1.346808585 | -0.704284514 | -1.522792398 | -0.015248107 | -0.484181147 | -0.21065439 | -0.330925578 | 0.640531426 | 0.472355251 | 0.059527563 | -1.217449682 | -0.698349929 | -0.345851235 | 0.152435161 | -0.148441924 | 0.259890923 | -0.052980227 | 0.262840575 | -0.046120849 | -0.10274659 |
| 442 | 1.768390261 | 0.468666658 | -0.329126664 | -0.028126197 | 0.027404292 | -0.04633104 | 0.494731024 | -0.097157824 | 0.253472241 | 0.323243178 | -0.490959092 | -0.813855055 | -0.153313798 | 0.182242047 | 0.035329914 | -0.019993736 | 0.049247647 | 0.055218161 | -0.169292497 | -0.010765167 |
| 453 | 1.796858662 | 0.228571285 | 0.056935772 | -0.255122422 | 0.32263577 | -0.17913493 | 0.565614993 | -0.136223509 | 0.037931073 | 0.223215415 | -0.21964651 | 0.036337467 | 0.251424728 | 0.019720683 | -0.011384715 | -0.112400067 | 0.129929975 | 0.125581765 | 0.106410009 | 0.001079353 |
| 455 | 1.348693941 | -0.181466488 | -0.333452823 | -1.232044037 | -0.12671307 | -0.10513194 | -0.29715382 | 0.33710041 | -0.104243409 | 0.739009201 | -0.005028882 | -0.559265072 | -0.327895248 | 0.114178608 | 0.285085984 | 0.028276537 | -0.102839836 | -0.148389588 | 0.031544571 | 0.060824786 |
| 461 | 1.570083134 | 0.094459511 | -0.662894242 | -0.840587668 | 0.969136032 | 0.558912461 | 1.425719132 | 0.565247063 | 1.074950154 | 0.965861932 | 0.717071763 | 0.020268268 | 0.265800697 | -0.393859912 | -0.086875315 | 0.379795684 | 0.030442079 | -0.109118136 | 0.118715428 | -0.090796399 |
| 466 | 1.815497556 | 1.896848187 | 0.779983201 | -0.263048492 | 0.865992306 | 0.179850321 | 0.176495599 | -0.310863143 | -0.109930901 | -0.187434981 | -0.525944885 | 0.024797986 | 0.283806143 | -0.001177805 | 0.026258142 | -0.004302238 | 0.243470182 | -0.059502793 | 0.003255574 | 0.073839842 |
| 480 | 1.184923195 | 0.959161343 | -0.515782185 | -0.051113003 | -0.237251567 | 0.163184674 | -0.415853896 | 0.057765263 | -0.263528277 | 0.212875209 | 0.096335816 | -0.085540201 | 0.294538074 | 0.141351887 | -0.069687513 | 0.21199914 | -0.084226828 | -0.103464451 | -0.117597581 | 0.116230842 |
| 1001 | 0.768516425 | -0.125165864 | 1.078604343 | 0.487664124 | -0.369726669 | 0.366736431 | -0.391733657 | -0.475461073 | -0.136094084 | -0.437874343 | -0.587947881 | -0.158406184 | -0.16215552 | 0.117223365 | -0.30469812 | -0.182768185 | 0.072686918 | 0.069711069 | -0.101099783 | -0.078232706 |
| 1002 | 0.259257323 | -1.402603449 | 1.9368409 | -0.467079232 | -0.350772952 | 0.161392071 | -0.027585948 | 0.505585077 | 0.017777499 | 0.02947765 | 0.043106972 | -0.18889099 | 0.208417943 | -0.158267543 | 0.098395198 | 0.384895079 | 0.082122285 | 0.252454951 | -0.159589204 | 0.019392706 |
| 1003 | 0.371221736 | -2.081688874 | 1.015407457 | -0.19916565 | 0.059905712 | -0.51407812 | 0.118407449 | -0.421726689 | -0.513243402 | -0.6681201 | 0.087343295 | -0.569225652 | 0.164137331 | 0.01648259 | -0.13971442 | 0.103770953 | -0.062479775 | -0.008859476 | -0.073931244 | -0.030368599 |
| 1004 | 0.385292313 | -1.546755974 | 1.25747908 | -0.289272928 | -1.159915547 | -0.59868868 | -0.287638676 | 0.081981053 | 0.117284425 | 0.352979105 | 0.40227273 | -0.573844535 | 0.076573334 | -0.429486048 | -0.248658791 | -0.042161433 | -0.071393855 | 0.028377455 | -0.142612393 | -0.007628522 |
| 1005 | -0.006551381 | 0.817653728 | 0.027888499 | 0.365128952 | 0.511778217 | 0.406259962 | -0.029011096 | -0.193008608 | -0.660488462 | 0.624039142 | 0.103758275 | -0.202982133 | -0.102401625 | 0.240208926 | -0.22300391 | 0.000778618 | 0.0445412 | -0.088494363 | -0.073364676 | 0.008616628 |
| 1006 | 0.233069345 | 0.096259056 | 0.246749161 | 0.080375874 | 0.09110071 | 0.567925171 | 0.133447982 | -0.545403395 | 0.711152865 | 0.454378229 | -0.012850997 | -1.189463939 | -0.083941794 | 0.087082079 | -0.549142575 | 0.300099711 | -0.123101352 | -0.022116054 | -0.103024431 | -0.192369945 |
| 1007 | 0.146399017 | -1.400243709 | 0.566941527 | 0.150861708 | -0.262868303 | 1.093827371 | 0.757197356 | 0.443141841 | 0.730890079 | -0.079663143 | 0.453343168 | 0.203640817 | -0.136244807 | 0.274816341 | -0.220050111 | -0.299705952 | 0.074892753 | 0.089892861 | 0.221256004 | -0.05181273 |
| 1008 | -0.199952726 | 0.068477526 | 0.763531212 | -0.102847082 | 0.097857338 | 0.348356617 | -0.428603804 | -0.026638445 | 0.728119447 | 0.082428154 | 0.348841345 | -0.230209438 | -0.110529479 | -0.270715236 | -0.393206752 | -0.089703065 | -0.08365498 | 0.035018608 | 0.147826332 | 0.156569889 |
| 1009 | 0.435698721 | -1.038719468 | 1.13689653 | 0.960148678 | 0.346620985 | 0.381338555 | 0.499268356 | 0.378284577 | 0.600172442 | -0.255479475 | -0.38723852 | -0.093847715 | -0.06276144 | 0.063731323 | 0.040012589 | -0.355740781 | -0.113469147 | 0.207389365 | 0.104880531 | 0.063157436 |
| 1010 | -0.812612557 | -0.845590372 | 1.278234045 | 0.282405313 | 0.521513148 | 0.810885006 | -0.724781483 | 0.190420881 | -0.186891062 | 0.212257545 | -0.004964458 | -0.408707064 | 0.07390435 | 0.411970788 | 0.273156152 | 0.21075391 | -0.070821603 | -0.10247446 | -0.045364242 | 0.106822586 |
| 1011 | 0.919257807 | -0.122868707 | -0.342829118 | -0.236137213 | -0.890130331 | -0.763245329 | 0.301474252 | 0.309816462 | -0.200738202 | -0.204643546 | 0.178297937 | 0.068281827 | -0.181504505 | -0.142568232 | -0.042466533 | 0.083377705 | 0.134260251 | 0.061041053 | 0.086795454 | -0.023999851 |
| 1012 | -0.2008741 | -0.246583095 | -0.098378217 | -0.155533589 | -0.143812283 | 0.411739073 | 0.031758434 | -0.007246382 | -0.151641702 | -0.248224695 | 0.100513776 | -0.227042936 | 0.144507 | 0.063565401 | -0.065060245 | 0.387936786 | -0.106072843 | 0.082867677 | -0.008836098 | 0.138296898 |
| 1013 | 0.908118499 | 0.090650313 | 0.33447085 | -0.58270815 | -0.614870236 | -0.436212296 | 0.221553587 | 0.551129847 | -0.458163106 | -0.38056341 | 0.07272977 | 0.142732753 | -0.19412053 | 0.007816877 | -0.330747376 | 0.285531951 | -0.019972198 | -0.07020521 | -0.005863778 | -0.033118544 |
| 1014 | -0.688004177 | -0.489247619 | 0.51627391 | 0.317008865 | -0.058234265 | 0.748707689 | 0.195308932 | 0.317715284 | 0.269419248 | -0.024252928 | 0.176938831 | -0.098996476 | -0.003819562 | 0.015497002 | 0.46157901 | 0.050969664 | 0.086142072 | 0.143527189 | 0.037297513 | 0.145879207 |
| 1015 | -0.389814974 | -1.244712495 | 1.383667831 | -0.921187647 | 0.574249058 | 0.156351266 | -0.629384616 | 0.372843536 | 0.144789152 | -0.575256106 | 0.031800471 | -0.206363036 | -0.152378896 | -0.033678102 | -0.331504888 | -0.280115148 | 0.260462087 | -0.049855625 | -0.088029104 | 0.0906624 |
| 1016 | 0.167356786 | -0.149577545 | 0.817908828 | 0.344860424 | 0.115937328 | 1.16284771 | -0.033295785 | 0.204083135 | 0.323815541 | 0.168276033 | 0.032836806 | 0.10027589 | -0.329856761 | -0.063955658 | 0.322639961 | -0.016601125 | 0.08002069 | 0.066224233 | -0.097948589 | 0.055344473 |
| 1017 | 0.268086295 | 0.414714163 | -0.084390445 | 0.004820203 | 0.300868995 | 1.347401093 | -1.020865606 | -0.38558397 | -0.264172152 | 0.170592186 | 0.371387248 | 0.067808059 | -0.132289734 | -0.056935152 | -0.416842888 | -0.114544768 | -0.251971981 | 0.137985347 | -0.098926026 | 0.103286568 |
| 1018 | 0.183426523 | -0.705533201 | 0.784413704 | 0.204361273 | -0.003768605 | 0.828944898 | 0.015072345 | 0.450391783 | 0.78774907 | -0.038704271 | -0.031800708 | -0.123567253 | 0.236608964 | -0.038240091 | 0.136813301 | 0.390186052 | -0.042515842 | 0.12186455 | 0.008066576 | 0.045008935 |
| 1019 | 0.456159882 | -0.964581674 | 0.14393025 | -0.043547889 | -0.206988436 | -0.484904707 | 0.561273939 | -0.043667699 | -0.248002799 | 0.075242872 | -0.149999137 | 0.396924795 | -0.241896518 | -0.128417645 | 0.014052876 | -0.094017792 | -0.05566503 | 0.212526664 | -0.011672556 | 0.080743841 |
| 1020 | 0.282965608 | 1.116155658 | 0.130519497 | 0.242655783 | 0.363015884 | 0.182567081 | -0.14819864 | -0.168423842 | 0.012386229 | -0.619983914 | -0.106506885 | 0.130839924 | -0.117466442 | -0.046539315 | 0.046554377 | 0.196076671 | 0.030911169 | 0.007467723 | -0.062633283 | 0.155848955 |
| 1021 | 0.454588452 | 0.067045438 | -0.423672281 | -0.265495433 | 0.140982746 | 0.248288975 | 0.305142731 | 0.019708447 | 0.134347953 | 0.105623452 | -0.314490345 | -0.970945737 | -0.035884252 | 0.199635779 | 0.080475368 | 0.238983953 | 0.023908937 | -0.06446892 | -0.118618383 | -0.140336685 |
| 1022 | -0.267960407 | 0.419251962 | -0.420450557 | 0.325005978 | -0.168250827 | 0.011213037 | -0.205243854 | 0.020664465 | -0.047516159 | -0.030926775 | 0.154660009 | 0.030973984 | 0.076049188 | 0.126618153 | -0.011187393 | -0.047065027 | -0.166836183 | -0.078053722 | -0.028187826 | -0.003087397 |
| 1023 | 0.474814001 | 0.726161184 | -0.761682574 | 0.207652324 | 0.326763503 | 0.158961789 | -0.253446547 | -0.220392003 | -0.694834676 | 0.012355927 | 0.070297453 | 0.02912883 | -0.013642381 | -0.034401716 | -0.112208016 | -0.066198355 | 0.151791553 | 0.128232747 | -0.115649728 | -0.002232134 |
| 1024 | -0.186144411 | 1.6138417 | -0.159183674 | 0.746126423 | 0.501847535 | -0.147767931 | -0.122238593 | -0.2278128 | -0.281659267 | 0.036518997 | 0.153888501 | -0.390698438 | -0.053307861 | 0.17739542 | -0.022054988 | 0.178165694 | -0.058676097 | -0.008612649 | 0.111539169 | 0.080829023 |
| 1025 | 0.175030304 | 0.944766516 | 0.040016506 | 0.172386019 | 0.310162668 | 0.026366767 | -0.268025684 | -0.046749956 | -0.11750679 | -0.481122708 | -0.098697859 | 0.06739452 | -0.057035407 | 0.08699722 | 0.005443873 | -0.219823345 | 0.019366932 | -0.000994944 | -0.063773803 | 0.009688203 |
| 1026 | 0.149805669 | -0.181105608 | -0.246192047 | -0.279136327 | -0.238174068 | -0.232628296 | -0.361141211 | 0.276507411 | -0.04792478 | -0.123482605 | 0.207446478 | 0.080037668 | 0.124559069 | -0.042486305 | 0.003847746 | -0.117273259 | 0.034350871 | 0.157617746 | -0.109168149 | 0.013335033 |
| 1027 | -0.704395719 | 0.341527304 | 0.721538569 | -0.232558215 | 0.321728869 | 0.26387479 | 0.648237876 | -0.310321232 | 0.033797804 | 0.212090748 | -0.465171024 | 0.625286458 | -0.267903356 | 0.20029955 | -0.051347057 | -0.108174451 | -0.493407157 | -0.207957466 | -0.157281971 | 0.050502462 |
| 1028 | -0.102684258 | -0.078137312 | -0.008453167 | -0.074849002 | 0.098581186 | 0.007229104 | -0.066886246 | 0.02194416 | 0.22792372 | -0.090588507 | 0.087300166 | -0.047668254 | 0.206224782 | 0.011723667 | 0.041147088 | -0.158277721 | 0.046156621 | 0.109748051 | 0.089991514 | -0.017955584 |
| 1029 | 0.021989958 | -0.229178767 | 0.165650782 | -0.234251127 | 0.197692101 | -0.256401563 | -0.121382188 | 0.218702165 | 0.060276443 | 0.141640045 | 0.030838099 | 0.081108981 | 0.016307728 | -0.00898612 | 0.051512656 | -0.052333948 | -0.013108215 | 0.066750009 | 0.150566489 | -0.015354228 |
| 1030 | -0.077705937 | -1.007915894 | 0.307638632 | -0.131277761 | -0.145439872 | -0.333156437 | 0.852736323 | -0.21102986 | -0.247119494 | 0.188637976 | -0.406257706 | 0.509350652 | -0.111691735 | -0.001442164 | -0.09364463 | 0.099006927 | -0.177921043 | 0.0462345 | 0.062272679 | -0.082347043 |
| 1031 | -0.009795342 | 0.009075368 | 0.127989812 | -0.127337028 | 0.183877058 | -0.187338691 | -0.035812809 | 0.124709962 | 0.099376997 | -0.009950756 | 0.068036886 | 0.081451608 | -0.067833936 | 0.091462128 | 0.058490273 | -0.295811851 | 0.057522497 | 0.009066664 | 0.024831696 | -0.09814904 |
| 1032 | -0.094889523 | 0.088511798 | -0.038984865 | -0.13430471 | 0.114258899 | -0.128347816 | -0.494007371 | 0.222330191 | 0.058832762 | -0.183910366 | -0.32190027 | 0.138215241 | 0.032792912 | 0.090690211 | -0.019455902 | -0.134150415 | 0.081138187 | -0.044925789 | 0.144062442 | -0.020312726 |
| 1033 | -0.003282712 | 0.953837393 | -0.265528361 | 0.252954784 | 0.158374405 | 0.068634739 | -0.387851554 | -0.109043437 | -0.378020883 | 0.295565526 | 0.269603634 | -0.080015487 | -0.136885216 | 0.080585032 | -0.068511123 | 0.007645152 | 0.123943303 | 0.102754812 | -0.056397827 | 0.075661128 |
| 1034 | 0.095325214 | 0.006463168 | -0.550404585 | 0.240873586 | -0.352605568 | -0.466829437 | -0.33834435 | 0.232321083 | -0.044846621 | -0.060546198 | 0.172565036 | -0.211183863 | 0.211085046 | 0.043958809 | 0.008723813 | 0.080893881 | 0.021060463 | 0.032311024 | 0.07650181 | -0.009430505 |
| 1035 | -0.131216107 | 0.277218308 | -0.582801451 | 0.061946948 | -0.062374454 | -0.038068085 | -0.598659777 | 0.112891176 | 0.030623836 | -0.100635619 | -0.113338242 | 0.045280279 | 0.181721313 | 0.123620241 | -0.076322035 | 0.104212013 | 0.078546385 | -0.078651219 | 0.067747888 | 0.021046132 |
| 1036 | -0.046959164 | -0.39433002 | 0.480476883 | -0.304756 | -0.221371782 | -0.493674724 | 0.288951518 | -0.238149292 | -0.20055292 | 0.079015694 | -0.201296426 | 0.209250993 | -0.0417379 | -0.027443872 | 0.007242994 | 0.177197036 | 0.110356846 | 0.210732581 | 0.169283746 | -0.018358711 |
| 1037 | 0.025211794 | -0.118199236 | -0.636773514 | 0.09495837 | -0.540786957 | 0.049814172 | -0.244896927 | 0.23942376 | -0.214891784 | 0.226733896 | 0.450893067 | 0.00409956 | -0.095110295 | -0.029740532 | 0.140328105 | -0.27348675 | -0.08321153 | -0.120086126 | -0.115484181 | -0.08868279 |
| 1038 | -0.133166959 | 0.312159102 | 0.148668089 | -0.044239892 | -0.046896588 | 0.053757907 | 0.296885951 | 0.019783828 | 0.227622785 | 0.380588698 | -0.268969749 | 0.324472581 | 0.031430328 | 0.107896156 | -0.116234013 | -0.072703423 | -0.233425173 | -0.053683722 | 0.04328701 | -0.00529602 |
| 1039 | -0.050576667 | 1.600233388 | 0.304182411 | 0.332003757 | 0.51249897 | 0.123557168 | -0.256864669 | -0.092186266 | -0.001309997 | -0.097920909 | -0.087868979 | -0.442759298 | -0.052457827 | 0.236569602 | 0.019063762 | -0.081033834 | -0.055598411 | -0.014872246 | 0.118439001 | 0.016560043 |
| 1040 | 0.038425071 | -0.908519344 | -0.035455828 | 0.087748419 | -0.256964509 | -0.343501386 | 0.441704662 | -0.117658281 | -0.339753993 | 0.145543582 | -0.042127138 | 0.115852597 | -0.30994289 | 0.102025031 | 0.122840996 | -0.083297638 | -0.168456458 | -0.024610666 | 0.022346023 | -0.075267206 |
| 1041 | 0.116909893 | -0.334284564 | -0.659493896 | 0.369391249 | -0.324920332 | -0.146858613 | -0.049360464 | 0.024939058 | -0.281822843 | -0.074513668 | 0.017207598 | -0.05325817 | -0.071805872 | -0.025709367 | 0.023079183 | 0.097183695 | 0.052128136 | -0.033716708 | 0.036671028 | -0.011772079 |
| 1042 | -0.097566569 | -0.146729135 | -0.778946483 | 0.675421658 | 0.011118973 | -0.152787316 | 0.086196644 | -0.18766518 | 0.16767326 | -0.525853944 | -0.291010663 | 0.112918041 | 0.087336365 | 0.053829875 | -0.043821413 | 0.198782259 | 0.00793819 | -0.084692013 | 0.022872658 | 0.027788577 |
| 1043 | -0.063187117 | -0.025008776 | -0.680042853 | 0.471729106 | -0.117372996 | -0.336068231 | -0.173499521 | -0.156174028 | -0.299051165 | -0.157880312 | 0.165331273 | -0.151360094 | 0.025960899 | -0.035367677 | 0.030466238 | 0.219329375 | 0.067781431 | 0.115240286 | 0.018052354 | 0.083279705 |
| 1044 | 0.095791195 | 0.334367942 | -0.271851084 | -0.046142364 | 0.052878188 | -7.28E-02 | -0.529326378 | 0.281775001 | -0.267854597 | 0.278898366 | -0.041424993 | -0.069815823 | -0.080695344 | 0.132108381 | -0.014358149 | -0.019855321 | -0.105681003 | -0.121972663 | 0.058554455 | 0.04590029 |
| 1045 | -0.204373583 | -0.386372952 | 0.08023764 | 0.171664004 | -0.072179535 | -0.18667926 | 0.517405739 | -0.088557921 | 0.160483317 | 0.278673858 | -0.052771591 | 0.049195957 | -0.083170411 | -0.029905909 | 0.218101776 | -0.673675836 | 0.088916815 | 0.062140493 | 0.147630864 | -0.068595327 |
| 2001 | 0.779571087 | 0.406285805 | 1.834245112 | -0.596453271 | 0.374308439 | 1.192810746 | -0.269120262 | 0.561433892 | 0.149051849 | 0.010377371 | -0.05376368 | 0.3551061 | -0.000570923 | 0.127776581 | 0.227861239 | 0.350153652 | -0.123375815 | -0.066875701 | -0.04040393 | -0.072279379 |
| 2002 | 1.281405408 | 0.421469463 | 0.629328831 | -0.609799911 | 0.481996516 | 0.689530413 | 0.533808316 | 0.25667118 | 0.628132852 | -0.679253727 | 0.708969673 | 0.045751598 | 0.123222845 | -0.023614191 | -0.3100336 | 0.299773074 | 0.164444164 | -0.080230325 | 0.16177684 | 0.038706054 |
| 2003 | 1.299380306 | -0.136299376 | 1.417470965 | -1.201769399 | -0.2551346 | -0.046995951 | 0.538547812 | 0.293143909 | -0.250577536 | 0.397009603 | 0.50630772 | 0.403365503 | -0.036008903 | 0.300757224 | -0.533858234 | -0.013027631 | -0.01808311 | -0.001668743 | -0.086308752 | -0.099071237 |
| 2004 | 1.395874473 | 1.610606779 | 0.167263816 | -0.079045762 | 0.166860138 | 0.7219042 | -0.529555243 | 0.097243668 | -0.1089355 | -0.892160031 | 0.27311201 | -0.16661625 | -0.202213121 | -0.392654159 | 0.256538417 | -0.093243187 | -0.189289728 | 0.055020336 | 0.045580724 | -0.216782956 |
| 2005 | 1.533118691 | 0.288972014 | -0.162776413 | -0.971720865 | -0.304464681 | 0.679074281 | 0.524073017 | 0.229605674 | 0.277301722 | -0.667697845 | 0.088559137 | 0.083404402 | -0.146958527 | 0.184490812 | -0.180346502 | -0.079618652 | -0.274553613 | -0.100932261 | 0.058435464 | 0.100801759 |
| 2006 | 1.571812674 | 0.710123079 | 0.311296317 | -1.155088415 | 0.335076462 | 0.614894944 | 0.145495869 | 0.327444182 | 0.40144333 | -0.255585809 | -0.178210169 | -0.513328605 | -0.272653931 | 0.078605264 | 0.100014594 | 0.077494005 | -0.012923126 | 0.130348071 | -0.077271529 | -0.116599545 |
| 2007 | 1.422658536 | 1.152115 | -0.174645101 | -0.366657937 | 0.056632456 | 0.499127056 | -0.939027556 | 0.231778418 | -0.357249018 | -0.761185044 | 0.232103682 | -0.14228861 | -0.486410096 | -0.45549312 | 0.408847266 | 0.064376365 | -0.132958738 | 0.094605617 | 0.102089707 | -0.304623605 |
| 2008 | 1.815197538 | -0.318071393 | -0.032938302 | -0.974765792 | -0.071405476 | 0.485571764 | 0.210733693 | 0.113641442 | -0.0109485 | -0.184718471 | 0.866649122 | 0.158350158 | -0.030162963 | 0.302944005 | -0.158423431 | -0.272430369 | 0.342849881 | -0.017801471 | -0.062412143 | 0.222437165 |
| 2009 | 1.339787424 | 0.961449041 | -0.126612398 | -0.682140245 | -0.258885178 | 0.422407714 | -0.419633958 | 0.032457006 | 0.291639485 | -0.699958396 | 0.075743952 | 0.228740223 | -0.137559194 | -0.099140157 | 0.155473934 | -0.220892196 | -0.081185913 | 0.086392926 | 0.008178313 | -0.365441485 |
| 2010 | 1.446790417 | 1.703563243 | 0.122660369 | -0.087451953 | 0.213077379 | 0.852262588 | -0.427101118 | 0.134137943 | -0.035134778 | -0.849958523 | 0.159668167 | -0.287466606 | -0.283202521 | -0.346108242 | 0.261552781 | -0.11773382 | -0.184749732 | 0.030427562 | 0.044456008 | -0.145752065 |
| 2011 | 1.296353968 | 0.066445704 | -0.02718488 | -1.175207432 | -0.413351248 | 0.596635572 | 0.211042821 | 0.290386523 | 0.532183892 | -0.424838541 | 0.05536111 | 0.126873515 | 0.243596715 | 0.50708329 | -0.299399337 | -0.192929951 | 0.149530274 | -0.052616042 | -0.091628701 | 0.113067488 |
