## Supplementary material for "Distinctive features of lipoprotein profiles in stroke patients": Fig. S1: index.htm

Support

  

### S1, 2 Fig: PCA

### S3, 4 Fig: box plots

### S5 Fig: Age of sample

### S6 Fig: Contribution of PCA

| TG and Cholesterol | Current Methods |
| --- | --- |

### S1 Table: fitting data

### S2 Table: PC for the samples

### S3 Table: PC for the items
