## Supplementary figures and images for "Distinctive features of lipoprotein profiles in stroke patients"

### age.png

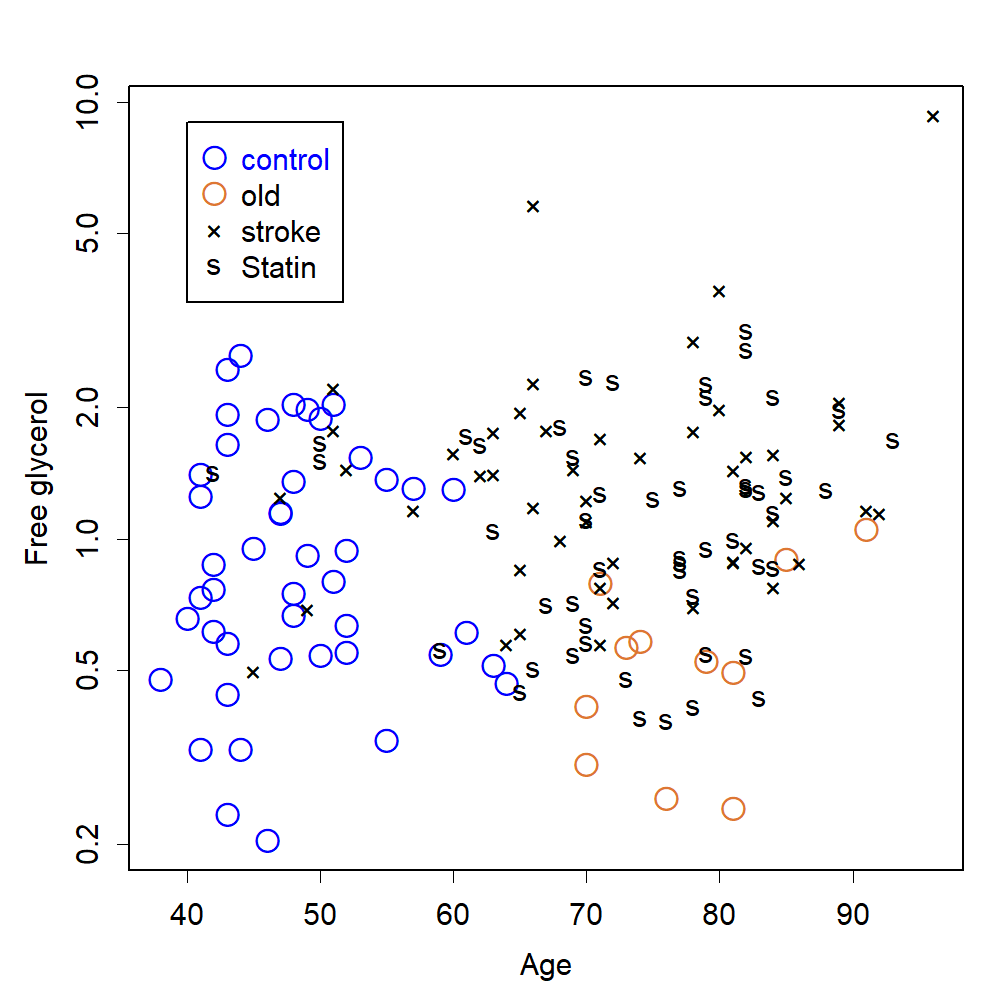

### agebox.png

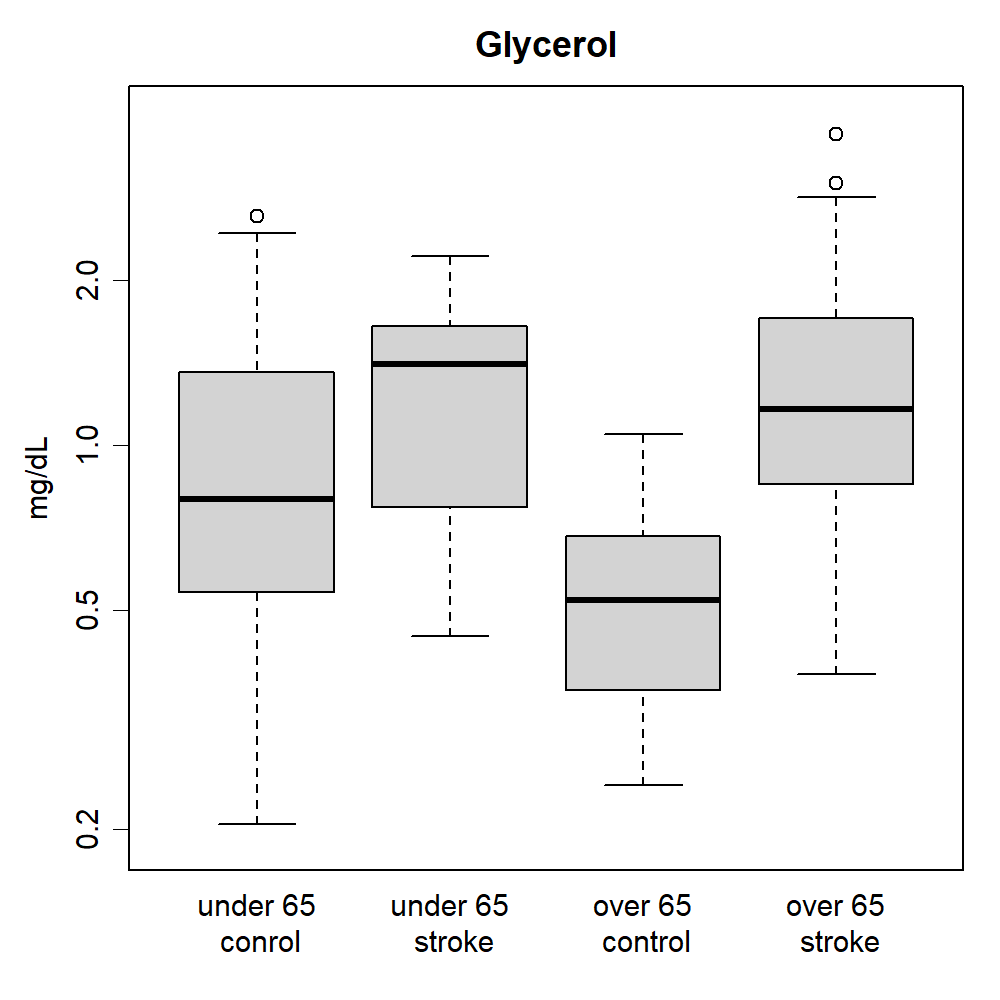

### Box2HDL.png

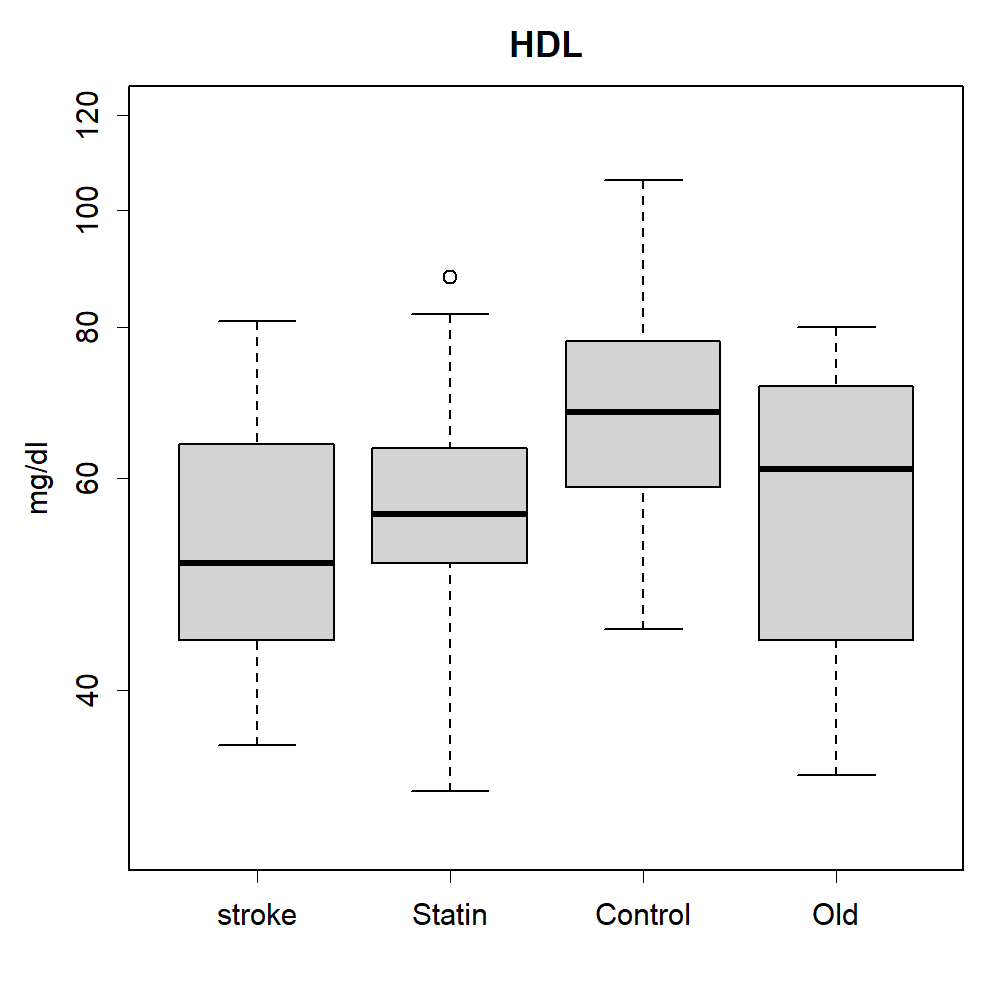

### Box2LDL.png

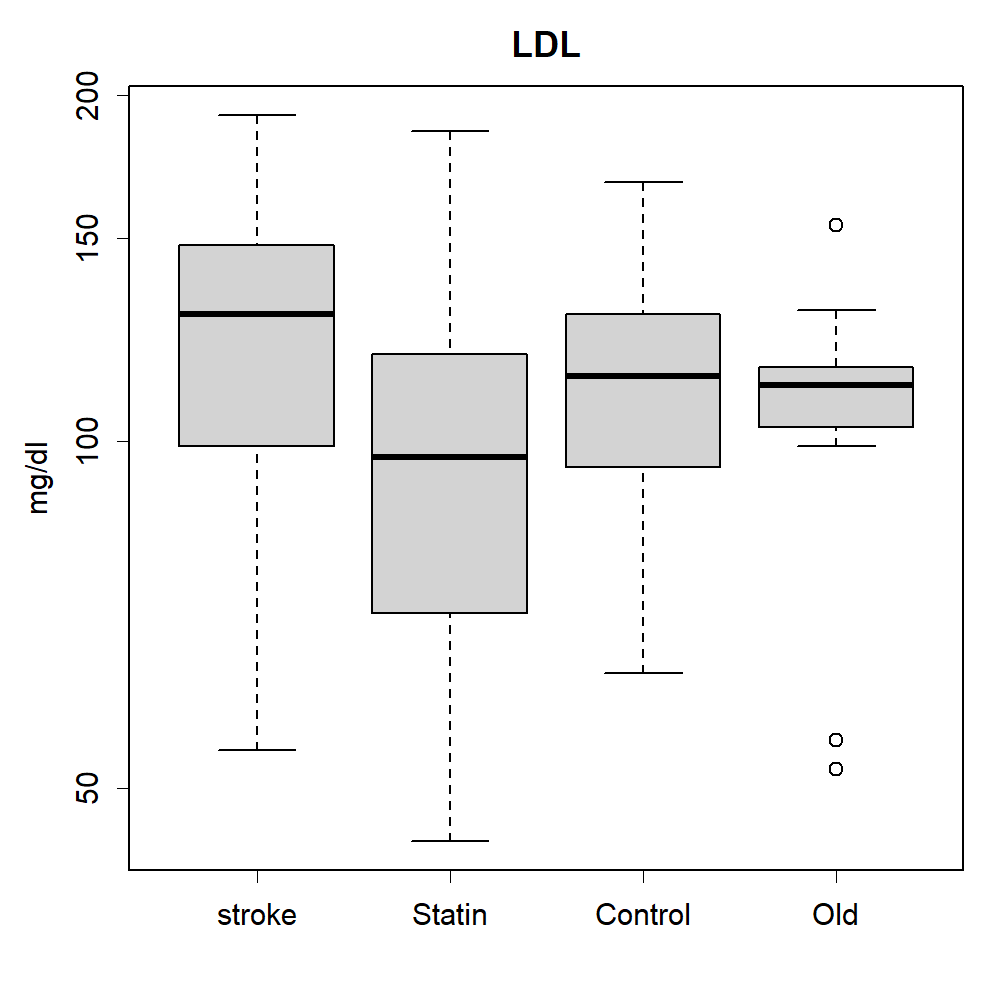

### Box2Tch.png

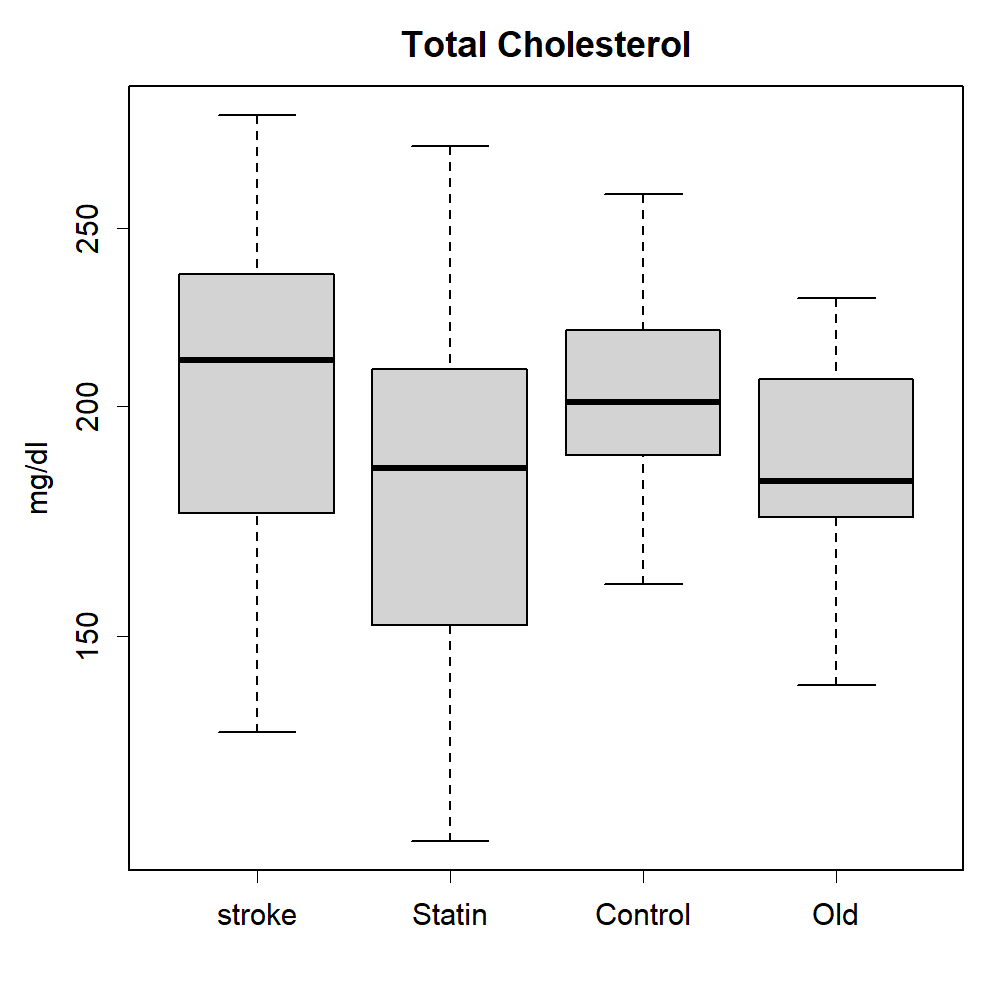

### Box2TG.png

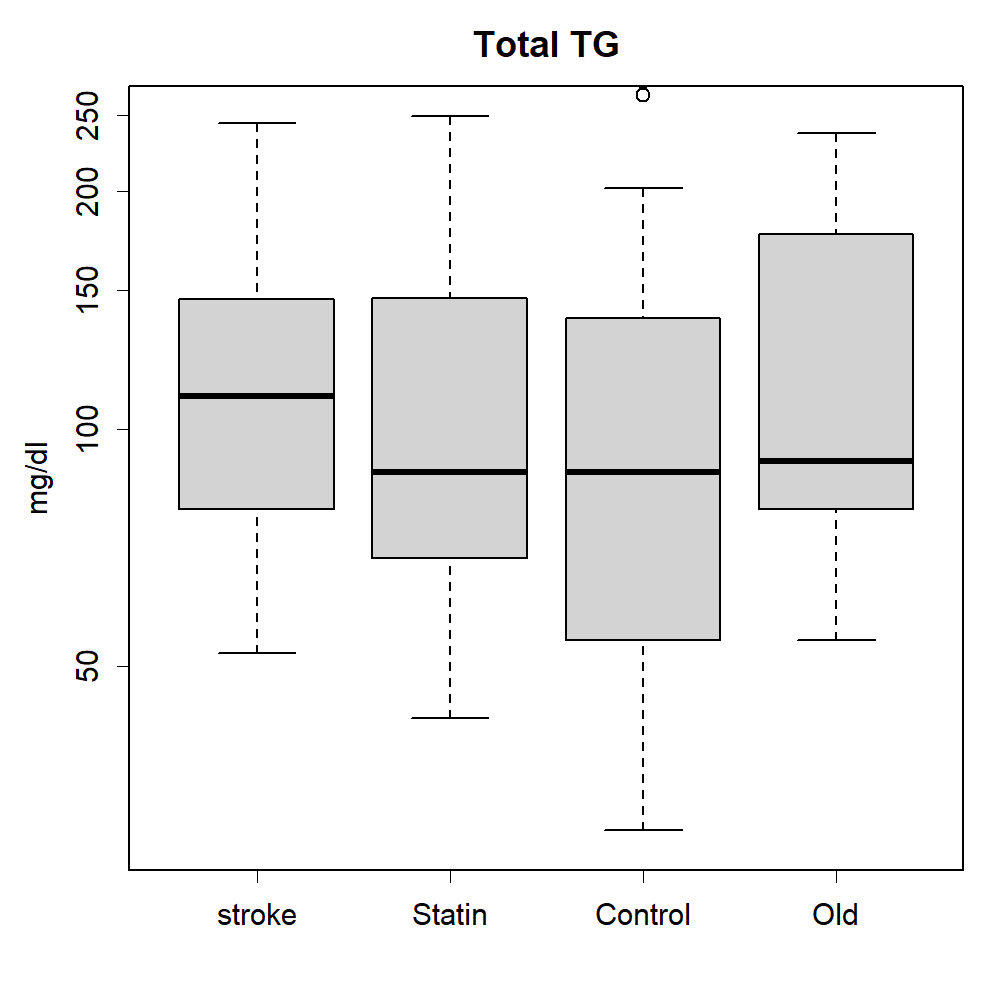

### CH_TGbox.png

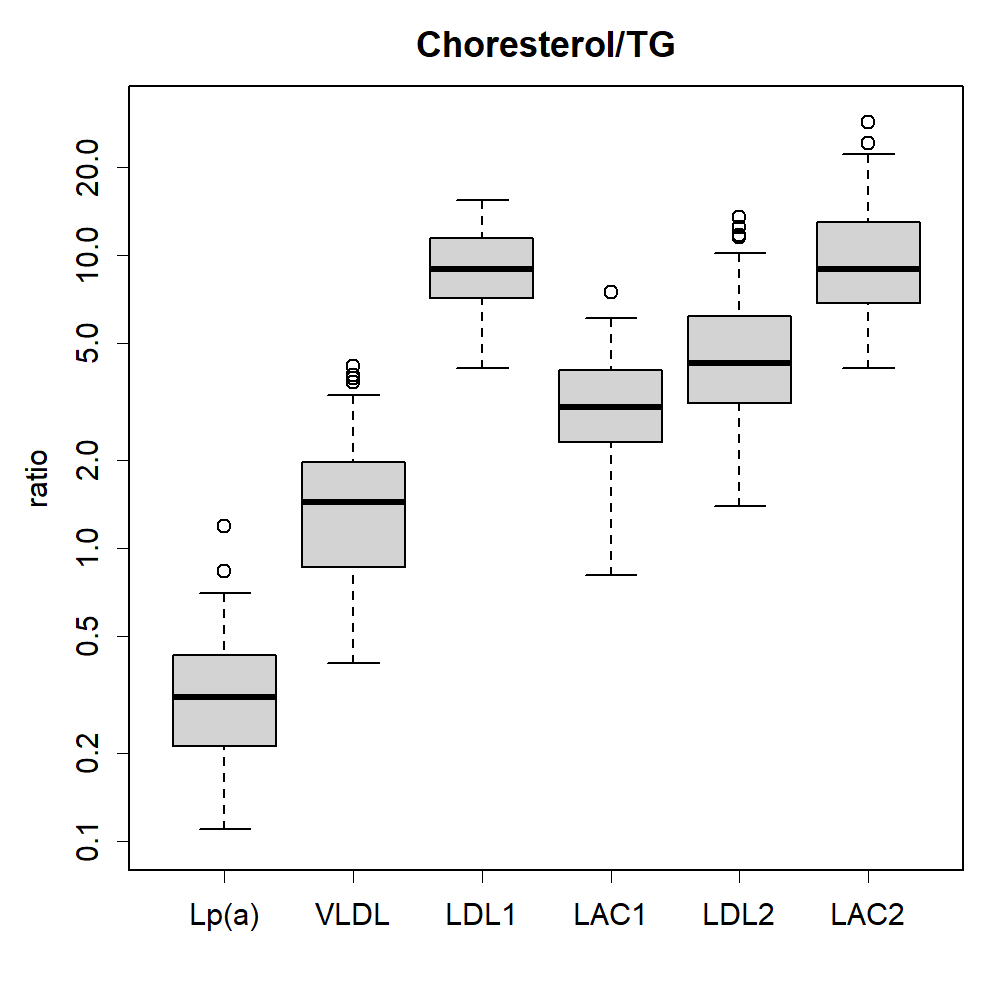

### CM1.png

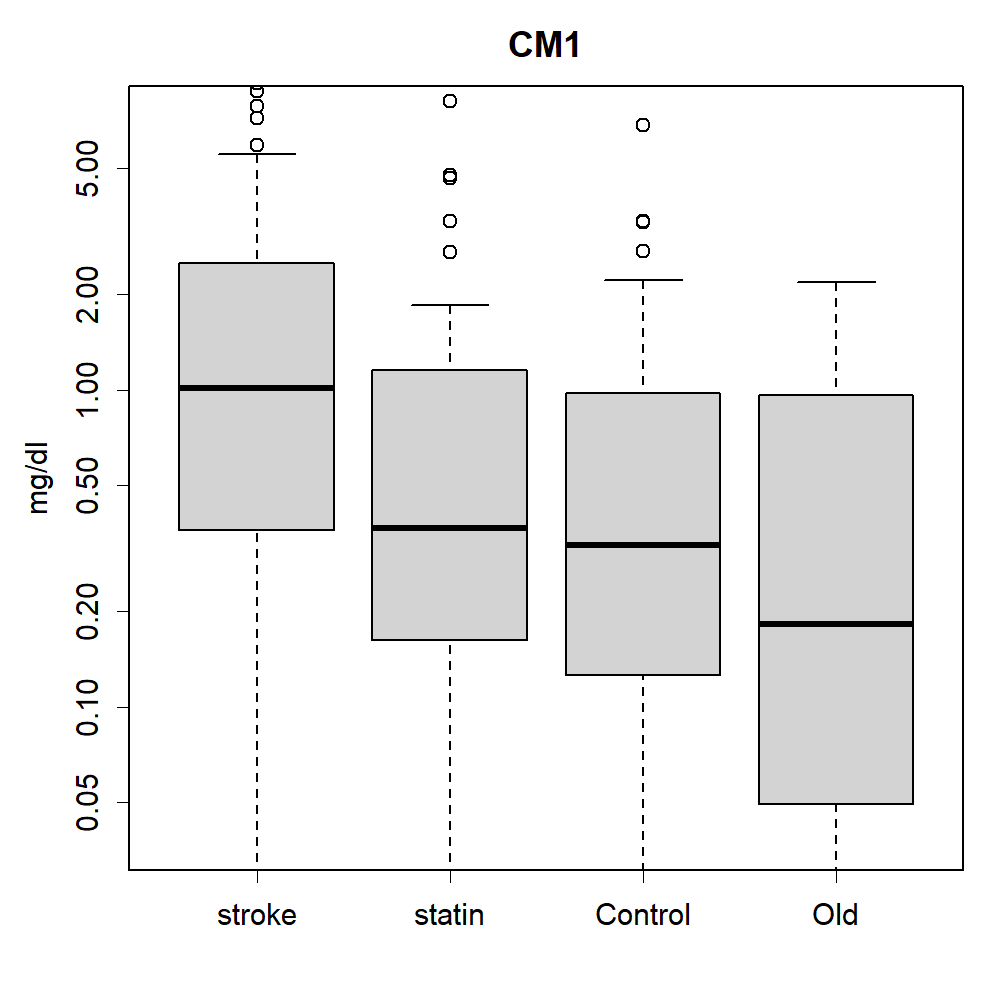

### CM1.png

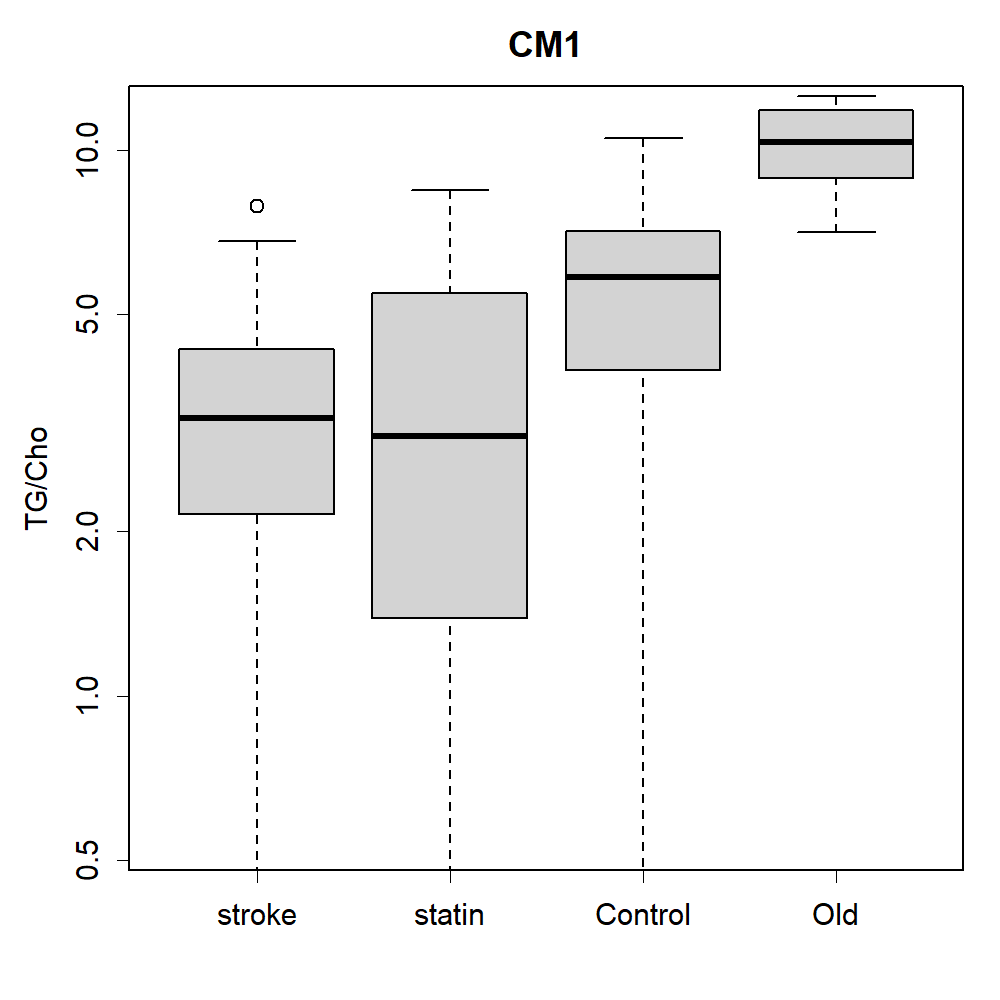

### CM1.png

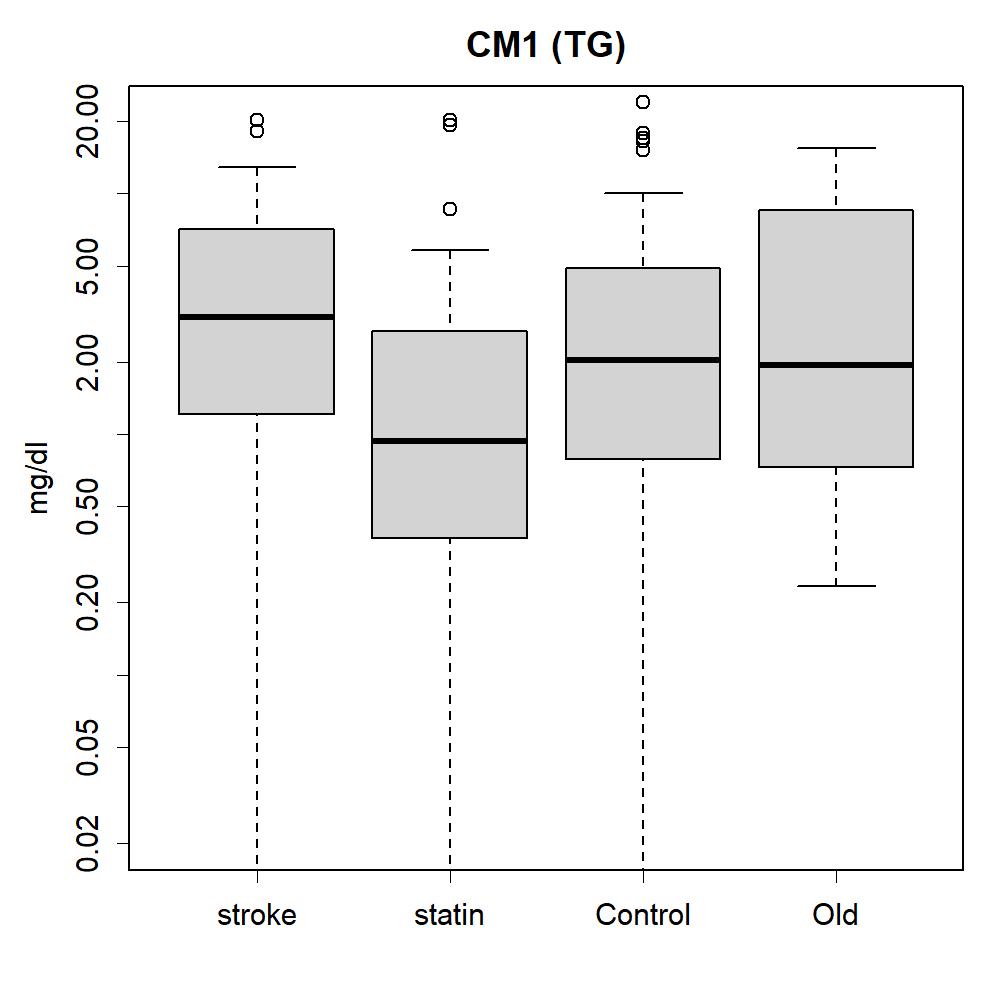

### CM2.png

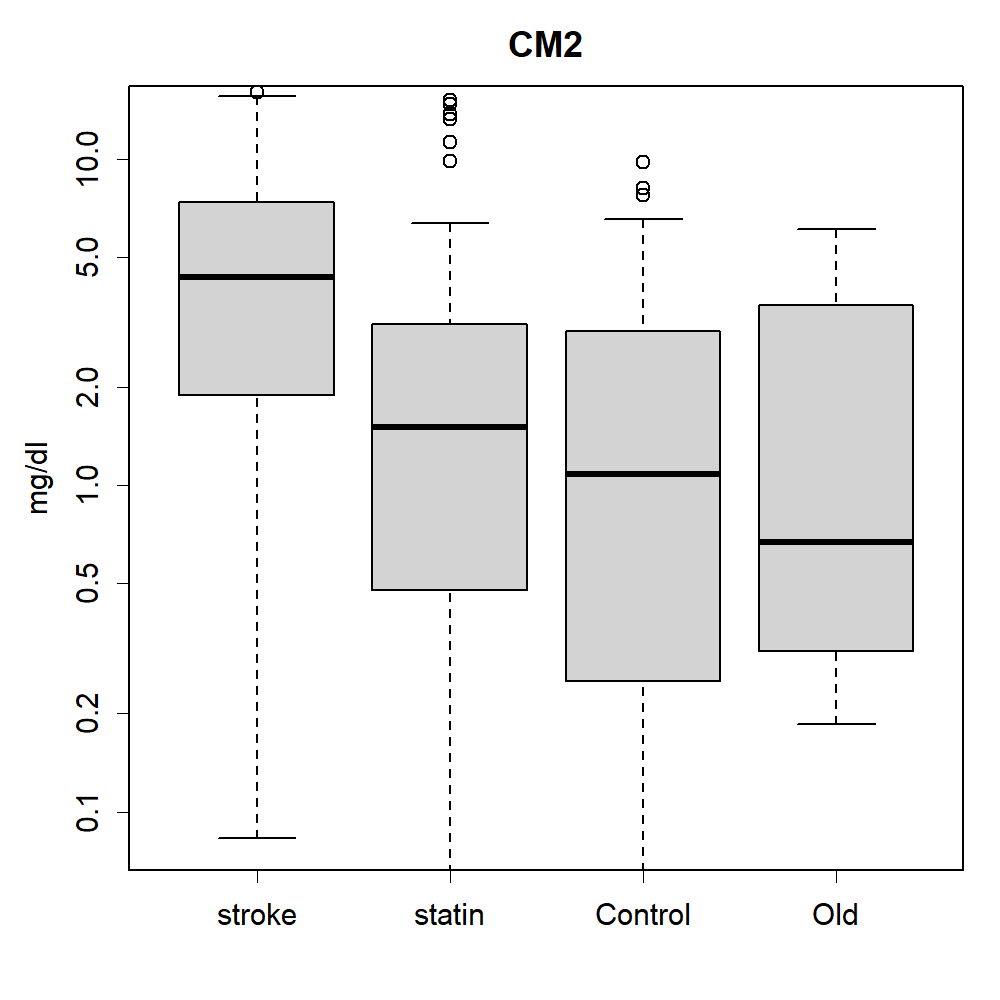

### CM2.png

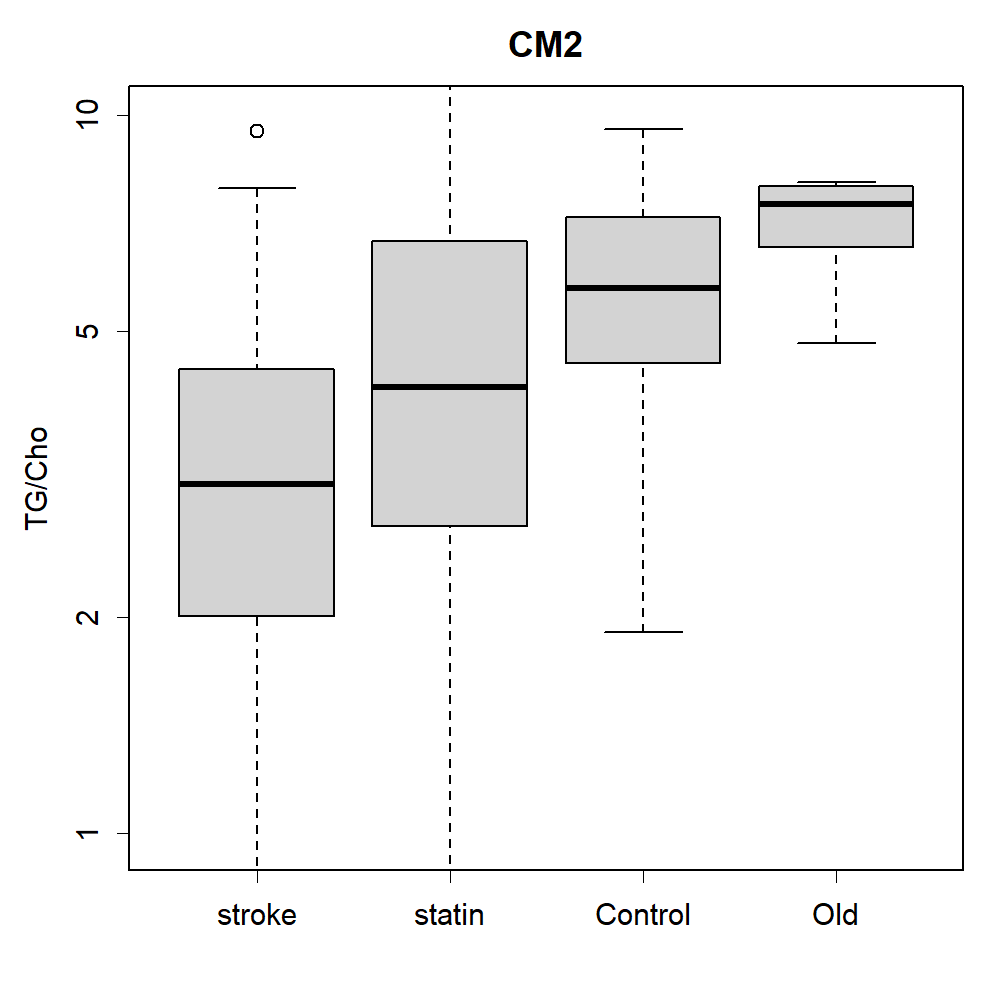

### CM2.png

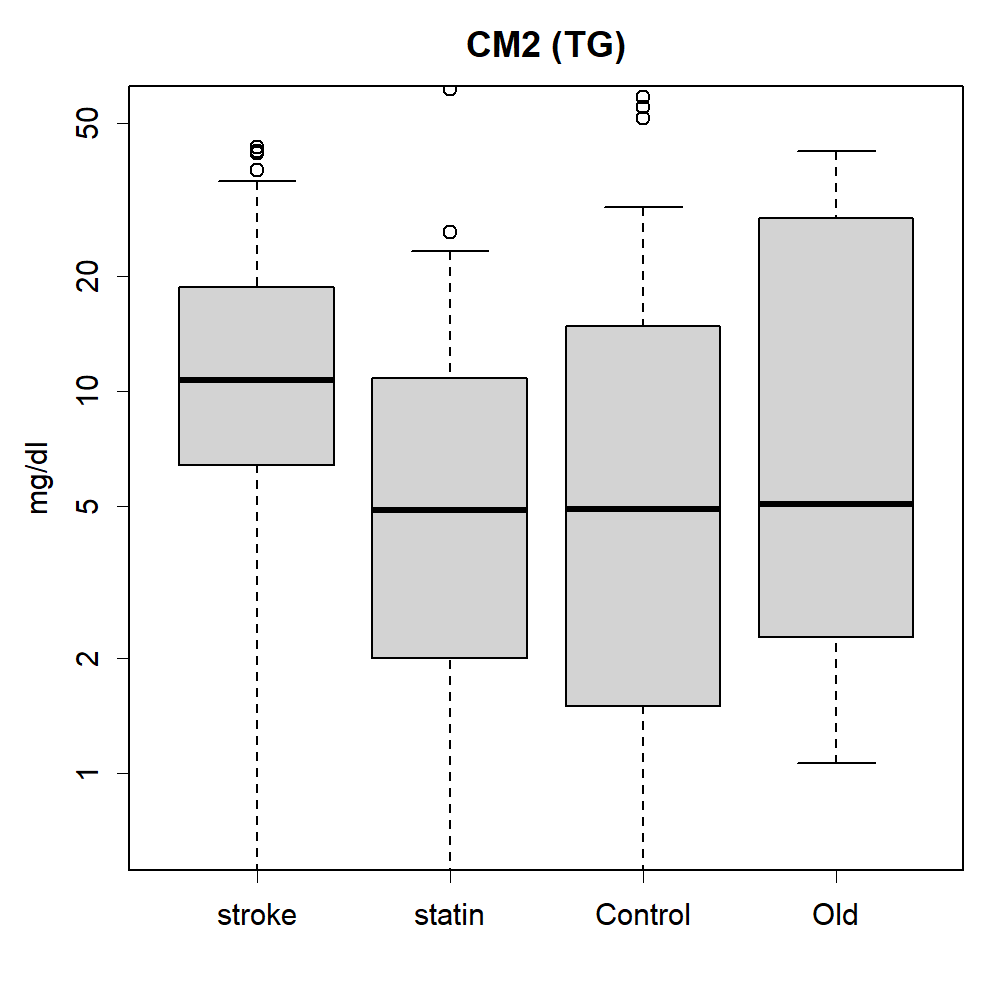

### contribution2.png

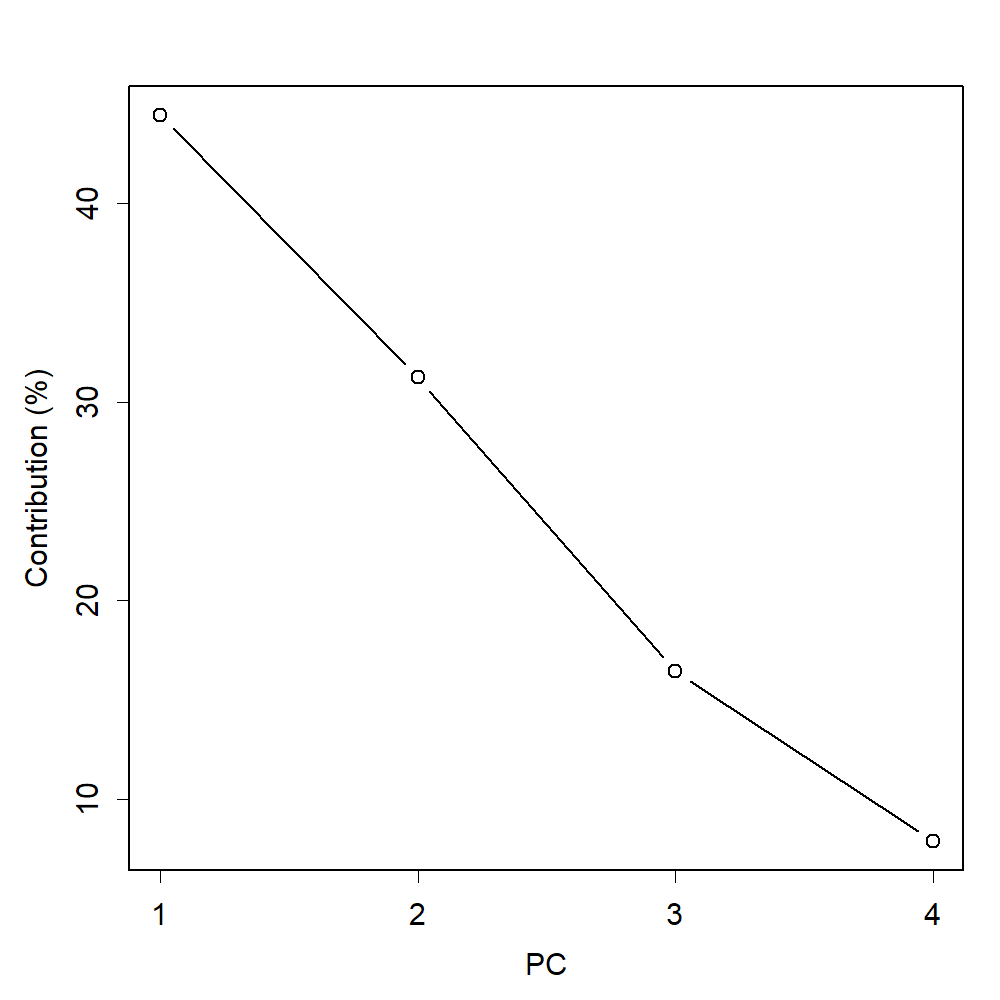

### contribution3.png

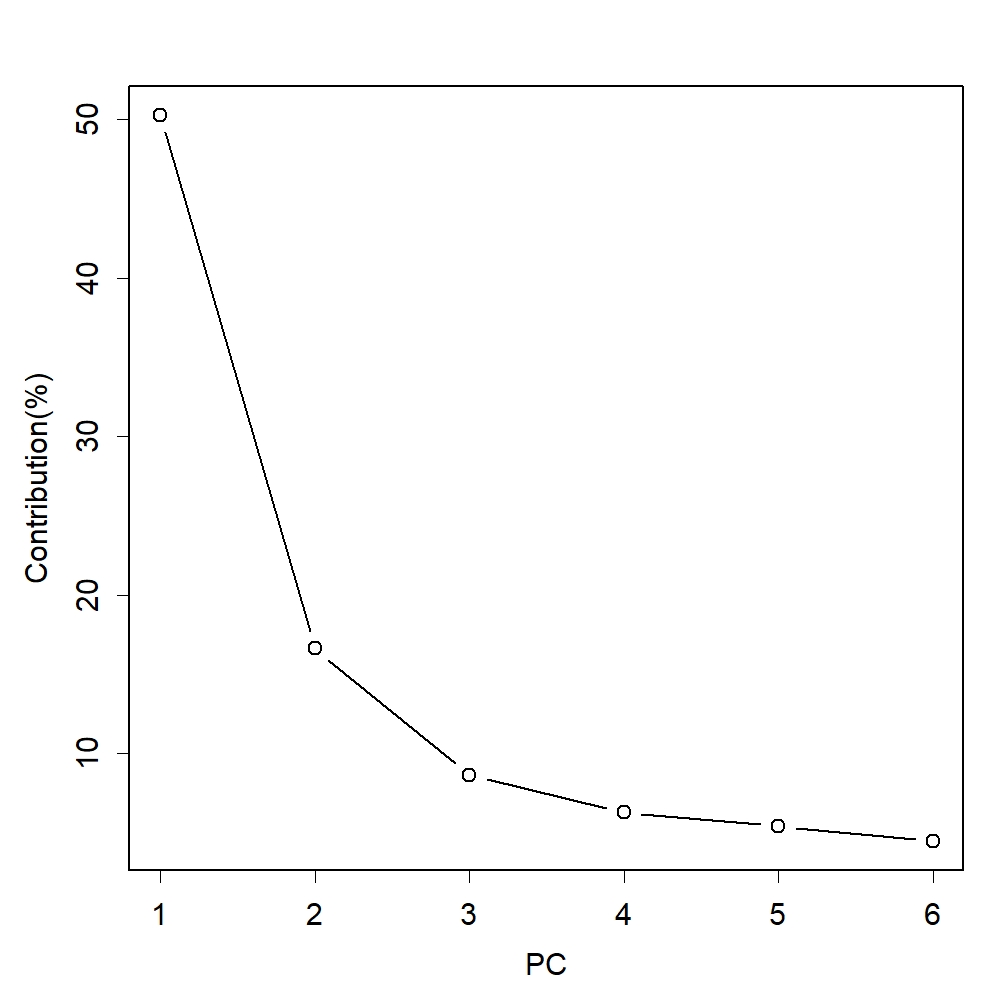

### contribution.png

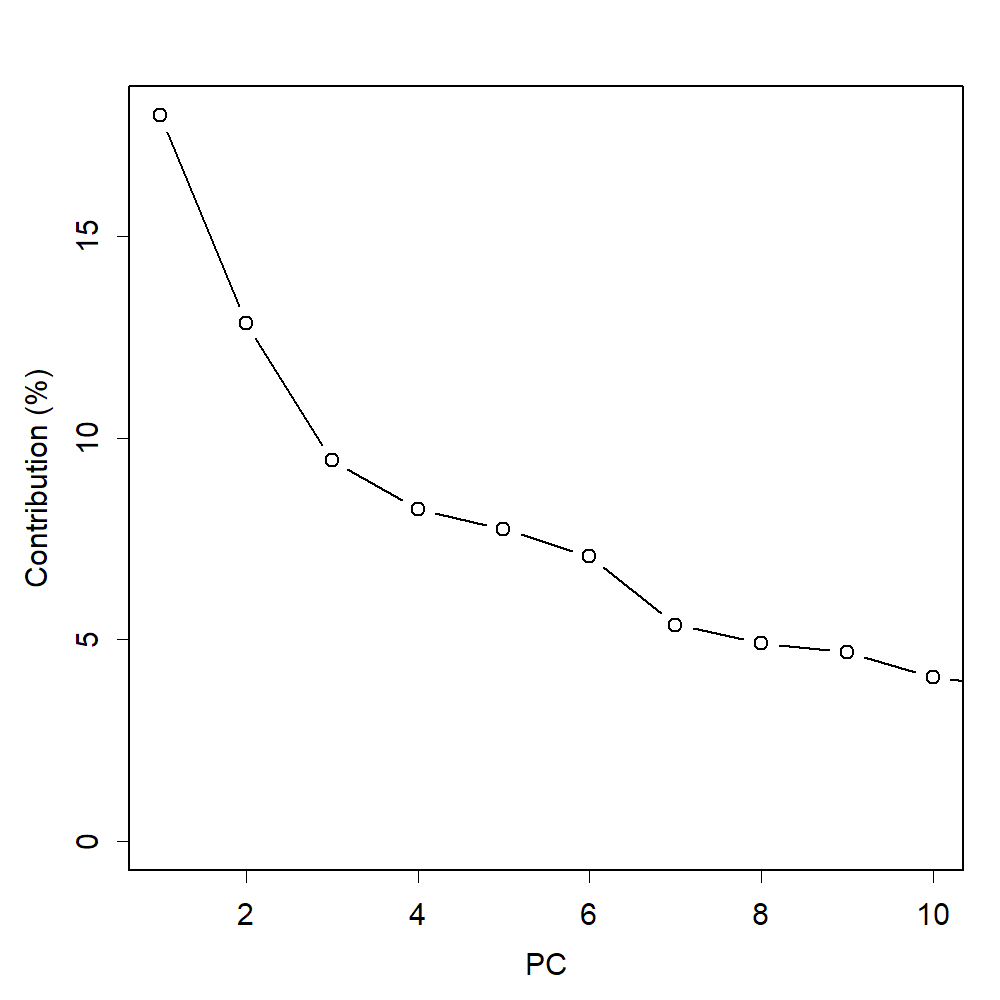

### gly.png

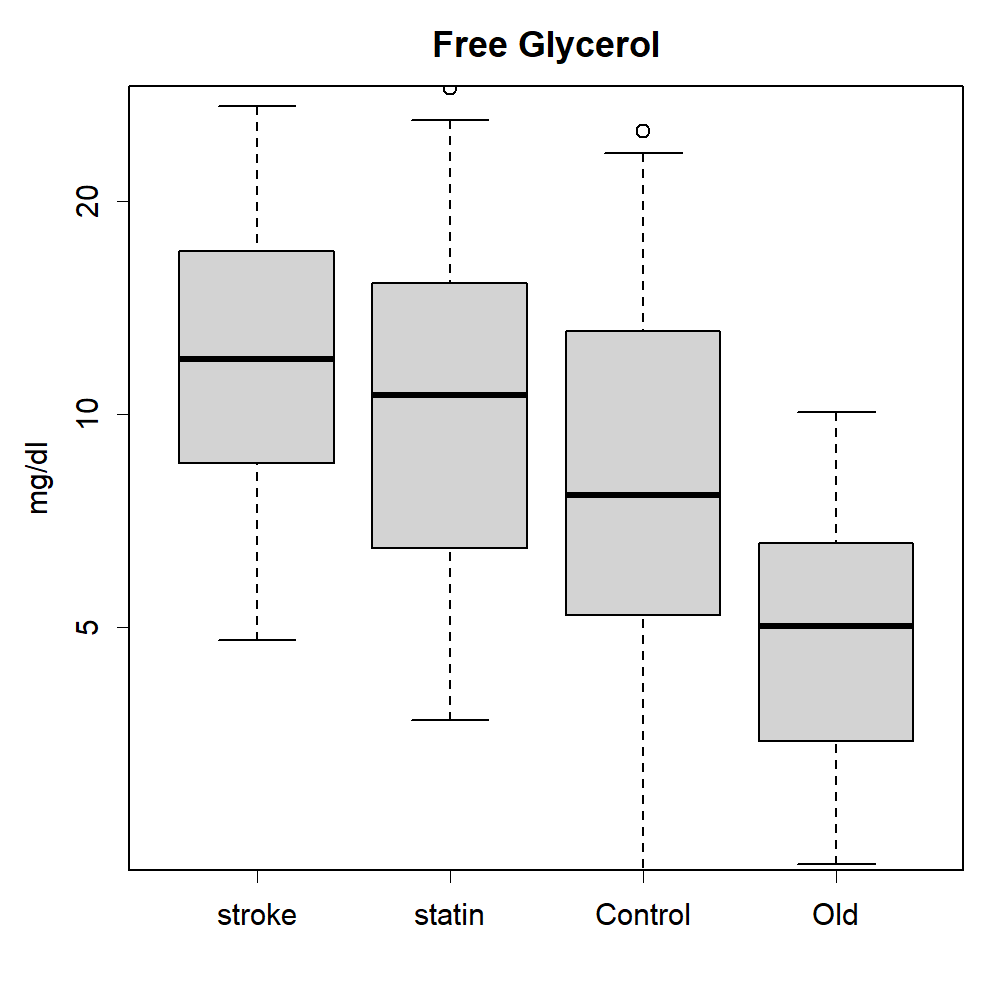

### glycerolLDL1.png

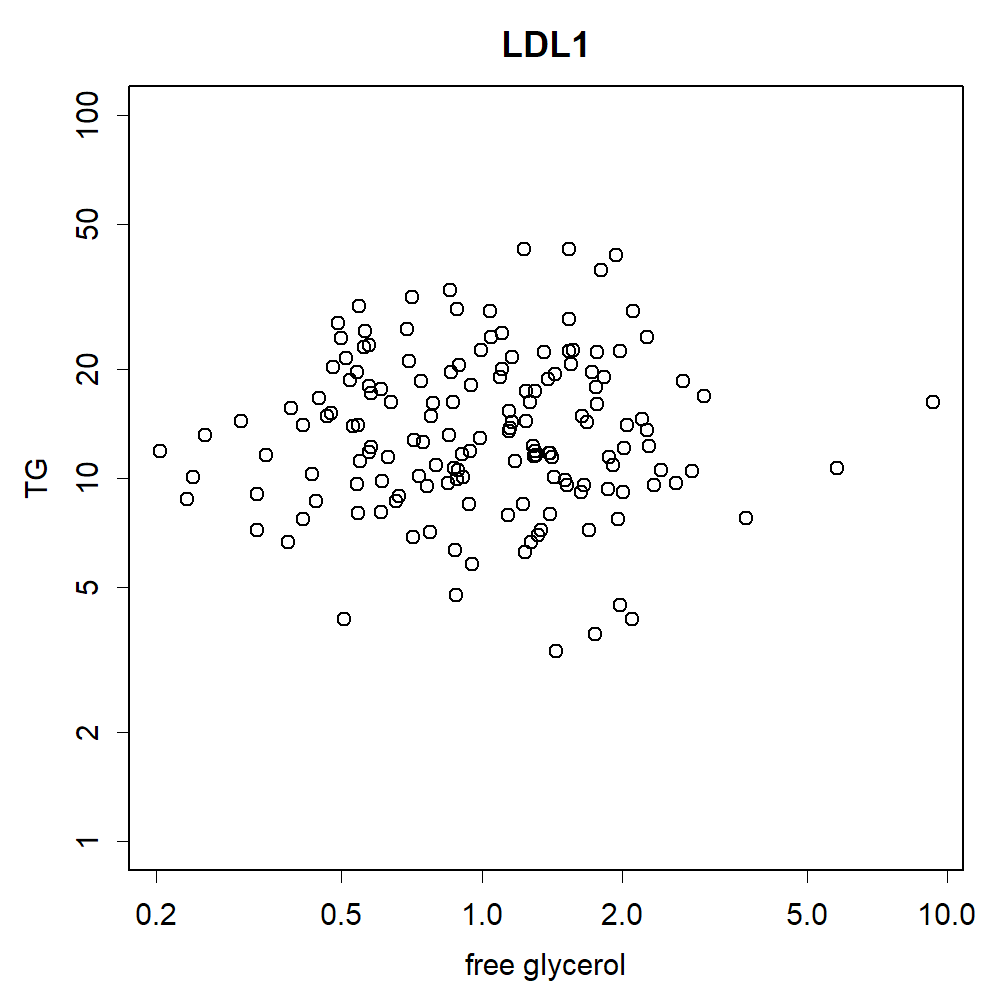

### glycerolLDL2.png

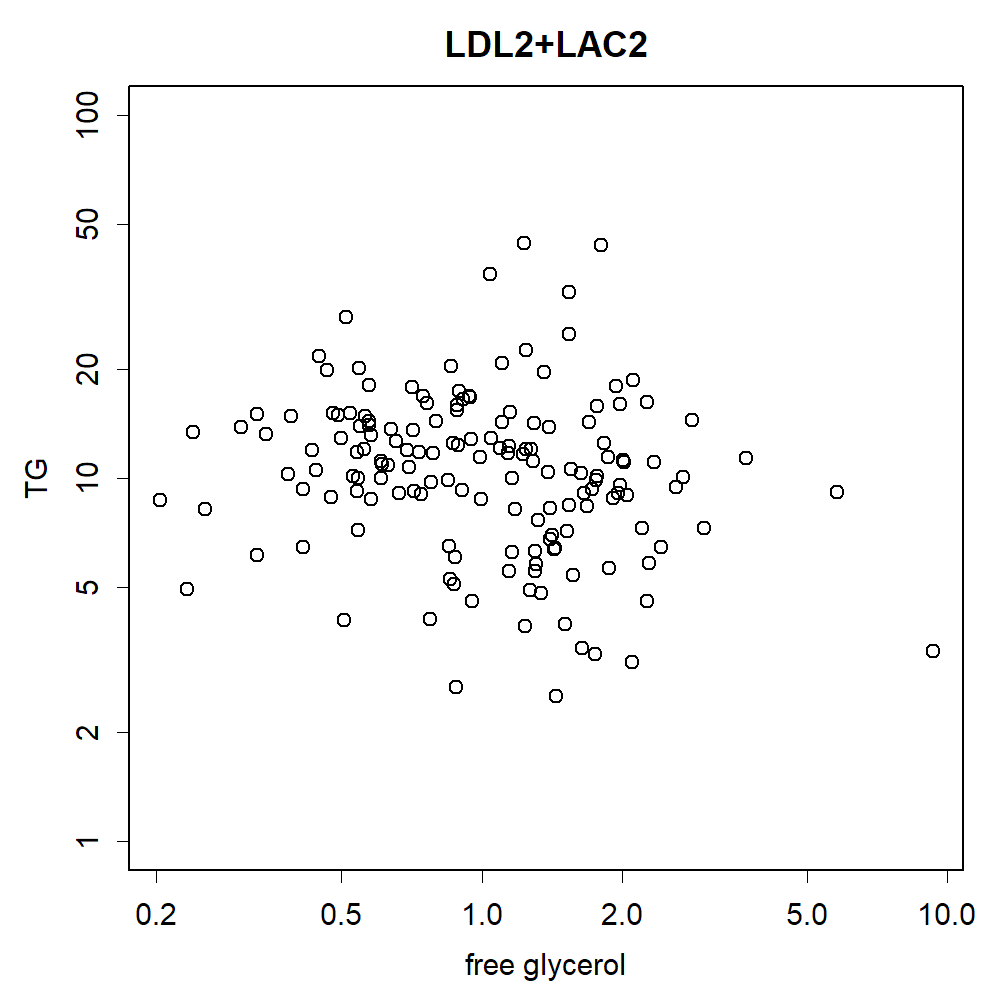

### glycerolratio.png

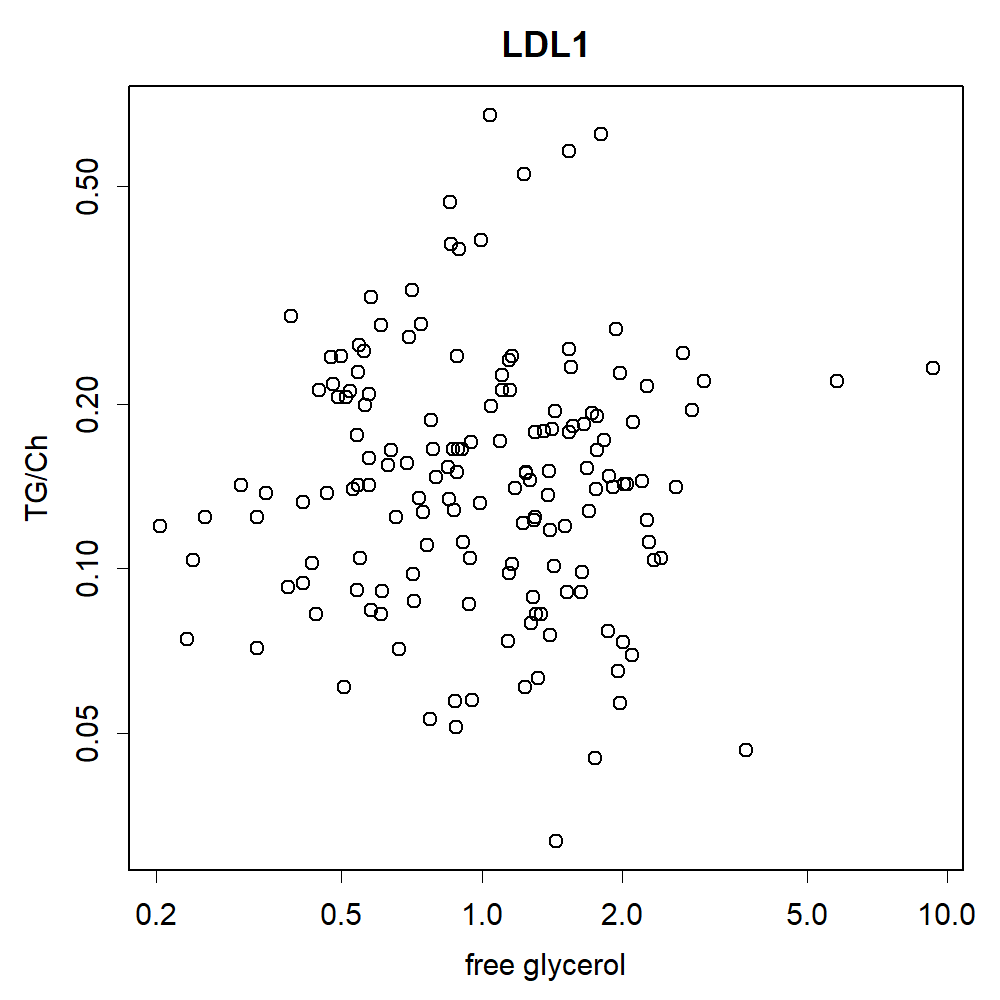

### glycerolratioCM1.png

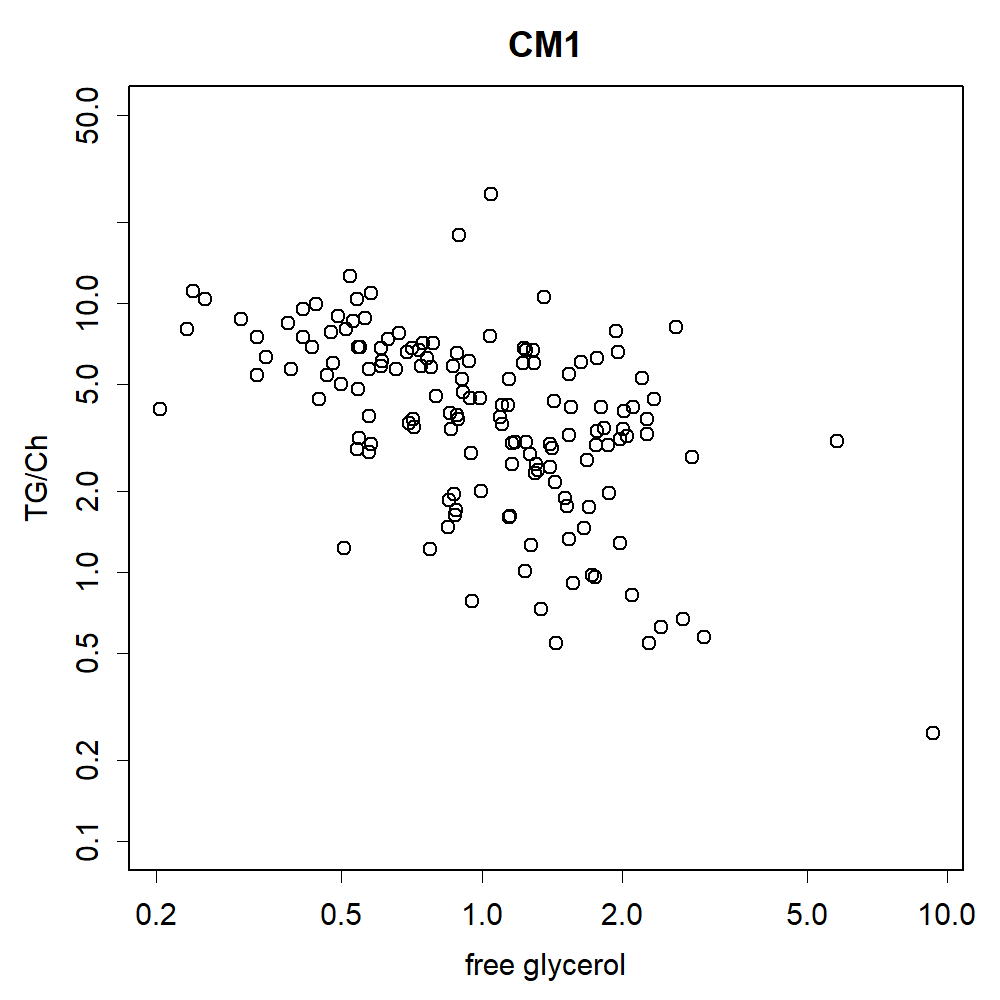

### glycerolTR.png

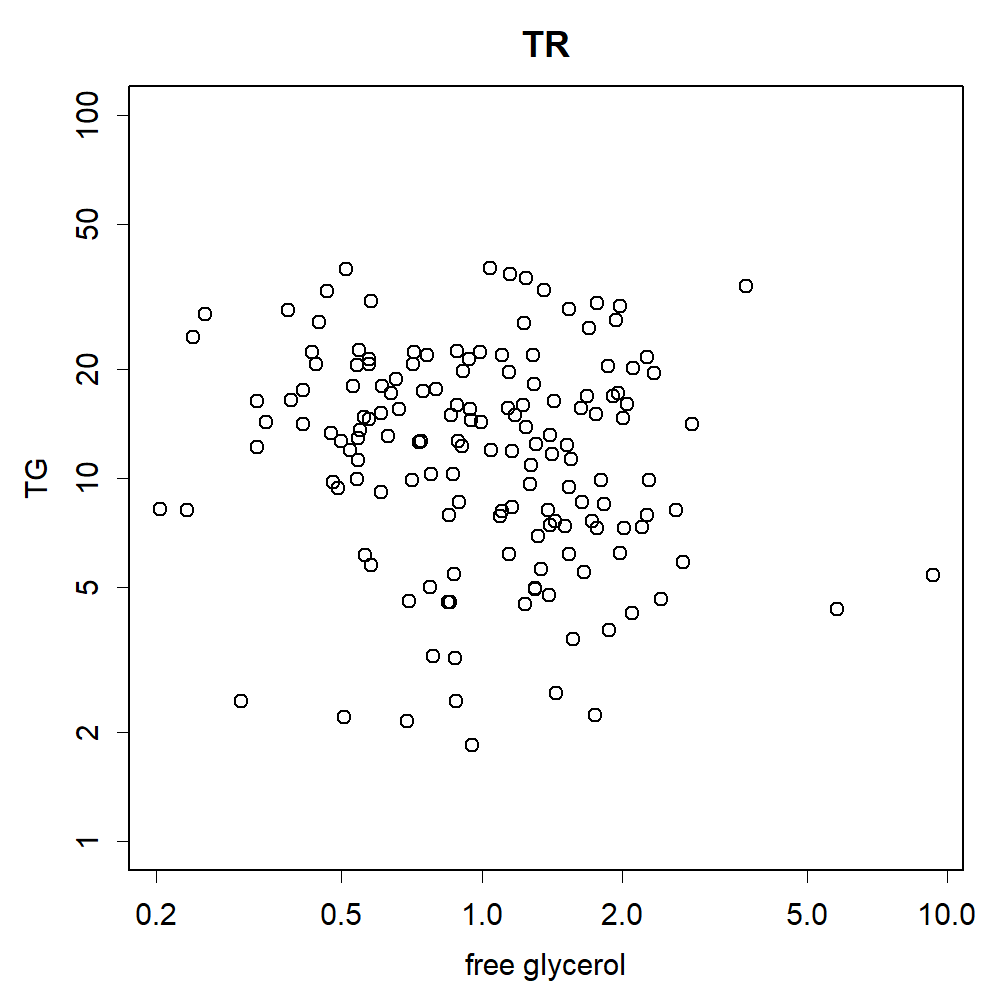

### HDL1.png

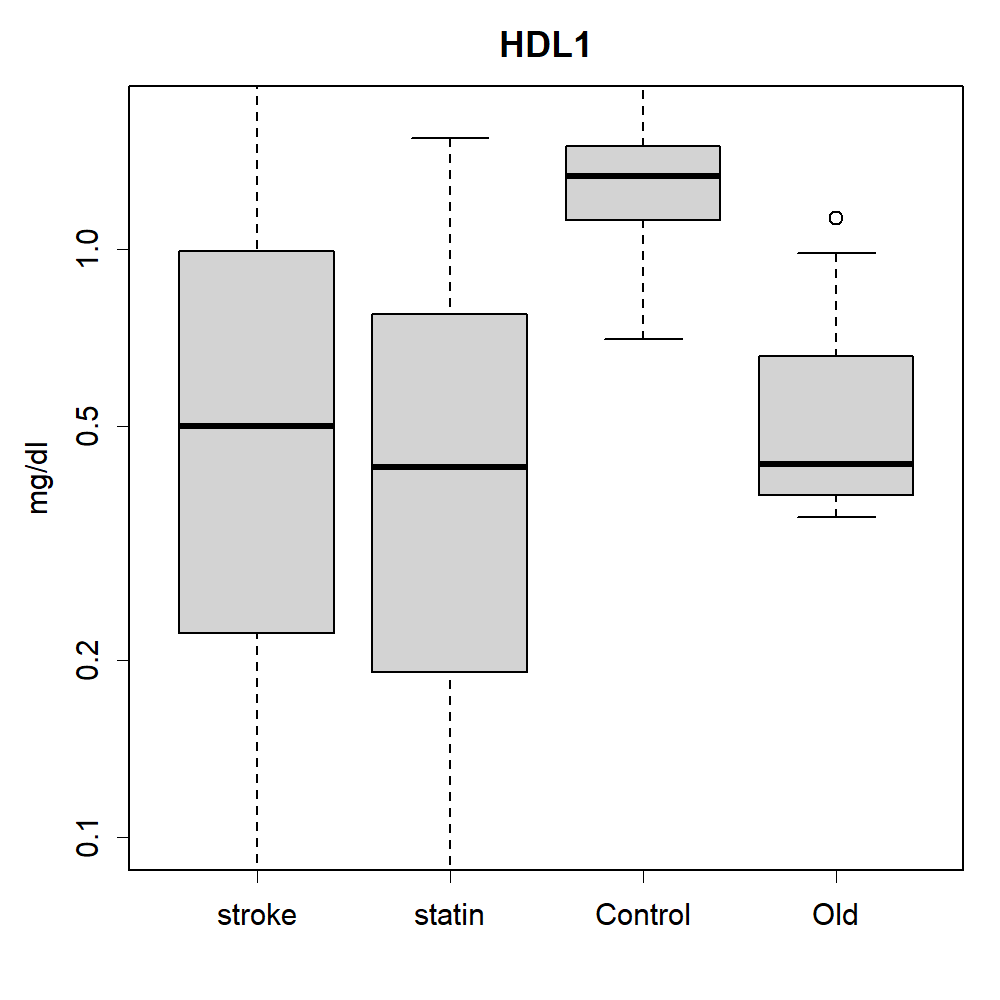

### HDL1.png

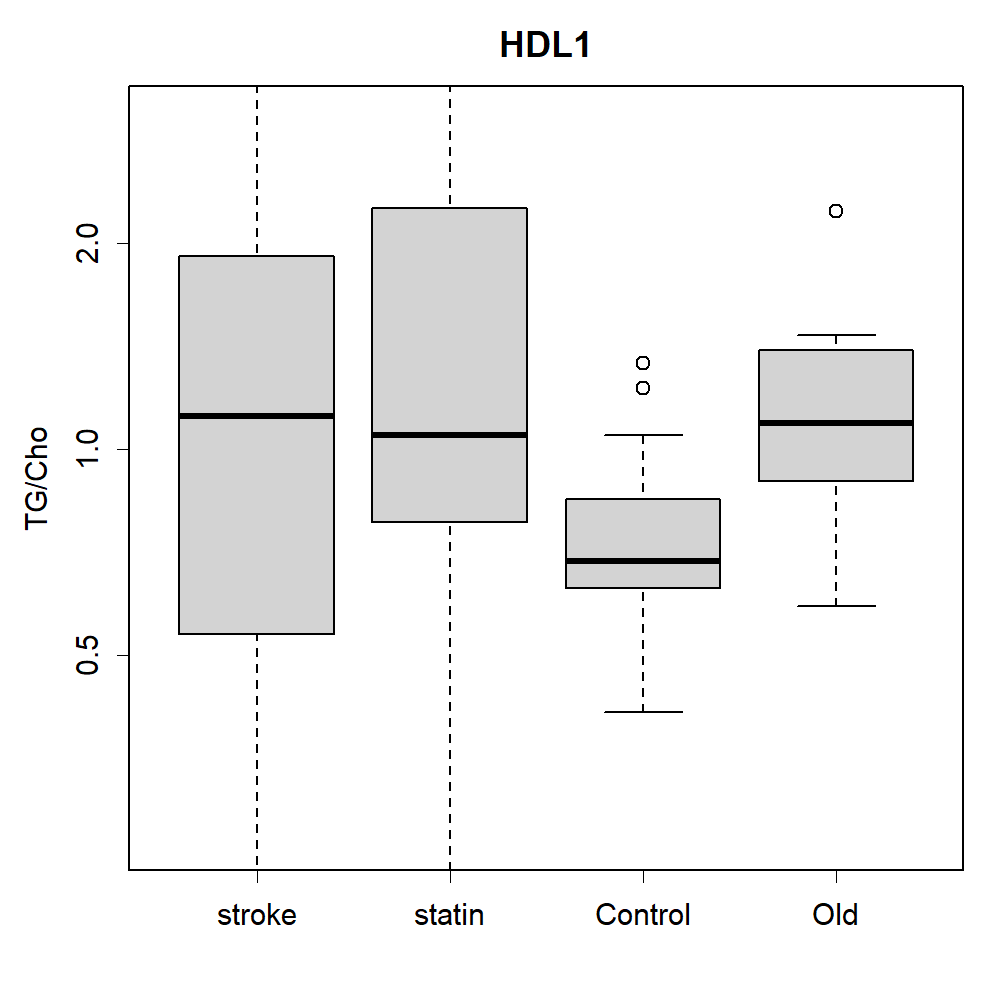

### HDL1.png

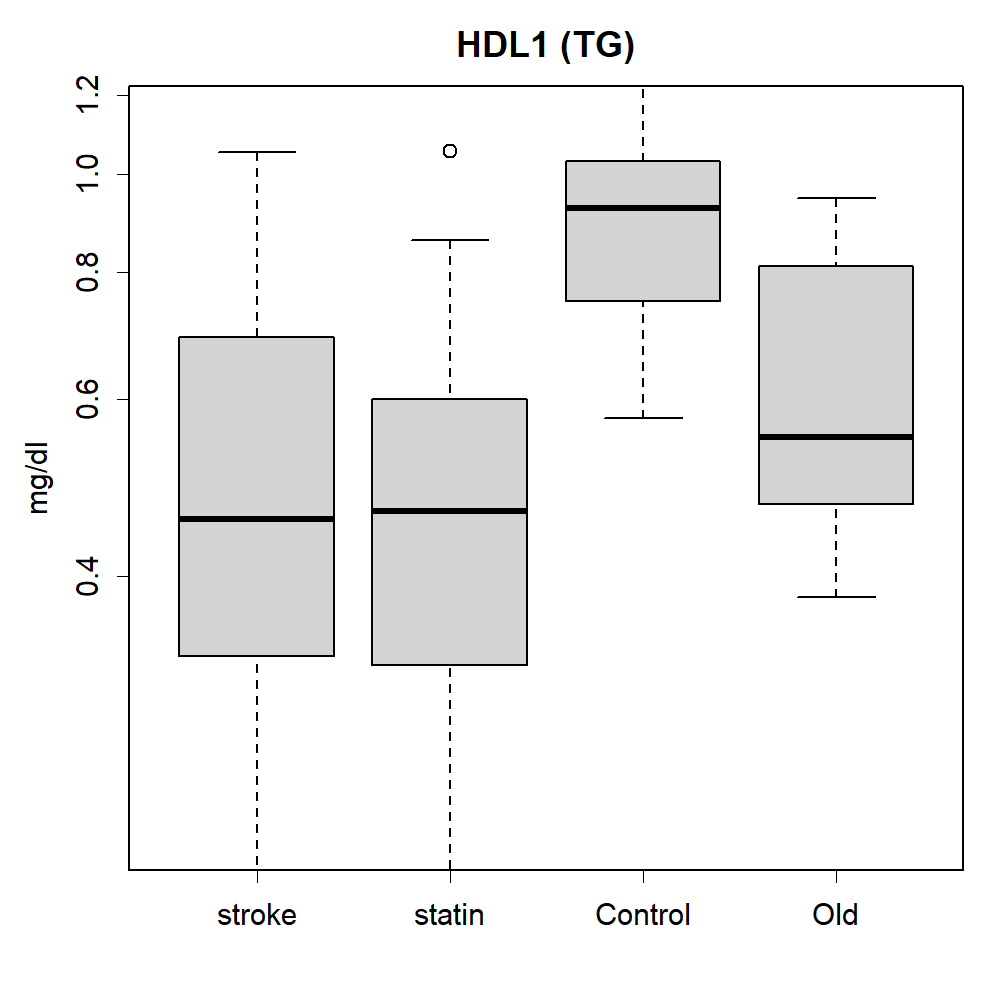

### HDL2.png

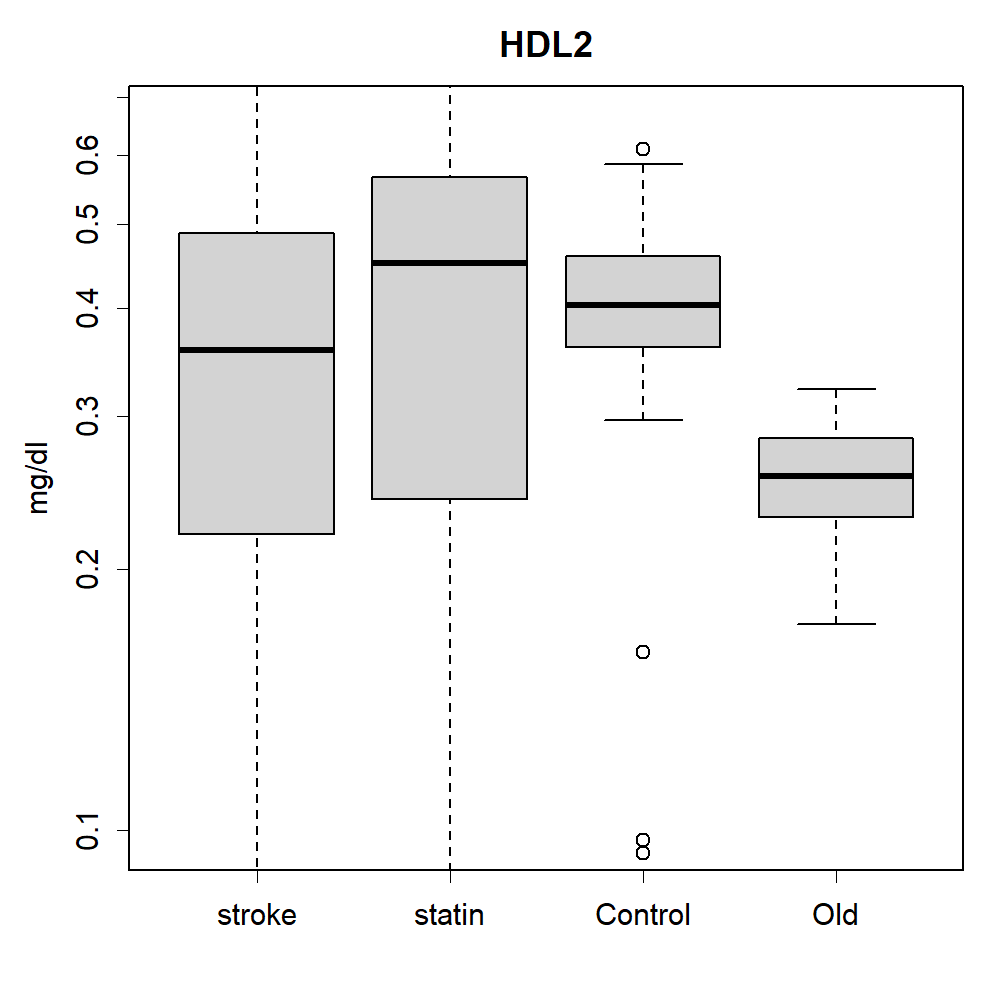

### HDL2.png

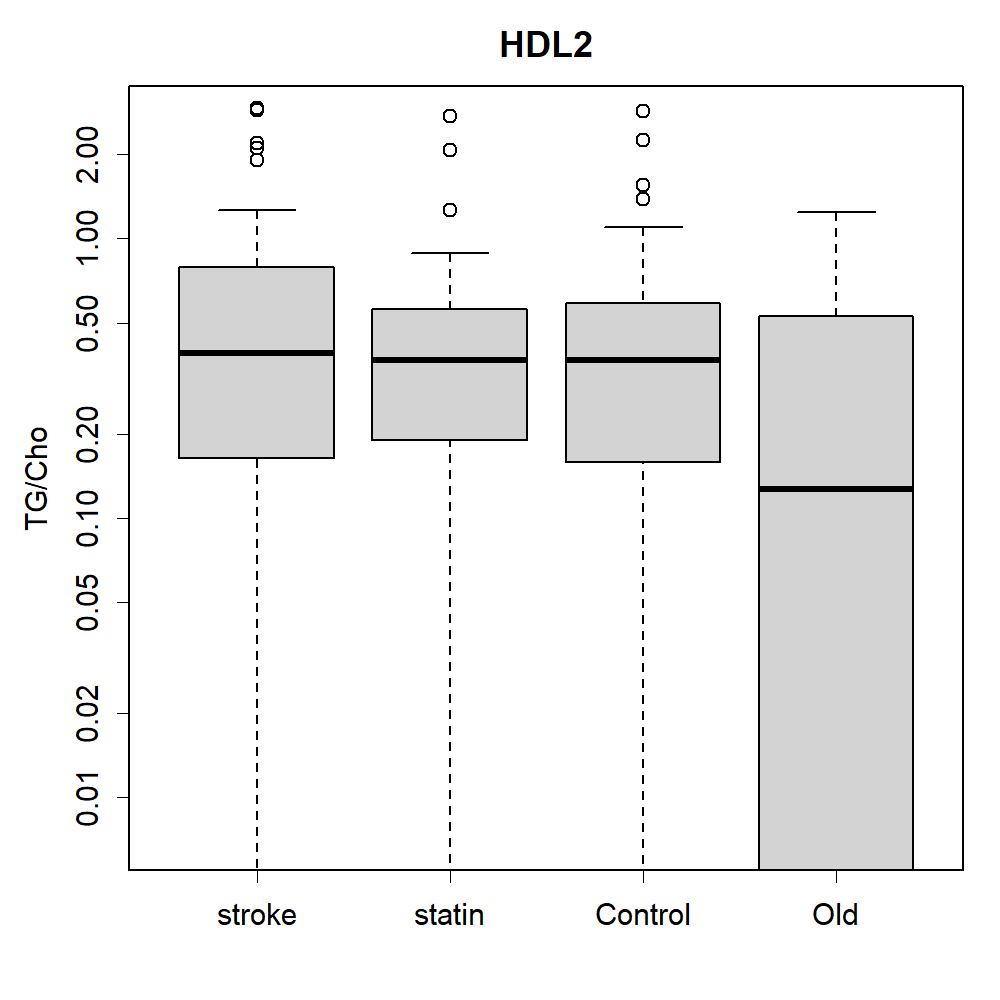

### HDL2.png

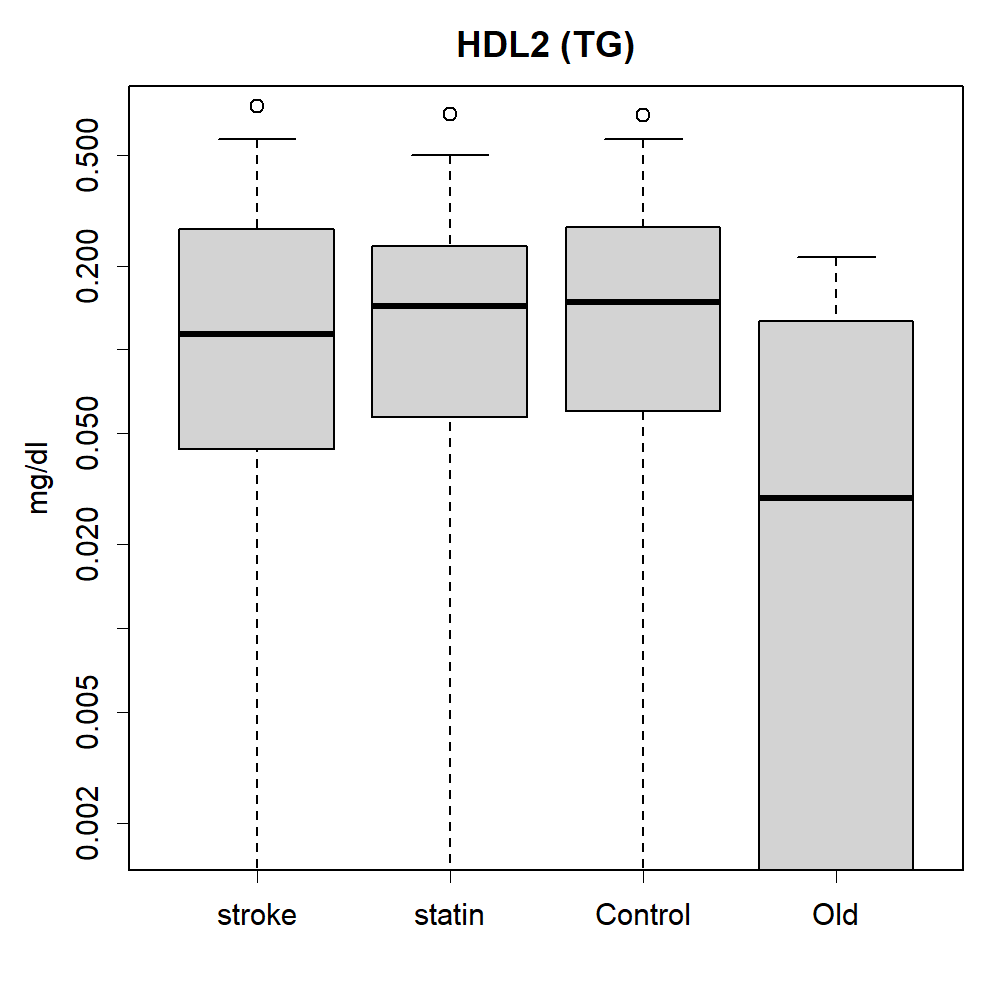

### histAge2.png

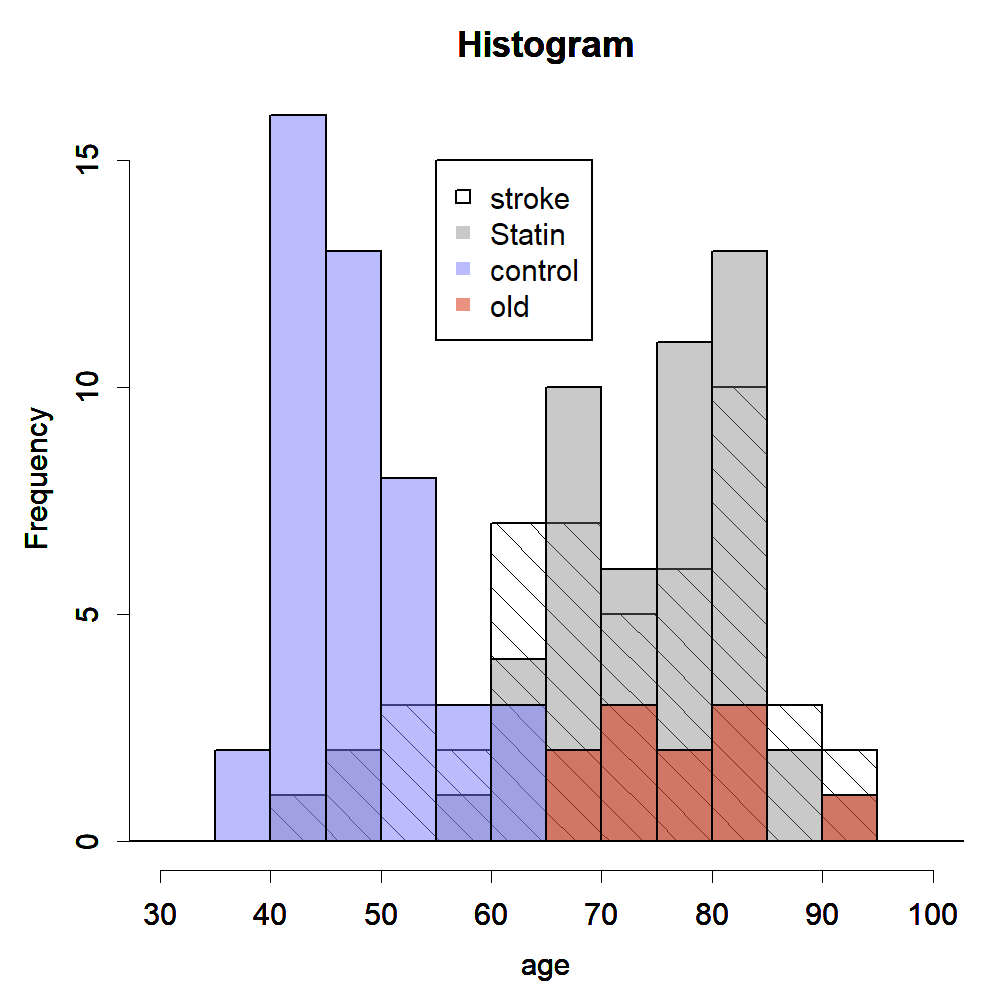

### LAC1.png

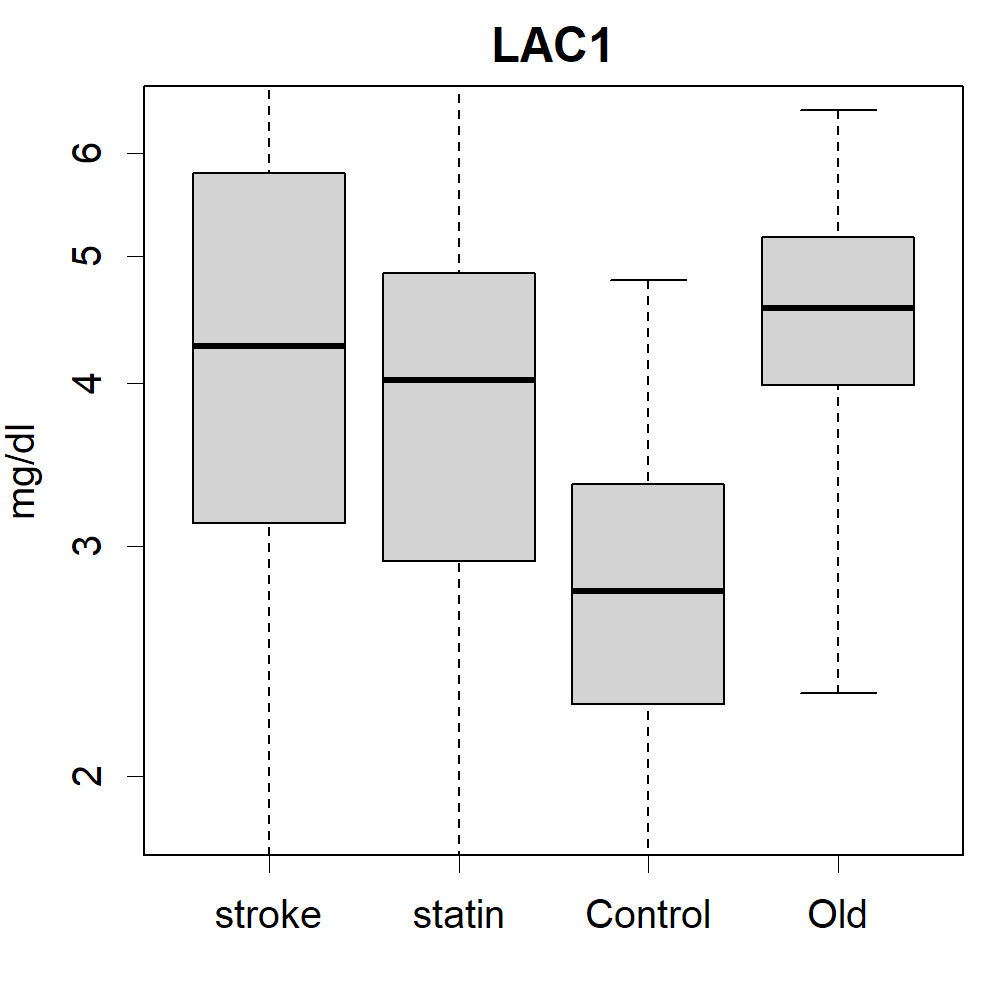

### LAC1.png

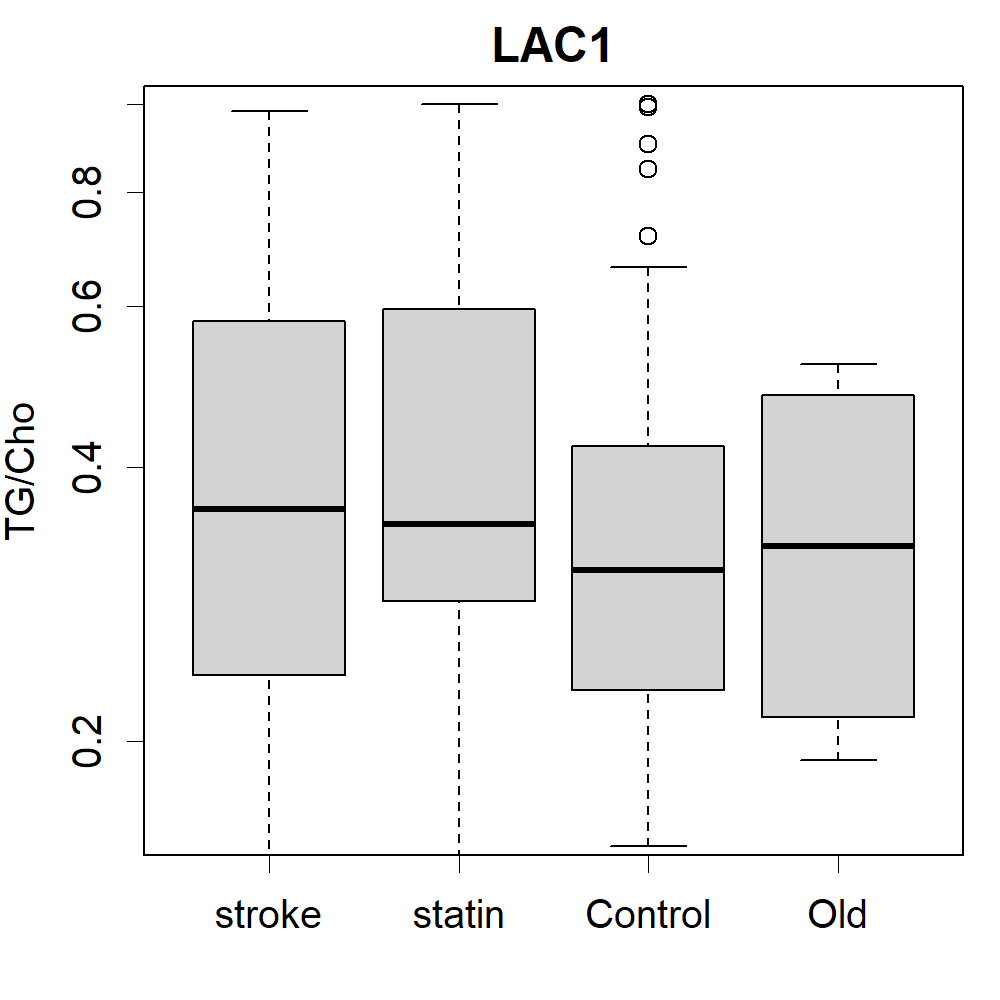

### LAC1.png

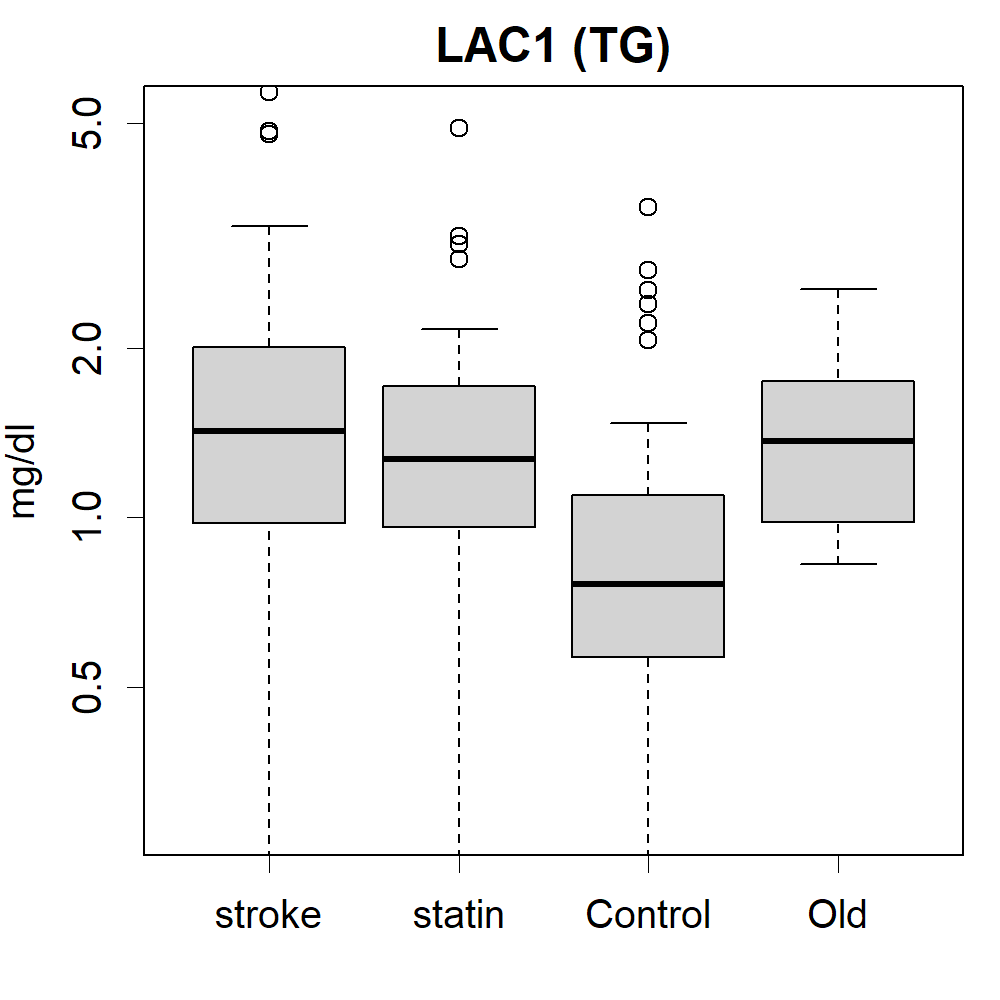

### LDL1.png

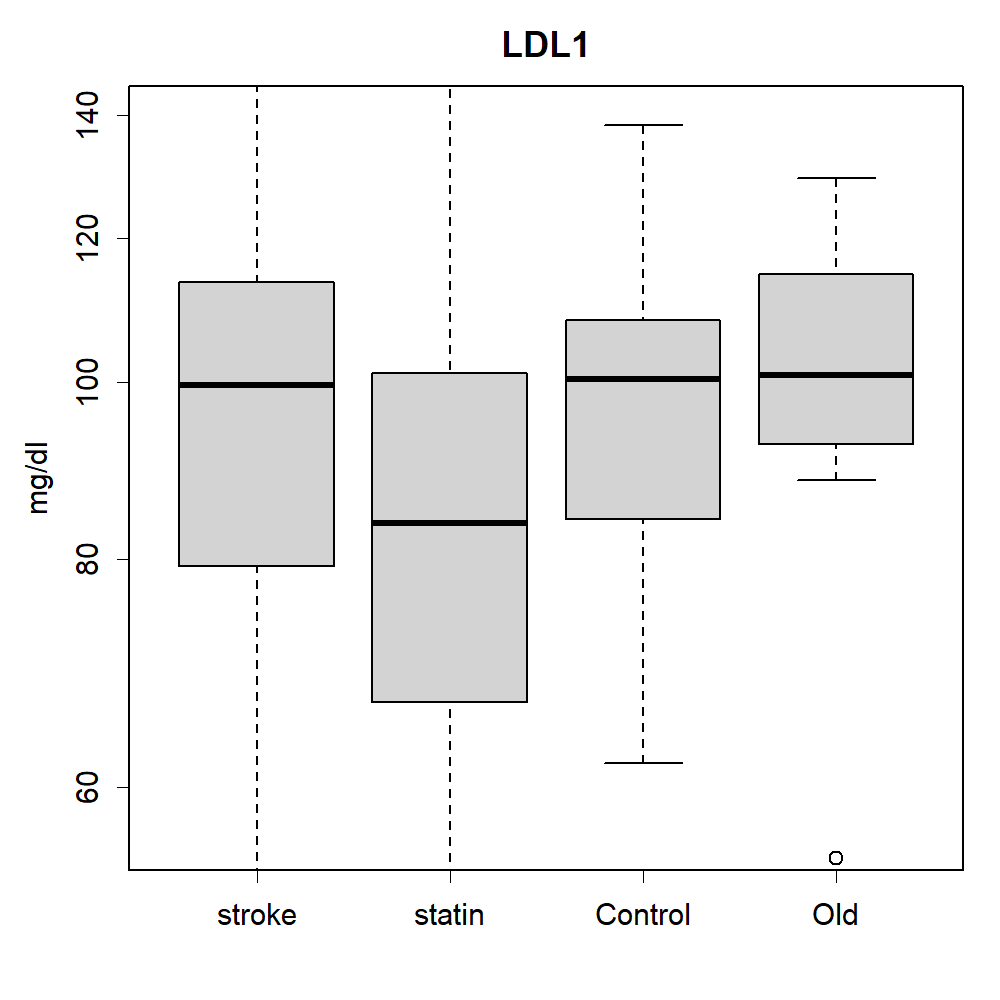

### LDL1.png

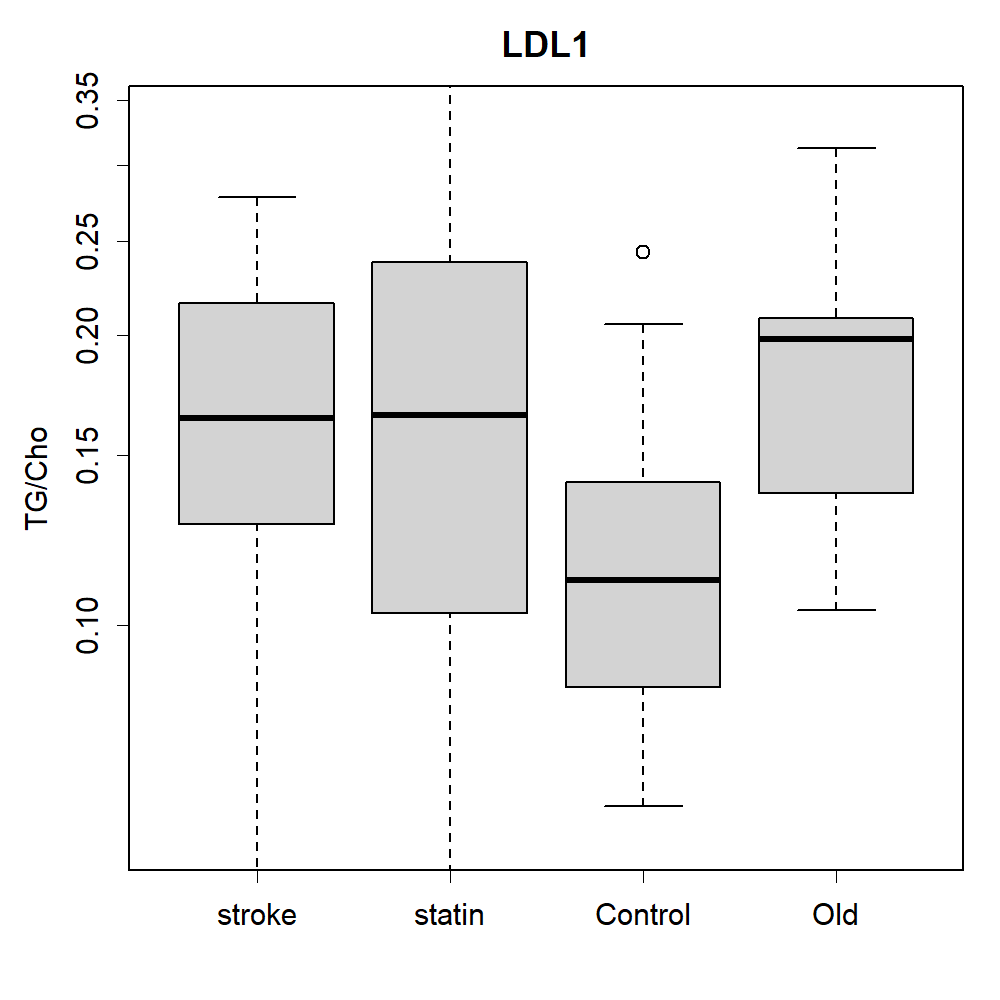

### LDL1.png

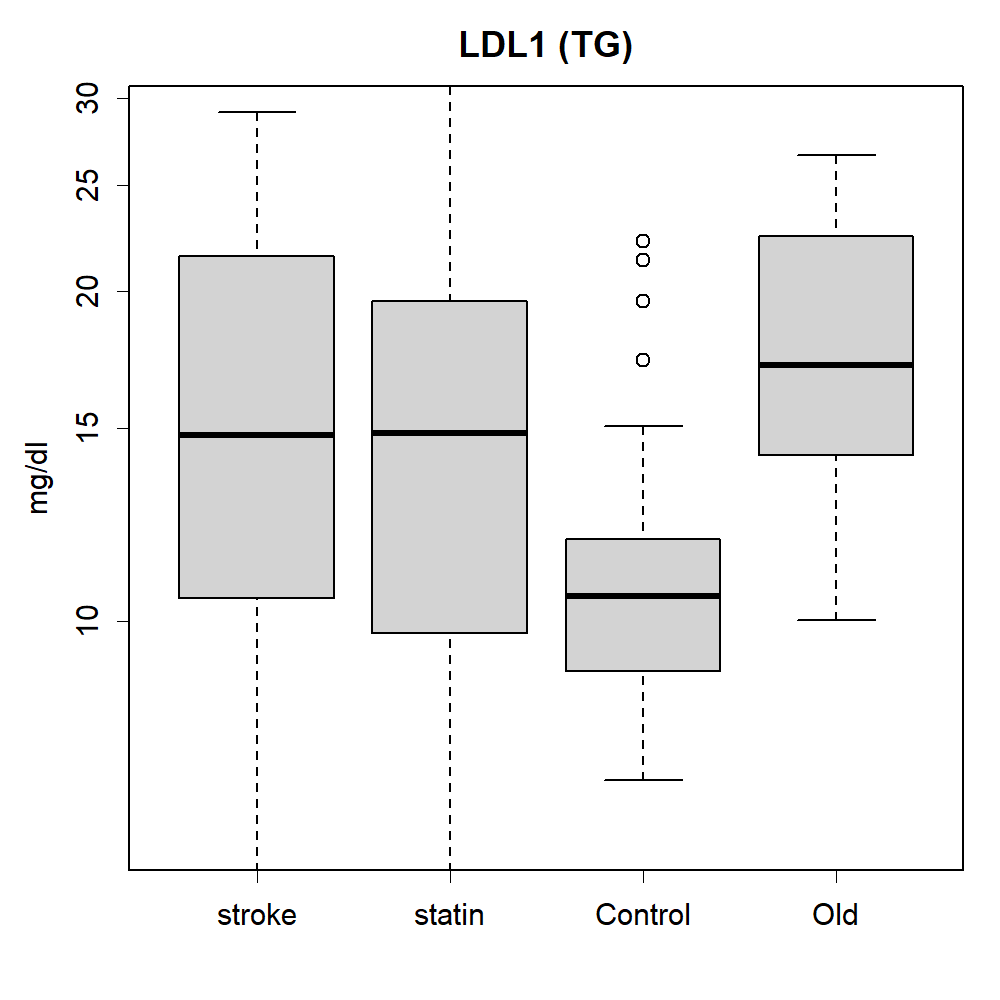

### LDL2+LAC2.png

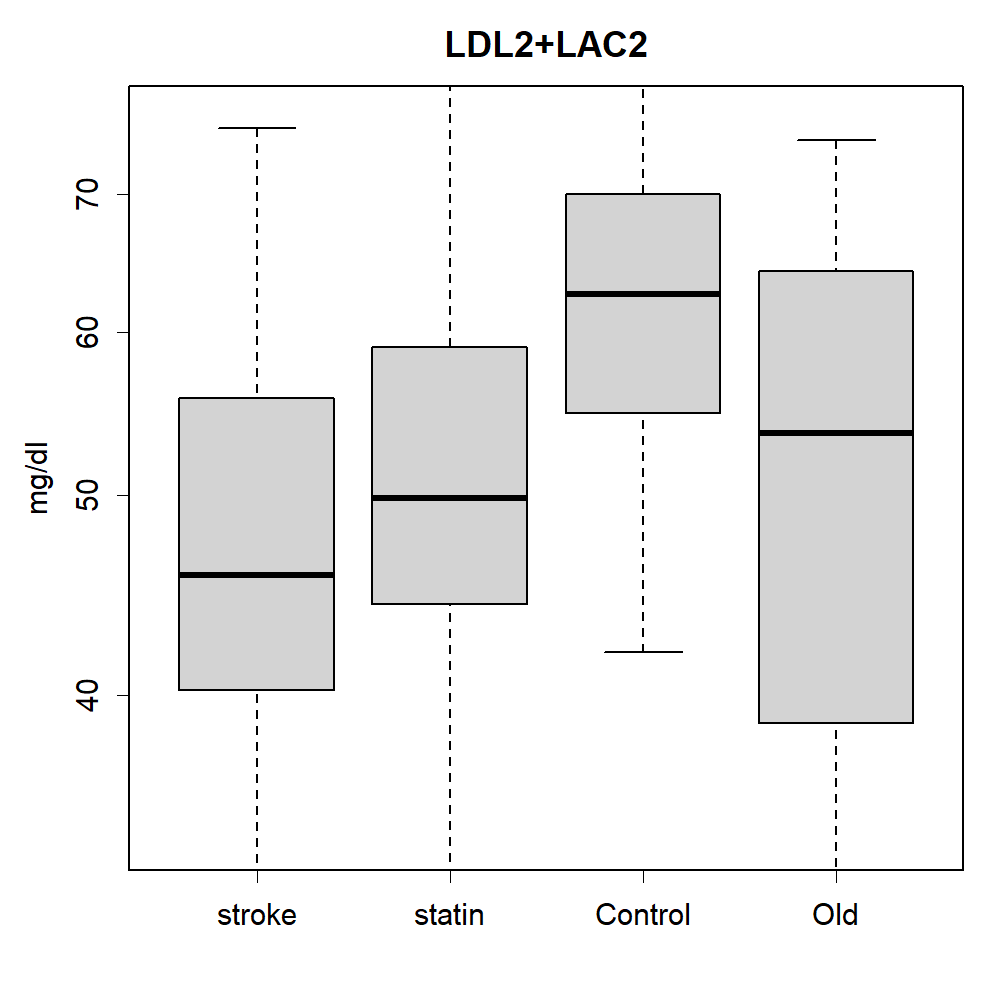

### LDL2+LAC2.png

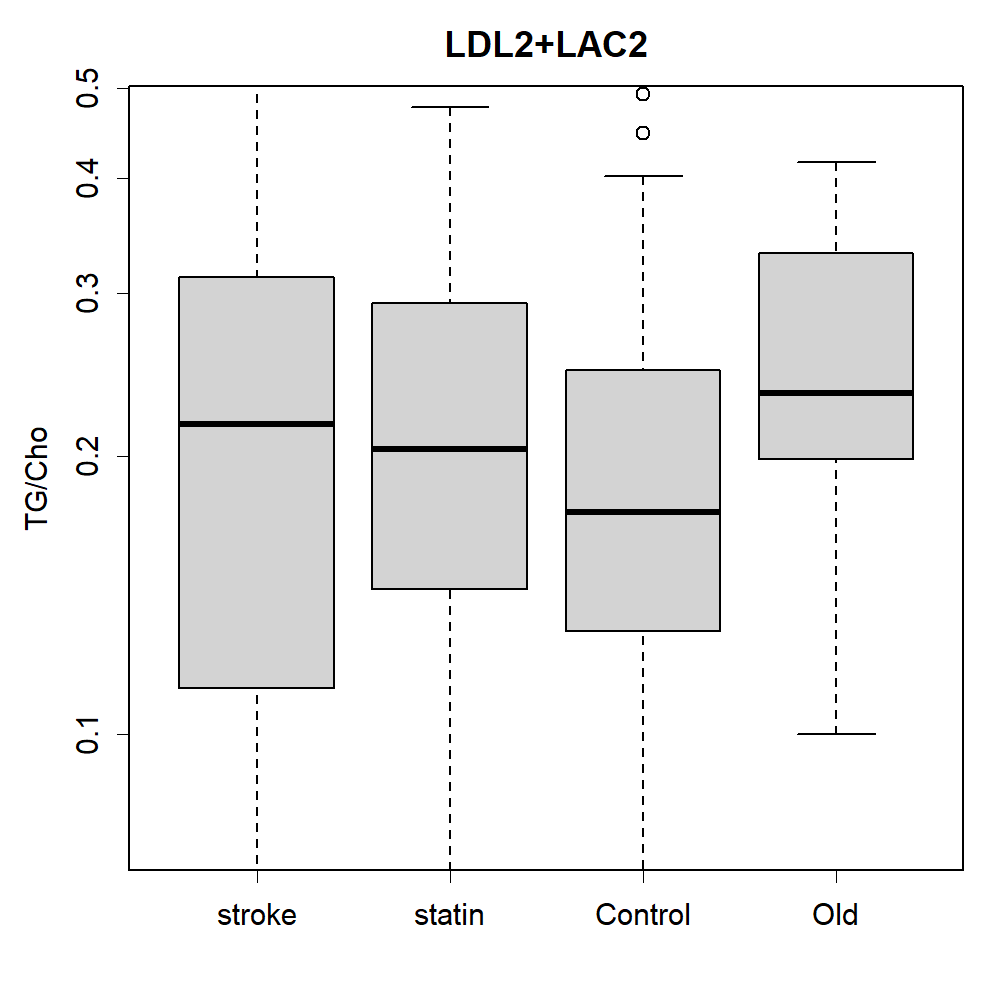

### LDL2+LAC2.png

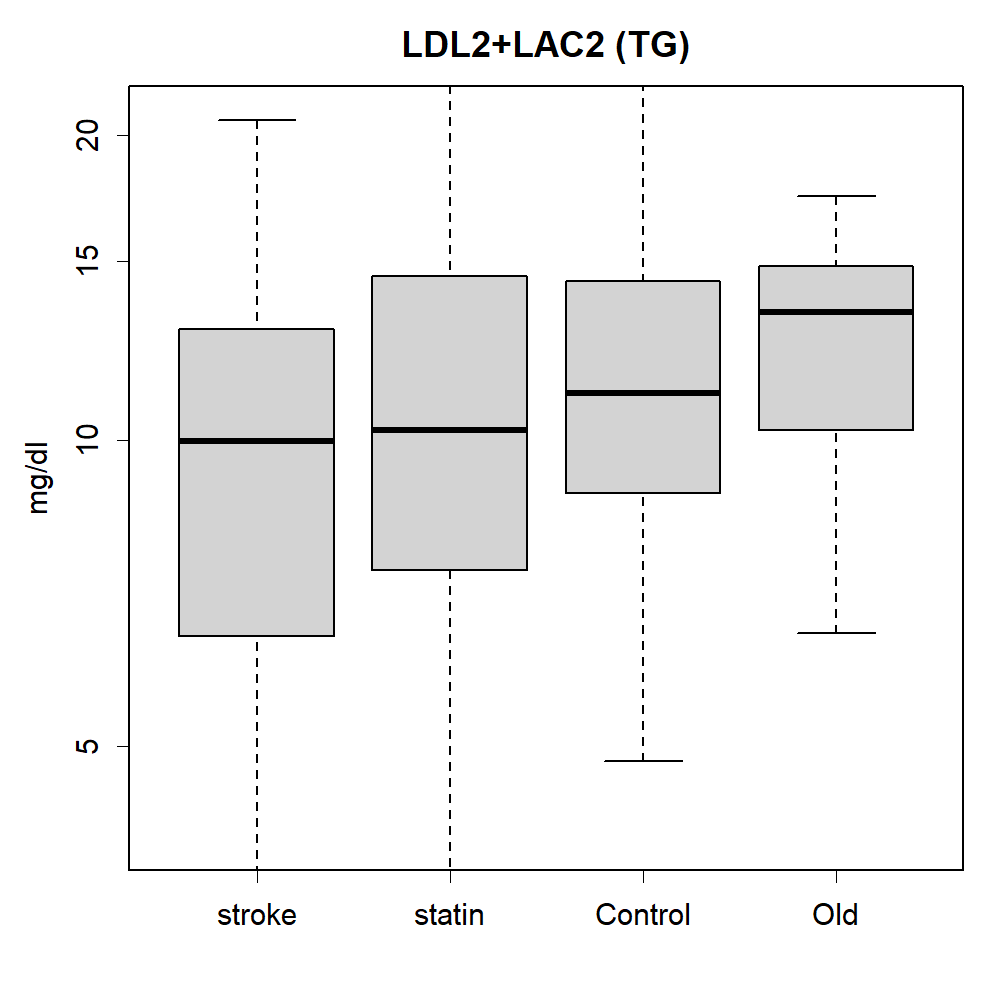

### Lp(a).png

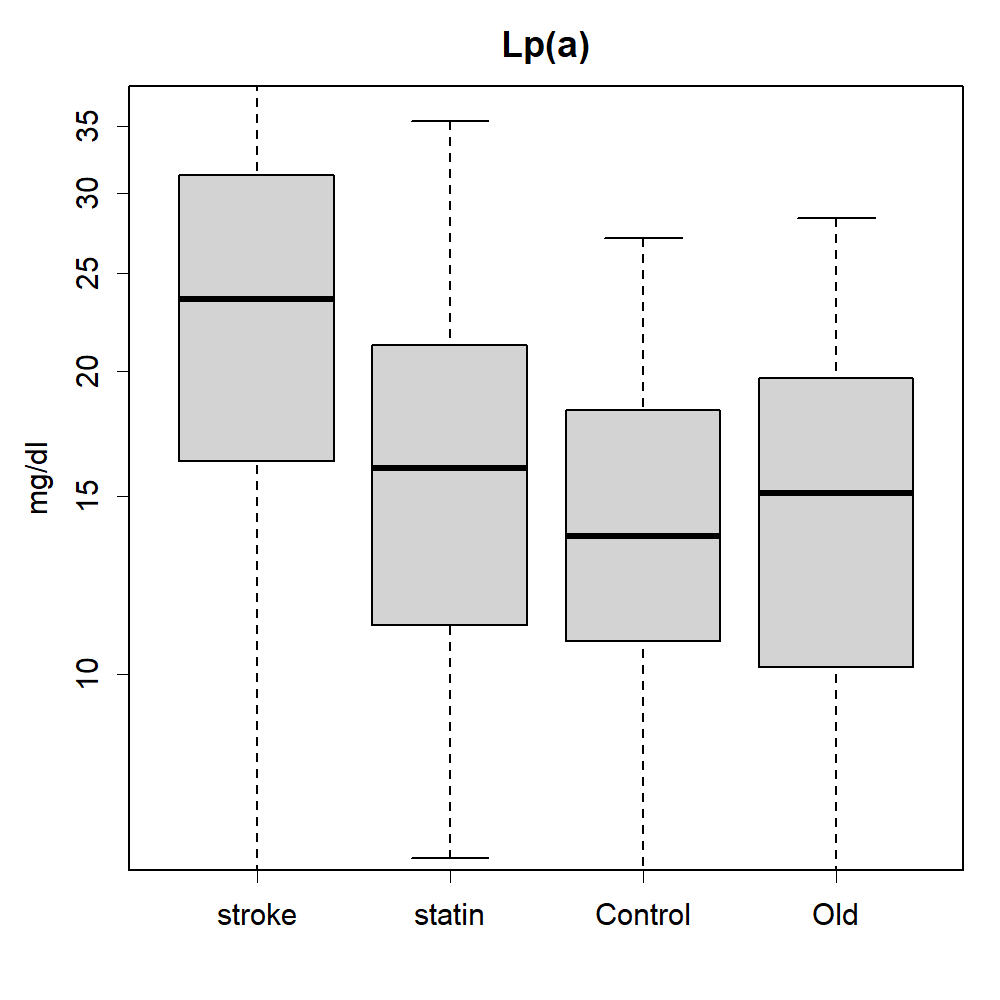

### Lp(a).png

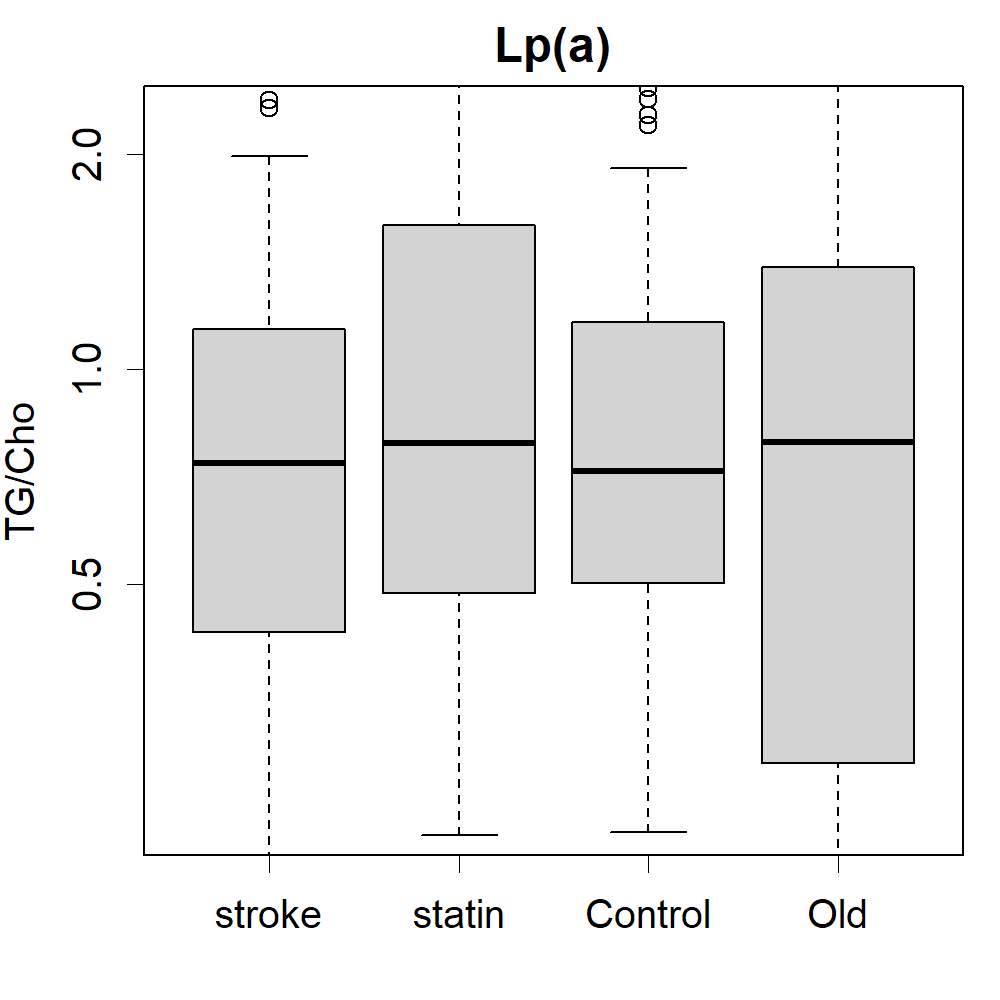

### Lp(a).png

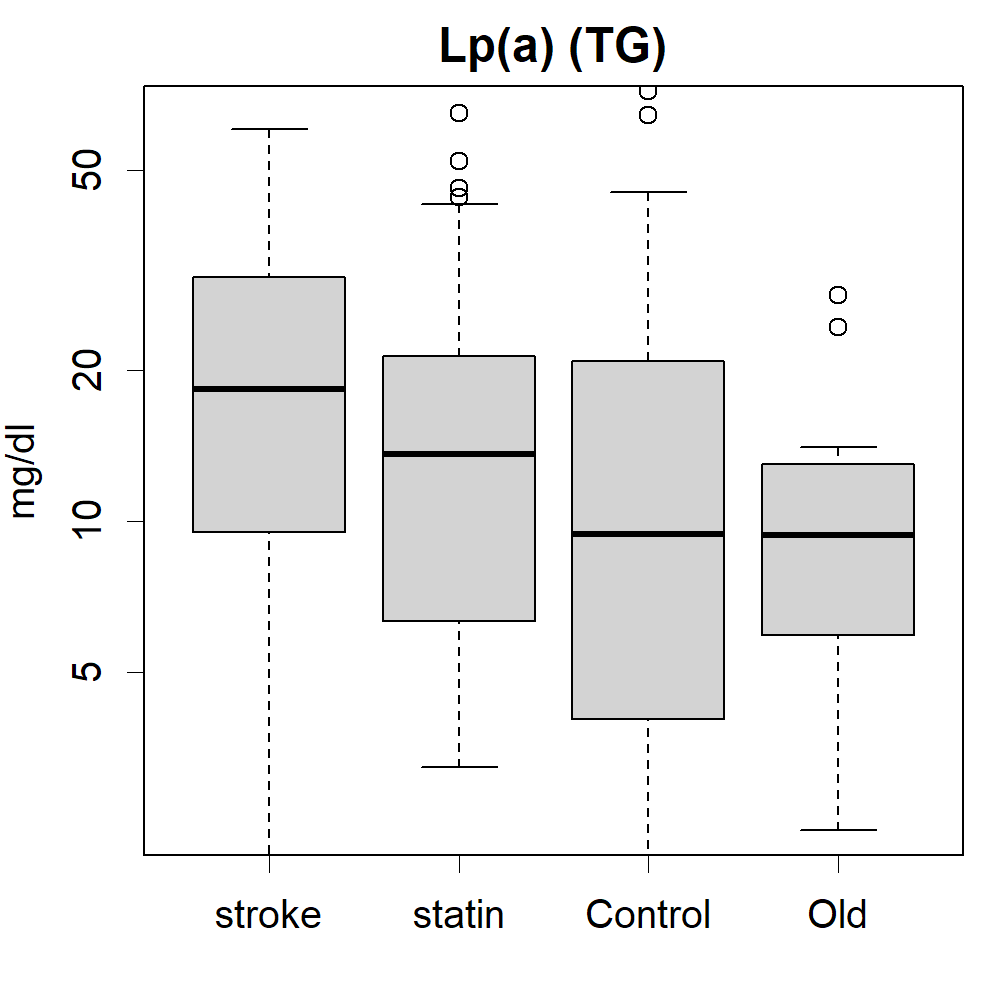

### mHDL.png

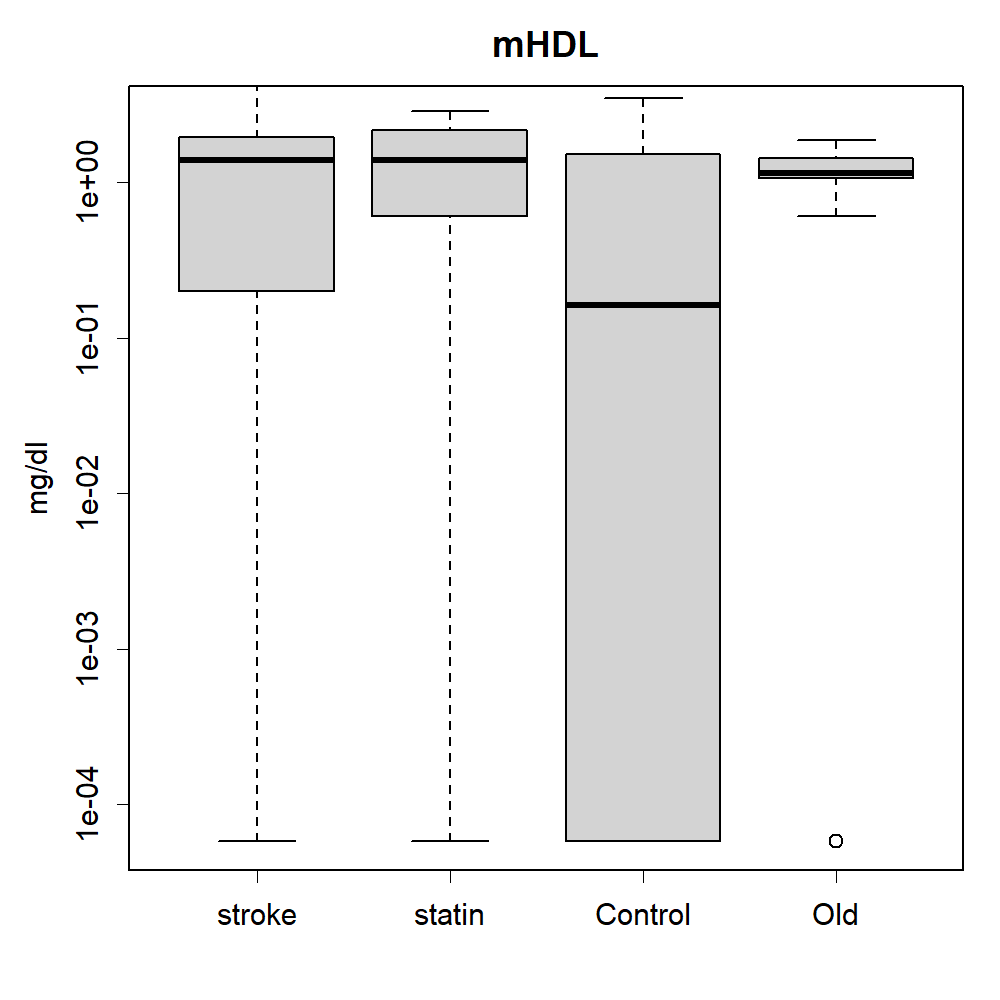

### mHDL.png

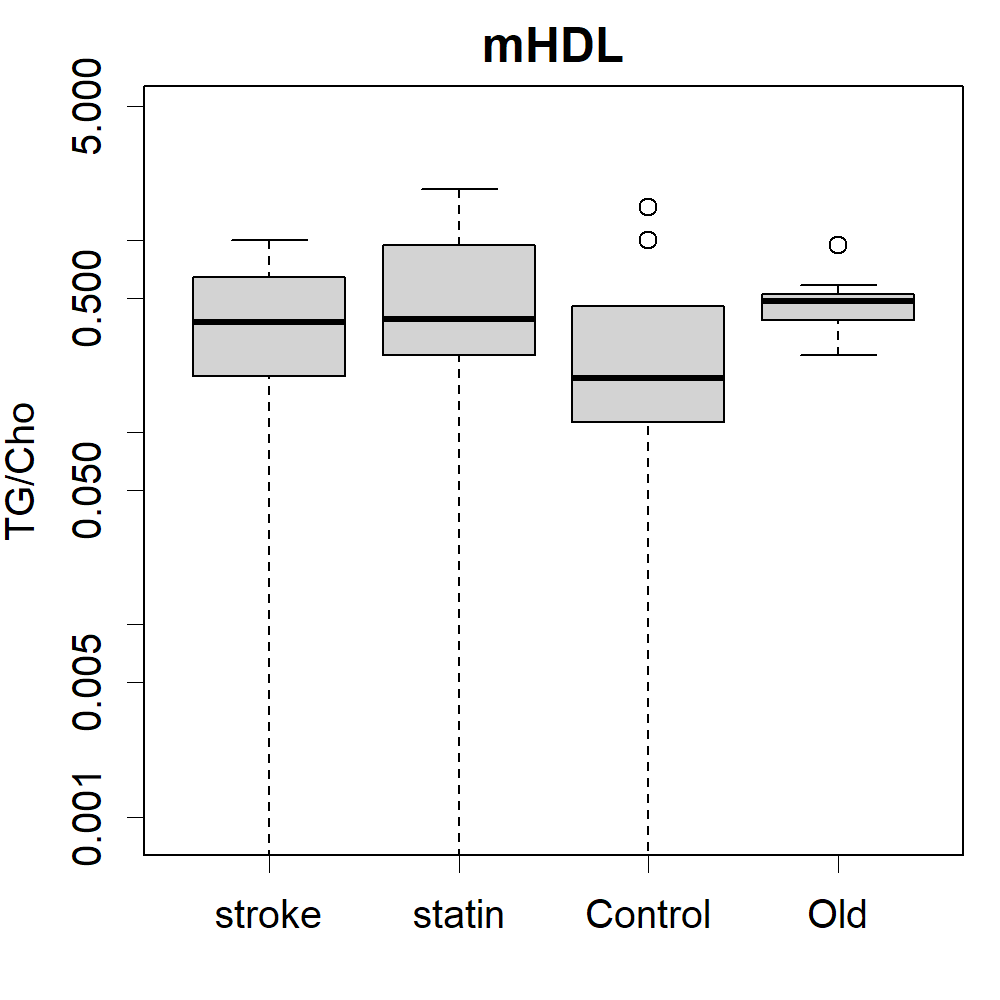

### mHDL.png

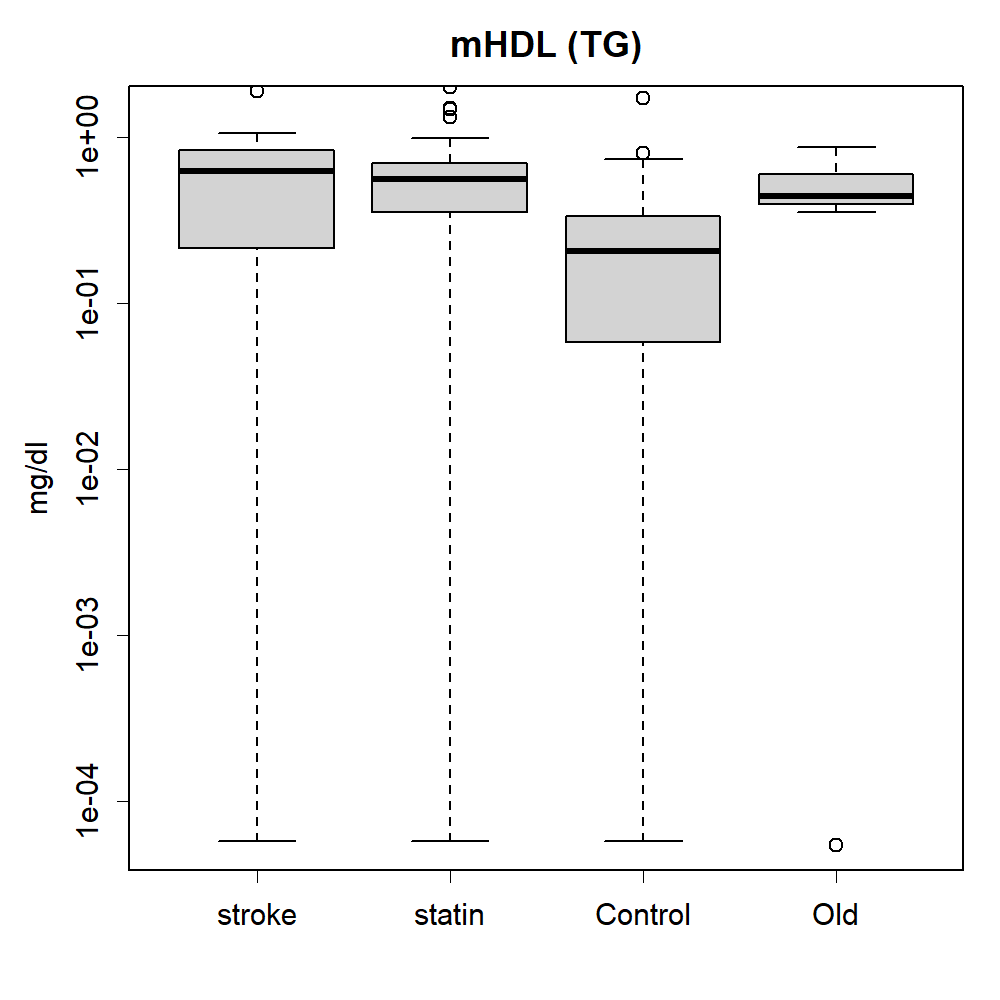

### PC12.png

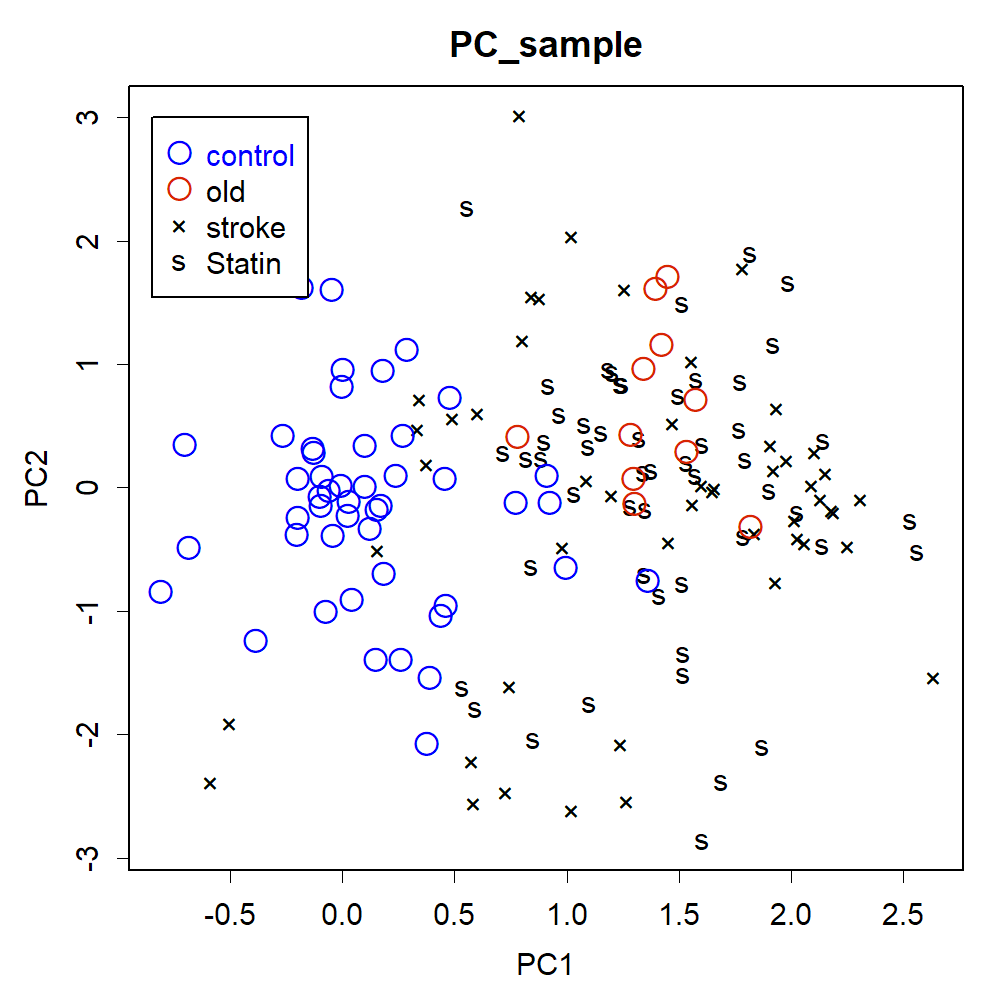

### PC12A.png

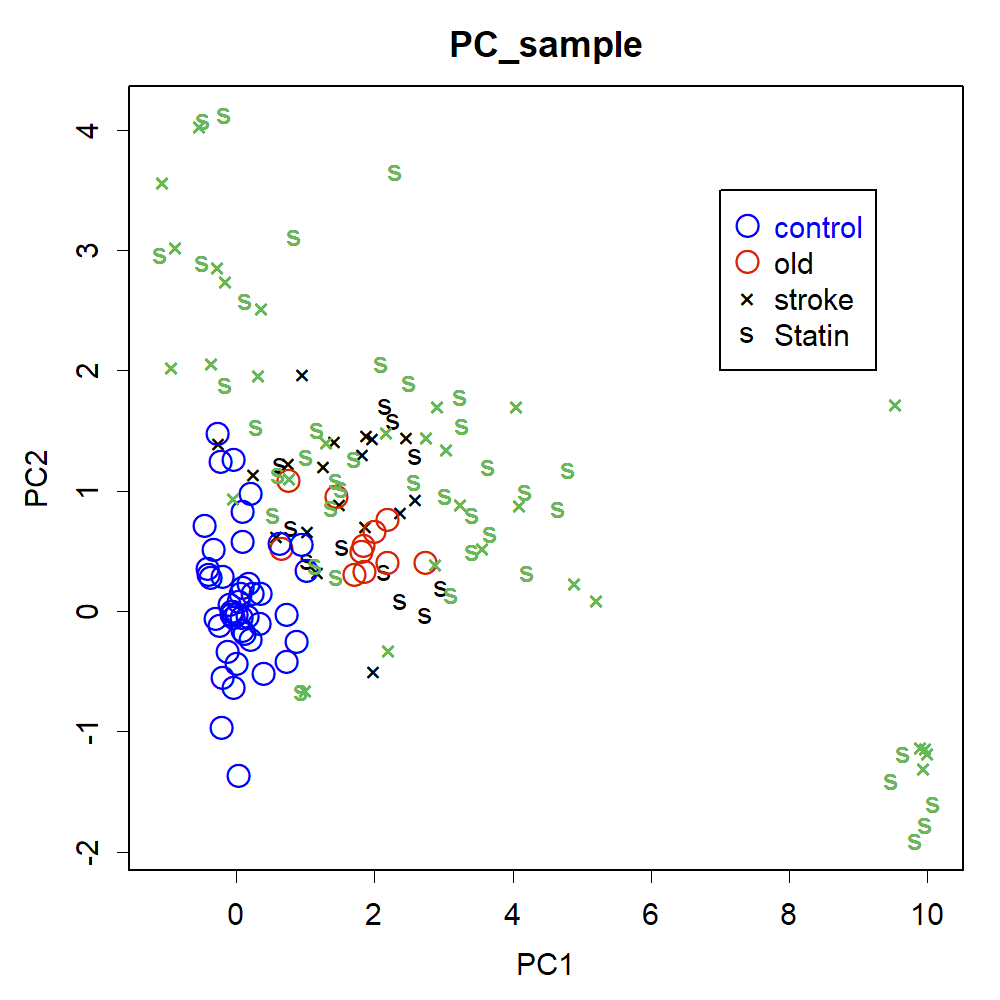

### PC12Furui.png

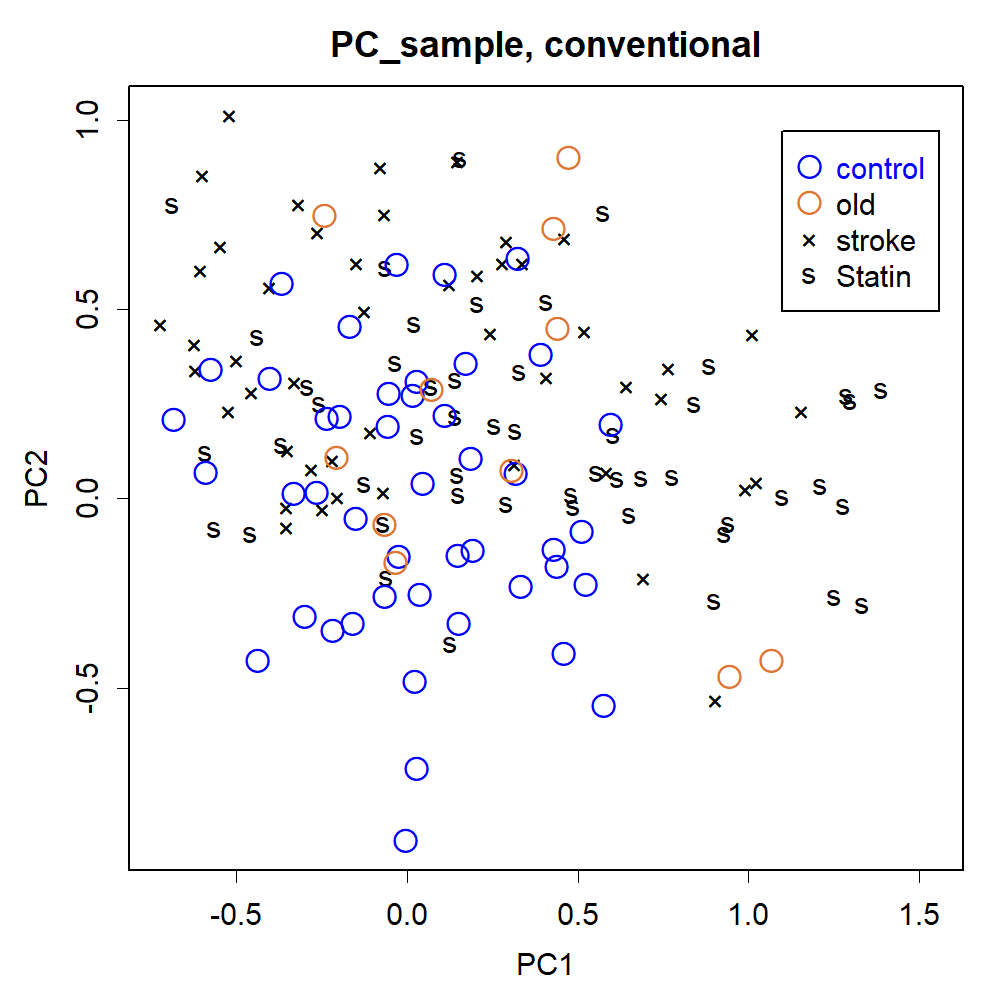

### PC12item.png

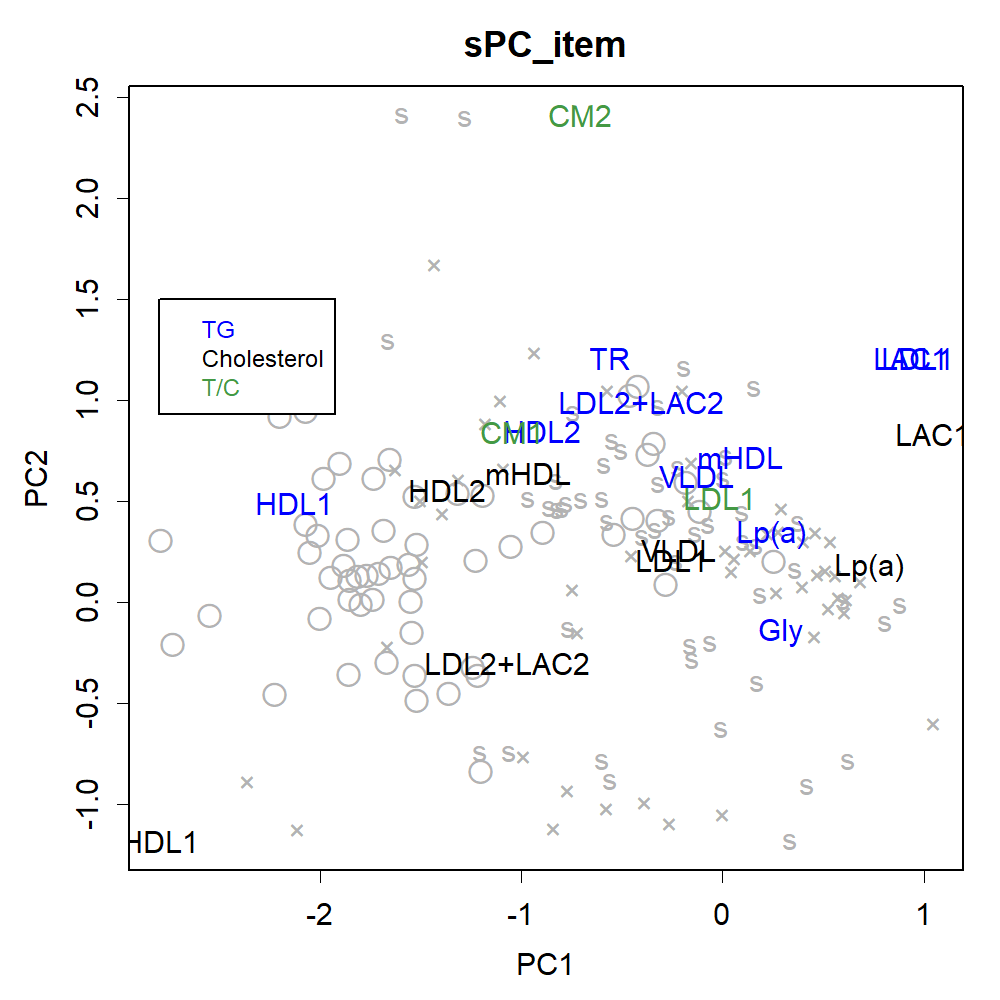

### PC12itemA.png

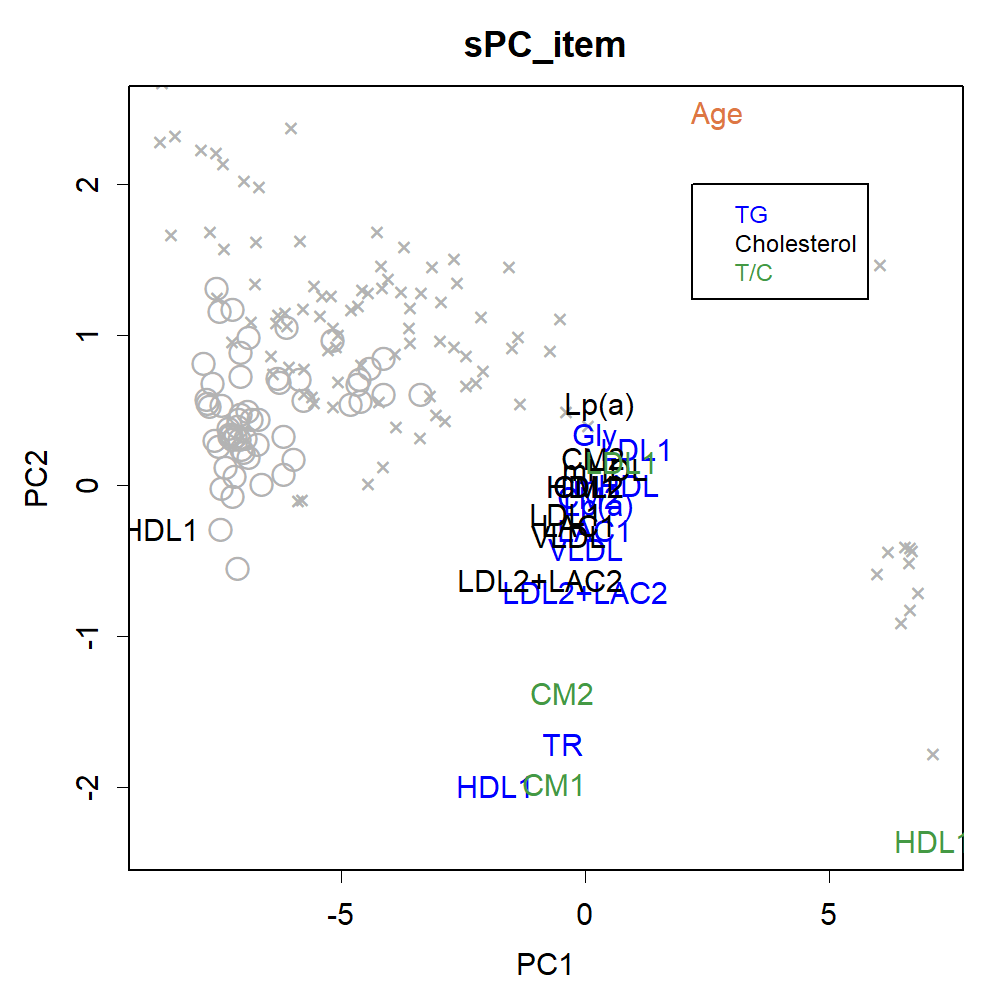

### PC12itemFurui.png

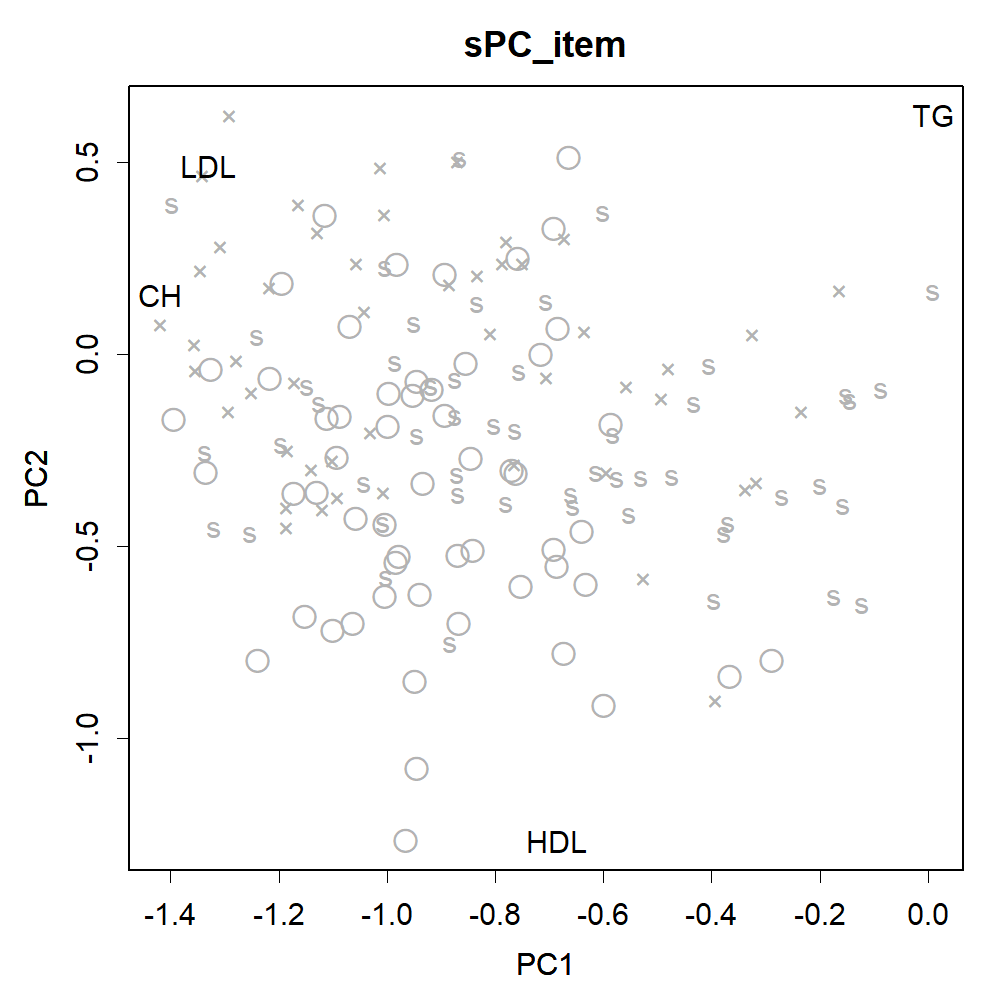

### PC12itemOOKISA.png

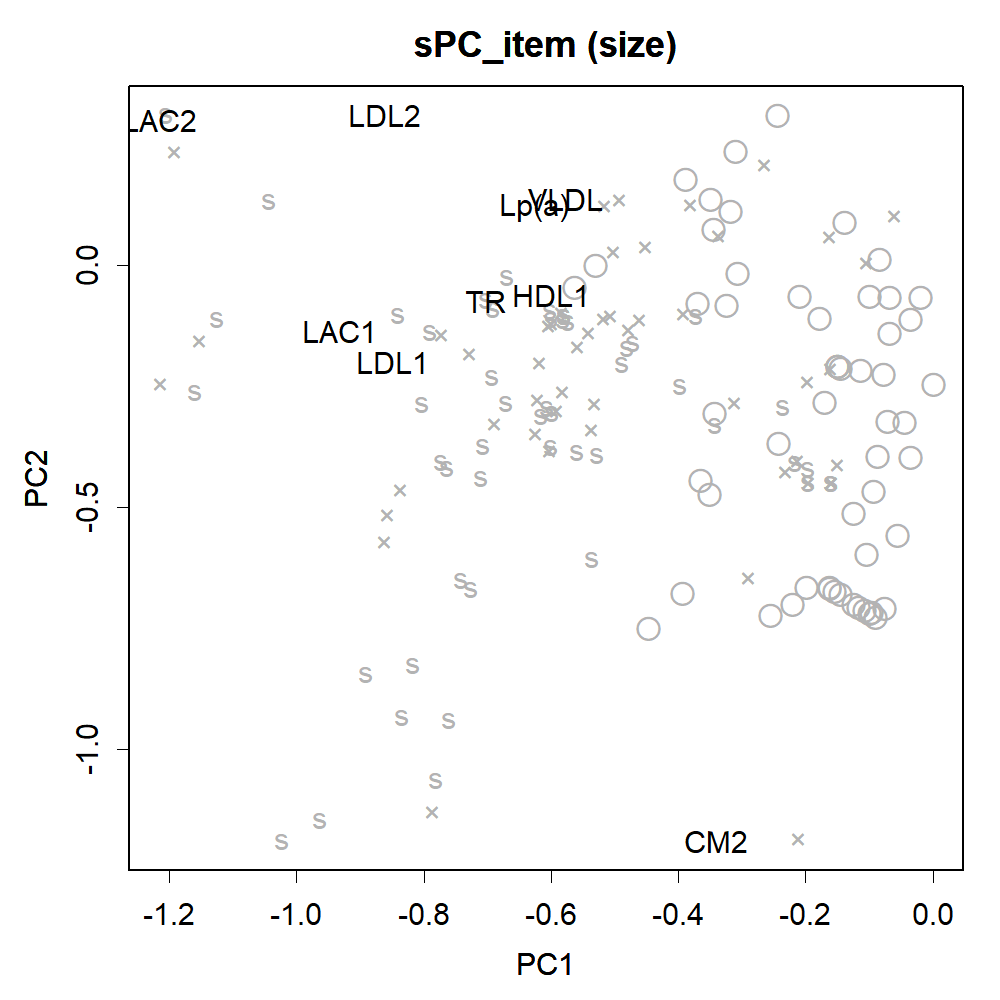

### PC12ookisa.png

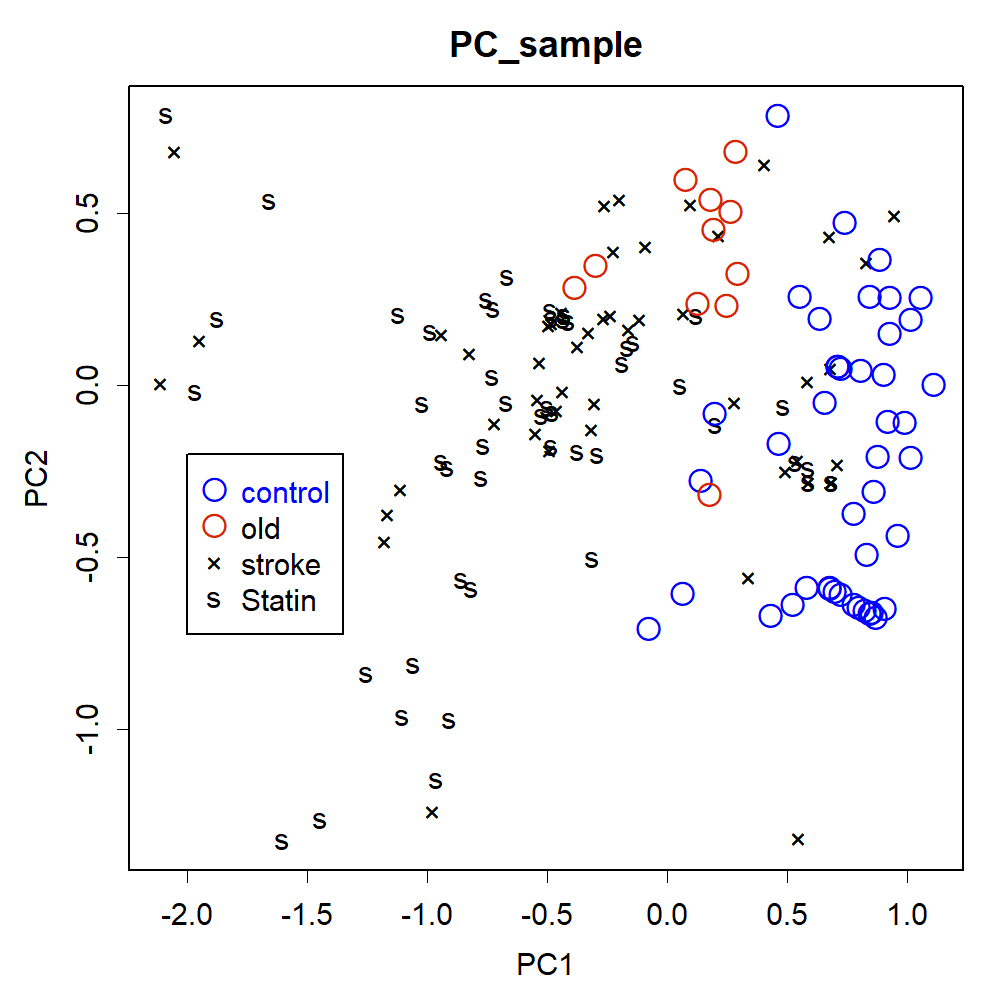

### PC12orig.png

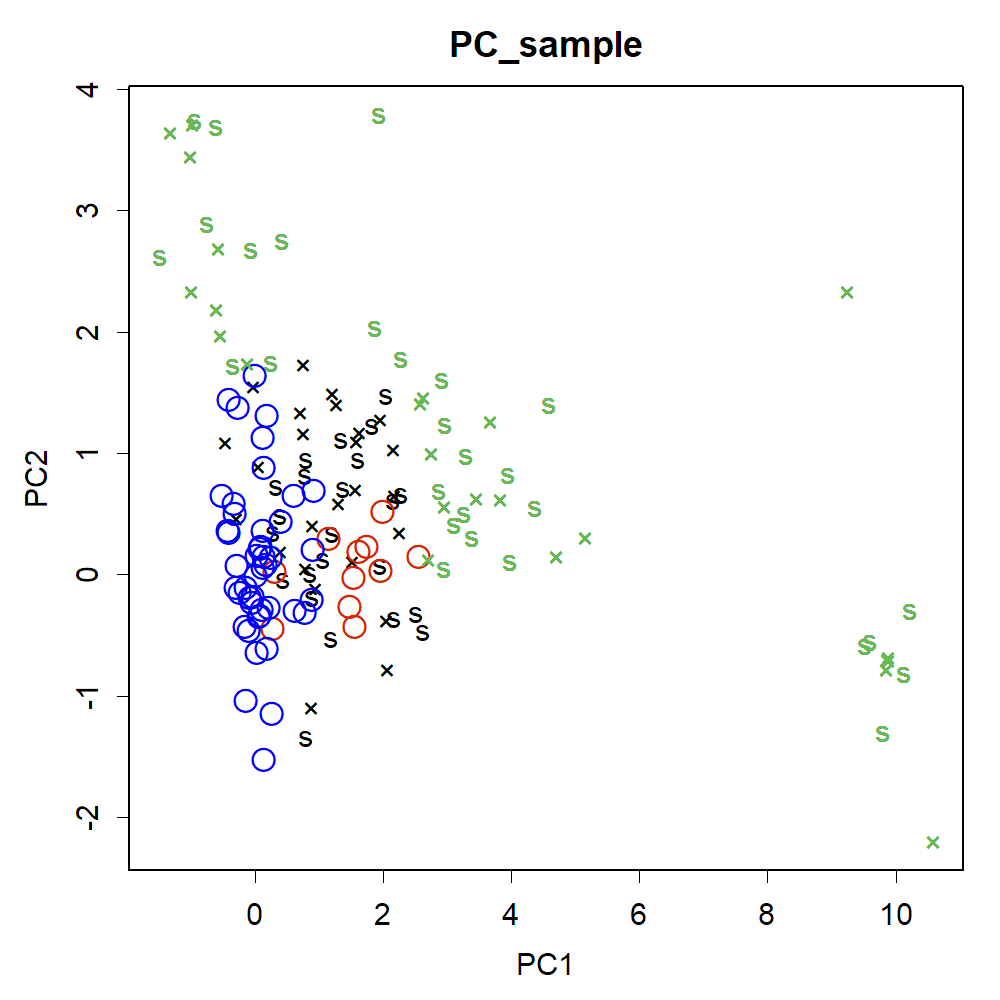

### PC34.png

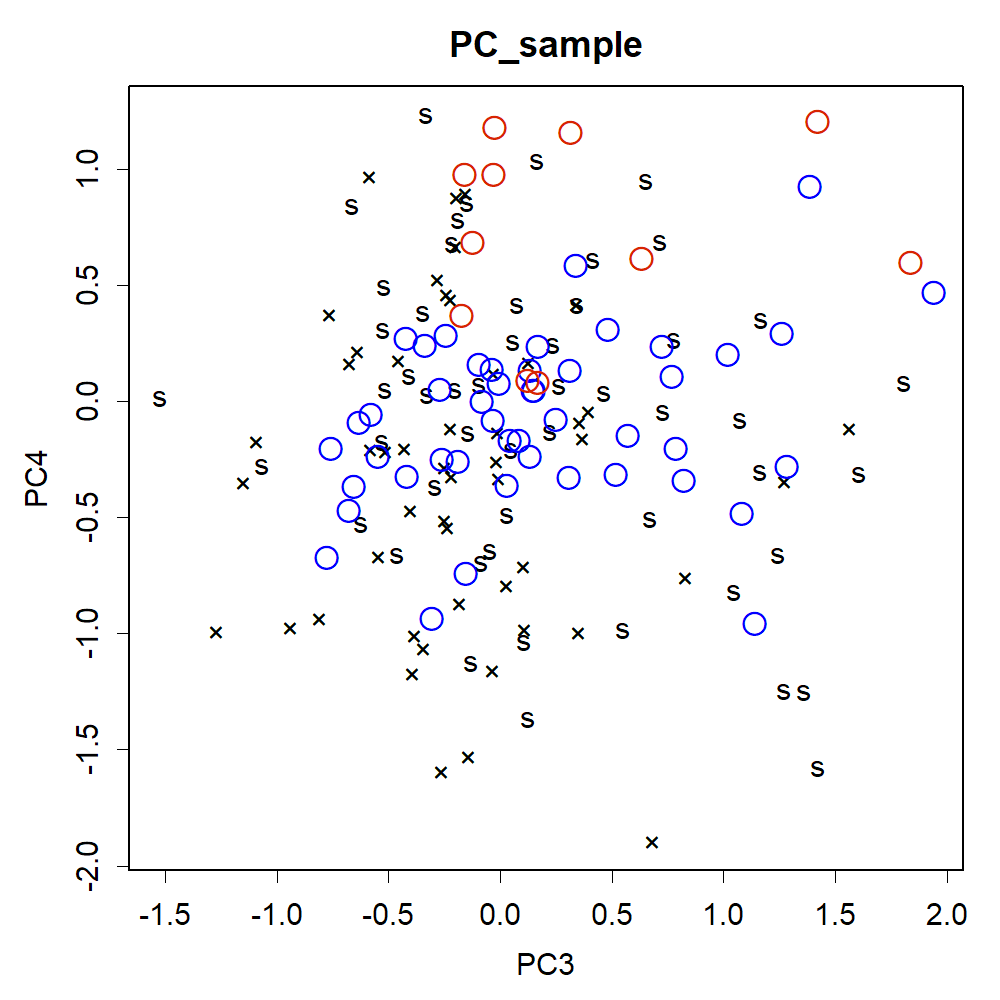

### PC34A.png

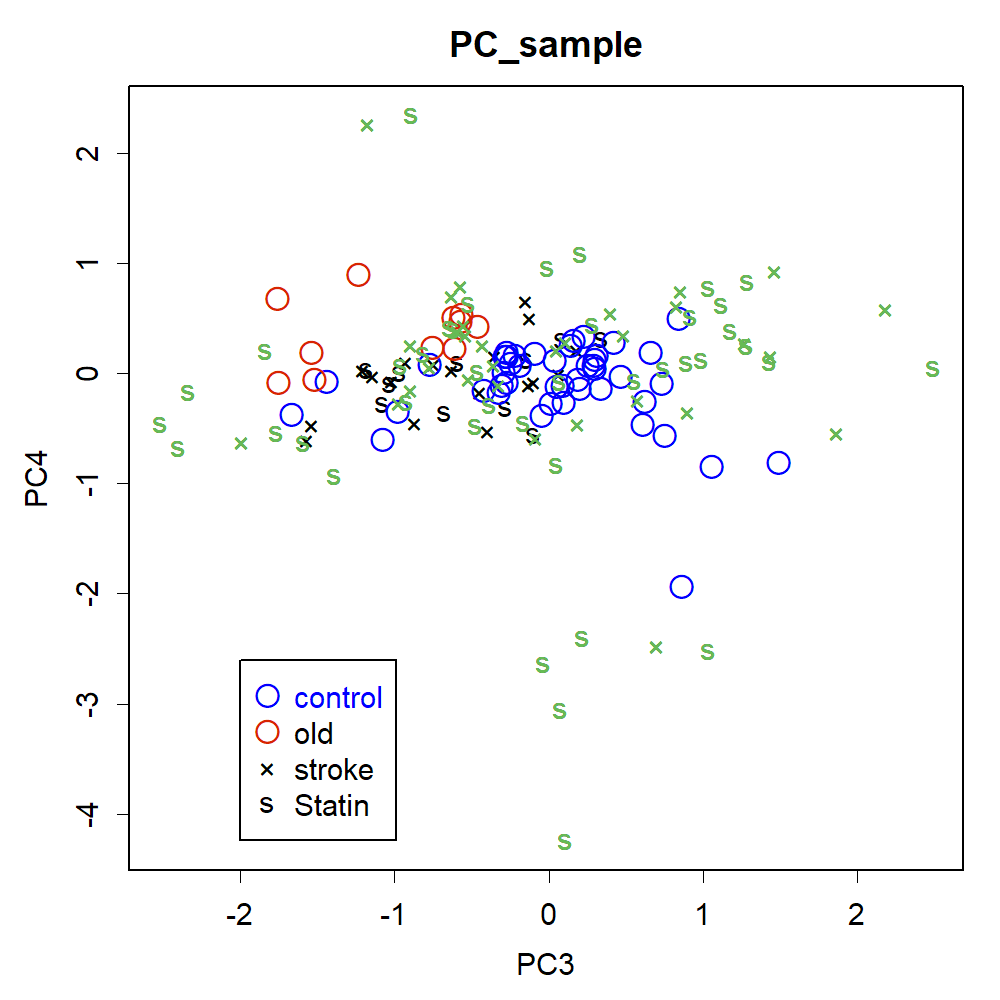

### PC34Furui.png

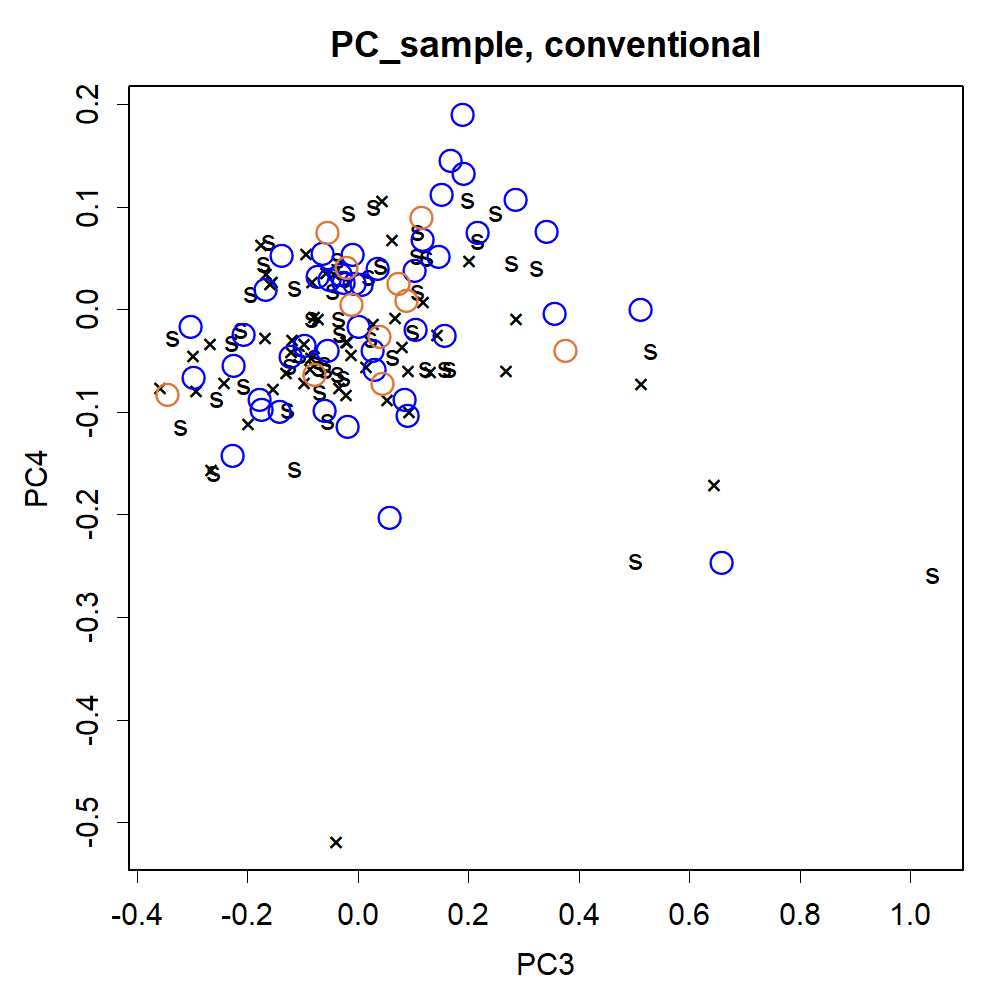

### PC34item.png

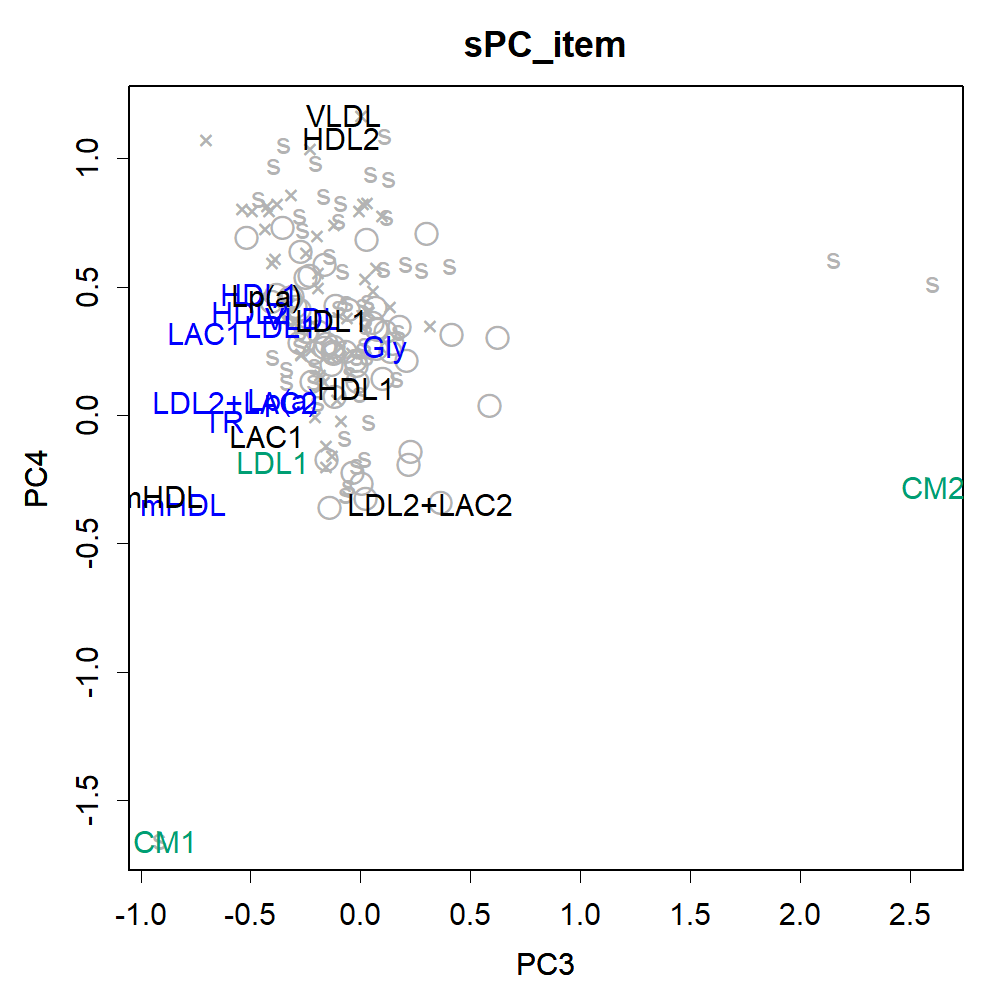

### PC34itemA.png

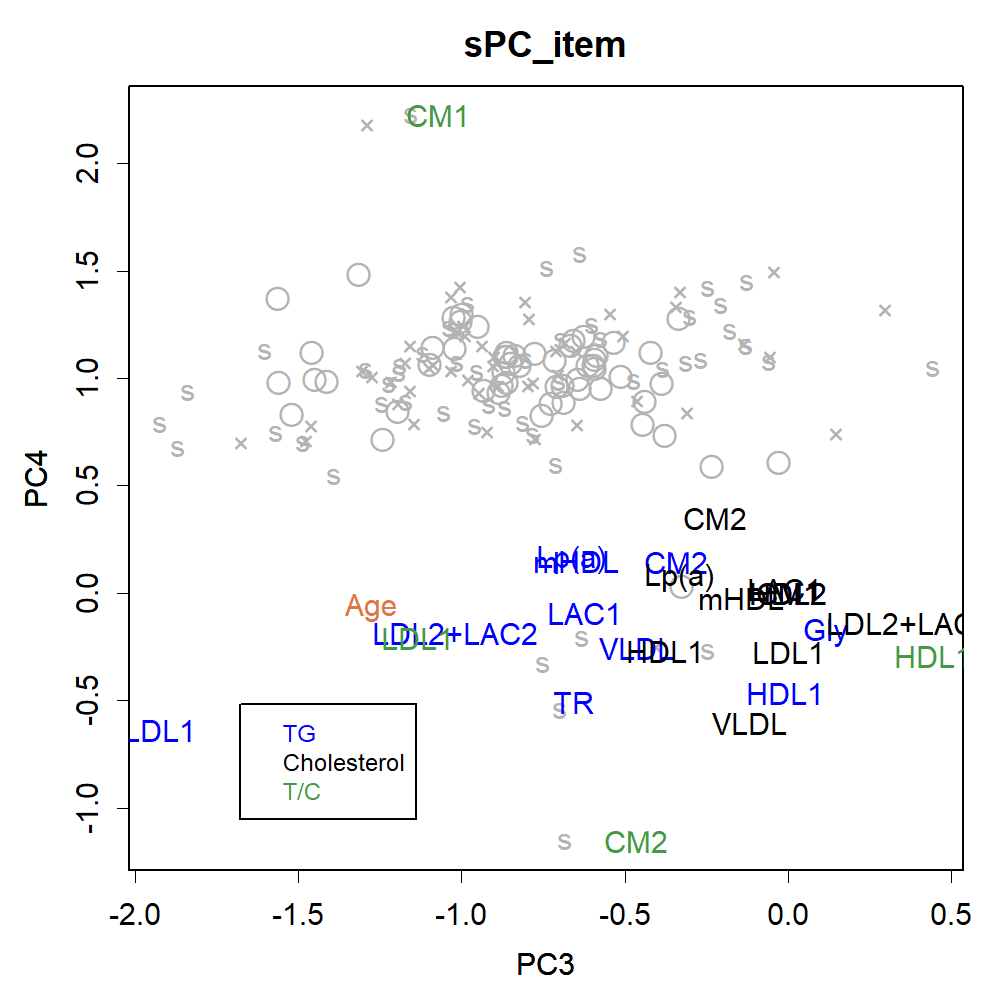

### PC34itemFurui.png

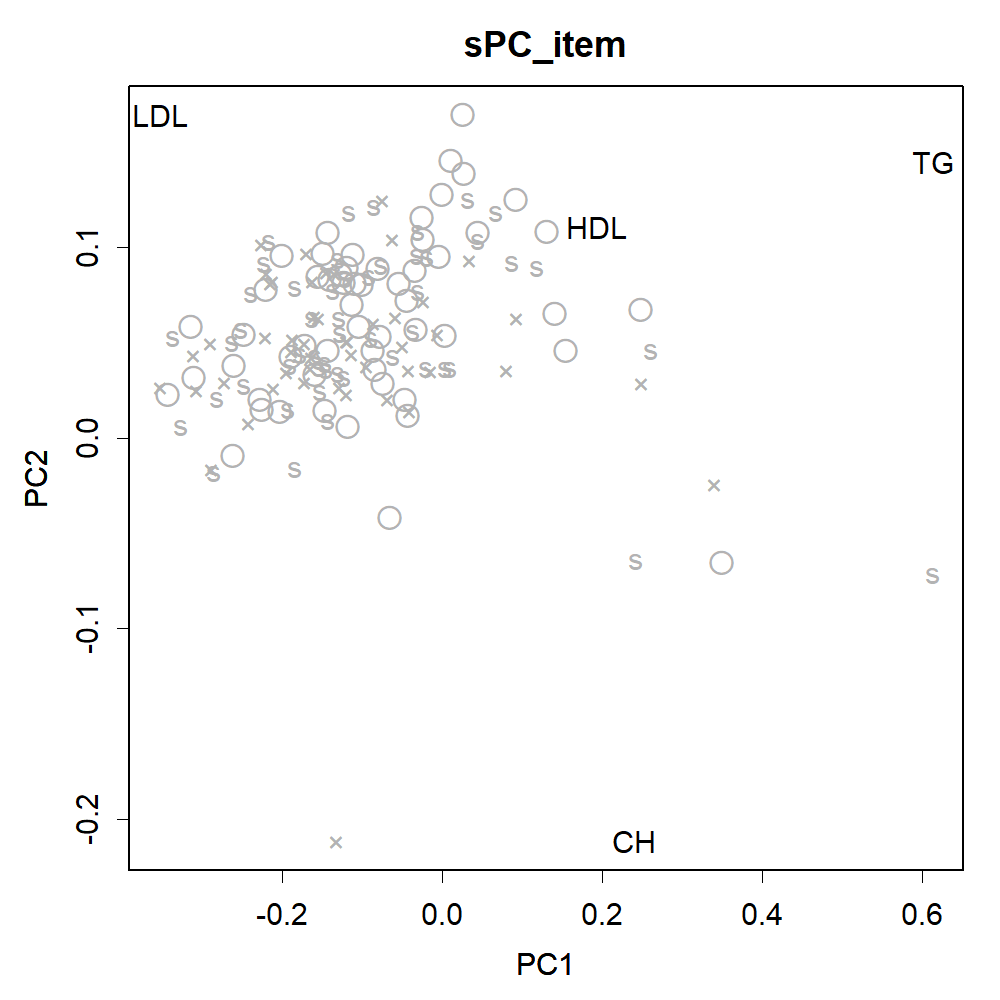

### PC34itemOOKISA.png

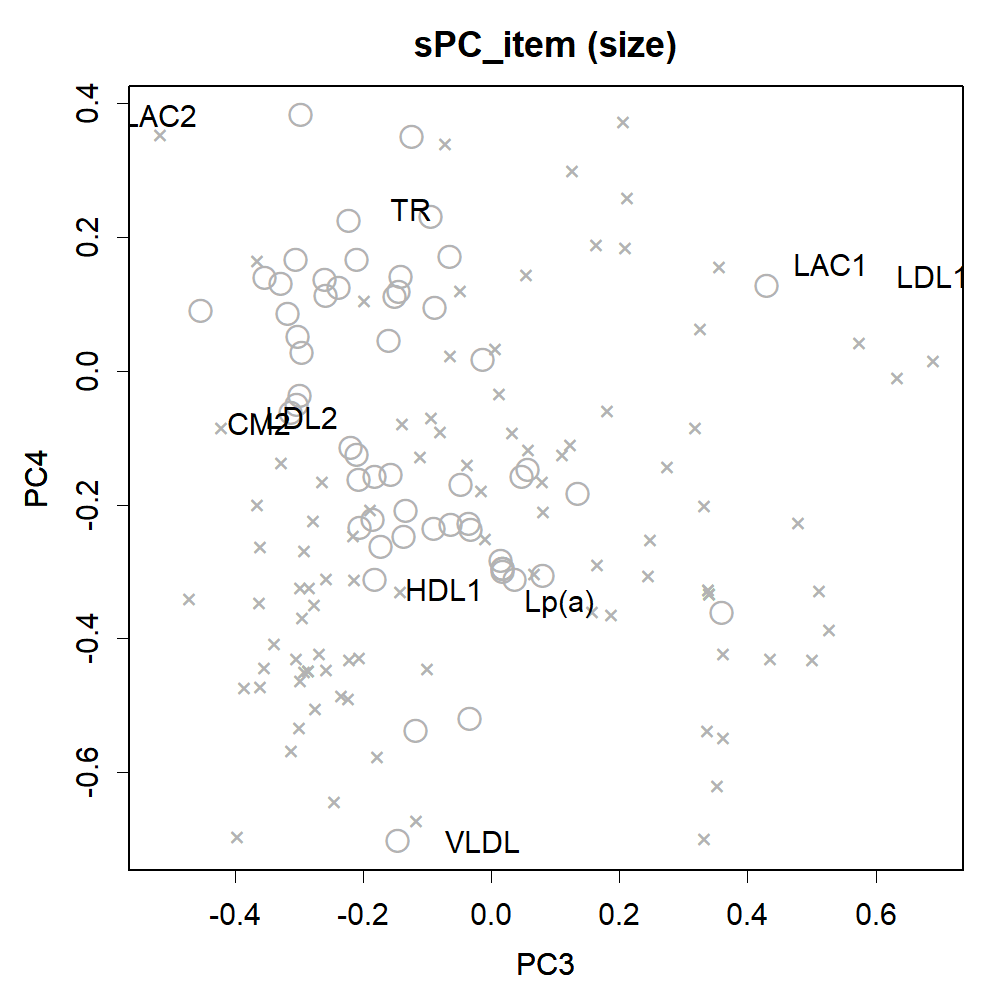

### PC34ookisa.png

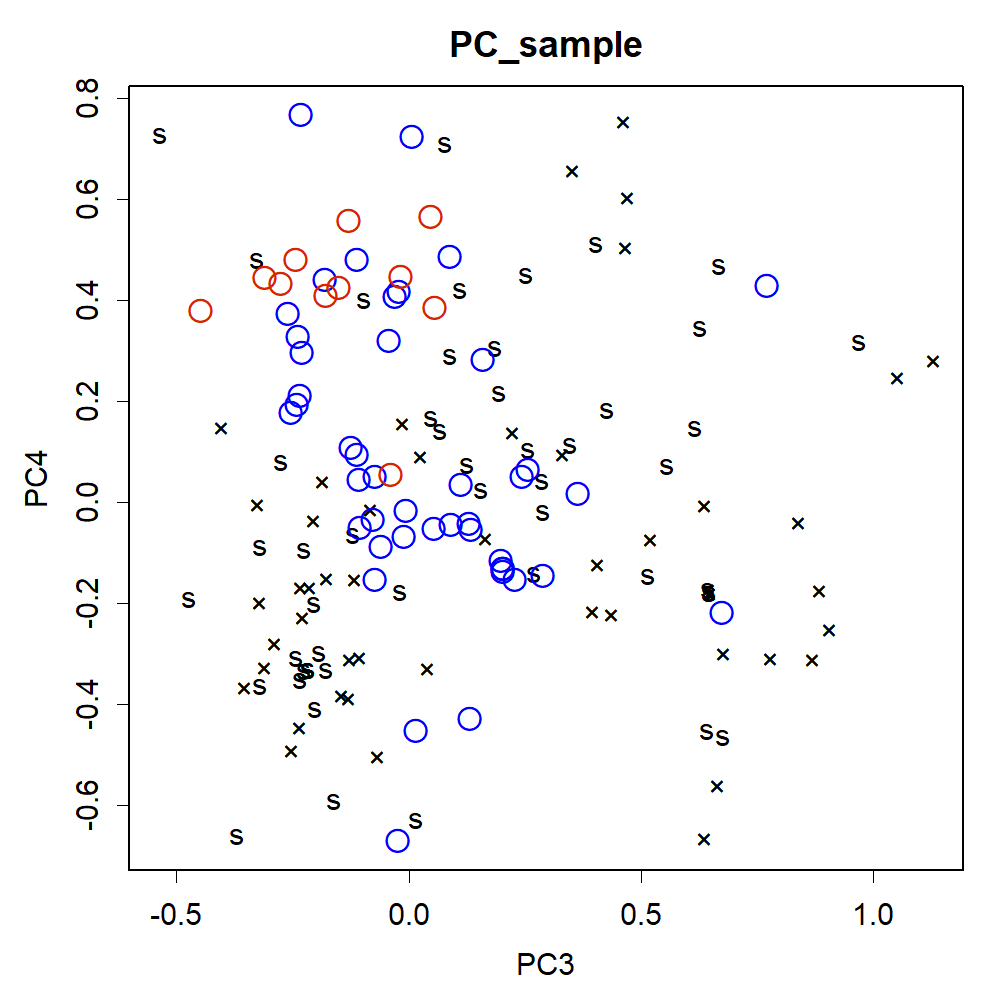

### PC34orig.png

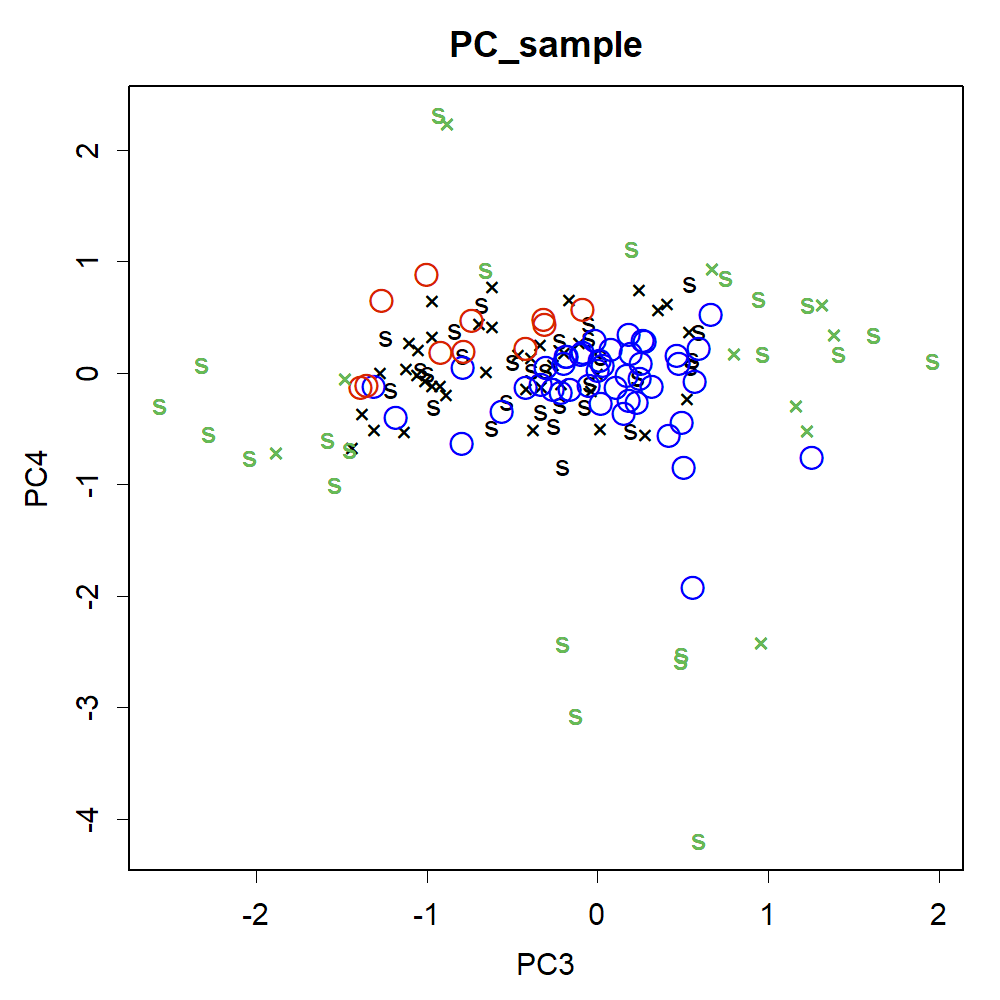

### PC56.png

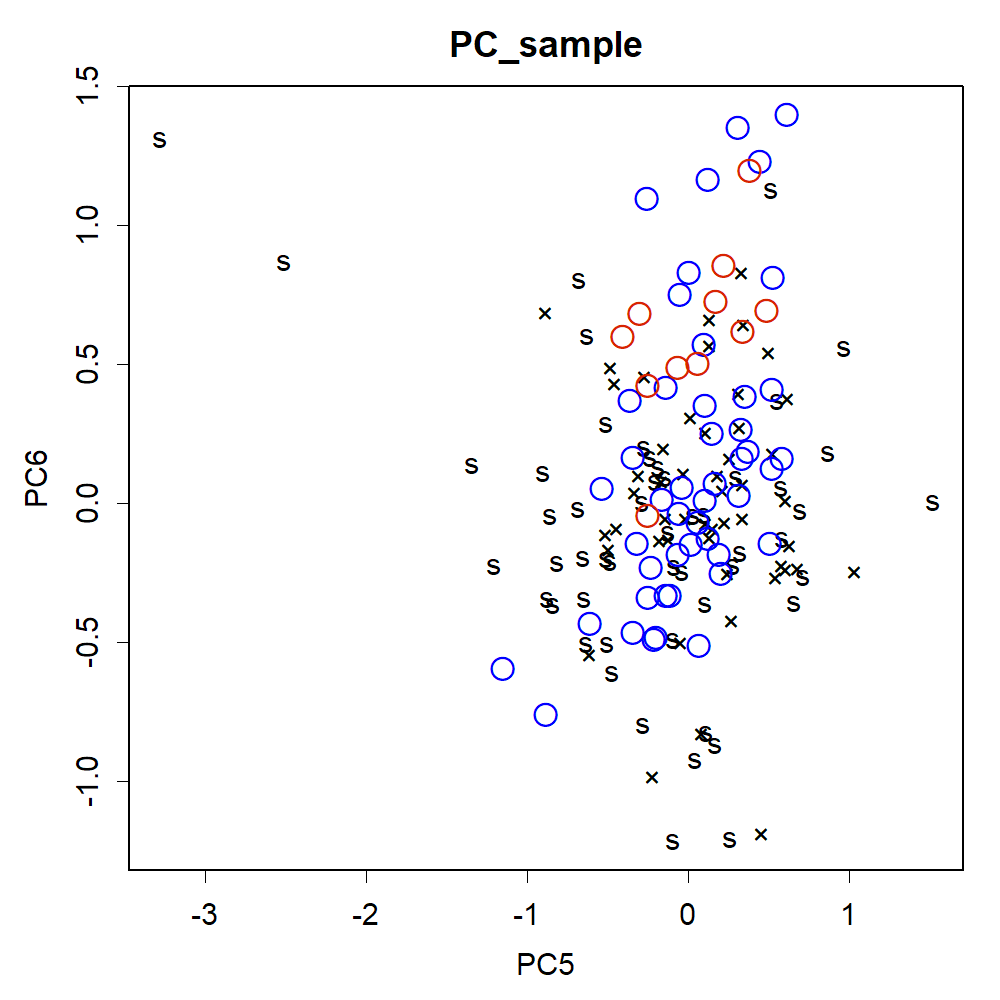

### PC56item.png

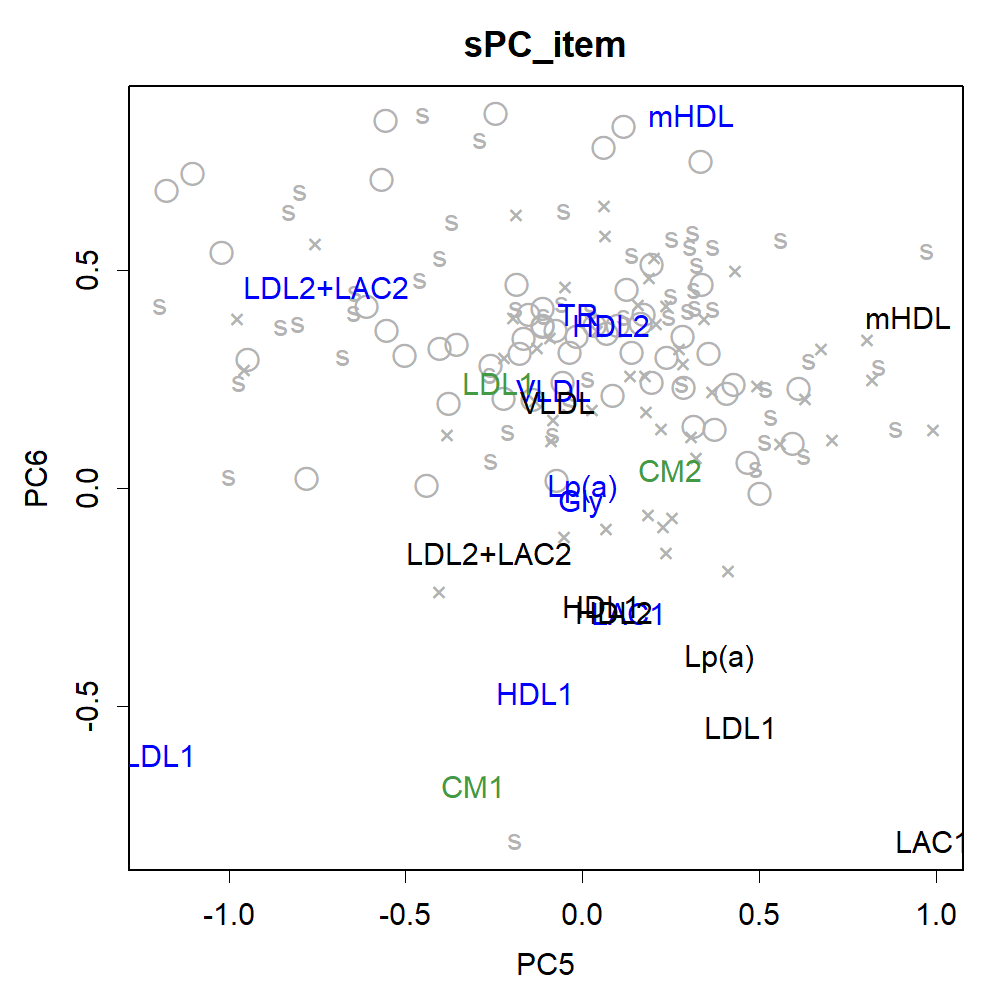

### PC56itemOOKISA.png

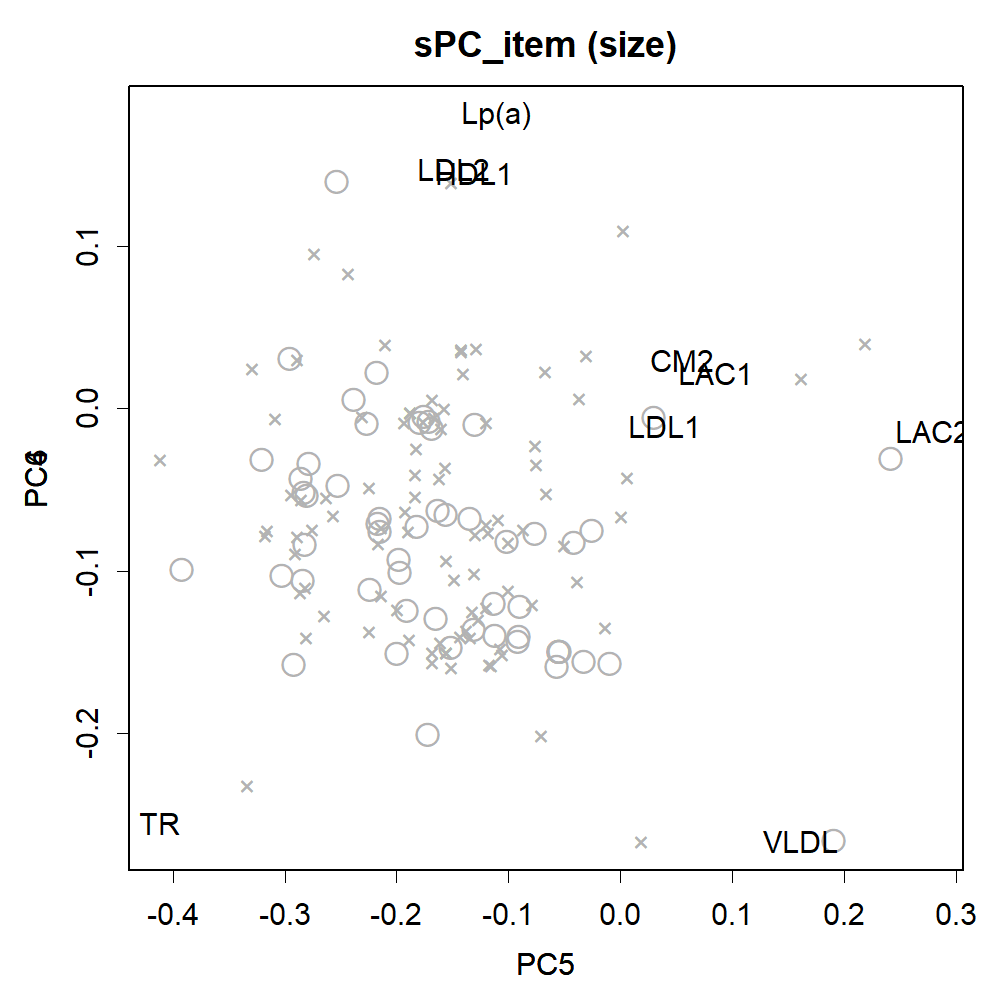

### PC56ookisa.png

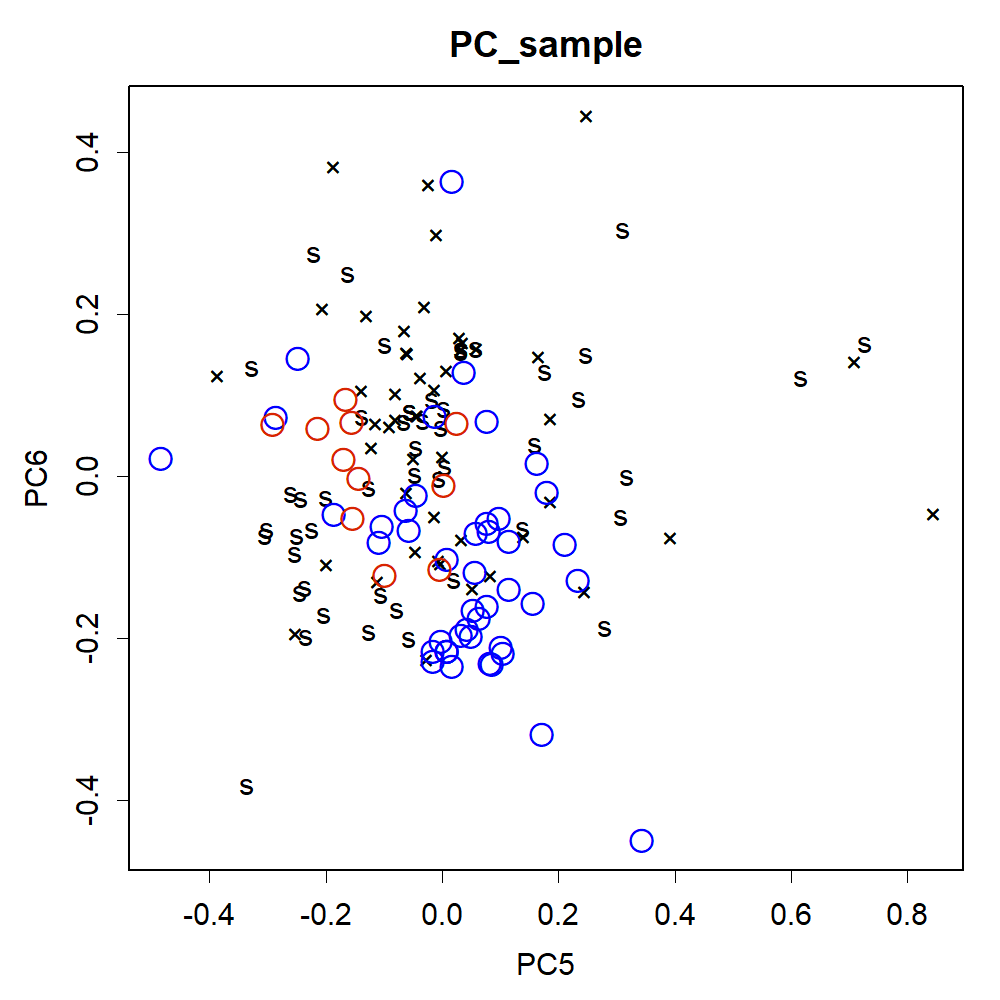

### PC56Orig.png

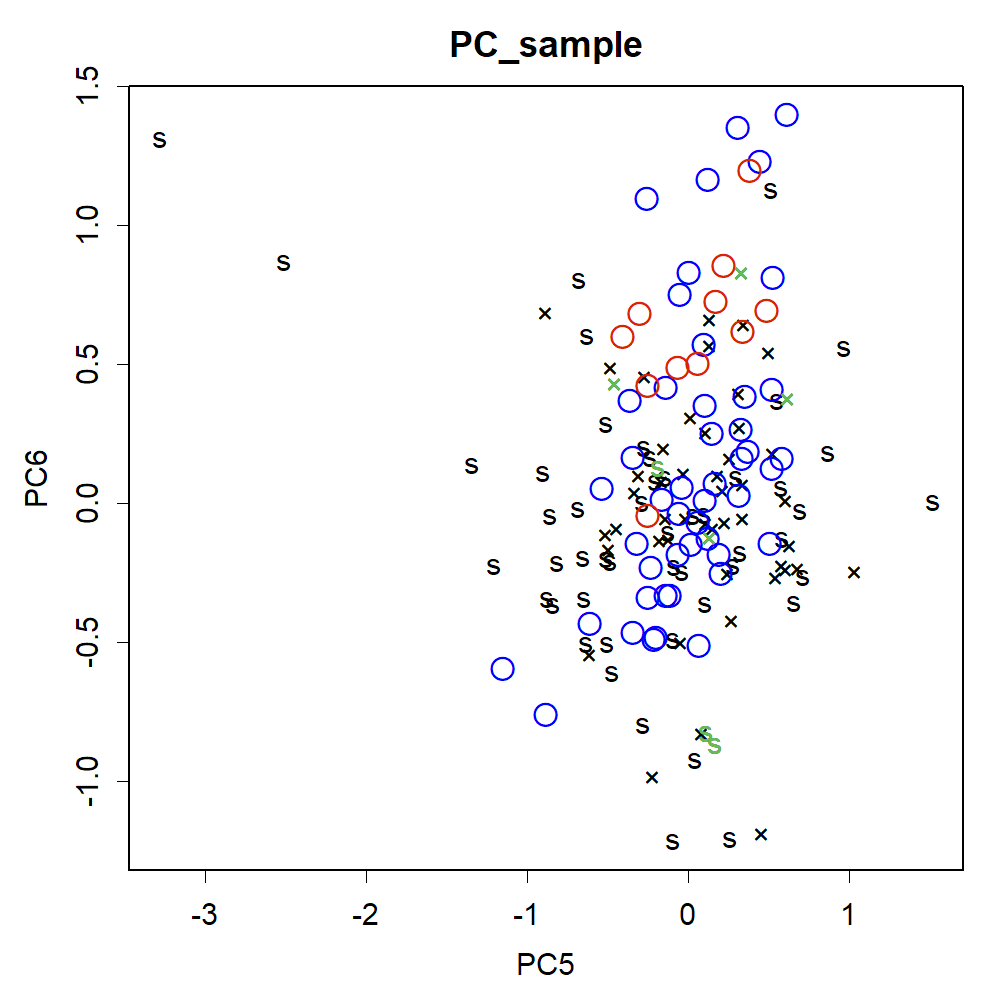

### PC78.png

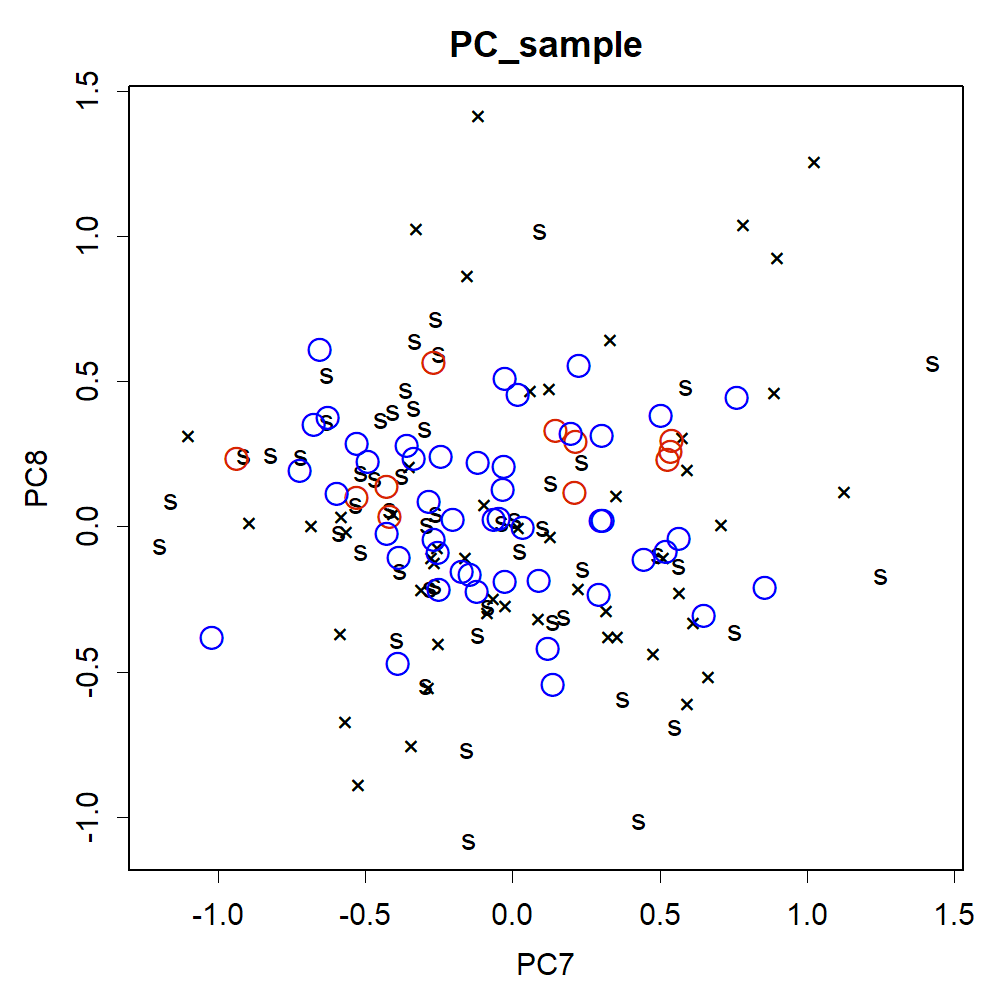

### PC78item.png

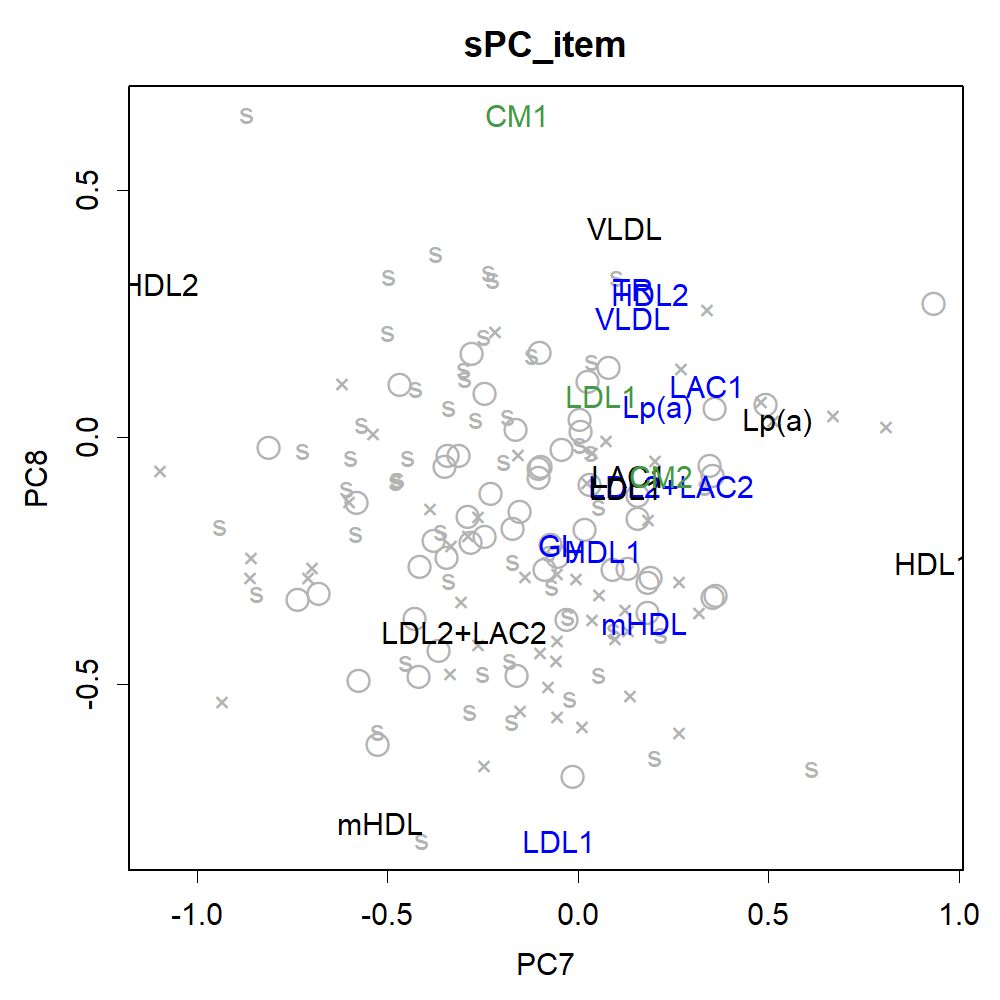

### PC910.png

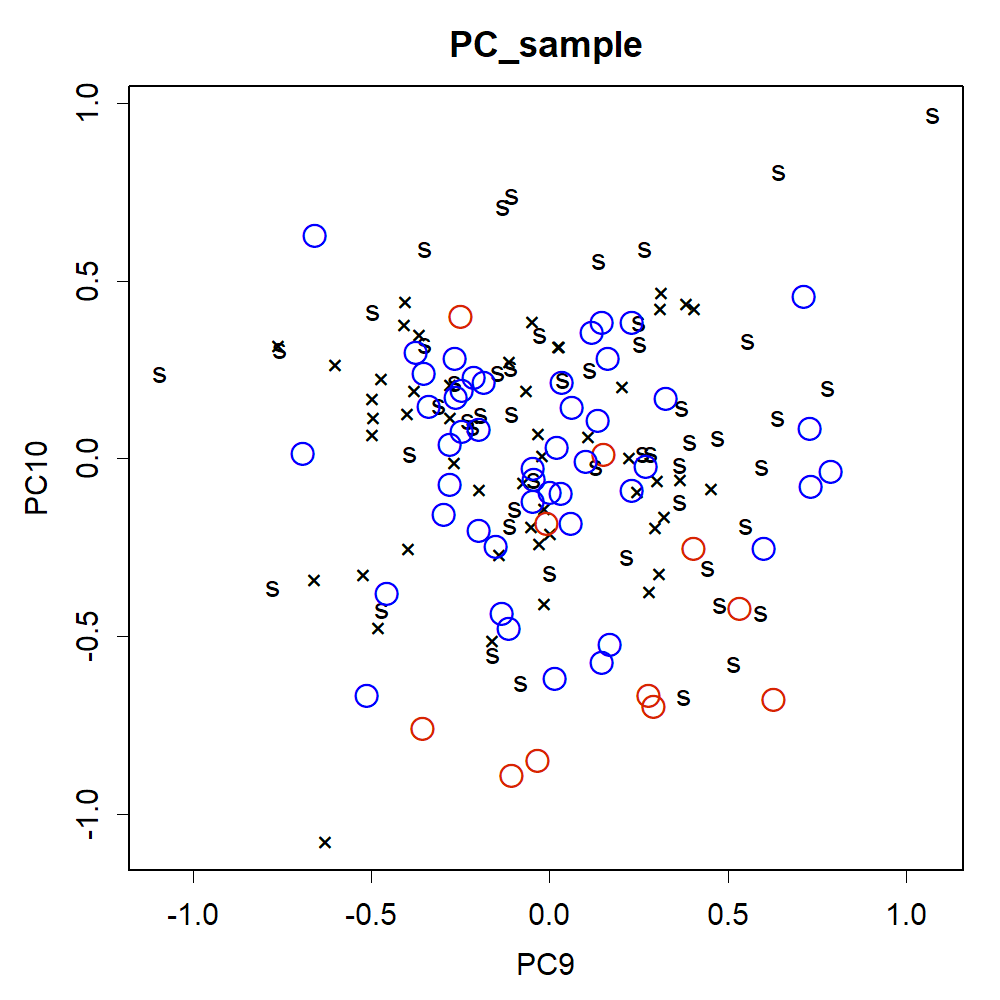

### PC910item.png

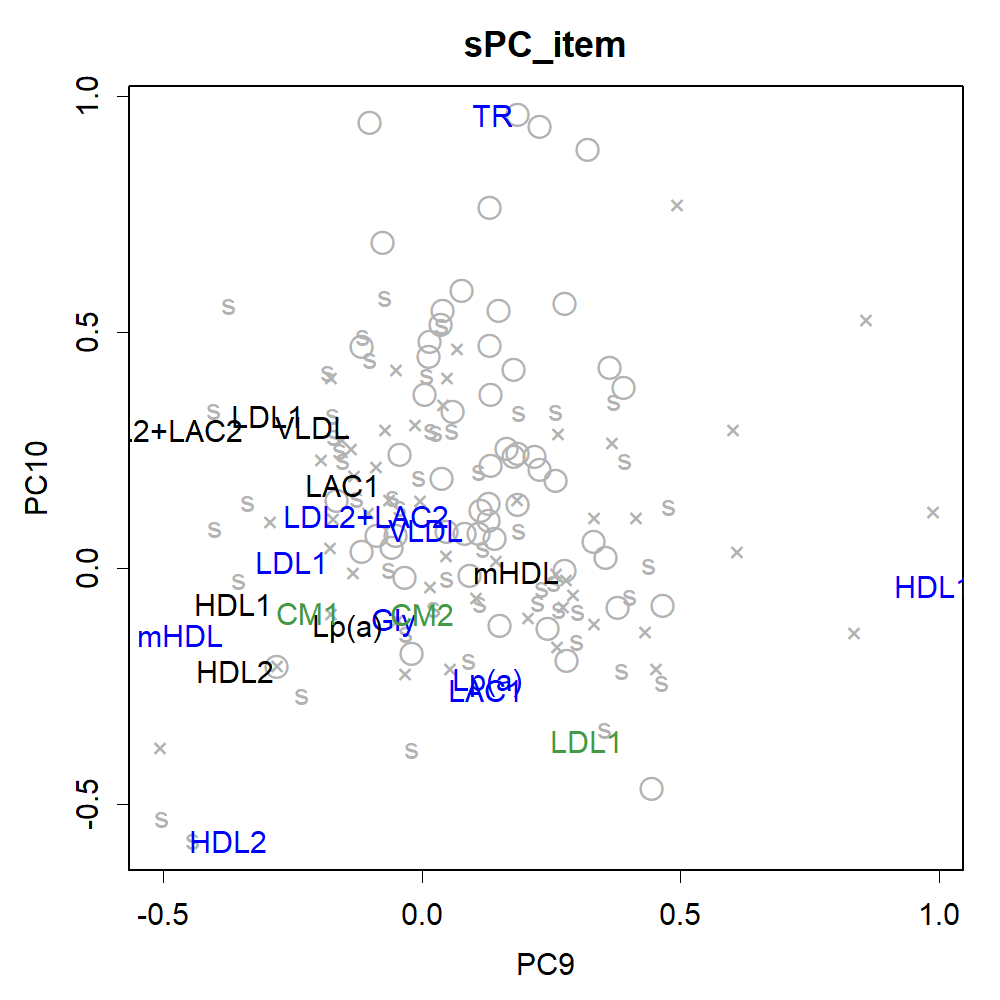

### QQ.png

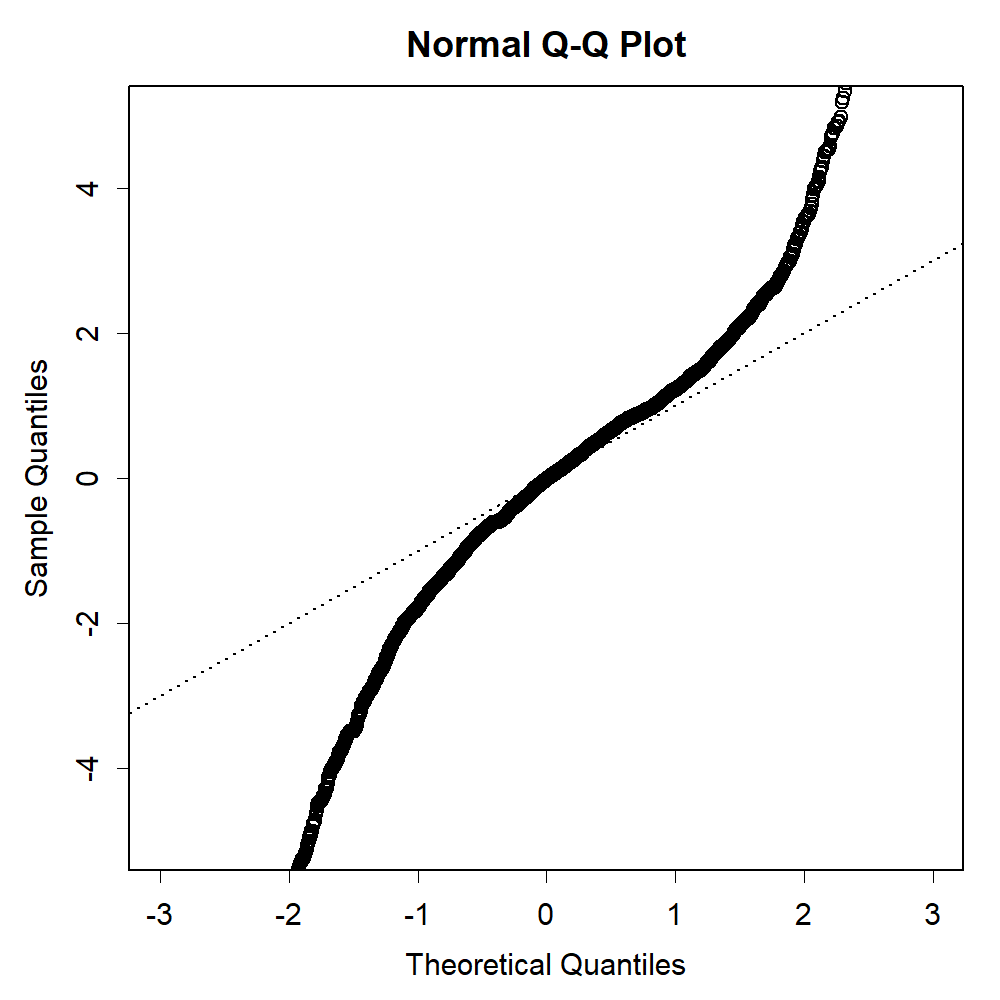

### sizetocontentLAC1.png

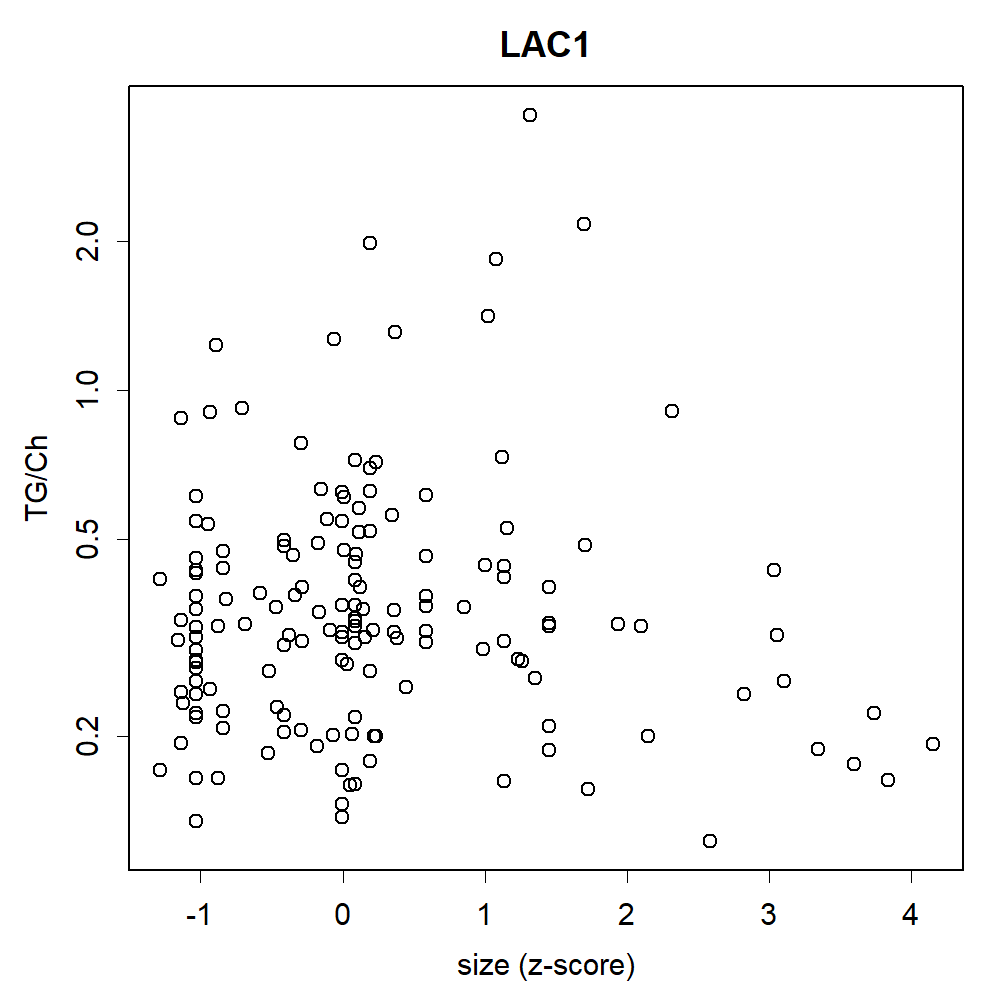

### sizetocontentLDL1.png

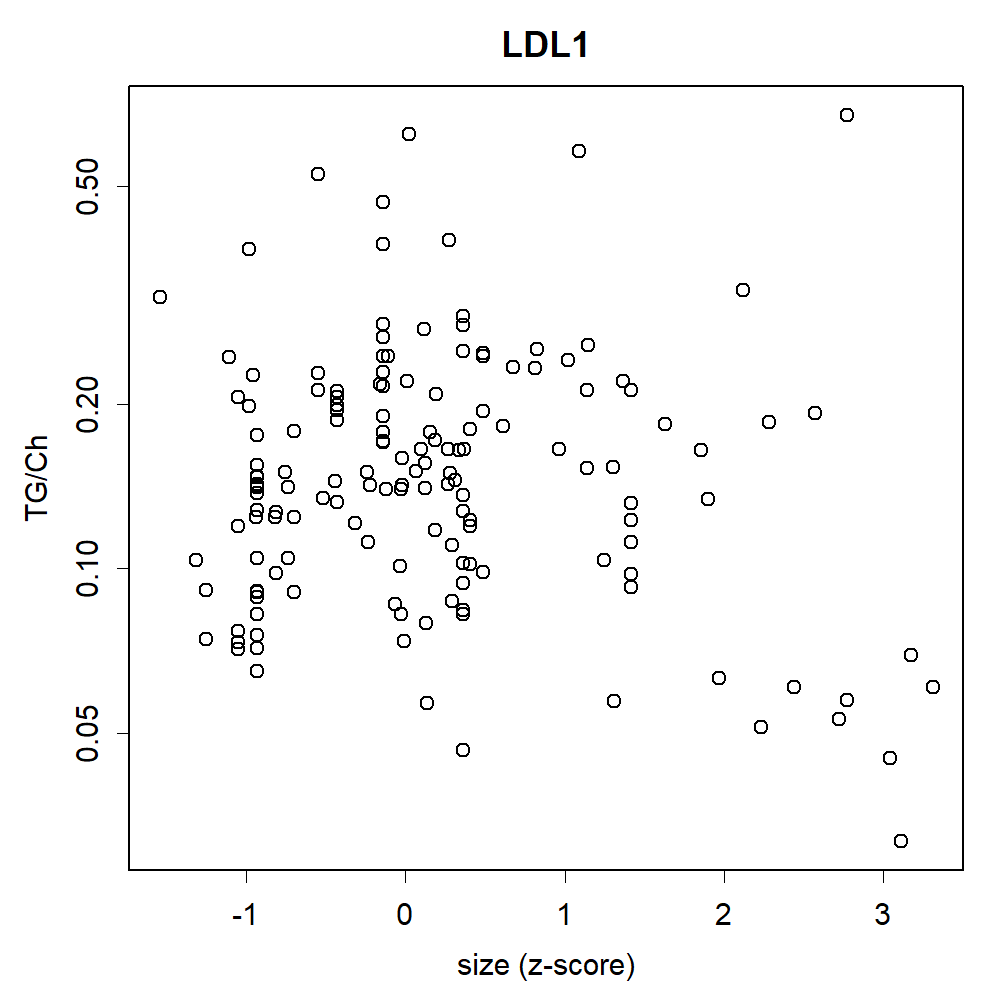

### TR.png

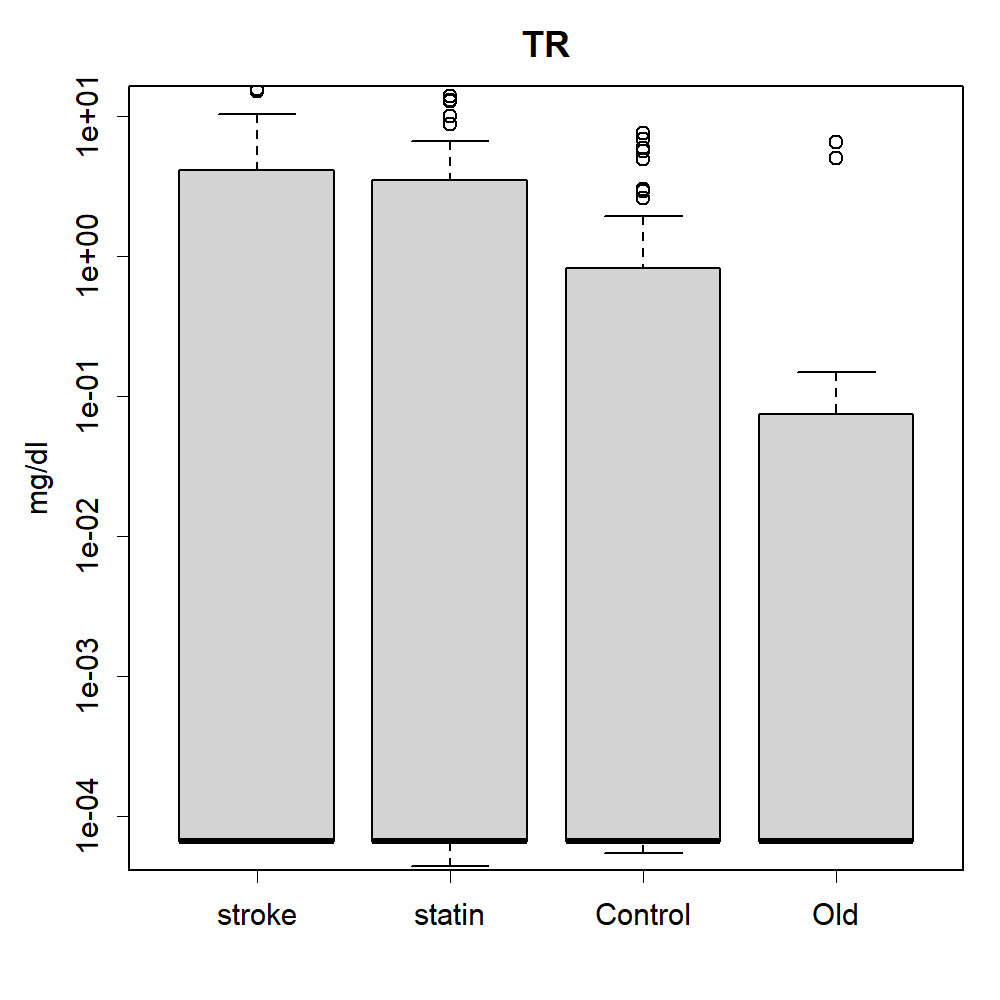

### TR.png

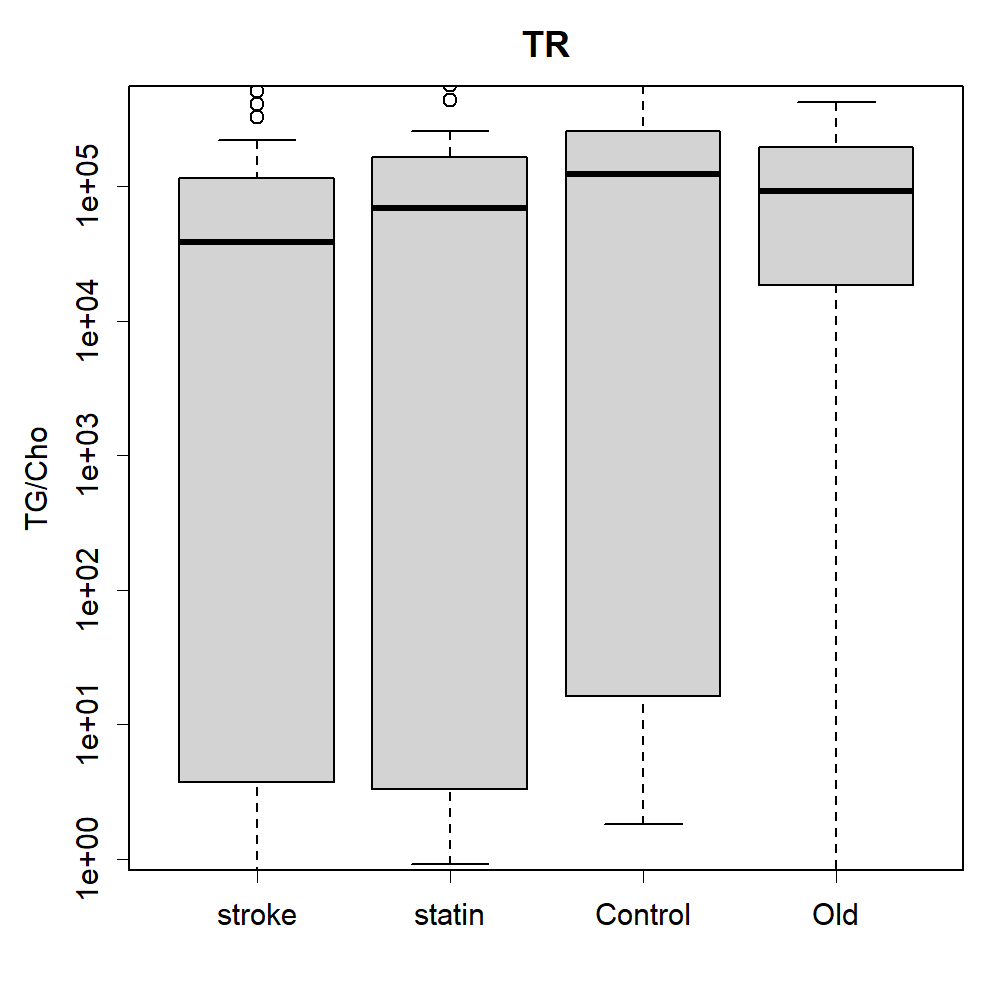

### TR.png

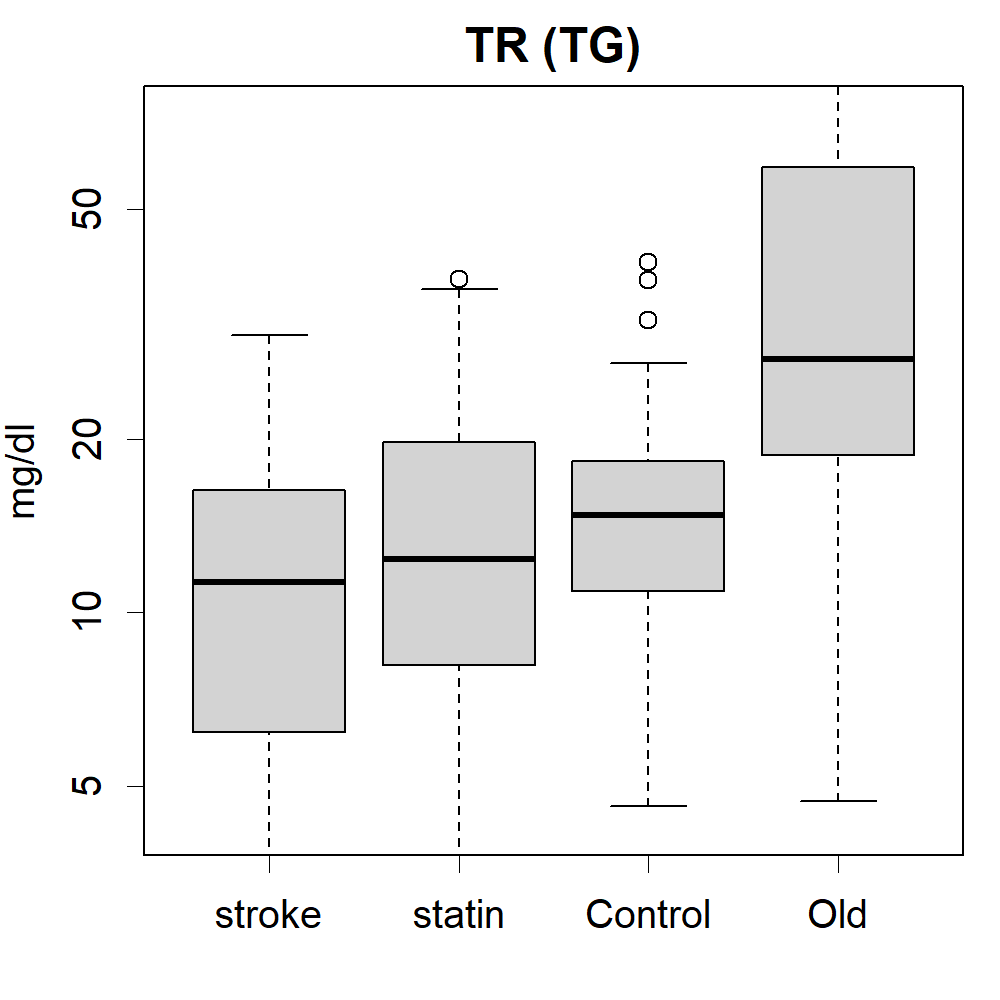

### VLDL.png

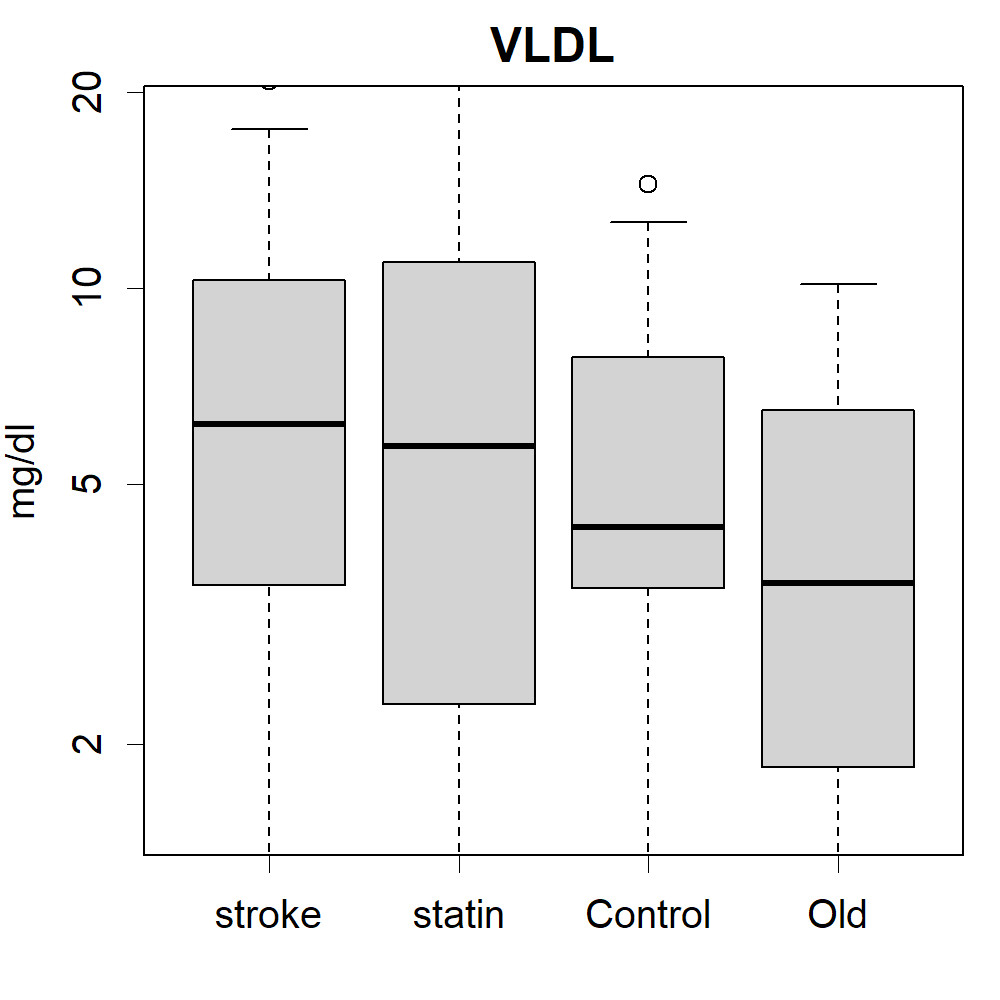

### VLDL.png

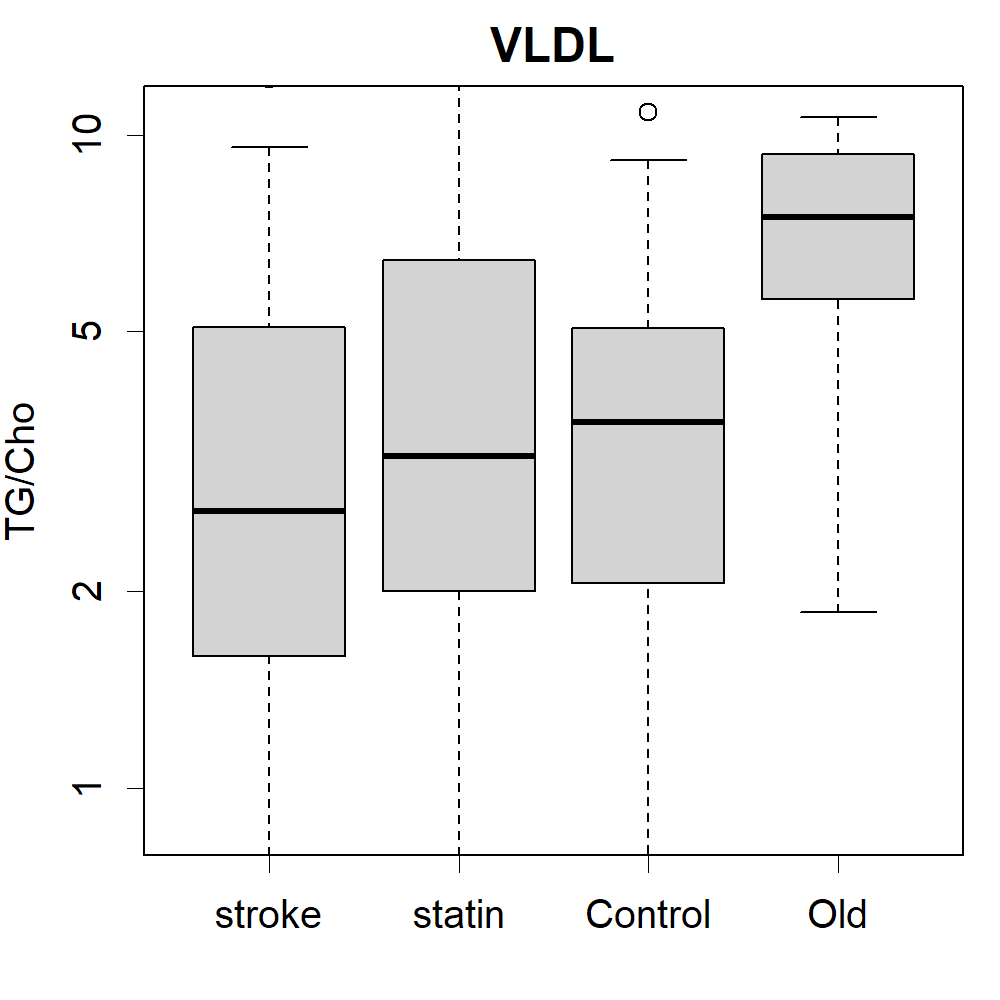

### VLDL.png

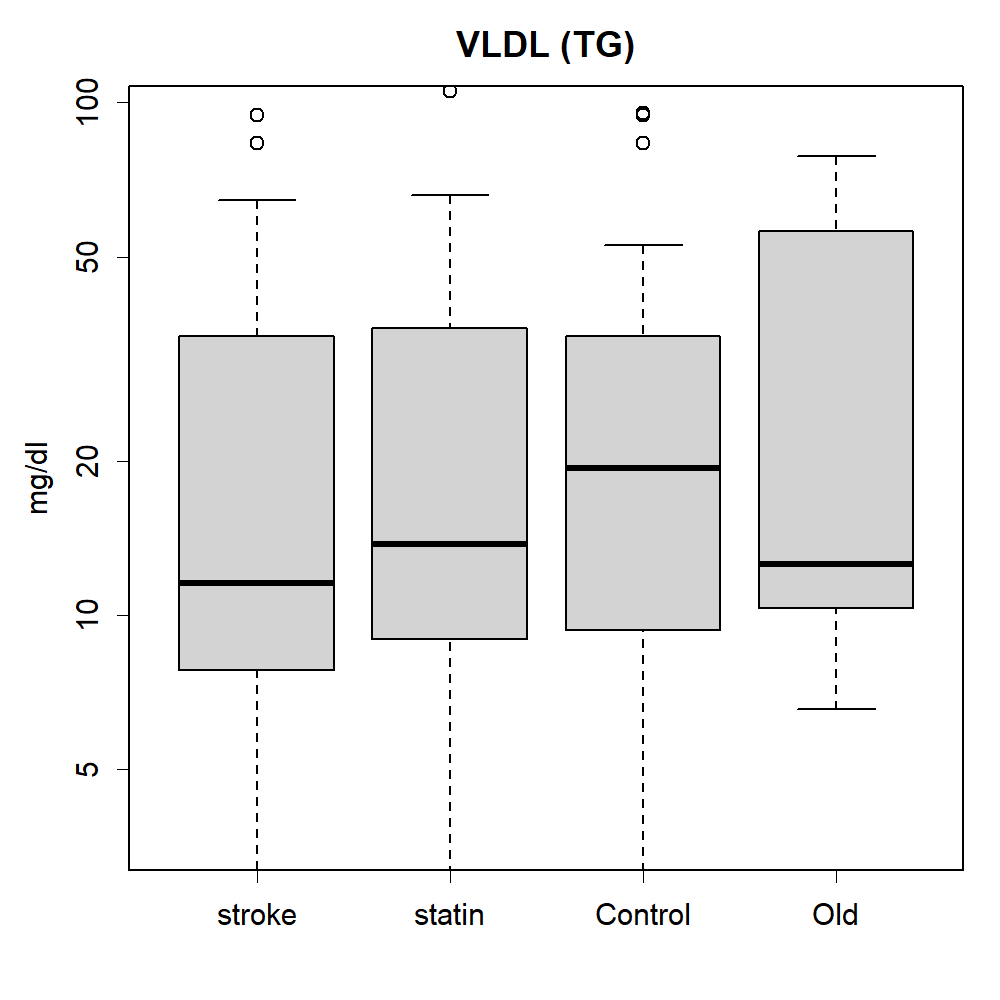
